# Multi-cancer early detection tests for general population screening: a systematic literature review

**DOI:** 10.1101/2024.02.14.24302576

**Authors:** Ros Wade, Sarah Nevitt, Yiwen Liu, Melissa Harden, Claire Khouja, Gary Raine, Rachel Churchill, Sofia Dias

## Abstract

**Background:** General population cancer screening in the UK is limited to selected cancers. Blood-based multi-cancer early detection (MCED) tests aim to detect potential cancer signals from multiple cancers in the blood. The use of an MCED test for population screening requires a high specificity and a reasonable sensitivity to detect early-stage disease, so that the benefits of earlier diagnosis and treatment can be realised.

**Objective:** To undertake a systematic literature review of the clinical effectiveness evidence on blood-based MCED tests for screening.

**Methods:** Comprehensive searches of electronic databases (including MEDLINE and Embase) and trial registers were undertaken in September 2023 to identify published and unpublished studies of MCED tests. Test manufacturer websites and reference lists of included studies and pertinent reviews were checked for additional studies. The target population was individuals aged 50 to 79 years without clinical suspicion of cancer. Outcomes of interest included test accuracy, number and proportion of cancers detected (by site and stage), time to diagnostic resolution, mortality, potential harms, health-related quality of life (HRQoL), acceptability and satisfaction. Risk of bias was assessed using the QUADAS-2 checklist. Results were summarised using narrative synthesis. Stakeholders contributed to protocol development, report drafting, and interpretation of review findings.

**Results:** Over 8000 records were identified. Thirty-six studies met the inclusion criteria: one ongoing randomised controlled trial (RCT), 13 completed cohort studies, 17 completed case-control studies and five ongoing cohort or case-control studies. Individual tests claimed to detect from three to over 50 different types of cancer. Diagnostic accuracy of currently available MCED tests varied substantially: Galleri® (GRAIL) sensitivity 20.8% to 66.3%, specificity 98.4% to 99.5% (3 studies); CancerSEEK (Exact Sciences) sensitivity 27.1% to 62.3%, specificity 98.9% to 99.1% (2 studies); SPOT-MAS™ (Gene Solutions) sensitivity 72.4% to 100%, specificity 97.0% to 99.9% (2 studies); TruCheck™ (Datar Cancer Genetics) sensitivity 90.0%, specificity 96.4% (1 study); CDA (AnPac Bio) sensitivity 40.0%, specificity 97.6% (1 study). AICS® (Ajinomoto) screens for individual cancers separately, so no overall test performance statistics are available. Where reported, sensitivity was lower for detecting earlier stage cancers (Stage I-II) compared with later stage cancers (Stage III-IV). Studies of seven other MCED tests at an unclear stage of development were also summarised.

**Limitations:** Study selection was complex; it was often difficult to determine the stage of development of MCED tests. The evidence was limited; there were no completed RCTs and most included studies had a high overall risk of bias, primarily owing to limited follow-up of participants with negative test results. Only one study of Galleri recruited asymptomatic individuals aged over 50 in the USA, however, study results may not be representative of the UK general screening population. No meaningful results were reported relating to patient relevant outcomes, such as mortality, potential harms, HRQoL, acceptability or satisfaction.

**Conclusions:** All currently available MCED tests reported high specificity (>96%). Sensitivity was highly variable and influenced by study design, population, reference standard test used and length of follow-up.

**Future work:** Further research should report patient relevant outcome and consider patient and service impacts.

**PROSPERO registration number:** CRD42023467901

**Funding:** This project was funded by the National Institute for Health and Care Research (NIHR) Evidence Synthesis programme (NIHR161758) and will be published in full in the Health Technology Assessment Journal; Vol. XX, No. XX. See the NIHR Journals Library website for further project information.

**PLAIN LANGUAGE SUMMARY:** Cancer screening is only available for some cancers. New tests that look for signs of cancer in blood (blood-based multi-cancer early detection tests) are being developed; they aim to detect multiple different cancers at an early stage, when they are potentially more treatable. Taking account of stakeholder feedback, we reviewed all studies assessing the effectiveness of blood-based multi-cancer early detection tests for cancer screening. We thoroughly searched for relevant studies and found over 8000 records. We included 30 completed studies and six ongoing studies of 13 different tests. None of the studies were good quality, mainly because they didn’t properly check whether the test result might have been incorrect and participants with a negative test result actually had cancer. Most studies included participants who are different from the general UK population that would be likely to have this type test for cancer screening. None of the studies reported meaningful results for patient-relevant outcomes, such as death, potential harms, quality of life and acceptability. We found 14 completed studies assessing six tests that are currently available: Galleri® (GRAIL), CancerSEEK (Exact Sciences), SPOT-MAS™ (Gene Solutions), TruCheck™ (Datar Cancer Genetics), CDA (AnPac Bio) and AICS® (Ajinomoto). All of the tests were quite good at ruling out cancer, but their accuracy for finding cancer varied a lot, mostly because of differences in the study methods and characteristics of the included participants. The tests were better at finding more advanced cancers, which are potentially less curable than early cancers, so more research is needed to know whether tests would actually save lives. Better designed studies including participants similar to those who might get the test in the real world, and which report on patient-relevant outcomes and properly consider patient experience and impact on services, are needed. Several new studies are planned or underway.

**SCIENTIFIC SUMMARY:** *Background:* General population cancer screening in the UK is limited to selected cancers (cervical, breast, bowel and, for some high-risk individuals, lung). Most other cancers are detected after presentation of symptoms, when the disease tends to be at a more advanced stage and treatment options may be more limited. Blood-based multi-cancer early detection (MCED) tests aim to detect potential cancer signals (such as circulating cell-free deoxyribonucleic acid [cfDNA]) from multiple cancers in the blood. The use of an MCED test as a screening tool in a generally healthy, asymptomatic population, requires a high specificity and a reasonable sensitivity to detect early-stage disease, so that the benefits of earlier diagnosis and treatment can be realised. An MCED test embedded within a national population-based screening programme, in addition to existing cancer screening programmes, may increase the number of cancers diagnosed at an earlier stage. However, identification of cancers with no effective treatments, even at an early stage, may have no improvement on mortality or health-related quality of life (HRQoL). In addition, early screening of healthy people for such a wide range of cancers, and the expected lengthy time to diagnostic confirmation, may create anxiety and lead to unnecessary follow-up tests, when false positive test results occur.

*Objectives:* The aim of this project was to conduct a systematic review to assess the accuracy and clinical effectiveness, acceptability and feasibility of blood-based MCED tests for population-based screening.

*Methods:* Comprehensive searches of electronic databases (including MEDLINE and Embase) and trial registers were undertaken in September 2023. Test manufacturer websites and reference lists of included studies and pertinent reviews were checked for additional relevant studies. Published and unpublished prospective clinical trials and cohort studies of blood-based MCED tests for screening were sought. Studies assessing blood-based tests for assessing prognosis or therapeutic decision-making in patients with cancer were not eligible for inclusion. The target population was individuals aged 50 to 79 years without clinical suspicion of cancer and who had not been diagnosed with, or received treatment for, cancer within the last three years. As insufficient relevant studies were identified within the target population, studies that included patients known to have cancer (i.e., case-control studies) and studies that included individuals with a different age range were included. Outcomes of interest were test accuracy (including sensitivity, specificity, positive and negative predictive values), number and proportion of cancers detected (by site and stage), mortality, time to diagnostic resolution, incidental findings, additional tests and procedures, potential harms, HRQoL, acceptability and satisfaction. A standardised data extraction form was developed and piloted. Data on the intervention(s), participant characteristics, setting, study design, reference standard test(s) used, and relevant outcomes were extracted from included studies by one reviewer and independently checked by a second reviewer. Risk of bias and applicability were assessed using the QUADAS-2 (Quality Assessment of Diagnostic Accuracy Studies) checklist by one reviewer and independently checked by a second. Disagreements were resolved through discussion. Results were summarised using narrative synthesis. Stakeholders contributed to protocol development, report drafting, and interpretation of review findings.

*Results:* The electronic searches identified 8,069 records; 228 full texts were further reviewed. Eleven additional records were identified from searching MCED test manufacturer websites. Study selection was complex; it was often difficult to determine whether studies assessed technologies at an early stage of development, or the final or near-final version of the test. Thirty-six studies, evaluating thirteen MCED tests or technologies, met the review inclusion criteria: one ongoing randomised controlled trial (RCT), 13 completed cohort studies, 17 completed case-control studies, four ongoing cohort studies and one ongoing case-control study. Studies assessed the following MCED tests: Galleri® test (GRAIL, Menlo Park, California), CancerSEEK (Exact Sciences, Madison, Wisconsin), SPOT-MAS™ (Gene Solutions, Ho Chi Minh City, Vietnam), TruCheck™ (Datar Cancer Genetics, Beyreuth, Germany), CDA (Cancer Differentiation Analysis; AnPac Bio, Shanghai, China) and AICS® test (AminoIndex Cancer Screening; Ajinomoto, Tokyo, Japan). Other MCED technologies included in the review, that were at an unclear stage of development and did not appear to be available for use, were: Aristotle® (StageZero Life Sciences, Richmond, Ontario), CancerenD24 (manufacturer unknown), OncoSeek® (SeekIn Inc, San Diego, California), SeekInCare® (SeekIn Inc, San Diego, California), OverC™ (Burning Rock Biotech, Guangzhou, China), Carcimun-test (Carcimun Biotech, Garmisch-Partenkirchen, Germany) and SpecGastro test (manufacturer unknown). Technologies that appeared to be at a very early stage of development did not meet the inclusion criteria for the review. Individual MCED tests and technologies claimed to detect from three to over 50 different types of cancer. Owing to the differences in the number of cancer types detected, study design and populations, statistical pooling of results was not considered appropriate.

*Studies of MCED tests available for use:* The risk of bias assessment identified substantial concerns with the included studies. Case-control studies have a high risk of bias in the QUADAS-2 ‘patient selection’ domain. Almost all of the studies had a high risk of bias in the ‘flow and timing’ domain, however, this is difficult to avoid when the reference standard for positive test results involves invasive testing, as it is not practical or ethical to undertake such invasive tests in participants with a negative MCED (index) test result. Only one study was undertaken in the UK, although this was in individuals in whom cancer was suspected, so not reflective of the general cancer screening population. Cancer risk and the availability of general population cancer screening programmes differ worldwide, which will impact the applicability of results of the included studies to the UK. Ethnicity and socioeconomic status of included participants was not well reported in the included studies. There were also concerns about the applicability of the CancerSEEK test, which has since been modified (now called Cancerguard™) and is undergoing further assessment. The applicability of the SPOT-MAS, Trucheck, CDA and AICS tests assessed in the included studies was unclear. Outcomes relating to MCED test performance (i.e., test accuracy and number of cancers detected by site and/or stage) were reported in most studies. Overall test sensitivity and specificity are not directly comparable across different MCED tests, owing to the differences in the number of cancer types each test claims to detect. Diagnostic accuracy varied substantially (95% confidence interval [CI] shown in brackets): **Galleri** (3 studies) Sensitivity: 20.8% (14.0 to 29.2) to 66.3% (61.2 to 71.1) Specificity: 98.4% (98.1 to 98.8) to 99.5% (99.0 to 99.8) **CancerSEEK** (2 studies) Sensitivity: 27.1% (18.5 to 37.1) to 62.3% (59.3 to 65.3) Specificity: 98.9% (98.7 to 99.1) to 99.1% (98.5 to 99.8) **SPOT-MAS** (2 studies) Sensitivity: 72.4% (66.3 to 78.0) to 100% (54.1 to 100) Specificity: 97.0% (95.1 to 98.4) to 99.9% (99.6 to 100) **TruCheck** (1 study) Sensitivity: 90.0% (55.5 to 99.7) Specificity: 96.4% (95.9 to 96.8) **CDA** (1 study) Sensitivity: 40.0% (95% CI 12.2 to 73.8) Specificity: 97.6% (95% CI 96.8 to 98.2) **AICS** screens for individual cancers separately; sensitivity ranged from 16.7% (95% CI 3.0 to 56.4) for ovary/uterus cancer to 51.7% (95% CI 34.4 to 68.6) for gastric cancer. Sensitivity by cancer stage was only reported in some studies of Galleri and CancerSEEK. Sensitivity was considerably lower for detecting earlier stage cancers (Stage I-II) compared with later stage cancers (Stage III-IV). Amongst the Galleri studies, sensitivity for detecting Stage I-II cancer ranged from 27.5% (25.3 to 29.8) to 37.3% (29.8 to 45.4) and sensitivity for detecting Stage III-IV cancer ranged from 83.9% (81.7 to 85.9) to 89.7% (84.5 to 93.6). The CancerSEEK cohort study reported sensitivity for detecting Stage I-II cancer of 12.7% (95% CI 6.6 to 23.1) and sensitivity for detecting Stage III-IV cancer of 53.1% (95% CI 36.4 to 69.1). One study of Galleri found that sensitivity was higher in an ‘elevated risk’ cohort (23.4%, 95% CI 14.5 to 34.4) than a ‘non-elevated risk’ cohort (16.3%, 95% CI 6.8 to 30.7). Studies of Galleri, CancerSEEK, SPOT-MAS, CDA and AICS reported sensitivity by cancer site and found that it varied substantially, although the total number of participants diagnosed with certain types of cancer was low, so results are difficult to interpret. <u>Screening programme availability:</u> The sensitivity of the MCED tests to detect solid tumour cancers without a current screening programme in the UK was generally higher than the sensitivity to detect cancers with a current screening programme in the UK (breast, cervical and colorectal). However, this was not the case in one study of Galleri and the study of the CDA test, where sensitivity for detecting solid tumour cancers without a current screening programme was lower than for cancers with a current screening programme in the UK. <u>Subgroup results by participant demographic characteristics:</u> One study each of Galleri and CancerSEEK reported MCED test performance by pre-specified subgroups of interest (i.e., age, sex and ethnicity). For CancerSEEK, sensitivity was slightly lower for participants less than 50 years of age, compared to participants aged 50 or over, while for Galleri sensitivity was very similar across the age categories presented. The sensitivity of Galleri was highest for Hispanic participants (63%), and it was lowest (43%) for the small number of participants classified as ‘Other’ ethnicity in the study. Sensitivity of CancerSEEK ranged from 50% in participants with unknown ethnicities to 70.4% in Asian participants (and cancer was correctly detected by the CancerSEEK test in one Hispanic participant resulting in a sensitivity of 100%). One study using an earlier version of the Galleri test reported results by age and sex for a subset of study participants; cancer signal detection rate was similar in males and females and increased with age for both sexes, however, few details were given on the subset of participants analysed. Only one study of Galleri reported data for participants with a low socioeconomic status. <u>Patient relevant outcomes</u> Only limited results relating to patient relevant outcomes, such as mortality, potential harms, HRQoL, acceptability and satisfaction of individuals screened, were reported in some studies of Galleri, CancerSEEK and AICS.

*Studies of MCED technologies at an unclear stage of development:* The risk of bias assessment identified substantial concerns. Most studies were case-control studies so had a high risk of bias in the ‘patient selection’ domain of QUADAS-2. Most studies also had a high risk of bias in the ‘index test’ and/or ‘flow and timing’ domains. All studies were considered to have high or unclear concerns relating to the applicability of study participants, index tests and reference standard tests. Outcomes relating to MCED test performance were reported in most studies. OncoSeek reported the lowest overall sensitivity across all cancer types (47.4%), and CancerenD24 reported the lowest sensitivity in detecting bladder cancer (38.0%). By stage, OverC and SeekInCare reported a sensitivity of 35.4% and 50.3%, respectively, for stage I cancer. The highest sensitivity overall came from the Carcimun-test (88.8%), however, the exclusion of individuals with inflammation is noted as a disadvantage. The SpecGastro test was only developed to detect three types of gastrointestinal cancer (colorectal, gastric, and oesophageal).

*Stakeholder engagement:* At the protocol stage, stakeholders highlighted issues with the implementation of MCED tests, including resource use, impact on existing diagnostic services and wider care pathways, the need to balance benefits with potential risks, and consideration of factors likely to affect test uptake. Stakeholders also reinforced the importance of patient relevant outcomes. Stakeholders commenting on the draft report noted that important details about the potential benefits, harms, and possible unintended consequences of implementing MCED tests in the UK were poorly reported, limiting the relevance of the available evidence for policy decision-making. Other feedback fell into six broad areas: poor applicability and generalisability of available evidence; limitations of the current evidence base; the potential impact of MCED tests on existing screening, diagnostic and treatment pathways; opportunities to enhance services to improve outcomes; acceptability and potential impact on populations offered and/or receiving screening, and; targeting specific groups to support early identification and improve outcomes.

*Conclusions:* Limited evidence is available on the potential for early detection of treatable cancers, and the consequences of introducing screening with an MCED test in a UK population. There were no completed RCTs identified for any of the MCED tests and most included studies had a high overall risk of bias, primarily owing to limited follow-up of participants with negative test results. There were concerns about the applicability of the participants in most studies. Only one study of Galleri recruited asymptomatic individuals aged over 50 years but it was conducted in the USA, therefore, study participants and results may not be representative of a UK screening population. All currently available MCED tests (Galleri, CancerSEEK, SPOT-MAS, TruCheck, CDA and AICS) reported high specificity (>96%) which is essential if an MCED test is to correctly classify people who do not have cancer. Sensitivity was variable and influenced by study design, population, reference standard test used and length of follow-up. Sensitivity also varied by cancer stage; where reported, MCED tests had considerably lower sensitivity to detect earlier stage cancers (Stage I-II). Sensitivity also appeared to vary substantially for different cancer sites, although results are limited by small patient numbers for some cancer sites. The sensitivity of most of the MCED tests to detect solid tumour cancers without a current screening programme in the UK was higher than their sensitivity to detect cancers with a screening programme in the UK (breast, cervical and colorectal). Where reported, differences in test accuracy by age and sex were small. Whilst some differences were observed by ethnicity, these results should be interpreted with caution as the majority of participants recruited to studies were White and numbers of participants from other ethnic groups were small. Evidence on seven MCED technologies which were at an unclear stage of development and did not appear to be available for use were briefly summarised; most were evaluated in case-control studies and had a high risk of bias, all studies had high or unclear applicability concerns. No meaningful results were reported relating to patient relevant outcomes, such as mortality, potential harms, HRQoL, acceptability or satisfaction.

*Recommendations for research:* RCTs with sufficiently long follow-up, reporting outcomes that are directly relevant to patients, such as mortality/morbidity, safety, and HRQoL, are needed and some are planned or underway. Research is also needed on the resource implications of MCED tests on NHS services, risk of over-treatment and cost-effectiveness of implementing MCED tests for screening in the UK.

*Study registration:* This study is registered as PROSPERO CRD42023467901.

*Funding details:* This project was funded by the National Institute for Health and Care Research (NIHR) Evidence Synthesis programme (NIHR161758) and will be published in full in the Health Technology Assessment Journal; Vol. XX, No. XX. See the NIHR Journals Library website for further project information.

## 1 BACKGROUND

Population-based cancer screening in the UK national health service (NHS) is currently limited to selected cancers (cervical, breast, bowel).^1^ Additionally, in some areas of England and Wales individuals at high risk of developing lung cancer can receive a lung health check.^2^ Most other cancers are detected after presentation of symptoms, many of which will be diagnosed at stages III and IV, where treatment options may be more limited. Breast, prostate, lung, and bowel cancers together account for just over half of all new cancers diagnosed.^3^

The Galleri® test (GRAIL, Menlo Park, California) is a multi-cancer early detection (MCED) blood test that uses genetic sequencing to detect potential signals of cancer and is currently recommended by the manufacturer for use in adults with an elevated risk of cancer, such as those aged 50 years or older.^4^ The assay is combined with a machine-learning-based classification algorithm that identifies patterns predictive of cancer and indicative of potential cancer site of origin. The test detects circulating cell-free deoxyribonucleic acid (DNA) (cfDNA) and is able to predict the most likely site, or sites, within the body that the signal is coming from (the ‘Cancer Signal Origin’), allowing for confirmatory follow-up tests. Galleri predicts up to two Cancer Signal Origins (CSO) by comparing the methylation pattern to the patterns of 21 possible CSO predictions. Predicting the origin of the cancer signal helps healthcare providers select the appropriate follow-up diagnostic tests. The CSO can be either an anatomic site (e.g., colorectal) or a cellular lineage (e.g., lymphoid).^5^

Another blood-based MCED test which detects cfDNA and protein biomarkers (such as cancer antigen 125, CA125) is CancerSEEK (Exact Sciences, Madison, Wisconsin).^6^ MCED tests based on detecting other cancer-related biomarkers in the blood are also available.^7^ For example, SPOT-MAS™ (Gene Solutions, Ho Chi Minh City, Vietnam) detects circulating tumour DNA (ctDNA) – a type of cfDNA – and applies machine learning algorithms to detect five types of cancer.^8^ TruCheck™ (Datar Cancer Genetics, Beyreuth, Germany) detects the presence of circulating tumour cells (CTCs) and their clusters, which are causatively associated with malignant tumours and are rare among asymptomatic populations.^9^ CDA (Cancer Differentiation Analysis; AnPac Bio, Shanghai, China) detects and analyses electrical biophysical signatures in whole blood samples and generates a CDA value (with higher values indicating higher cancer risk), rather than focusing on specific cells.^10^ The AICS® test (AminoIndex Cancer Screening; Ajinomoto, Tokyo, Japan) uses plasma-free amino acid profiles as biomarkers for six different types of cancer, but rather than giving an overall prediction, the test ranks participants on the probability of having each of the cancers tested (grouped into A, B, or C, with C as the high-risk group).^11^ A recent review summarised these different MCED technologies and provides an overview of the type of biomarkers (e.g., cfDNA, CTC, protein or metabolites) that can be used to differentiate a variety of cancers.^12^

The NHS Long Term Plan ambition seeks to diagnose 75% of cancers at stage I or II, to enable more effective treatment.^13^ An MCED test embedded within a national population-based screening programme, in addition to existing cancer screening programmes, may increase the number of cancers diagnosed at an earlier stage, potentially improving the likelihood of treatment success and consequent survival rates. However, the identification of cancers with no effective treatments even at an early stage may have no improvement on mortality or health-related quality of life (HRQoL). It is also unclear whether detecting some cancers earlier impacts cancer-specific mortality since they might still have been detected and successfully treated using existing screening and referral pathways, without MCED testing.^14^

In addition, early screening of healthy people for such a wide range of cancers, and the expected lengthy time to diagnostic resolution, may create anxiety and lead to unnecessary follow-up tests, when false positives occur.^15, 16^ The potential for overdiagnosis of cancers at such an early stage that they might never have advanced enough to require treatment, may also lead to unintended harms.^17^ Communication of a negative MCED test result might also lead to false reassurance and reduce uptake to other existing screening programmes or lead to delays in individuals presenting to their GP with symptoms, even though it is recommended that regular screening is continued regardless of MCED test result.^18^

The aim of this project was to assess the accuracy and clinical effectiveness, acceptability and feasibility of blood-based MCED tests for population-based screening of individuals aged 50 to 79 years without clinical suspicion of cancer and who have not been diagnosed with cancer or received treatment for cancer within the last three years.

The objective was to conduct a systematic literature review of the clinical effectiveness evidence on blood-based MCED tests for screening.

## 2 METHODS

The systematic review was conducted following the general principles recommended in the Centre for Reviews and Dissemination’s guidance and reported in accordance with the Preferred Reporting Items for Systematic Reviews and Meta-Analyses (PRISMA) statement (Appendix 1, Table 9 and Table 10).^19, 20^ The systematic review was registered with PROSPERO, registration number CRD42023467901.

### 2.1 Inclusion criteria

#### 2.1.1 Population

The target population was individuals aged 50 to 79 years without clinical suspicion of cancer and who had not been diagnosed with cancer or received treatment for cancer within the last three years. As insufficient relevant studies were identified within the target population, studies including patients known to have cancer (i.e., case-control studies) and studies that included individuals with a wider age range than 50 to 79 years were considered for inclusion.

Subgroups of interest were individuals at elevated risk of cancer (e.g., smoking history, genetic predisposition or personal history of malignancy), and patients diagnosed with different cancer types (i.e., primary site) and at different cancer stages, where diagnostic accuracy may differ. Where possible, we also planned to examine differences in demographic characteristics such as age and sex, as well as potentially important characteristics associated with health inequalities, such as ethnic group and socio-economic status.

#### 2.1.2 Interventions

This review included blood-based MCED tests for cancer screening, where these tests aim to detect multiple types of cancer. Studies assessing blood-based tests for assessing prognosis (e.g., risk-stratification, tumour staging and genotyping) or therapeutic decision-making (e.g., guiding precision therapy or monitoring response to treatment) in patients known to have cancer were not eligible for inclusion.

Technologies are also being developed to detect cancer signals in other bodily fluids, such as urine.^12^ However, such technologies are in a much earlier stage of development than blood-based tests so we only focused on blood-based MCED tests in this review.

#### 2.1.3 Comparators

The comparator was no MCED test but individuals should still be offered relevant existing screening programmes and clinical follow-up of symptoms. Uncontrolled studies were also eligible for inclusion if relevant outcome data were provided.

#### 2.1.4 Outcomes

Outcomes of interest related to test performance were

- accuracy of the test; including sensitivity, specificity, positive and negative predictive values, and reference standard test used to determine true disease status, if any
- accuracy of the CSO
- number and proportion of cancers detected (by site and stage), including the proportion of cancers targeted by the test which were detected

Patient-relevant outcomes of interest were:

- mortality (all-cause and disease-specific);
- time to diagnosis (or exclusion) of cancer;
- incidental findings;
- additional tests and procedures;
- potential harms;
- HRQoL;
- acceptability to individuals screened
- satisfaction of individuals screened.

#### 2.1.5 Study designs

Prospective clinical trials (including randomised and other controlled trials) and cohort studies were sought. As insufficient relevant trials and prospective cohort studies were identified, we included case-control studies, including patients known to have cancer, if relevant outcome data were reported.

For case-control studies, only the following outcomes were relevant: accuracy of the test (sensitivity and specificity); accuracy of the CSO; number of cancers detected (by site and stage); acceptability to individuals tested; and satisfaction of individuals tested.

Early development studies (e.g., pre-clinical studies using biobank samples, studies training, evaluating or refining algorithms) which did not recruit participants with the aim of assessing diagnostic accuracy or clinical effectiveness of the tests were not eligible for inclusion.

### 2.2 Search strategy for identification of studies

The aim of the search was to systematically identify published and unpublished studies of MCED tests used for the purposes of population screening. Comprehensive searches of electronic databases, trial registers, examination of relevant websites and reference checking of included studies and systematic reviews was undertaken.

A search strategy was designed in Ovid MEDLINE by an Information Specialist (MH) in consultation with the review team. The strategy combined terms for multiple cancer, terms for liquid biopsy or blood tests, and terms for screening or early detection. Searches of the title and abstract fields of database records along with relevant subject headings were included in the strategy. Specific phrases for the tests such as “multi-cancer early detection tests” were also included in the strategy as well as the brand names of individual tests (e.g., Galleri, PanSEER, and CancerSEEK). The search was limited to records published from 2010 onwards to reflect the recent development of these technologies. No further limits were applied. The MEDLINE strategy was peer reviewed by a second Information Specialist and any necessary adjustments or corrections made. The strategy was then adapted for use in all other databases and resources searched.

The following databases were searched in September 2023: MEDLINE ALL (Ovid), Embase (Ovid), Cochrane Central Register of Controlled Trials (CENTRAL, Wiley), the Science Citation Index (Web of Science), Cochrane Database of Systematic Reviews (CDSR, Wiley), Database of Abstract of Reviews of Effects (DARE), and KSR Evidence (Ovid).

Unpublished, ongoing or grey literature was identified through searching the health technology assessment (HTA) database, International HTA database (INAHTA), websites of international HTA organisations, Conference Proceedings Citation index – Science (Web of Science), ClinicalTrials.gov, WHO International Clinical Trials Registry portal, and PROSPERO. All search results were imported into EndNote 20 reference management software and deduplicated.

After screening records identified by electronic searches, manufacturers of MCED tests were identified from the included studies and their websites examined to identify further references published from 2020 onwards. The reference lists of included studies and relevant systematic reviews were also checked for any relevant references.

The full search strategies can be found in Appendix 2.

### 2.3 Study selection

All references identified by the electronic searches were uploaded into EPPI-Reviewer. The machine learning and text mining tool in EPPI-Reviewer (priority screening) was used to prioritise titles and abstracts for screening.^21^ All titles and abstracts were assessed by one reviewer (CK, GR, RW, SD, SN or YL) with the first 10% of prioritised records assessed by two reviewers to ensure consistency; disagreements were resolved by discussion or, if necessary, a third independent reviewer. The full texts of potentially eligible studies were assessed independently by two reviewers (CK, GR, RW, SD, SN or YL), using the same process for resolution of disagreements as outlined above. Studies published as pre-prints or conference abstracts reporting relevant outcome data were eligible for inclusion. Foreign-language publications were eligible for inclusion and translated for data extraction, if applicable. Eligible ongoing studies (e.g., reported in protocols and trial registers) without relevant outcome data reported at the time of data extraction were included.

### 2.4 Data extraction

A standardised data extraction form was developed and piloted. Data on the intervention(s), patient characteristics, setting, study design, reference standard test(s) used, and relevant outcomes were extracted from included studies by one reviewer (RW, SN or YL) and independently checked by a second reviewer (CK, GR, RW, SN or YL). Disagreements were resolved through discussion.

The following values were extracted or calculated for each MCED test:

- the number of true positives (TP) which is the number of people with a positive cancer signal (i.e., a positive result of the MCED test) who do have cancer, i.e., the number of people correctly identified by the MCED test as having cancer
- the number of false positives (FP) which is the number of people with a positive cancer signal who do not have cancer, i.e., the number of people incorrectly identified by the MCED test as having cancer
- the number of true negatives (TN) which is the number of people with a negative cancer signal (i.e., a negative result of the MCED test) who do not have cancer, i.e., the number of people correctly identified by the MCED test as not having cancer
- the number of false negatives (FN) which is the number of people with a negative cancer signal who do have cancer, i.e., the number of people incorrectly identified by the MCED test as not having cancer

Measures of test accuracy are as follows:

- Sensitivity = TP / (TP + FN) This is the true positive rate which is the probability that an individual with cancer receives a positive MCED test result; in other words, the ability of a test to correctly classify a person with cancer.
- Specificity = TN / (TN + FP) This is the true negative rate which is the probability that an individual without cancer receives a negative MCED test result; in other words, the ability of a test to correctly classify a person without cancer.
- Positive predictive value (PPV) = TP / (TP + FP) This is the probability that a person who receives a positive MCED test result has cancer.
- Negative predictive value (NPV) = TN / (TN + FN) This is the probability that a person who receives a negative MCED test result does not have cancer.

These test accuracy measures were extracted or, where not directly reported, calculated from other reported data using package epiR^22^ in RStudio version 4.3.1,^23^ where appropriate. Calculated measures, as opposed to directly reported measures, are identified as such in all results tables. Sensitivity was also extracted or calculated where possible by cancer site and stage. Specificity was not presented by site and stage as it is important that a test correctly classifies that a person does not have cancer of any type or stage.

### 2.5 Critical appraisal

Risk of bias and applicability were assessed using the QUADAS-2 (Quality Assessment of Diagnostic Accuracy Studies) checklist^24^ by one reviewer (RW or YL) and independently checked by a second reviewer (CK, GR, RW or YL). Disagreements were resolved through discussion.

### 2.6 Data synthesis and investigation of heterogeneity

Data were not suitable for pooling in a meta-analysis due to the difference in study designs, populations, and interventions. The results of data extraction are presented in a series of structured tables grouped by MCED test and visualised using the R package “ggplot2”^25^ where appropriate. Narrative summaries of differences in study designs, populations and MCED tests as well as narrative summaries of MCED test performance and patient relevant outcomes by MCED test and across MCED tests are presented, where appropriate.

Certainty in the body of evidence was considered in terms of the study design (e.g., cohort vs case-control), the type of reference standard test used, the extent and length of follow-up for true and false negatives, and the relevance of the population (e.g., asymptomatic vs symptomatic population).

#### 2.6.1 Subgroup analyses

A narrative summary of results relevant to subgroups of interest (individuals at elevated risk of cancer, diagnosed with different cancer types, age groups, ethnic group and sex) and differences in accuracy by CSO is presented, where available.

Results by cancer types with and without current screening in the UK are also presented. Although this was not a pre-specified subgroup of interest in the protocol,^26^ stakeholders commented that this was a useful summary of the available evidence.

### 2.7 Stakeholder involvement

We ensured that relevant perspectives were properly considered during protocol development and as part of the process of understanding, interpreting and contextualising the findings of this review. In developing the protocol, we worked with a range of content experts involved in the cancer screening and care pathway, including general practitioners and cancer screening and diagnostic research and implementation experts, as well as representatives from the UK National Screening Committee. We also worked with the manager at Healthwatch York^27^ to ensure that issues raised by patients and public communities were considered at an early stage.

Upon completion of the review, a draft copy of the final report was shared with a selected group of stakeholders (as outlined in the Acknowledgements). Comments and feedback from these stakeholders were incorporated into the final draft of the report.

Several further consultation exercises were then undertaken to explore the broader views of patients and the public about the use of MCEDs as part of a general population screening programme. These open discussions (one group involving 11 participants, and two individual consultations with separate informants) took place remotely via Zoom, to maximise opportunities for involvement, and lasted between 1 and 1.5 hours. The group discussion was led by one of the co-authors (RC) and our Healthwatch York partner, whilst the individual consultations were undertaken by our Healthwatch partner alone. At the start of each discussion, participants were given some brief context about MCEDs and an outline of the purpose of the session (based on the information provided in the invitation – see Appendix 3). A brief verbal description of the review undertaken, based on the Plain Language Summary, was also provided. With the support of several organisations, including Healthwatch York, the TRANSFORM platform and Involve Hull,^28^ the Humber and North Yorkshire Cancer Alliance,^29^ we were able to involve people from across the UK with lived experience of a cancer diagnosis, carers, as well as people who would meet the inclusion criteria for general population screening using MCEDs in this review.

## 3 RESULTS

### 3.1 Studies included in the review

The electronic searches identified a total of 8,069 records after deduplication between databases. The full texts of 228 records were ordered for further review; 176 were excluded at full paper stage and are listed in Appendix 4, along with the reasons for their exclusion. No additional records were identified from screening reference lists of included studies and relevant systematic reviews. Eleven additional records were identified from searching MCED test manufacturer websites.

Sixty-three records reporting results from 36 individual studies, evaluating 13 tests or technologies met the review inclusion criteria. There was a considerable amount of duplicate reporting in e.g., multiple conference abstracts and posters, in addition to the main journal article describing a study. One ongoing randomised controlled trial (RCT), 13 completed cohort studies, 17 completed case-control studies, four ongoing cohort studies and one ongoing case-control study were included. Figure 1 presents the flow of records through the selection process.

**Figure 1.**
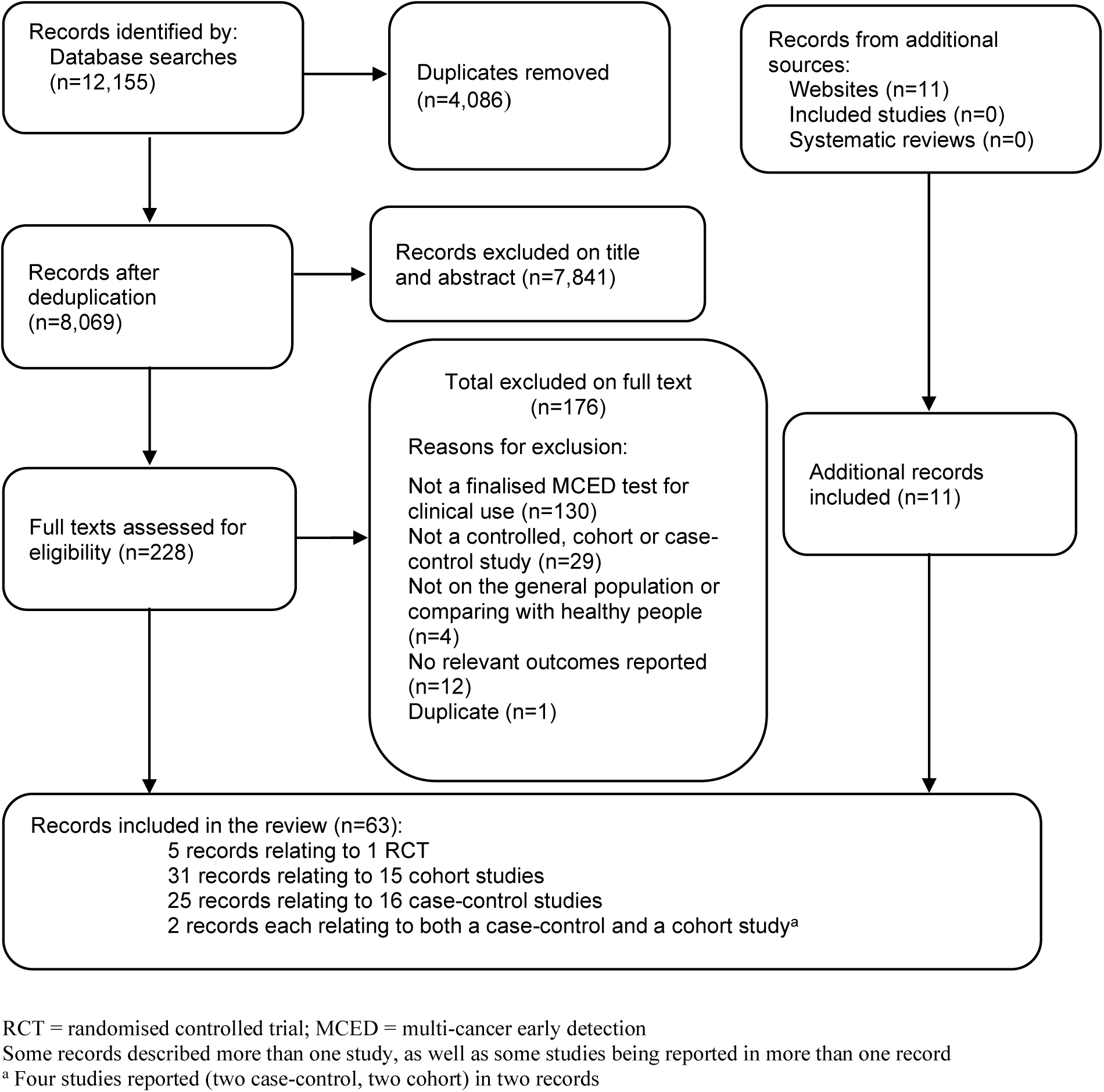
Flow diagram of the study selection process.

Study selection was complex. In particular, it was often difficult at full-text screening to determine whether studies were reporting results for technologies at an early stage of development, or whether studies were assessing the final or near-final version of the test.

Where known, we only included studies that appeared to assess tests that were in the final stages of development, e.g., for the GRAIL MCED test, we only included studies that assessed the “refined MCED test” (Galleri), i.e., the PATHFINDER study^30^ and the Circulating Cell-free Genome Atlas (CCGA) substudy 3,^31^ but not CCGA sub studies 1 and 2 that assessed an earlier version of the test.^32^ For CancerSEEK, completed studies assessed what appears to be an earlier version of the test.^6, 33^ A modified version of the test, now called Cancerguard™ is undergoing further assessment but no completed eligible studies were found.^34^ Studies were also included reporting data on SPOT-MAS,^35^ Trucheck,^36^ CDA,^37^ and AICS^38^ which are blood-based MCED tests currently available for use, according to the manufacturer’s websites.

Studies of other MCED technologies at an unclear stage of development, which do not seem to be available for use at the date of submission of this report, were also included in this review: Aristotle® (StageZero Life Sciences, Richmond, Ontario), CancerenD24 (manufacturer unknown), OncoSeek® (SeekIn Inc, San Diego, California), SeekInCare® (SeekIn Inc, San Diego, California), OverC™ (Burning Rock Biotech, Guangzhou, China), Carcimun-test (Carcimun Biotech, Garmisch-Partenkirchen, Germany) and SpecGastro test (manufacturer unknown).

Technologies that appeared to be at a very early stage of development and, therefore, not meeting the inclusion criteria for the review are described in Section 3.6.

The main references for completed and ongoing studies included for each test or technology are summarised in Table 1, along with the number of cancer types detected or targeted by each test. Additional records reporting supplementary information for studies in Table 2,^39-57^ are detailed in Appendix 5, Table 11.

**Table 1.**
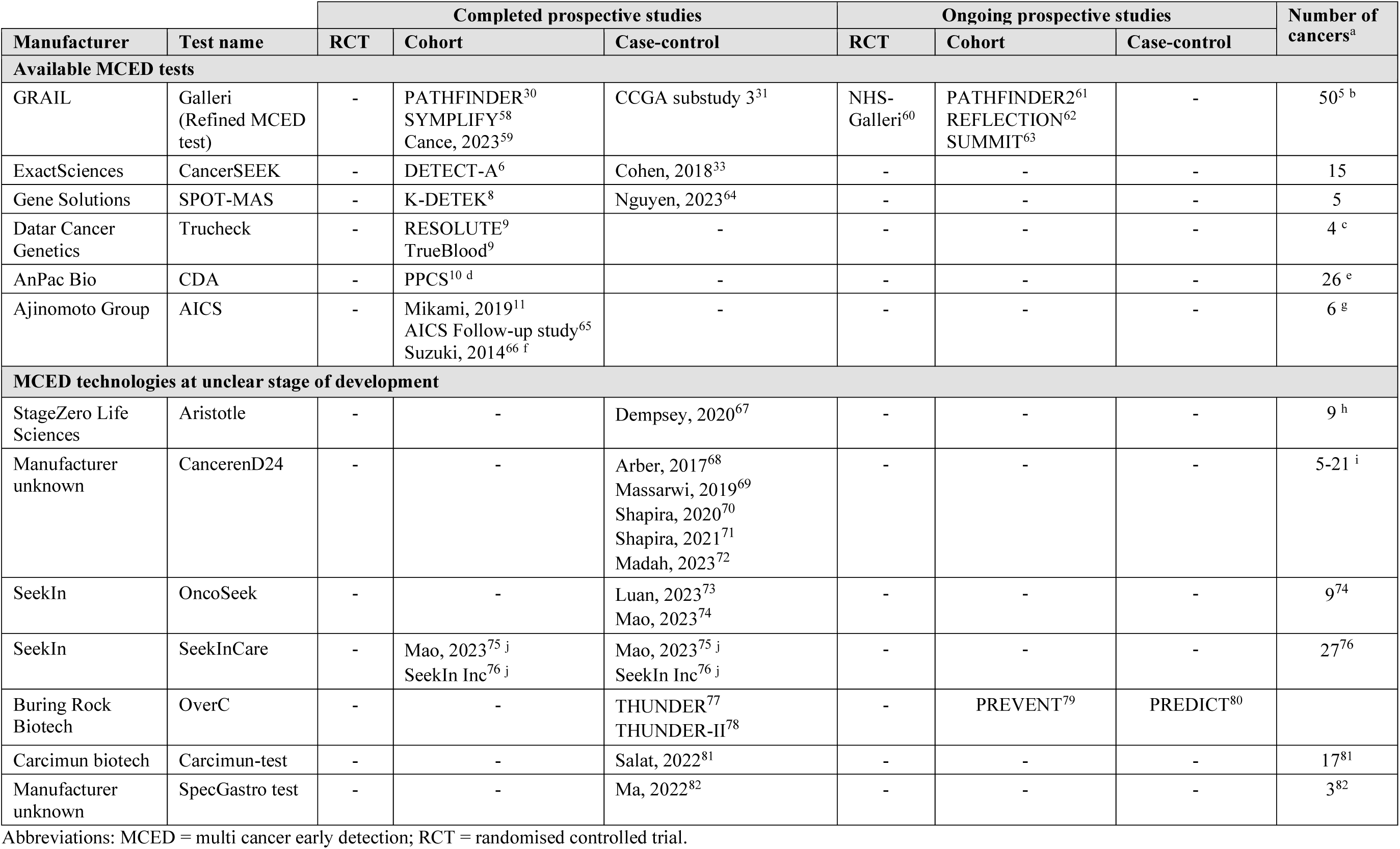
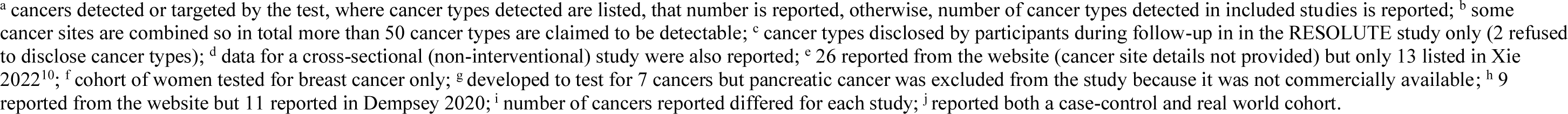
Completed and ongoing studies available for each test and number of cancers detected or targeted by each test.

**Table 2.**
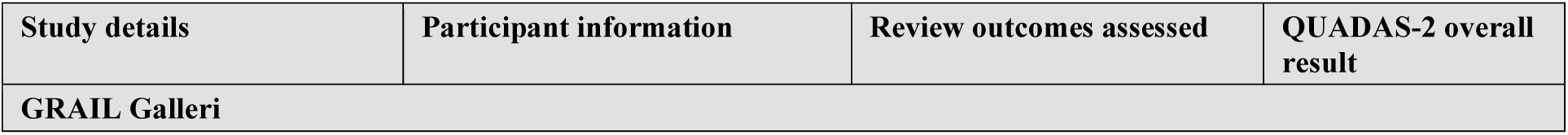
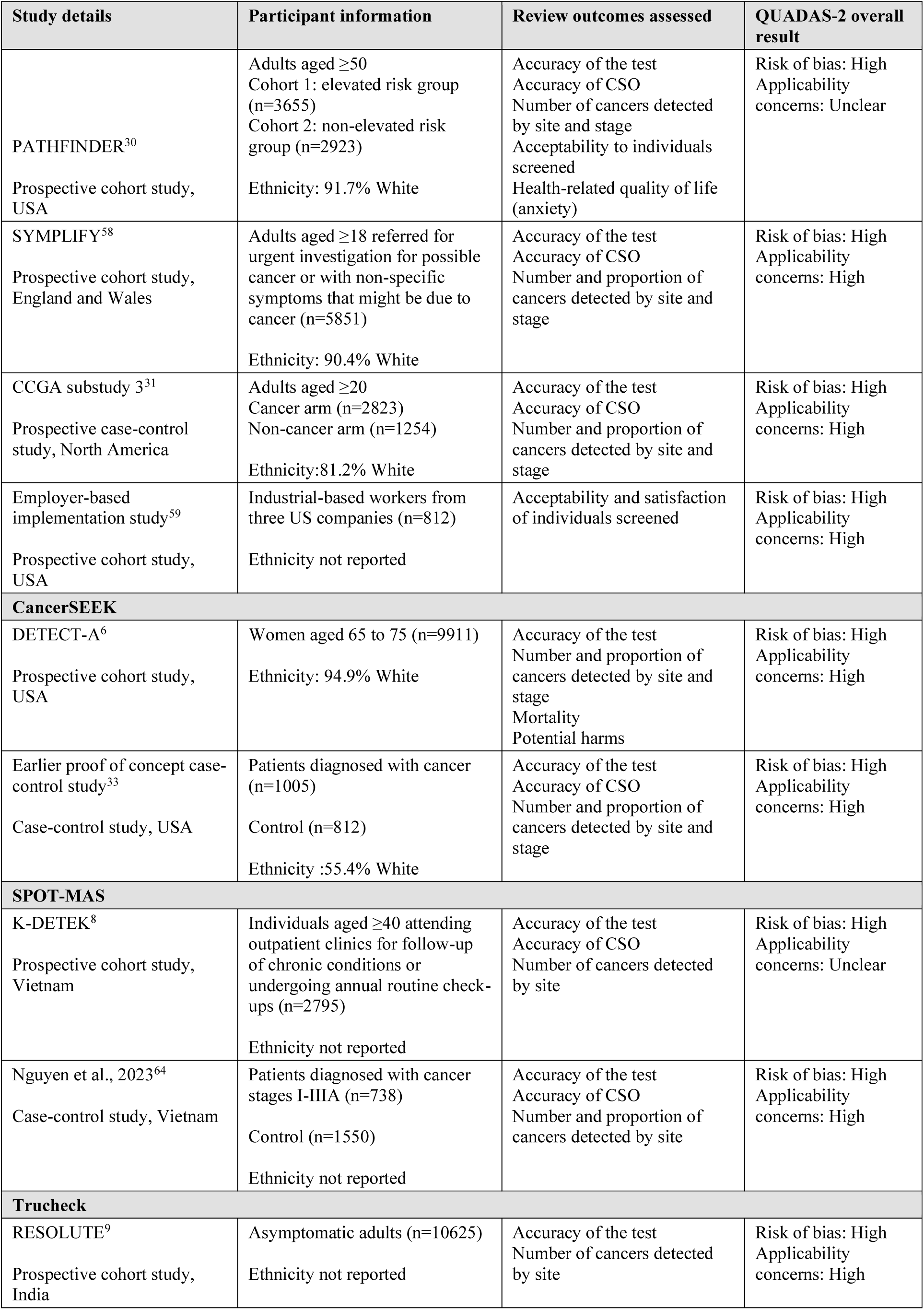
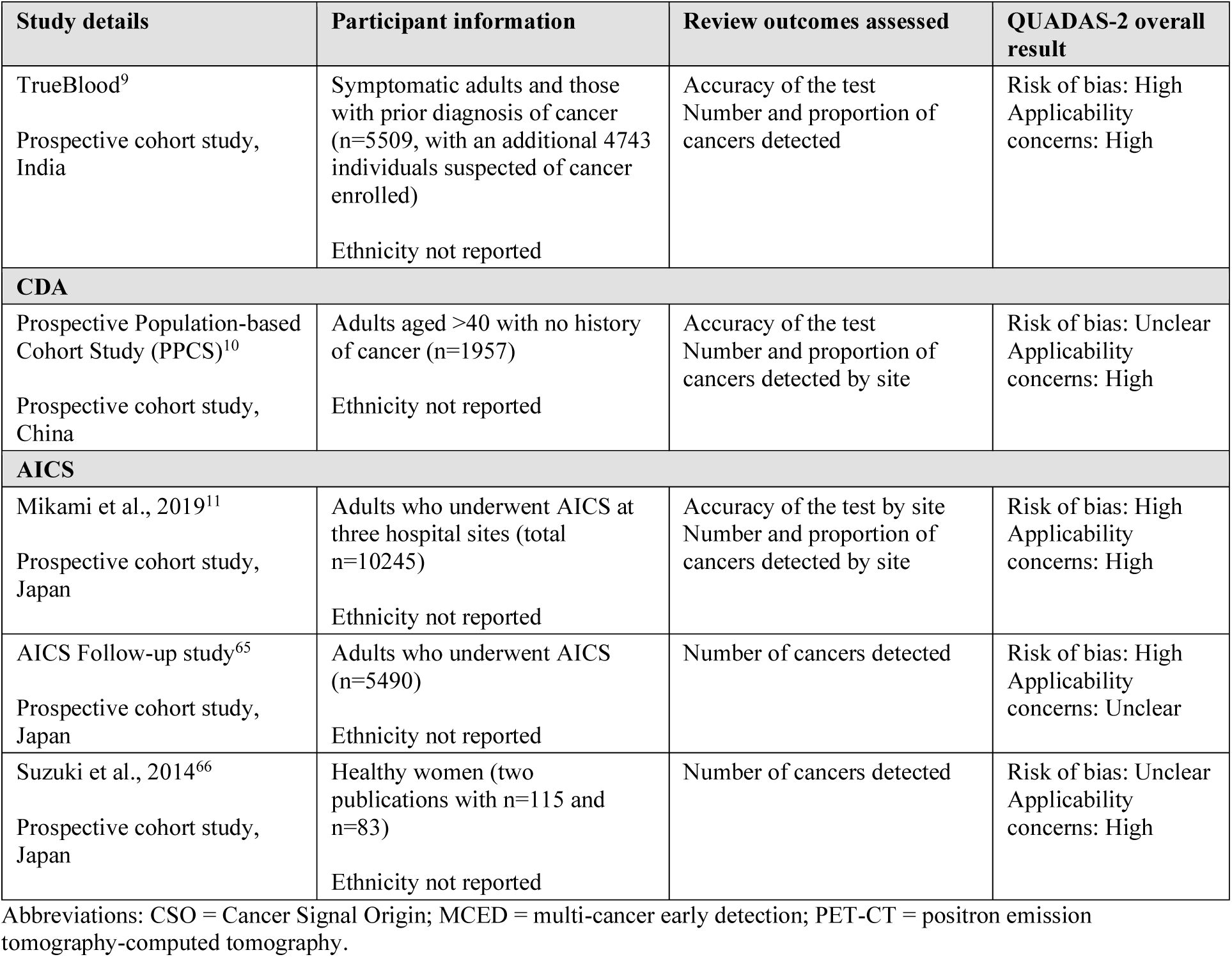
Summary of the included studies for each MCED test.

### 3.2 Characteristics of the included studies

Table 2 summarises study characteristics for each of the MCED tests currently available for use. Further details of each study and technology are presented in Appendix 5, Table 11.

There were no completed prospective RCTs identified for any of the MCED tests. All studies were either prospective cohort studies or case-control studies. Only one study (SYMPLIFY^58^) was undertaken in the UK (sites in England and Wales), although this was in individuals in whom cancer was suspected. Cancer risk and the availability of general population cancer screening programmes differ worldwide, which will impact the applicability of results of the included studies to the UK. Most of the prospective cohort studies and control groups in all case control studies recruited participants without any known history of cancer. The ‘elevated-risk’ cohort of the PATHFINDER study included 1622 participants (41%) who had a history of invasive or haematological malignancy with treatment completed >3 years prior to enrolment (Appendix 5, Table 11). Participants with a cancer history with treatment completed >3 years prior were also eligible for enrolment into the SYMPLIFY^58^ study, but the number of recruited participants with a history of cancer was not reported.

Ethnicity of included participants was not well reported across the included studies and socioeconomic status was reported in only one study of Galleri that recruited individuals with low socioeconomic status. Only the three Galleri studies,^30, 31, 58^ and the two CancerSEEK studies^6, 33^ reported on participants’ ethnic backgrounds (Appendix 5, Table 11). The majority of participants included in these studies were from White Caucasian background (81.2% to 91.7% in three studies of Galleri^30, 31^ and 55.4% to 94.9% in the two studies of CancerSEEK).^6, 33^ The case-control CancerSEEK study further included 17.8% of participants of Asian ethnicity,^33^ compared with DETECT-A which only included 0.4%,^6^ and the Galleri studies (PATHFINDER: 1.9%, SYMPLIFY: 4.2% [including South Asian and Chinese], CCGA substudy 3: 1.8%).^30, 31, 58^

Outcomes relating to the MCED test performance (i.e., accuracy of the test, accuracy of CSO and number of cancers detected by site and/or stage) were reported in most studies. Very limited patient relevant outcomes, such as mortality, potential harms (e.g., relating to adverse effects of additional tests and procedures undertaken), HRQoL (e.g., anxiety), acceptability and satisfaction of individuals screened were reported only in studies of Galleri and CancerSEEK.

MCED technologies that appear to be at an earlier stage of development and for which it is unclear whether the finalised test version is being evaluated, or if they may still undergo further modification (i.e., Aristotle, CancerenD24, OncoSeek, SeekInCare, OverC, Carcimun-test and SpecGastro test), are presented in Section 3.4.3 with study characteristics presented in Appendix 6, Table 12.

### 3.3 Quality of the included studies

#### 3.1.1 Available MCED tests

An overall summary of QUADAS-2 assessments of the studies of MCED tests currently available for use is presented in Figure 2 (using the R packages “robvis”^83^ and “ggplot2”^25^). The risk of bias assessment identified substantial concerns with the included studies. Patient selection and the reference standard were poorly reported in most of the studies, resulting in an ‘unclear’ risk of bias and/or applicability judgement. Almost all the included studies had a high risk of bias in the ‘flow and timing’ domain of QUADAS-2. However, this is difficult to avoid in studies where the reference standard for positive test results involves invasive testing, as it would not be practical or ethical to undertake such invasive tests in participants with a negative MCED (index) test result.

There was a high applicability concern relating to the ‘patient selection’ domain of QUADAS-2 for several studies as the included participants did not reflect the target population of interest for this review. The index test was also poorly reported across several studies, resulting in an “unclear” applicability concern in this domain.

**Figure 2.**
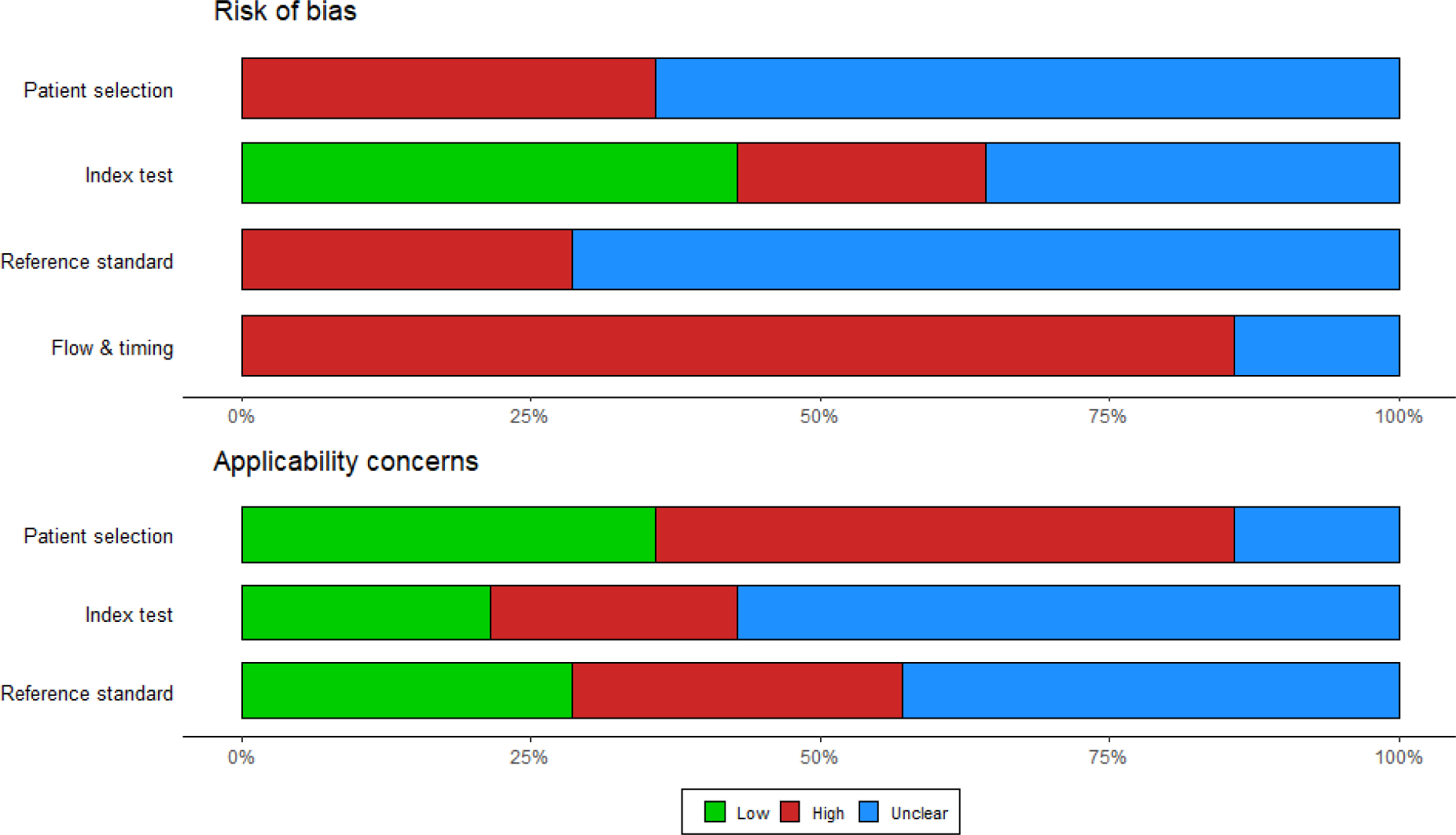
QUADAS-2 overall summary.

QUADAS-2 assessments for each study of MCED tests currently available for use are summarised in Table 3. One study of each of Galleri,^31^ CancerSEEK,^33^ and SPOT-MAS^64^ were case-control studies, which are considered to have a high risk of bias in the ‘patient selection’ domain of the QUADAS-2 checklist.^24^ There was a high concern regarding the applicability of the index test for the studies evaluating CancerSEEK, as this test has been modified (now called Cancerguard™) and is undergoing further assessment.^34^

**Table 3.**
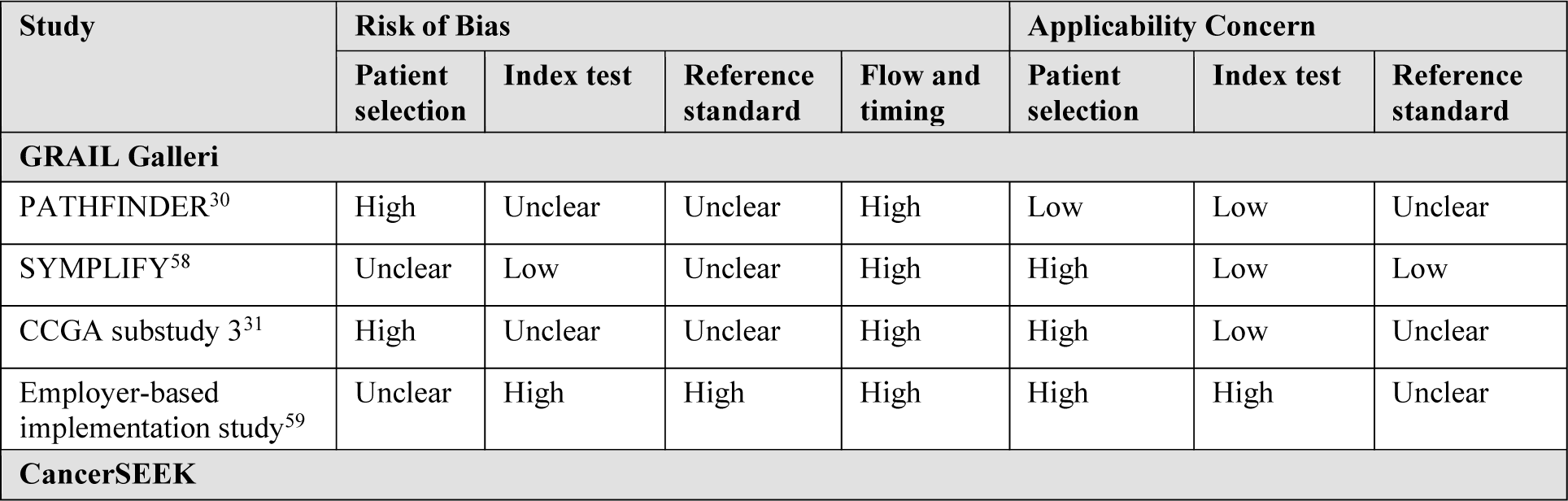
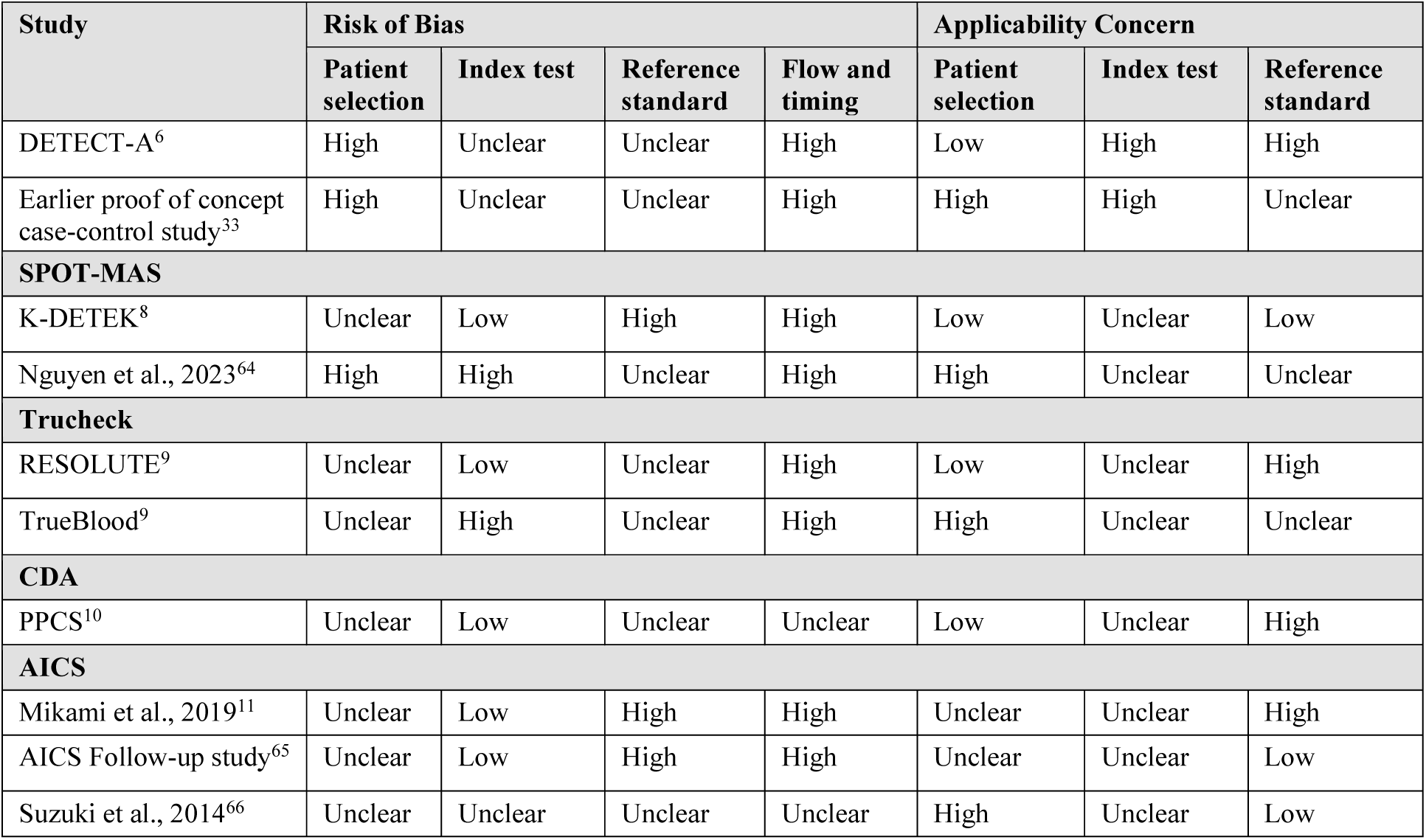
QUADAS-2 assessment results for studies of each MCED test.

#### 3.1.2 MCED technologies at an unclear stage of development

The QUADAS-2 assessment of the studies of MCED technologies at an unclear stage of development is summarised in Appendix 6, Table 13. All of the studies had a high risk of bias and/or applicability concerns; most were case-control studies, and there were also concerns regarding whether the index test was the finalised version of the MCED test, as well as the lack of follow-up reported for healthy controls in some studies.

### 3.4 Outcomes reported in the included studies

No completed RCTs were found for any of the included MCED tests. The Galleri test has an RCT ongoing (see Section 3.5), which plans to report interim results at one year of follow-up. However, no data are currently available for this RCT, so data were extracted from prospective cohort and case-control studies evaluating the refined GRAIL MCED test (Galleri). For other MCED tests, no planned RCTs were identified and only data from prospective cohort and case-control studies were extracted, where available (see Table 1).

Due to the substantial differences in the number of cancers detected by the included tests, study design and populations, statistical pooling of results was not considered appropriate. Results for all MCED tests are presented within tables, described, and compared, where appropriate.

Relevance, validity and comparability of the different outcomes depends upon study design, whether a reference standard test was used to diagnose true disease status when a cancer signal was detected (i.e., positive MCED test result returned), and which additional tests, if any, were conducted on study participants without a cancer signal detected (negative MCED test result returned), to determine true disease status. The most appropriate reference standard test, or combination of tests, is also dependent on the cancer site being investigated and these may differ in their diagnostic accuracy. In addition, overall test sensitivity and specificity are not directly comparable across different MCED tests as the number of cancers each test claims to detect are different (see Table 1, and Appendix 7, Table 14).

Another key issue is that accurate classification of true and false negatives will depend on the extent and length of follow-up in prospective studies. A short follow-up will result in estimates of sensitivity that are higher than they would be if a perfect reference standard test was used to rule out cancer in all study participants with a negative test result. Sensitivity of the MCED test will therefore be subject to bias when only participants with a positive MCED test result undergo further diagnostic investigations during the study. In such cases, FN test results might be missed, unless detected at future routine screening, or clinical investigation after presentation with symptoms, which may not occur for all participants during the follow-up period of the study. In other words, negative results of an MCED test may incorrectly be assumed to be ‘true’ negative results, due to a lack of further testing and a short follow-up time.

Additionally, within studies that include patients known to have cancer (i.e., case-control studies) or who have been referred due to a suspicion of cancer (e.g., symptomatic individuals), estimates of sensitivity will be higher than they would be for the target population of asymptomatic individuals. Estimates of PPV and NPV in these studies will not be reflective of an asymptomatic screening context and are therefore not directly relevant to our target population.

Comparability and relevance of test accuracy measures collected from different studies will therefore be dependent on:

- Which reference standard test (or combination of tests) was used to diagnose cancer in individuals with a positive MCED test result, and the accuracy of that reference standard test (classification of TP and FP).
- Whether further investigations, beyond follow-up, were carried out to rule out cancer in individuals with a negative MCED test result (classification of TN and FN).
- Whether length of follow-up of individuals with a negative MCED test result would be sufficient to detect cancers present at the time of MCED test (classification of TN and FN).
- The prevalence of cancer in the study population in relation to the target population for screening (interpretation of PPV and NPV).
- The location of the studies affects the generalisability of results to UK clinical practice for most of the technologies. Differences in participants’ ethnicity, cancer risk factors, and characteristics of the healthcare system (including existing screening programmes and referral pathways) can impact the prevalence of different cancers.

These issues should be kept in mind when interpreting and comparing the results presented in this section.

#### 3.4.1 Test performance in the included studies

Accuracy of the test and accuracy of the CSO of the Galleri, CancerSEEK, SPOT-MAS, TruCheck and CDA tests is presented in Table 4. Test performance for AICS is not included in Table 4, as each cancer is tested for separately, so no overall results are available. Accuracy of first or second CSO was measured only in the PATHFINDER^30^ and SYMPLIFY^58^ cohort studies of Galleri and in the Cohen 2018^33^ case-control study of CancerSEEK. Where measured, other studies only assessed the accuracy of the first CSO.

Number of cancers detected by the MCED tests by stage is reported in Table 5. Cancer stage was reported for Galleri and CancerSEEK only and total cancers detected by cancer stage were not reported in the PATHFINDER study for the refined MCED test (Galleri). Sensitivity of Galleri and CancerSEEK by cancer stage, where reported, is shown in Figure 3.

**Figure 3.**
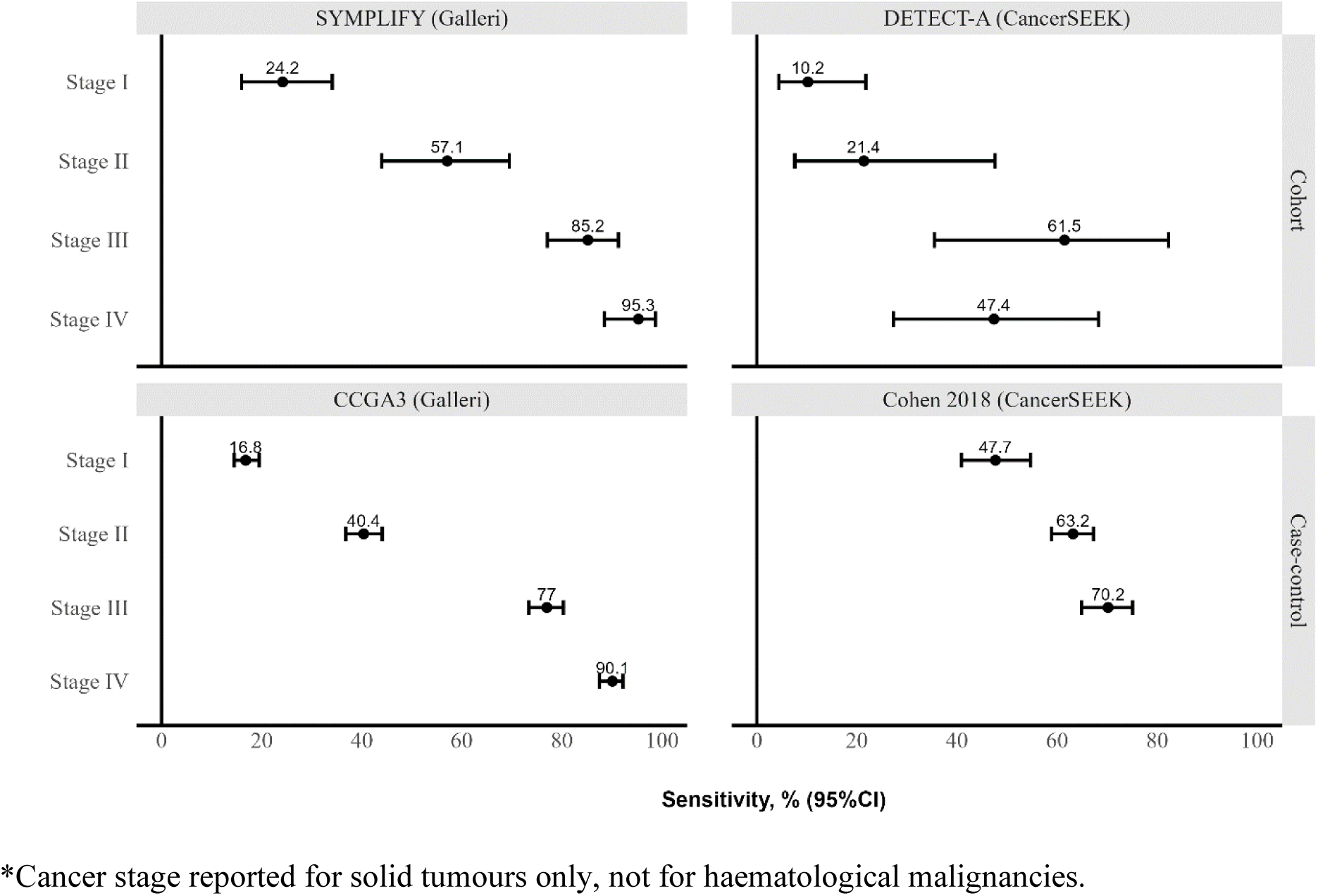
Performance (sensitivity) of MCED tests by cancer stage*.

**Table 4.**
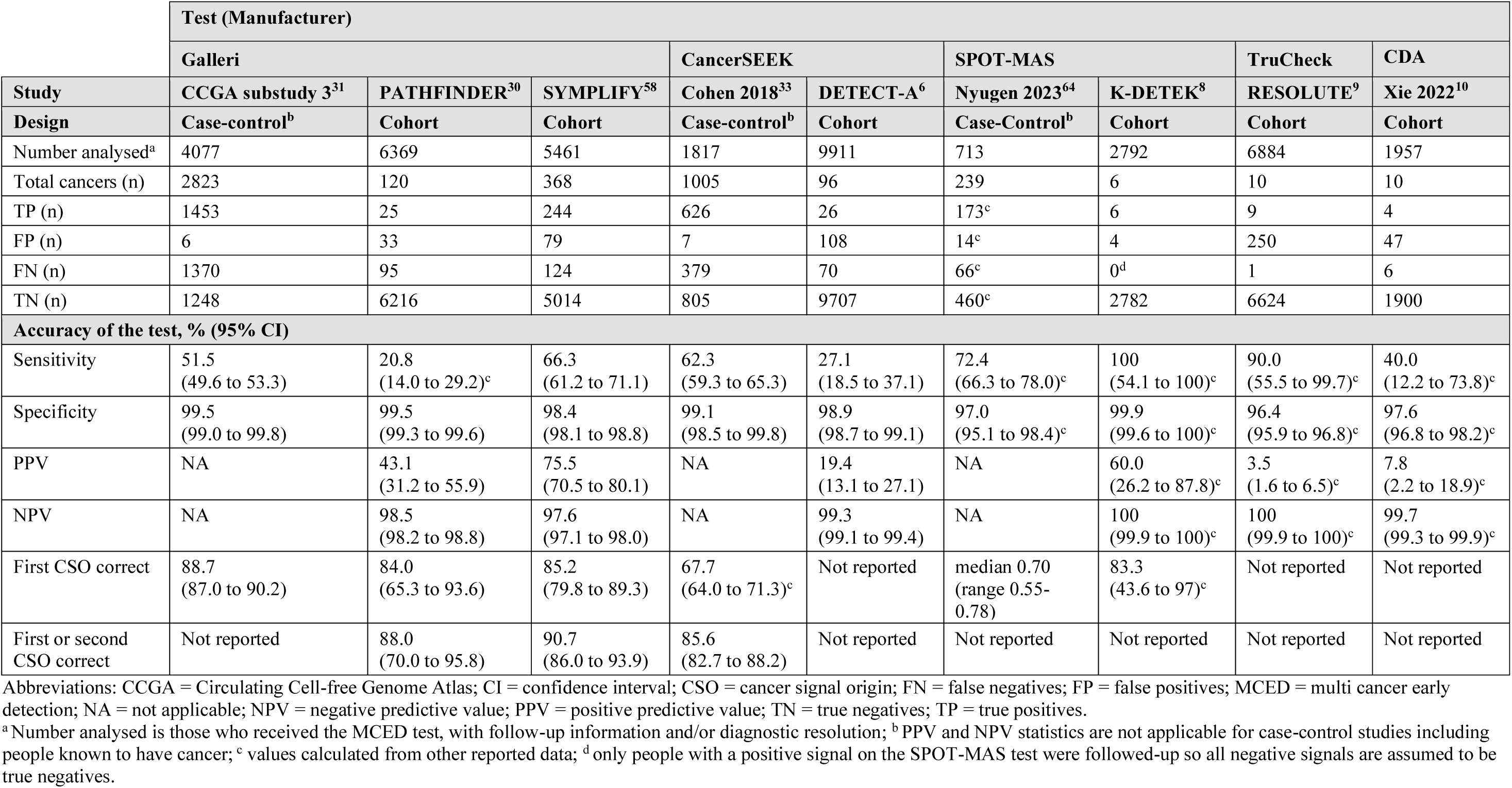
Test performance and accuracy of the tests.

**Table 5.**
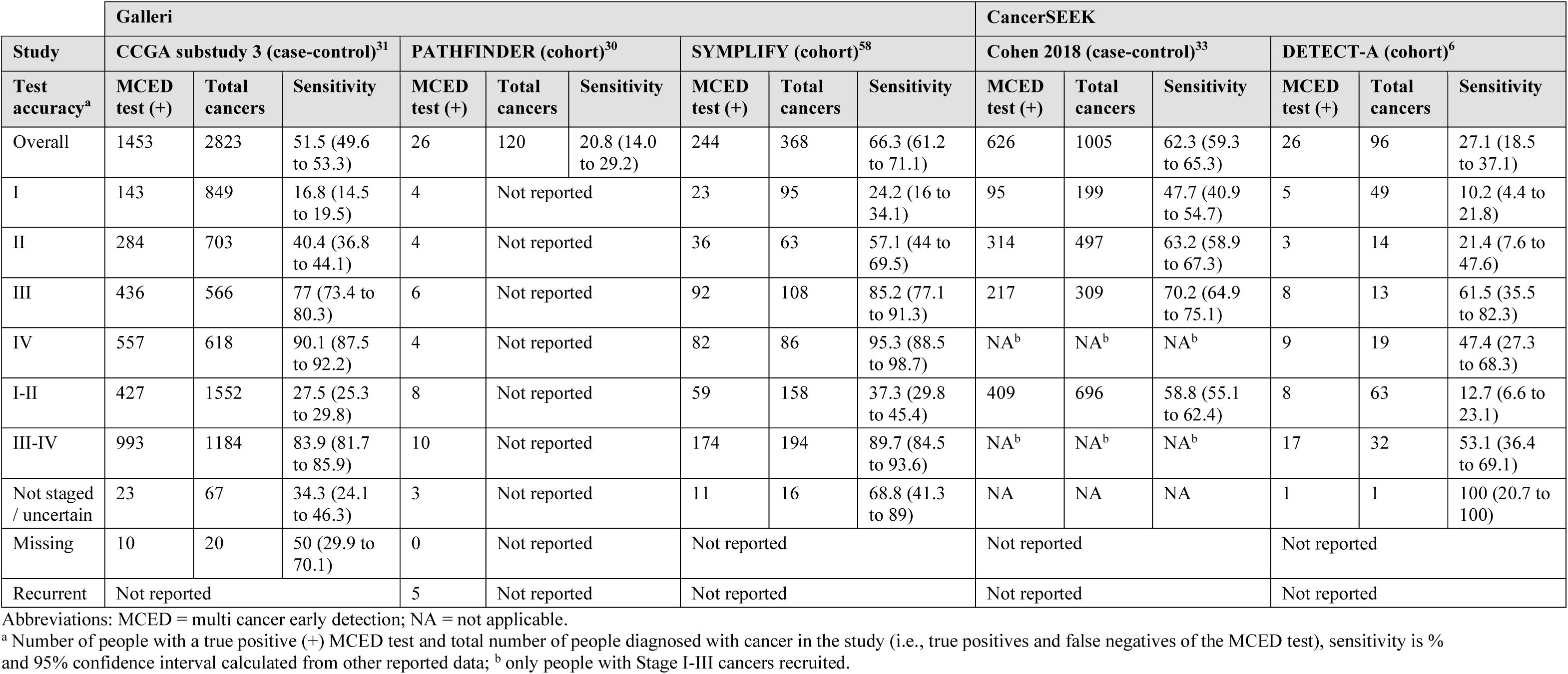
Number and proportion of cancers detected by the MCED tests by stage.

##### 3.4.1.1 Galleri

PATHFINDER^30^ recruited two cohorts: one included participants considered at elevated risk of cancer (n=3655) and another included participants without an elevated cancer risk (n=2923). The primary aim of this study was to assess the accuracy of an old version of the MCED test produced by GRAIL. However, analysis of blood specimens with the refined MCED test (Galleri) was also carried out. The refined MCED test results were not returned to physicians or participants and did not influence diagnostic evaluation. The number of positive cancer signals detected on both the old and refined versions of the MCED test was 41 out of 92 (44.0%, 95% CI 34.2 to 54.2%). The refined MCED test detected fewer positive signals overall and most discordant negatives (42/51; 82.4%) had a haematological MCED cancer signal CSO prediction. The old and refined test versions agreed on 99.7% (95% CI 98.7% to 99.2%) of negative signals (Figure S4 in Schrag et al.^30^). Although carried out in the USA, the participants recruited to this study are reflective of our target population in terms of age and recruited participants are broadly representative of a screening population (i.e., asymptomatic) with some individuals expected to be at higher or lower risk of cancer. However, it is unclear whether the proportions of individuals with and without additional cancer risk factors recruited to the PATHFINDER study are reflective of the UK target screening population.

Table 4 presents results for the refined MCED test (Galleri). Only 120 cancers were detected in 6369 analysed participants (a cancer detection rate of 1.9%), reflecting the asymptomatic population recruited to this study. Sensitivity was low (20.8%, 95% confidence interval [CI] 14.0% to 29.2%) although the first CSO was correct in 84.0% of cancers detected (95% CI 65.3% to 93.6%) increasing to 88.0% (95% CI 70.0% to 95.8%) for first or second CSO. The PPV was 43.1% (95% CI 99.3% to 99.6%). Specificity was high (99.5%, 95% CI 99.3% to 99.6%) and the NPV was also high (98.5%, 95% CI 98.2% to 98.8%), although a short follow-up and lack of reference standard testing on participants with a negative MCED test limits the interpretation of these results.

Fifteen different cancer types were identified. The number of participants with each cancer type are presented in Appendix 7, Table 15 and the number of cancers identified by the MCED test by stage are presented in Table 5. However, the total number of cancers diagnosed (including FN of the MCED test) for each cancer type and at each stage was not reported so these results are difficult to interpret and the sensitivity of the MCED test by different cancer types and stages is unknown.

Performance of the refined MCED test in the elevated and non-elevated risk cohorts are presented in Appendix 8, Table 19. In the elevated risk cohort, 77 cancers were detected in 3532 participants (2.2%); in the non-elevated risk cohort 43 cancers were detected in 2837 participants (1.5%). Sensitivity was lower for the non-elevated risk cohort (16.3%, 95% CI 6.8% to 30.7%) than for the elevated risk cohort (23.4%, 95% CI 14.5% to 34.4%) but specificity remained high for both groups. The proportions of correct first, and first or second CSO, and PPV were lowest for participants without additional cancer risk but specificity and NPV were similar across groups (Appendix 8, Table 19).

Included patients in the SYMPLIFY study^58^ were symptomatic, so not reflective of the target population of interest for this review. Participants were investigated according to current NHS practice and without knowledge of the MCED test results, which were not returned to clinicians or study participants. Results for 5,461 participants with an evaluable MCED test and diagnostic test results are presented in Table 4, of which 368 (6.7%) had a cancer diagnosis. A sensitivity of 66.3% (95% CI 61.2% to 71.1%) and specificity of 98.4% (95% CI 98.1% to 98.8%) were reported, with first CSO correct in 85.2% (79.8% to 89.3%) of cases, rising to 90.7% (86.0% to 93.9%) for first and second CSO.

Sensitivity of the MCED test increased with cancer stage, 37.3% (95% CI 29.8% to 45.4%) for stages I-II and 89.7% (95% CI 84.5% to 93.6%) for stages III-IV (full results by stage are presented in Table 5 and Figure 3). Sensitivity, specificity, and accuracy of CSO also varied by cancer site. Although specificity remained high for all cancer sites (ranging from 96.2% to 100%^58^), sensitivity varied substantially by cancer site although the total number of participants diagnosed with certain types of cancer was low, so results are difficult to interpret (Appendix 7, Table 15). SYMPLIFY also reported sensitivity of the MCED test by cancer site and stage (Supplementary material, page 8 of Nicholson et al.^58^) and shows that sensitivity of the CSO increases by cancer stage for all cancer sites, although there are very small numbers is some categories making results difficult to interpret. The accuracy of the first CSO was higher for stages III-IV and lower for stages I-II (Supplementary material, page 21^58^).

CCGA substudy 3^31^ was a case-control study recruiting 4077 participants where 2823 were known to have cancer (cases, 69%) and 1254 were confirmed not to have cancer at one year follow-up (controls, 31%). Test performance is presented in Table 4. Specificity was high (99.5%, 95% CI 99.0% to 99.8%) with 51.5% sensitivity (95% CI 49.6% to 53.3%) and the first CSO was correct in 88.7% of cases (95% CI 87.0% to 90.2%). Sensitivity of the MCED test increased with cancer stage, being relatively low 27.5% (95% CI 25.3% to 29.8%) for stages I-II but higher for stages III-IV (83.9%, 95% CI 81.7% to 85.9%; full results by stage are presented in Table 5 and Figure 3). Sensitivity also varied by cancer site although the total number of participants diagnosed with certain types of cancer was low, so results are difficult to interpret (Appendix 7, Table 15). Sensitivity of the MCED test by cancer site and stage is also reported in CCGA substudy 3 (Supplementary Table S5 of Klein et al.^31^) and shows that sensitivity of the CSO increases by cancer stage for all cancer sites, although there are very small numbers is some categories, making results difficult to interpret. The accuracy of the first CSO is also reported by cancer type and shows great variability (from 0% to 87%, Supplementary Table S7 of Klein et al.^31^).

##### 3.4.1.2 CancerSEEK

DETECT-A^6^ was a prospective cohort study in the USA, which recruited 9911 women who were followed up for 12 months. In total, 96 women (0.97%) were diagnosed with cancer during the study, 26 of which were first detected by the CancerSEEK test (sensitivity 27.1%, 95% CI 18.5% to 37.1%, Table 4). Specificity was 98.9% (95% CI 98.7% to 99.1%; PPV 19.4%, 95% CI 13.1% to 27.1%; Table 4). The accuracy of the CSO was not reported. The majority of the cancers (65.6%) diagnosed during the DETECT-A^6^ study were Stage I-II (Table 5), however the sensitivity of the CancerSEEK test to detect Stage I-II cancers was lower (12.7%, 95% CI 6.6% to 23.1%) than the sensitivity to detect Stage III-IV cancers (53.1%, 95% CI 36.4% to 69.1%) (full results by stage are presented in Table 5 and Figure 3).

A case-control study of CancerSEEK recruited 1005 patients diagnosed with stage I-III cancers (see Appendix 7, Table 16 for the cancer types among recruited patients) and 812 healthy controls.^33^ Specificity was high (99.1%, 95% CI 98.5 to 99.8%) with only 7 out of 812 (0.9%) healthy controls receiving FP test results (Table 4). However, 379 out of 1005 cancers were not detected by the test (sensitivity 62.3%, 59.3% to 65.3%, Table 4) and first CSO was correct in 67.7% of positive tests across all cancer types, with first CSO correct in less than 50% of cases for liver, lung, and upper gastrointestinal cancers (Table S8 and S10 of Cohen et al., 2018^33^). The proportion of first or second CSO being correct was higher, 85.6% across all cancer types (Table 4). Sensitivity of the CancerSEEK test increased with advancing cancer stage (Figure 3).

The number and proportion of each cancer type detected by the CancerSEEK test is provided in Appendix 7, Table 16. Sensitivity of the CancerSEEK test was highest to detect ovarian cancer and lowest to detect breast cancer in both studies.^6, 33^

The participants included in the DETECT-A study^6^ are closer to the target population of interest in this review (asymptomatic screening 50 to 79 years old), than the participants in Cohen 2018,^33^ although DETECT-A was limited to women aged 65 to 75 years.

##### 3.4.1.3 Other MCED tests

The SPOT-MAS test was evaluated in the K-DETEK^8^ cohort study which recruited 2792 participants over the age of 40 without clinical suspicion of cancer or history of cancer from outpatient clinics in Vietnam. Only the 10 participants (0.36%) who had a positive signal on the SPOT-MAS test were followed-up (for six months) for further diagnostic investigations. Therefore, all negative signals of the SPOT-MAS test were assumed to be TN in the K-DETEK^8^ study. Sensitivity and NPV of the SPOT-MAS test are calculated as 100% (Table 4), although this is unlikely to reflect true test performance. Out of the 10 positive signals, six were confirmed to be cancer (PPV, 60%, 95% CI 26.2% to 87.8%) and four were FP (specificity 99.9%, 95% CI 99.6% to 100%; Table 4). First CSO was correct for 83.3% (five out of six) of the cancers; the types of cancer detected by the SPOT-MAS test in the K-DETEK^8^ study are presented in Appendix 7, Table 17. A case-control study^64^ recruited 239 patients diagnosed with Stage I-IIIA cancers (see Appendix 7, Table 17 for the cancer types among recruited patients) and 474 healthy controls as a validation cohort for the SPOT-MAS test. Specificity of the SPOT-MAS test was high (97.0, 95% CI 95.1% to 98.4%) with only 14 out of the 474 healthy controls with FP results. Sensitivity was 72.43% (95% CI 66.3% to 78.0%) and first CSO was correct for a median of 70% of cancers across all cancer types (Table 4).

Two studies, RESOLUTE and TrueBlood, evaluated the performance of the TruCheck test.^9^ In total, 10 participants out of 6884 (0.15%) were diagnosed with cancer during the RESOLUTE study,^9^ 9 of which were detected by the TruCheck test (sensitivity 90%, 95% CI 55.5% to 99.7%; Table 4; see Appendix 7, Table 18 for the types of cancer detected by the TruCheck test). An FP signal was also returned in 250 participants who were found not to have cancer (specificity 96.4%, 95% CI 95.9% to 96.8%; PPV 3.5%, 95% CI 1.6% to 6.5%; Table 4). In the TrueBlood study,^9^ the sensitivity of the TruCheck test was 93%, correctly detecting cancer in 9224 out of 9920 participants with known or suspected (later confirmed) cancer.

The CDA test was evaluated in the Prospective Population-based Cohort Study (PPCS)^10^ where 1957 were followed-up for a median duration of 15 months (range 12 to 20 months). In total, 10 participants (0.51%) were diagnosed with cancer, 4 of which were detected by the CDA test (sensitivity 40%, 95% CI 12.2% to 73.8%; see Appendix 7, Table 18 for the types of cancer detected by the CDA test). An FP signal was also returned in 47 participants who were found not to have cancer (specificity 97.6%, 95% CI 96.8% to 98.2%; PPV 7.8%, 95% CI 2.2% to 18.9%, Table 4).

The AICS test was evaluated in a cohort study^11^ which followed participants for up to 6.2 years. Sensitivity by cancer type is presented in Appendix 7, Table 18. AICS was also evaluated in the AICS follow-up study^65^ Out of 622 participants with a Rank C (high risk for cancer on the AICS test) who had received a detailed examination in an interim analysis, two cases of prostate cancer and one case of each of lung, colorectal and breast cancer were detected. In another study,^66^ up to 115 healthy women were tested for breast cancer using AICS in Japan, and the authors recommended that where rank B or C is returned from the AICS test, further inspection with mammography should be carried out.

The number of cancers detected by the SPOT-MAS, TruCheck, CDA and AICS tests and the total number of cancers diagnosed in cohort studies was very low due to limited follow-up investigations and/or short follow-up periods (Appendix 7, Table 17 and Table 18). Therefore, sensitivity of these tests and any differences in the sensitivity of the tests by specific cancer types and stages are difficult to interpret. Furthermore, stage of cancer detected was not reported for any of the SPOT-MAS, TruCheck, CDA and AICS studies, and accuracy of CSO was not reported for the TruCheck, CDA or AICS tests.

##### 3.4.1.4 MCED test performance for cancers with and without a current screening programme

The number and proportion of cancer types with and without a current screening programme in the UK detected by the tests are reported in Table 6 and Figure 4, where they could be extracted or calculated. The numbers and proportions of each specific cancer type detected by each MCED test are presented in Appendix 7, Table 15 to Table 18.

**Figure 4.**
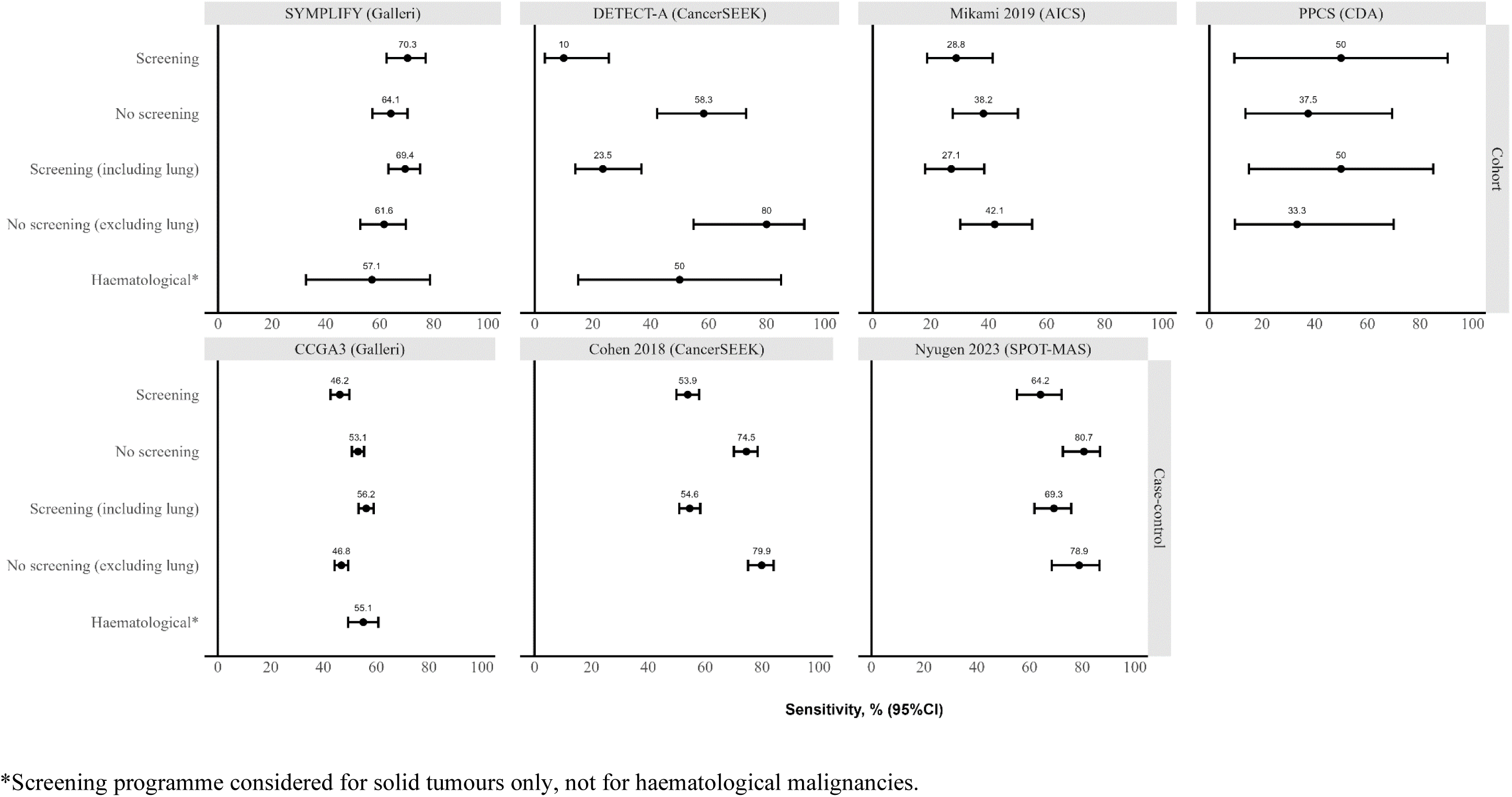
Performance (sensitivity) of MCED tests by the availability of screening programme in the UK.

**Table 6.**
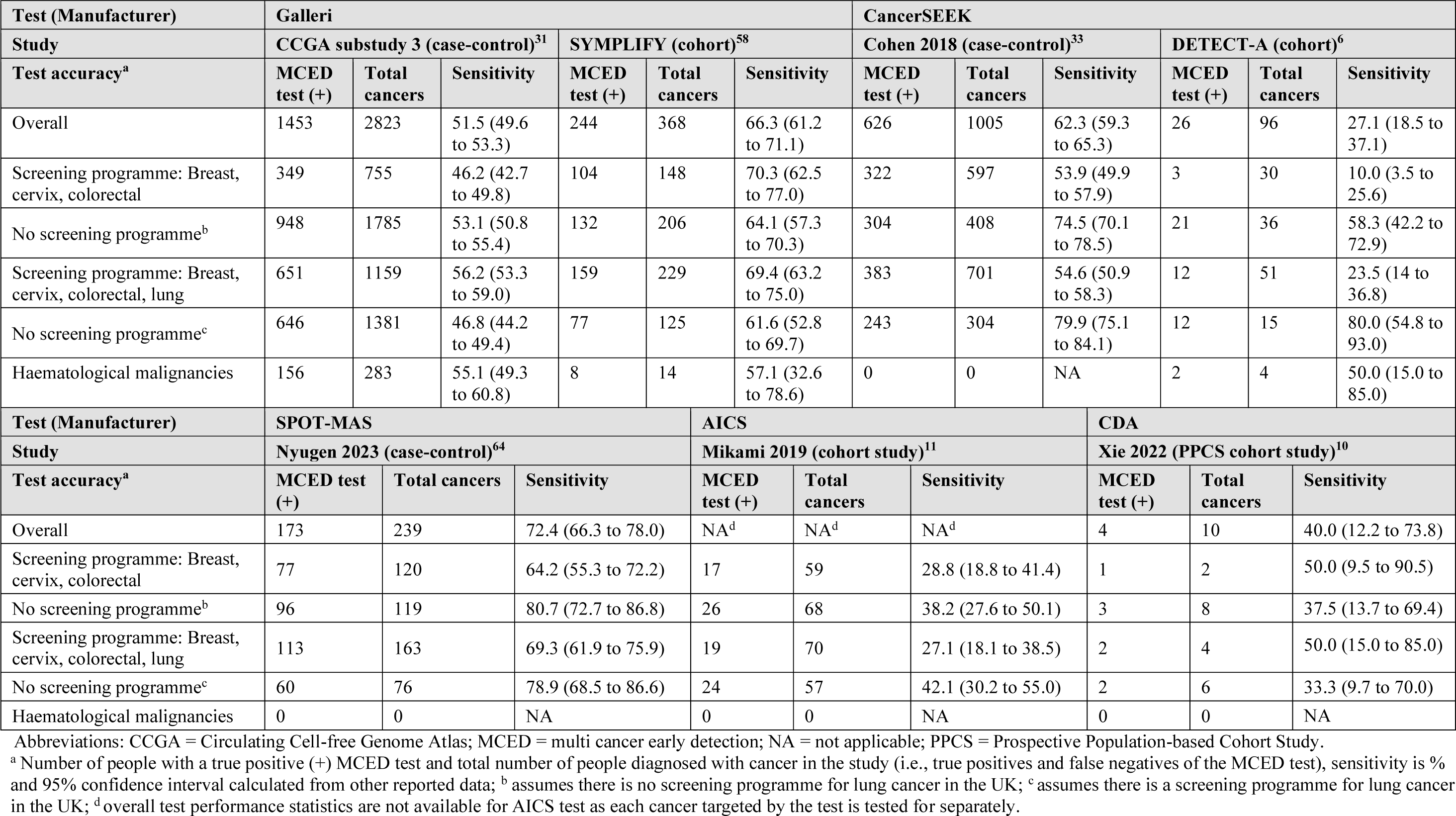
Number and proportion of cancers detected by MCED tests by cancer types with and without a current screening programme in the UK.

In one of the studies of Galleri (CCGA substudy 3^31^) and both studies of CancerSEEK, sensitivity of the tests to detect solid tumour cancers without a current screening programme in the UK was higher than their sensitivity to detect cancers with a current screening programme (breast, cervical and colorectal) in the UK (Table 6, Figure 4). However, when lung cancer is considered covered by existing screening programmes, sensitivity of the Galleri test is higher for solid tumour cancers with a current screening programme in both CCGA substudy 3^31^ and SYMPLIFY^58^ (Table 6, Figure 4), which can be explained by the relatively high sensitivity of the Galleri test to detect lung cancer, compared to its overall sensitivity (Appendix 7, Table 15).

The sensitivity of the SPOT-MAS and AICS tests to detect solid tumour cancers without a current screening programme in the UK was higher than the sensitivity of these MCED tests to detect cancers with a current screening programme in the UK, but the sensitivity for solid tumour cancers without a current screening programme was lower for the CDA test (Table 6, Figure 4).

Haematological malignancies were diagnosed for two studies of Galleri and one study of CancerSEEK (Figure 4). In CCGA substudy 3^31^ and SYMPLIFY^58^ sensitivity of the Galleri test for detecting haematological malignancies was similar to its overall sensitivity. Four haematological malignancies were diagnosed during the DETECT-A study, two of which were detected by the CancerSEEK test (sensitivity 50%, 95% CI 15% to 85%) which is higher that its overall sensitivity (Table 6). No haematological malignancies were diagnosed during the studies of the SPOT-MAS, AICS and CDA tests (Figure 4), although neither the SPOT-MAS nor the AICS test claim to be able to detect these cancers (Appendix 7, Table 14).

##### 3.4.1.5 MCED test performance by subgroups

MCED test performance by pre-specified subgroups of interest (i.e., age, sex, and ethnicity) was reported in or could be calculated from studies of Galleri and CancerSEEK. Subgroup results by socioeconomic status were not reported in any of the included studies.

Test performance results (sensitivity, specificity and first CSO accuracy, where available) by age and ethnicity subgroups from the CCGA substudy 3^31^ of Galleri and the CancerSEEK case-control study^33^ are presented in Appendix 8, Table 20. Specificity was high for all age and ethnicity subgroups in both studies. For CancerSEEK, sensitivity and CSO accuracy were slightly lower for participants less than 50 years of age compared to participants aged 50 years or above while for Galleri, sensitivity and CSO accuracy were very similar across the age categories presented.

Sensitivity of Galleri was highest for Hispanic participants (63%), although with a slightly lower specificity than for other ethnic groups (98% compared to 99 to 100%), and it was lowest (43%) for the small number of participants classified as ‘Other’ in the study. Sensitivity of CancerSEEK ranged from 50% in participants with unknown ethnicities to 70.4% in Asian participants (and cancer was correctly detected by the CancerSEEK test in one Hispanic participant resulting in a sensitivity of 100%). However, any differences in the sensitivity of the Galleri or CancerSEEK test by ethnicity should be carefully interpreted as the majority of participants recruited to studies were White and other ethnic subgroups have much smaller numbers of participants.

A poster presentation reported the number of signals for cancers detected by age categories and sex (male/female, as defined in the publication) as well as the CSO distribution and prediction of accuracy by sex for a subset of the PATHFINDER participants, using the earlier version of the GRAIL MCED test.^84^ Reported data showed that the cancer signal detection rate was similar in males and females and increased with age for both, however few details were given on the subset of participants analysed and a now superseded version of the MCED test was used, so these findings should be interpreted with caution. No subgroup data by ethnicity were reported and no subgroup data were available for the refined MCED test.

#### 3.4.2 Other outcomes reported in included studies

Additional outcomes relevant to this review were reported in studies of Galleri and CancerSEEK, including mortality, potential harms, acceptability, and satisfaction of individuals screened. One study of the AICS test also reported very limited information on survival.

##### 3.4.2.1 Mortality

The DETECT-A study reported mortality outcomes among individuals who received a positive CancerSEEK test result, 4.3 years after the initial study.^85, 86^ Among the 26 participants with a true positive test result, half were in remission; sixteen (62%) were alive (5 at stage I, 4 at stage II, 5 at stage III and 2 at stage IV), of which seven had cancers where no standard screening options are currently available.^86^ All deceased participants had stage III (n=3) or IV cancer (n=7) at the time of diagnosis. Among participants with FP test results, only two developed cancer: one was diagnosed with stage I breast cancer 2.7 years after the test, and one with stage III ovarian cancer 2.9 years after the test.^85^

Information on mortality was reported for the PATHFINDER study for only two participants who were followed-up for more than one year after diagnosis.^87^ One had stage I renal cell carcinoma and stage II head and neck cancer, and after a combination of treatments, was alive and cancer free at ≥502 days after diagnosis.^87^ One had stage IIIB lung cancer, and was alive at ≥683 days post-diagnosis, but metastatic disease had developed.^87^

Survival information for four participants who had the AICS test in the AICS follow-up study^65^ and were diagnosed with cancer (two detected by AICS and two not detected by AICS) was obtained from a cancer registry. All participants were alive at the time the information was obtained and were undergoing treatment.

##### 3.4.2.2 Potential harms and impact on healthcare systems

In the PATHFINDER study, results reported for the earlier version of the GRAIL MCED test showed diagnostic resolution was achieved after initial evaluations in 82% (32 out of 39) of participants with a positive test result, and additional testing was only required for individuals with a cancer history and negative initial evaluation or an equivocal initial evaluation. Whole body imaging was required in 69% cases (27 out of 39), but only contributed to diagnosis in under half of cancer cases (49%), and was only useful when detecting the presence of non-localised cancer.^88^ The median time to diagnostic resolution was 57 days (IQR 33 to 143 days) for TP results and 162 days (IQR 44 to 248 days) for FP results.^30^ Overall, 52% (17/33) participants with a TP test result had at least one clinic visit (average number 0.9 among the 33 participants), compared to 32% (18/57) participants with a FP result (average number 1 among the 57 participants); 79% (26/33) of participants with TP results had at least one lab test (average number of tests 3.7), compared to 88% (50/57) with FP results (average number 4); and 91% (30/33) individuals with TP results had imaging tests (average number 1.5), compared to 93% (53/57) of those with FP results (average number 1.9).^30^ More participants with TP test results had surgical and non-surgical procedures compared to those with FP results (82% vs 30%).^30^ Similar findings might be expected for participants testing positive with the refined version of the GRAIL MCED test (Galleri). In the SYMPLIFY study, median time to diagnosis was 35 days (lower quartile 20, upper quartile 57 days, Supplementary material, page 21 in Nicholson et al.^58^). However, this outcome is not directly relevant to the asymptomatic screening population considered in this review, since included patients had already been referred, and investigations were not triggered by positive MCED test and CSO results.

In the DETECT-A study, no adverse events were reported from the CancerSEEK test directly. 101 participants with a positive MCED test result underwent confirmatory positron emission tomography-computed tomography (PET-CT), a form of CT with radiation exposure more than twice as high as the exposure from standard CT; 62% required no further follow-up, while 16% had non-invasive procedures, 19% had minimally invasive procedures, and 3% (3 cases) required surgery.^6^ Another potential source of harm may come from decreased adherence to standard of care screening following a negative result. However, no differences in the proportion of participants who had a mammogram before and after enrolment in the study were found.^6^

##### 3.4.2.3 Acceptability and satisfaction

Both Galleri and CancerSEEK were reported as being generally acceptable to participants, with 97.1% and 95.0% of participants reporting high satisfaction with MCED testing or with participating in the study, respectively.^6, 89^ Among DETECT-A participants who received false results on the CancerSEEK test (false positives/negatives), 0.8% reported dissatisfaction and 1.7% would not participate in the study again, compared to 0.2% and 1% of those who received the correct results (true positives/negatives).^6^ Similarly, in the PATHFINDER study, 17.6% of participants who had a FP result reported dissatisfaction with MCED testing, compared to 8.0% of those with a TP result, and 2.8% of those with a negative result on the Galleri test.^89^

The PATHFINDER study additionally reported changes in anxiety levels of participants receiving the GRAIL MCED test, across different stages of the study (at pre-test, return of results, diagnostic resolution, and end of study).^89^ Although the overall mean levels of anxiety did not change substantially, the proportion of participants who reported increased anxiety (defined as scoring 3 points or more on the PROMIS [Patient-Reported Outcomes Measurement Information System] anxiety scale) changed between different stages of the study. However, the study did not examine anxiety levels whilst participants were waiting for test results, only after they had received the results. When participants received results of the MCED test, around half of those who received a positive result reported increased anxiety (57.9% for TP, 46.4% for FP).^89^After diagnostic resolution, the proportion with increased anxiety levels halved for the FP group (24.2%), and was similar to those with a negative test result (27.4%), while the proportion remained the same for the TP group (56.0%).^89^

A study to evaluate the implementation of the Galleri test as an employee benefit among individuals with low socioeconomic status, identified a number of factors that were important for test uptake, including: the test being an on-site event; having trusted long-term employees on site that spoke the same language and helped with any translations as necessary; the test results being explained in their native language; and the ability to administer the test without computer or digital equipment.^59^ However, this evaluation relied on employer insight, employee feedback and observations of GRAIL staff, so there is a potential for considerable bias in these results.

#### 3.4.3 MCED technologies at an unclear stage of development

The type of outcomes reported for MCED technologies at an unclear stage of development varied across studies and were not directly comparable with one another. Therefore, a short summary is provided below, and further details on study characteristics as well as any results reported can be found in Appendix 6, Table 12.

All studies were case-control studies, except for two publications from SeekInCare^75, 76^ which reported both case-control and prospective cohort studies including ‘real-world’ cohorts. A case-control study of CancerenD24,^70^ reported that cancer patients and healthy controls were matched on ethnicity but did not include a description of the different ethnic groups. Ethnicity was not reported in any other study. All studies reported on the accuracy of the MCED technology, including sensitivity and specificity, and some studies also reported these for each stage/type of cancer detected. OncoSeek reported the lowest overall sensitivity across all cancer types (47.4%),^73^ and CancerenD24 reported the lowest sensitivity in detecting bladder cancer (38.0%).^70^ By stage, OverC and SeekInCare reported a sensitivity of 35.4% and 50.3%,^75, 77^ respectively, for stage I cancer. The highest sensitivity overall came from the Carcimun-test (88.8%),^81^ however, the exclusion of individuals with inflammation is noted as a disadvantage of the technology as a screening tool in the general population. The SpecGastro test was only developed to detect three types of gastrointestinal cancer (colorectal, gastric, and oesophageal).^82^

### 3.5 Ongoing studies of included technologies

The NHS-Galleri trial^60^ is the only ongoing RCT identified. An interim analysis of NHS-Galleri at one-year post-randomisation is planned for late 2023/early 2024,^90^ which is expected to report on the number of stage IV cancers detected in each study arm. NHS-Galleri and PATHFINDER2 both plan to recruit healthy volunteers over 50 years of age (up to age 77 in NHS-Galleri), which is reflective of the target screening population for this review.^60, 61^ A list of ongoing studies identified for the included technologies is presented in Table 7.

**Table 7.** Summary of ongoing studies for included technologies.

| Study details | Participant information | Intervention | Outcomes |
| --- | --- | --- | --- |
| **GRAIL Galleri** |  |  |  |
| NHS-Galleri<sup>60</sup><br>Clinical trial identifier: ISRCTN91431511<br>NCT05611632<br>Randomised controlled trial<br>Estimated completion date: 28/02/26 | Healthy volunteers aged 50 to 77 | Blood collection and Galleri (MCED) test<br>Comparator: blood collection and storage for potential future evaluation | Incidence rate of stage III and IV cancers adjusted by the follow-up time in the intervention arm compared with the control arm; test performance; safety; impact on healthcare resource utilisation; cancer-specific mortality; potential impact of overdiagnosis |
| PATHFINDER2<sup>61</sup><br>Clinical trial identifier: NCT05155605<br>Prospective single-arm trial<br>Estimated completion date: 30/07/26 | Individuals aged $\geq 50$ | GRAIL MCED blood test | Safety; test performance; anxiety; participant-reported intention to follow guideline recommended cancer screening procedures; cancer detection rate of PET-CT; number and type of diagnostic evaluations; radiation exposure; accuracy amongst subgroups; perceptions of the MCED test |
| REFLECTION<sup>62</sup><br>Clinical trial identifier: NCT05205967<br>Prospective cohort study<br>Estimated completion date: 23/08/26 | Individuals aged $\geq 22$ who have opted to be screened with Galleri MCED test | Galleri blood based MCED test | Signal detection and cancer detection among participants; feasibility and acceptability; healthcare resource utilisation associated with cancer diagnostic workups for participants with signal detected |
| SUMMIT<sup>63</sup><br>Clinical trial identifier: NCT03934866<br>Prospective cohort study<br>Estimated completion date: August 2030 | Individuals aged 55 to 77 who are at high risk for lung cancer due to a significant smoking history | GRAIL blood test and low dose CT scan at the same visit | Test performance of GRAIL blood test and of delivering a low-dose CT screening service |
| **OverC – Burning Rock Biotech** |  |  |  |
| PREVENT<sup>79</sup><br>Clinical trial identifier: NCT05227534<br>Prospective cohort study<br>Estimated completion date: 31/12/28 | Asymptomatic participants with cancer risk, aged 40-75 | OverC multi-cancer detection blood test | Accuracy of the test after 1, 3 and 5 years; health-related quality of life; acceptability (satisfaction with the test) |
| PREDICT<sup>80</sup><br>Clinical trial identifier: NCT04817306<br>Prospective case-control study<br>Estimated completion date: 31/03/23 | Cancer patients, those with benign diseases, and healthy controls aged 40-75 | OverC multi-cancer detection blood test | Accuracy of the test; accuracy of CSO |

Abbreviations: CT = computed tomography; MCED = multi-cancer early detection; PET-CT = positron emission tomography-computed tomography.

### 3.6 Technologies excluded from the review

As discussed in Section 3.2, full-text screening identified several blood-based MCED technologies currently at an early stage of development and not ready to be implemented, which did not meet the inclusion criteria for this review. Given the fast-moving pace of research in this area, some of these may become available in the near future. We therefore provide a brief, non-exhaustive summary of some of these technologies below including: DELFI (DNA evaluation of fragments for early interception) developed by DELFI Diagnostics (Baltimore, Maryland), with two ongoing clinical trials evaluating its use in detecting lung cancer;^91, 92^ Aurora (AnchorDx, Guangzhou, China), which detects five types of cancer and has a planned clinical trial in asymptomatic populations;^93, 94^ PanTum (Zygnum AG, Darmstadt, Germany), with two ongoing clinical trials in China^95^ and India;^96^ LUNAR-2 (Guardant Health, Palo Alto, California), with an ongoing trial in individuals at high-risk of cancer;^97-99^ and HarbingerHx (Harbinger Health, Cambridge, Massachusetts), with an ongoing case-control study and expected product launch date in 2025.^100^ Further details can be found in Table 8.

**Table 8.** Summary of technologies excluded from the review.

| Technology | Manufacturer | Description | Completed/ongoing studies |
| --- | --- | --- | --- |
| Adela's MCED tests | Adela Bio (Foster City, California) | A genome-wide methylome enrichment platform that combines cfDNA with machine learning. | <u>Ongoing:</u><br>CAMPERR<sup>101</sup> is an ongoing case-control study to evaluate the test across 20 types of cancer. |
| Aurora | AnchorDx (Guangzhou, China) | A targeted methylation profiling platform capturing cancer-specific DNA methylation signatures across five cancer types (lung, breast, colorectal, gastric, esophageal) | <u>Completed:</u><br>Pre-clinical studies used plasma samples in a training/validation cohort.<sup>93, 94</sup><br>A large prospective clinical trial is planned in asymptomatic populations. |
| CAPP-Seq | Diehn Lab at Stanford (Stanford, California) | Cancer personalised profiling by deep sequencing – a method for quantifying circulating tumour DNA (ctDNA). | <u>Completed:</u><br>Initially implemented for non-small cell lung cancer.<sup>102</sup> Generalisable to other tumour types and work is ongoing to establish its clinical utility as an early detection tool for cancer. |
| DELFI | Delfi Diagnostics (Baltimore, Maryland) | DNA evaluation of fragments for early interception. Uses a machine learning algorithm to detect abnormalities of cfDNA across the genome. | <u>Completed:</u><br>Pre-clinical study using plasma samples to detect 7 types of cancer.<sup>91, 92</sup><br><u>Ongoing:</u><br>Two ongoing clinical trials on lung cancer: DELFI-L101<sup>103</sup> and DELFI-L201<sup>104</sup> (also known as CASCADE-LUNG), and one ongoing clinical trial (DETECT study,<sup>105</sup> past completion date) to detect cancer in liver transplant recipients. |
| Dxcover | Dxcover Limited (Glasgow, United Kingdom) | A blood-based test using infrared spectroscopy combined with machine learning to screen for eight types of cancer (brain, breast, colorectal, kidney, lung, ovarian, pancreatic, and prostate) | <u>Completed:</u><br>Discovery stage study using biobank samples to differentiate non cancer symptomatic from cancer patients.<sup>106</sup> |
| Elypta's MCED test | Elypta (Solna, Sweden) | A metabolism-based liquid biopsy using profiles of human glycosaminoglycans (GAGome). | <u>Ongoing:</u><br>An ongoing study<sup>107</sup> is assessing the performance of the test measured in plasma, in urine, or both in a prospective cohort of firefighters. |
| HarbingerHx | Harbinger Health (Cambridge, Massachusetts) | A platform that combines ctDNA with machine learning for early detection of cancer. Expected to launch in 2025.<sup>108</sup> | <u>Ongoing:</u><br>CORE-HH<sup>100</sup> is an ongoing case-control study to assess the performance of the platform in detecting cancer. |
| LUNAR-2 | Guardant Health (Palo Alto, California) | A blood-based test initially designed to detect colorectal cancer, but ongoing trials are evaluating its use in other types of cancer. | <u>Ongoing:</u><br>SHIELD<sup>97-99</sup> is an ongoing study of individuals at high-risk of cancer (first cohort will be focused on lung cancer). |
| MERCURY | Geneseeq Technology (Toronto, Canada) | A blood-based test using cfDNA features for multi-cancer early detection. | <u>Completed:</u><br>Evaluated in a case-control study of 3 types of cancer (liver, colorectal, lung).<sup>109</sup><br><u>Ongoing:</u><br>The Jinling cohort<sup>110</sup> is an ongoing prospective cohort study evaluating the use of MERCURY test in an average risk population. |
| MNALDI | Not reported | Multiplexed nanomaterial-assisted laser desorption/ionization for cancer identification | <u>Completed:</u><br>Pre-clinical study using plasma samples from two hospitals in China to detect six different cancers (liver, lung, pancreatic, colorectal, stomach, thyroid).<sup>111</sup> |
| PanSeerX | Singlera Genomics (San Diego, California) | A blood-based cancer screening test based on cancer-specific methylation signatures. | <u>Ongoing:</u><br>The FuSion Programme<sup>112-114</sup> is an ongoing prospective cohort study of asymptomatic individuals to evaluate the performance of the PanSeer assay. |
| PanTum | Zygnum AG (Darmstadt, Germany) | EDIM (Epitope detection in monocytes) technology focuses on the detection of two biomarkers (Apo10 and TKTL1) in monocytes, tested in 8 different types of cancer.<sup>115</sup> | <u>Completed:</u><br>Early case-control study evaluating its use in 3 types of cancer (bile duct, colorectal, pancreatic).<sup>116</sup><br><u>Ongoing:</u><br>Two ongoing clinical trials in China<sup>95</sup> and India.<sup>96</sup> |
| Raman Spectroscopy | Epigeneres Biotech (Mumbai, India) | Identifies cancer using biochemical fingerprints of Raman Spectroscopy and expression patterns of polymerase chain reaction. | <u>Ongoing:</u><br>An ongoing clinical trial<sup>117</sup> to assess the feasibility of Raman Spectroscopy as a screening tool for cancer detection in India. |
| TEC-Seq | Not reported | Targeted error correction sequencing of cfDNA from 58 genes, based on four types of cancer (colorectal, lung, ovarian, and breast). | <u>Completed:</u><br>Initial validation was done using plasma samples of patients and healthy controls.<sup>118</sup> |
| YiDiXue | Shenzhen Keruida health technology (Shenzhen, China) | A blood-based multi-cancer early detection test. | <u>Ongoing:</u><br>SZ-PILOT Study<sup>119</sup> is an ongoing case-control study to evaluate the clinical efficacy of the YiDiXue test. |

## 4 STAKEHOLDER ENGAGEMENT

At the protocol stage, early discussion with stakeholder representatives acknowledged the potential value of early diagnosis where this might result in improved treatment outcomes and survival rates, but this consultation also highlighted the importance of taking account of issues with the possible implementation of these tests including:

- Resource use and potential impact on existing diagnostic services (including any resulting need for further investigation/confirmation and waiting times between diagnosis and treatment, as well as planned frequency of testing)
- Impact on wider care pathways (including primary care)
- The need to balance any benefits with potential risks to patients and the public (including anxiety, the risks associated with both false positive and false negative test results, the potential active identification of cancers that might otherwise prove unproblematic for screened individuals, and the possible lack of effective treatment)
- Consideration of factors likely to affect test uptake (including possible health inequalities, such as ethnic group and socio-economic status).

Comments received at protocol stage also reinforced the importance of patient relevant outcomes, resulting in the inclusion of outcomes related to potential harms, HRQoL, acceptability to individuals screened, and satisfaction of individuals screened.

The initial stakeholder group (as listed in the Acknowledgements) were also invited to comment on a draft version of the final report, particularly to check technical descriptions, handling of available tests and tests in development, and presentation of study details for each test. All agreed these were appropriate, particularly in view of the early stage of development of these technologies and the rapidly growing evidence base. Those consulted also noted that important details about the potential benefits, harms, and possible unintended consequences of implementing these tests in the UK were often not reported, limiting the relevance of available evidence for policy decision-making. Concerns were expressed about the limitations of the current evidence base and the need for improved understanding of the natural history of, and treatment outcomes for, early-stage cancers detected by MCED tests in healthy individuals at different ages, particularly older people. Several stakeholders also expressed concerns at the high risk of bias ratings for all of the studies, and commented on the wide variation in the nature of the MCED tests as well as variability in the study findings, noting inherent difficulties in distinguishing between the quality of the study, the context in which it was undertaken, and the value of the test itself. Other feedback fell into three broad areas relating to the poor applicability and generalisability of the available evidence, the potential impact of MCED screening on existing screening, diagnostic and treatment pathways and the acceptability and potential impact on populations offered and/or receiving screening.

Following conclusion of the systematic literature review work, additional PPI consultation explored the broader views of patients and the public about the use of MCED tests as part of a general population screening programme.

Feedback from all stakeholder engagement is summarised below under six main themes

1. **Poor applicability and generalisability of available evidence**
  - *Population of interest:* Where reported, substantial differences between study participant characteristics and the target population for this review (the anticipated UK screened population), including population age range, ethnicity and cancer stage and type, were noted.
  - *Relevance to UK context:* Given that the review only identified one UK based study, and that substantial differences in the organisation and resourcing of services exist across the different healthcare environments in which studies were undertaken, the applicability of the current evidence base was questioned.
2. **Limitations of the current evidence base**
  - *Effectiveness of MCED tests in identifying cancers.* Whilst recognising the early stage of this research, contributors wanted reassurance that MCED tests actually worked, and that high quality evidence was available to decision-makers before general population screening programmes were considered. Several PPI contributors queried whether tests claiming to identify a very broad spectrum of cancers might actually be less appropriate to the NHS than tests that claim to identify fewer, treatable, cancers with a good prognosis and higher likelihood of recovery (especially where these were not already covered by an existing screening programme). They also raised concerns about how test effectiveness was being measured and whether an appropriate spectrum of outcomes is being considered.
  - *Balancing effectiveness and cost-effectiveness.* PPI contributors wanted much more detailed information about the variety of tests available, their respective cost, and accompanying claims about the numbers and types of cancers targeted, expressing concerns about both the commercial sector support for existing research and the cost-effectiveness of MCED tests for the NHS. They highlighted the need for better quality information and evidence from future independently conducted research and evaluation.
3. **Potential impact of MCED screening on existing screening, diagnostic and treatment pathways**
  - *Unknown effect on existing screening programmes*. Concerns were raised about the lack of evidence around implementation of MCED screening alongside existing, potentially duplicative, cancer-specific screening programmes. In the event of a negative MCED result, the potential to reduce participation in already established and demonstrably effective screening programmes (particularly where the screening process might be less appealing to patients) was highlighted. This could actually result in a reduction in the detection of early stage disease and the potential for increased mortality.
  - *Likely increased pressure on existing screening and diagnostic services.* Although little is known about plans for the implementation of an MCED screening programme in the UK, many issues were raised around the possible impact on already stretched blood testing services and diagnostic pathways. It was also noted that the current evidence base provides little to guide decisions about the appropriate frequency of MCED screening and optimal length of follow-up, especially in the context of existing cancer-specific screening programmes and taking account of patient characteristics, such as increasing age.
  - *Likely increased pressure on existing treatment and support services and resources.* The possible impact on primary, secondary and tertiary care was raised; the consequences of screening a large proportion of the healthy population should not be underestimated given the potential increase in NHS/healthcare system costs.
  - *Implications for general practice.* The practical implications for general practice were of particular concern, especially given current appointment difficulties and limited consultation time. All stakeholders noted that many cancer symptoms are also common in benign conditions, making them difficult to discriminate and potentially resulting in missed opportunities for early diagnosis, and that there may already sometimes be a lack of consistency in screening and referral decisions by GPs. PPI contributors suggested that clear guidance could be formulated to clarify the circumstance in which GPs should be able to refer patients for MCED screening, particularly if the introduction of these tests results in patients becoming less willing to report symptoms to GPs in case they might become ineligible for screening.
  - *Timely and appropriate communication of results.* PPI contributors, in particular, highlighted the considerable anxiety experienced by both patients and families awaiting test results, but also the importance of good support when results are communicated. Concerns were raised about the variety of ways that MCED test results might be shared with screened individuals, and the potentially damaging impact of some of these regardless of outcome. Furthermore, the likely need for increased anxiety management and support required after a positive result, especially in the case of a false positive finding, was acknowledged. The importance of evaluating and establishing resulting effects on general practice workload was considered a priority, especially in view of current pressures.
4. **Opportunities to enhance services to improve outcomes**
  - *Implementation of decision support tools and improved education for GPs.* Having better support systems in place for GPs was considered critical. One content expert cited experience with other screening programmes where challenges had been experienced in separating out use of tests for screening in asymptomatic populations and in populations with symptoms. Additional training on the appropriate use of MCED tests on the diagnostic testing pathway might also be required, especially if GPs were able to make referrals for screening tests. The value of clear and properly applied decision support systems in this context was highlighted.
  - *Appropriate health service design and resourcing.* Contributors acknowledged that the proposed implementation of an effective MCED test as part of a general population screening programme could, in theory, improve existing services if properly integrated, but that this would inevitably result in increased NHS costs, bringing more people into the system, resulting in the need for further testing, and placing additional burden on an already stretched system. PPI contributors noted the potential to improve efficiency, patient experience and screening uptake, were screening programmes to be integrated, perhaps via dedicated and suitably located community screening and diagnostic hubs to maximise opportunities for access. Additionally, the involvement of nurses, physician associates, and community pharmacists to support accurately and clearly communicating screening test results, potentially alongside general health checks and advice, was strongly favoured. It was acknowledged that the effectiveness, cost-effectiveness and acceptability of any accompanying service design changes would need to be properly evaluated in future research.
  - *Integrating general population screening with targeted health checks.* PPI contributors noted
  the positive impact that contact with cancer services has on lifestyle behaviours, and that implementing a general population cancer screening programme of this sort could also provide an excellent opportunity for prevention initiatives, for example, through undertaking general health checks and providing lifestyle advice and information. This might be especially important in the case of a negative test result.
5. **Acceptability and potential impact on populations offered and/or receiving screening**
  - *Acceptability of the MCED screening test.* All stakeholders agreed that acceptability was paramount, and that, whilst the acceptability of a simple blood test might be quite high, little evidence is available to confirm this. The likelihood that acceptability and uptake would not be distributed evenly across the population eligible for screening and the associated potential for exacerbating existing health inequalities was noted. PPI contributors also noted that regular MCED testing might, in some groups, actually reduce uptake of other possibly less acceptable screening tests. The need to properly demonstrate improved outcomes as a result of MCED screening across all populations was considered a priority.
  - *Acceptability of MCED test outcomes.* Stakeholders repeatedly observed the possibility that MCED tests could have the potential to detect early-stage cancers that, for many, might never result in symptoms or significant morbidity, particularly in older people. Consideration of the impact of unnecessary distress and potentially invasive intervention is currently absent from the existing evidence base.
  - *The effects of false positives on those screened.* Although the information provided to those invited for screening might be critical to uptake, concerns were expressed about the possible impact of a false positive test result, both in terms of unnecessary anxiety and distress caused, and also on subsequent confidence in screening programmes and diagnostic services. The need to better understand the impact on MCED screened individuals and their families was noted.
  - *The effect of a negative MCED test outcome on those screened.* The potential for undue reassurance and changes in other health-related behaviours (including routine screening uptake) following a negative MCED test result was noted, with the possible impact greater in some groups; again, the need to better understand the wider effects of different MCED test outcomes was highlighted.
  - *Poorly reported or missing patient relevant outcomes.* The need for improved collection of patient relevant outcomes in future research was emphasized by all stakeholders, but especially given their importance in cost-effectiveness assessments. In particular, the vital need to assess the performance of MCED screening using mortality endpoints was emphasised, not only due to its importance for patients, but also because of known inaccuracies in existing staging investigations at diagnosis, and the possibility that MCED tests might exacerbate these problems due to their mechanism of action (detection of evidence of cancer in the circulating blood).
6. **Targeting specific groups to support early identification and improve outcomes** PPI contributors highlighted a number of considerations around the adoption of MCED testing, noting the need to balance test accuracy and cost with the likelihood of improving outcomes for NHS patients. In particular they were interested in exploring options for a more focused approach to MCED screening, for example, use of MCED tests that targeted:
  - cancers not currently covered by existing cancer screening programmes (even if these tests identified fewer cancers)
  - cancers that are treatable/stageable where outcomes might be improved (even if these tests identified fewer cancers)
  - groups recognised as being at high risk of certain cancers (rather than in the general population)
  - groups less likely to engage with health services (to facilitate earlier identification)
  - younger age groups of 30-40 years or younger (to facilitate earlier identification)
  - people in remission following successful cancer treatment (where appropriate/feasible)

## 5 PATIENT AND PUBLIC INVOLVEMENT

As part of this study, we aimed to include the perspectives of patients and the public (along with other stakeholders) in both our protocol development and to help us better understand, interpret and contextualise the findings from the review.

We used a range of methods to achieve this, including inviting and receiving comments on the draft protocol prior to undertaking the review, incorporating limited feedback and observations on the draft final report, and hosting both group and individual discussions with representatives from the wider community, including people with lived experience of a cancer diagnosis, carers and those potentially eligible for screening.

Feedback at protocol stage resulted in the inclusion of additional patient relevant outcomes (including potential harms, HRQoL, acceptability to individuals screened, and satisfaction of individuals screened). It also highlighted the importance of broader issues of consideration to the implementation of such a screening programme. Subsequent PPI consultation was designed to further explore the issues identified through earlier stakeholder feedback, including resource use and potential impact on existing services, the need to balance any benefits with potential risks to patients and the public, and consideration of factors likely to affect test uptake.

The group PPI session provided an opportunity for a more reflective discussion on the issues raised, offering a more nuanced interpretation of these, as well as raising several additional themes, including limitations in the current evidence base, accompanying opportunities to enhance services to improve outcomes, and the potential for a more targeted population approach for MCED screening.

The nature of the evidence synthesis brief necessitated a focus on the existing evidence base to support future decision-making, primarily in terms of developing subsequent research. The short timeframe allowed for this work impacted the feasibility of stakeholder involvement generally and patient and public involvement in particular. We had an opportunity to involve PPI contributors at the conclusion of this work, and designed a process in collaboration with our partner organisation, Healthwatch York, to maximise involvement as we were able to give potential contributors only short notice to join a group discussion. We targeted a number of different organisations and individuals in our network (as listed, with grateful thanks, in our Acknowledgements), many of whom provided exceptional support with recruiting potential participants. Using Zoom, we were able to involve people from a range of backgrounds and geographical locations, and with a wide variety of experiences. All PPI contributors actively engaged in the discussions, enriching our understanding of considerations around the implementation of these tests as part of a general population screening programme.

The findings of this review raised many questions for stakeholders, and the PPI consultation emphasised the vital importance of good communication with patients and the public about our understanding of the current evidence base for these tests. Our project engagement work to date has provided a strong foundation for effective dissemination through existing PPI contributors, as well as strengthening and fostering relationships with key organisations via our Healthwatch channels and cancer related PPI groups.

In line with University of York Policy (Payment of Individuals for Involvement with and Contribution to Research), all PPI contributors were offered honoraria in the form of a gift voucher to acknowledge their time and contribution. By agreement, all PPI contributors were also either acknowledged by name or in association with the organisations with which they were affiliated.

The reporting of our patient and public involvement is aligned with the Guidance for Reporting Involvement of Patients and the Public Short Form (GRIPP2)^120^ as detailed in Appendix 9.

## 6 EQUALITY, DIVERSITY AND INCLUSION (EDI)

The independent research team for this project comprised a range of experience and expertise, and included both junior and senior methodologists. This review was designed to inform decisions about future research on the use of MCED tests as part of a general population screening programme, specifically focusing on people without cancer symptoms aged 50-79 years. As such, we took a pragmatic approach to stakeholder engagement, inviting protocol and report feedback from content experts with specific knowledge of equity and diversity considerations in screening, as well as inviting contributions and input from people meeting the population criteria for the review and people and carers with lived experience of a cancer diagnosis. Comments, views and feedback from organisations and people across the UK representing these populations were included in the review.

The available evidence has limited generalisability to the population of interest in this review and no directly applicable evidence was available to indicate the impact of an MCED screening programme on different groups. However, all stakeholders emphasised the potential, without appropriate mitigations, for an MCED based screening programme to exacerbate existing health inequalities. The concerns raised reflected recognised differences in motivation, willingness and practical difficulties in taking up the offer of screening amongst different groups (for example, working mothers or those with childcare responsibilities who may not prioritise their own health, those without a permanent address or who are homeless and therefore not registered with a GP, and those in particular types of employment where flexibility is limited). Likely differences in uptake and outcomes amongst different ethnic and socio-economic groups were also emphasised. Finally, the importance of considering overall patient burden was noted, particularly in terms of convenience of access to screening and any subsequent diagnostic testing (especially in remote areas), and the necessary travel time and associated cost (in both urban and rural areas) associated with this. All these observations mirror the evidence for differential access and uptake in other cancer screening programmes.^121-125^ From the evidence reviewed and the accompanying stakeholder feedback, it is clear that the feasibility, accessibility and impact of such a screening programme on a broad range of different groups requires detailed evaluation and mitigations may be required.^126-128^

The evidence in this field can be complex and difficult to understand, but every effort was made to ensure the language and terminology used in our report was accessible and understandable. The report was edited in response to feedback about terminology or concepts necessary to understand the evidence base and, where necessary, more detailed explanation was incorporated in the text. Additionally, several visual representations were incorporated to simplify presentation of some complex results and findings.

Acknowledging the tight timetable for delivery of this project, to maximise opportunities for engagement and reduce burden on PPI contributors, we circulated invitations to participate in a meeting without any expectation of preparation. We instead provided a platform for remote participation and open discussion, whilst offering compensation for time and contribution. Due to the short notice provided, or for reasons of digital exclusion, it is possible that some groups might not have been able to participate. However, the report will be shared with all participants (subsequent to the necessary permissions), and further co-production work is planned with patients, the public and third sector advocacy groups to support ongoing communication to a wider audience about the current evidence base for MCED tests.

## 7 IMPACT AND LEARNING

This systematic review has highlighted significant gaps in the evidence for MCED tests as they might be applied in a UK context. We have identified and reported on a wide variety of research needs, some of which are likely to be addressed in UK projects that are already planned or underway. The relationships fostered as part of our review and consultation work, with both stakeholder organisations and patients and the public, have also yielded opportunities for involvement in all future UK research projects of which we are currently aware.

This review has already had direct impact on three planned or early stage MCED test research projects. It is directly informing a project supported by the UK National Screening Committee (UK NSC) which has recently commissioned an evidence review of MCEDs (led by Bethany Shinkins, University of Warwick and Jason Oke, University of Oxford). These findings will be incorporated into the UK NCS evidence review and supplemented by a review of the methodological literature, with the overarching aim of identifying issues uniquely related to MCEDs and developing criteria for the UK NSC to use in the evaluation of MCEDs tests in a screening context. Members of the research team for this project will contribute to this work wherever possible and appropriate. This review is also being used as foundation work to underpin a project being led by the Centre for Health Economics (CHE) in partnership with the Centre for Reviews and Dissemination, University of York, that will support the understanding of the economic impacts of these tests and technologies. Finally, it has already contributed to planning and is informing the design of an HTA project NHS-Galleri RCT^60^.

To proactively communicate the findings of this review, we are now beginning to work with partners Healthwatch,^27^ Involve Hull,^28^ Yorkshire and Humber Cancer Alliance^29^ and our combined wider networks, to co-produce and develop dissemination resources explaining the current state of the evidence in a form accessible for target audiences

Finally, in view of the rapidly developing evidence in this field, enabling prompt public access to the findings of this review will maximise its impact, particularly for non-UK based projects. For example, the review could be valuable in informing the USA National Cancer Institute Vanguard Study on Multi-Cancer Detection study^129, 130^ which is due to begin a pilot study in 2024.

## 8 DISCUSSION

In terms of accuracy, the use of an MCED test as a screening tool in a generally healthy, asymptomatic population, alongside existing cancer screening programmes, requires a high specificity and high accuracy of the predicted CSO. It also requires reasonable sensitivity to detect early-stage disease so that the benefits of earlier diagnosis, where treatment options exist, can be realised, compared with a later-stage diagnosis where symptoms may be present and treatment options may be more limited. A potential advantage of MCED tests would be if they are able to detect cancer earlier, with test results used to intervene with therapies with intent to cure, thus positively impacting on mortality and HRQoL.

Limited evidence is available on the potential for early detection of treatable cancers, and the consequences of introducing screening with an MCED test in a UK population. In particular there is some concern that MCED tests may tend to identify cancers with an increased risk of late recurrence, meaning that even if a patient is initially diagnosed as having early stage cancer and treated, the disease may later recur leading to no improvement in survival.^18, 131, 132^

However, there is ongoing debate about whether detecting cancer at an earlier stage always leads to an improvement in mortality. The recent UK Collaborative Trial of Ovarian Cancer Screening (UKCTOCS) trial,^133^ which randomised women aged 50 to 74 years to annual multimodal screening or transvaginal ultrasound screening or no screening, found a significant reduction in the incidence of late-stage ovarian cancer with screening, but no benefit in terms of mortality. This is not an isolated case; the Cluster Randomized Trial of PSA Testing for Prostate Cancer (CAP) trial^134^ made the same observation for prostate cancer and this has been discussed widely in the literature.^14, 18, 132^

### 8.1 Summary of findings

This review summarised existing evidence on MCED test performance and patient-relevant outcomes. We included 36 studies evaluating 13 technologies that reported relevant outcomes for this review, and risk of bias assessment identified substantial concerns with the included studies. We found no completed RCTs or prospective cohort studies carried out in a UK asymptomatic population reporting accuracy measures, morbidity or mortality outcomes. Limited evidence on acceptability to patients and the potential impact on health services was found, although none in a UK setting. Ongoing studies may provide relevant evidence within the target screening population once their findings are published.

Of the 30 completed studies reporting results, only SYMPLIFY^58^ (evaluating the Galleri test) was conducted in the UK (England and Wales), although participants recruited had been referred for urgent investigation of possible cancer and are therefore not reflective of the population of asymptomatic individuals aged 50 to 79 years, which was the target population of this review. Prospective cohort studies that recruited asymptomatic individuals outside the UK included PATHFINDER^30^ (Galleri test, USA), DETECT-A^6^ (CancerSEEK, USA), K-DETEK^8^ (SPOT-MAS, Vietnam), RESOLUTE^9^ (TruCheck, India), PPCS^10^ (CDA, China), and Suzuki et al.^66^ (AICS, Japan) (see Appendix 5 for further study details). Of these, studies recruiting participants deemed to be most similar to the target population for this review (in terms of age and sex) were PATHFINDER,^30^ K-DETEK^8^ and PPCS,^10^ although the studies’ location affects generalisability of results to UK clinical practice due to potential differences in cancer prevalence, healthcare systems and population ethnicity.

Test accuracy and number of overall cancers detected will be different if included participants are at high risk of cancer (e.g., in a population already being investigated due to symptoms), asymptomatic with or without risk factors for cancer, or whether they are already known to have cancer (such as in case-control studies). The length of prospective follow-up and extent of further diagnostic investigations conducted for all participants, with or without a positive signal on the MCED test, will also impact on the total number of cancers diagnosed. Prospective follow-up within the included cohort studies ranged from six months to six years, and total numbers of cancers diagnosed were relatively low, impacting on MCED test accuracy estimates.

All currently available MCED tests had high specificity (>96%), an essential requirement of an MCED test to correctly classify people who do not have cancer. Diagnostic test sensitivity is inversely proportional to specificity, therefore an MCED tests with high specificity may have lower sensitivity. Sensitivity of the MCED tests was variable and influenced by study population, study design, reference standard test used and length of follow-up. Sensitivity also varied by cancer stage; generally, MCED tests had lower sensitivity to detect earlier stage cancers (Stage I-II) compared with later stage cancers (Stage III-IV). Where reported, accuracy of CSO was variable, ranging from 67.7% to 70% in case-control studies of CancerSEEK and SPOT-MAS to 85% to 90% in cohort studies of Galleri (Table 4).

The sensitivity of most of the MCED tests to detect solid tumour cancers without a current screening programme in the UK (excluding lung cancer) was higher than their sensitivity to detect cancers with a current screening programme in the UK, except for CDA and in one of the Galleri studies (Table 6). Similar results were found when lung cancer is considered to be covered by existing screening programmes; except for the Galleri test, since the sensitivity of the Galleri test to detect lung cancer is higher than its overall sensitivity. The sensitivity of the MCED tests to detect haematological malignancies was around 50%, although not all of the MCED tests claim to be able to detect these cancers.

The probability that an individual who receives a positive cancer signal has cancer (PPV) from the three cohort studies recruiting asymptomatic participants with ages similar to the target population of this review (and not focusing exclusively on women) ranged from 7.8% for CDA,^10^ to 60.0% for SPOT-MAS^8^ although 95% CIs were wide, so there is considerable uncertainty in these estimates (Table 4). The probability that an individual who receives a negative test result does not have cancer (NPV) ranged from 98.5% for Galleri^30^ to 100% for SPOT-MAS^8^ (Table 4). However, PPV and NPV values are directly related to the prevalence of the disease in the population being tested; PPV will increase and NPV will decrease with increasing prevalence. Prevalence of different cancers is variable across countries (due to ethnicity, risk factors, and healthcare system differences, among others) meaning these results are unlikely to be directly relevant to the UK screening population. The lack of a perfect reference test for all types of cancer may have led to some FP being inaccurately classified (e.g., if tumour too small to be detected by imaging), which may also bias PPV results.The lack of a reference standard test that can be applied to all study participants with negative test results, further limits interpretation.

No important differences in test accuracy by age, sex or ethnicity were observed for Galleri or CancerSEEK; however, studies of these tests recruited a majority of participants from White backgrounds, so results may not be applicable to other ethnic subgroups. No subgroup results were available for the SPOT-MAS, TruCheck, CDA and AICS tests and no subgroup results were available by socio-economic status for any of the MCED tests included in the review.

Limited results for patient relevant outcomes were reported for Galleri and CancerSEEK, and these are unlikely to reflect the target population of asymptomatic individuals aged 50 to 79 years old in a UK setting. Mortality data were available for a very small number of participants, mostly from case reports with follow-up of up to four years post cancer diagnosis. No adverse events were reported for either test; however, for the earlier version of the GRAIL MCED test, time to diagnostic resolution was shorter for those with a TP result compared to a FP result and over 90% of participants with FP results required further imaging tests.

No increase in anxiety levels across participants was reported at different stages of the PATHFINDER study (Galleri test).^30^ This does not, however, rule out that individual participants may have experienced substantial increases in anxiety whilst awaiting test results, and while awaiting diagnostic resolution particularly for those with a positive signal on the MCED test, as anxiety was not measured at these key times..

An additional seven MCED technologies which were at an unclear stage of development and did not appear to be currently available for use were included in the review. Most were evaluated in case-control studies and did not report relevant outcome data for the target population of interest. Many other blood-based MCED technologies which appeared to be at an early stage of development were identified but excluded from the review. These MCED tests and technologies may undergo further development and modification and become available for use in the future.

### 8.2 Strengths and limitations

The literature review was undertaken using systematic methods, reducing the potential for errors and bias. Comprehensive searches were undertaken to identify relevant evidence, including searches of manufacturers’ websites, which identified recent emerging findings from the included studies in conference posters and presentations; this was an important process in such a fast-moving field. The inclusion criteria were clearly defined in advance and full texts were assessed against the inclusion criteria by at least two experienced reviewers. The validity and applicability of the included studies were assessed using an appropriate quality assessment tool for diagnostic accuracy studies. A data extraction tool was developed and piloted; data extraction and validity assessment processes were independently checked for accuracy.

The systematic review was conducted by an independent team of experienced reviewers, statisticians, and information specialists, who were free of potential conflicts of interest. The project benefited from stakeholder input from a range of independent content experts, healthcare professionals, and patient and public representatives, which strengthened the protocol, and the presentation and interpretation of findings in the final report.

The review was limited by weaknesses in the evidence base. There were no completed RCTs identified for any of the MCED tests. Only one study^30^ (of the Galleri test), recruited individuals aged over 50 years without a clinical suspicion of cancer. However, this study was conducted in the USA, therefore, participants and results may not be representative of the UK screening population of interest in this review. Most studies were considered to have a high overall risk of bias, in addition to concerns regarding applicability. The variability in test specifications, study designs and included populations meant that meta-analysis was not appropriate.

The aim of this project was to identify available MCED tests for population-based screening, rather than to review all MCED technologies. However, reporting of many of the identified studies and technologies was limited, adding to the complexity of the study selection process; it was often difficult to determine the stage of development of technologies and whether studies were reporting results for tests at an early stage of development, or assessing a final or near-final version of the test that could be used for screening. In addition, the limited reporting made it difficult to assess the risk of bias and applicability of some of the included studies.

In addition to our review of blood-based MCED tests that are currently available for use, we have included evidence on technologies for which it is unclear whether they are fully developed tests, and presented a non-exhaustive list of technologies at a very early stage of development.

A recent review^135^ (published after our pre-specified search date) of MCED technologies identified 20 studies across various phases of development including four studies not identified by our search strategy. As these four studies all described tests at an early stage of development, which would not have been eligible for inclusion in our review, this serves as reassurance that our search terms were sufficiently broad to capture the most relevant records.

### 8.3 Implications for future research

It is important that the principles that underpin existing disease screening programmes in the NHS are also applied to MCED tests. It is essential that appropriately designed studies assess the natural history of early-stage cancers detected by MCED tests in healthy individuals at different ages, before MCED tests are introduced into a screening programme. Such studies should be designed to enable the development of decision models to direct clinical treatment towards those asymptomatic patients most likely to benefit from, and least likely to be harmed by, treatments.

The most promising and studied blood-based MCED tests are based on detecting cancer-related alterations in cfDNA. However, concentrations of cfDNA are relatively low at early cancer stages so it is unclear whether a simple blood draw would ever contain cfDNA in sufficient amounts to detect very small tumours.^7, 136^ Data collection from large RCTs is needed to evaluate the ability of currently available MCED tests to detect early stage cancers and whether acting on a positive MCED test improves mortality.^14^ The NHS-Galleri RCT^60^ is being conducted in an asymptomatic UK population aged 50 to 77, but its primary objective is to evaluate whether there is a significant reduction in incidence of advanced stage cancer (stages III-IV) in the intervention arm compared to control, three years post-randomisation. Mortality outcomes will also be collected as secondary outcomes. Sampling of participants for the NHS-Galleri RCT aims to ensure that the recruited sample is representative of the wider population in terms of age and socioeconomic status, and that sufficient numbers are recruited from groups typically under-represented in clinical trials, such as those from ethnic minority backgrounds.^137^ As such, when fully reported, data from this trial may provide high quality, direct evidence on the efficacy of this MCED test in a UK screening context.

However, the impact of MCED tests on NHS services including the practicalities of implementing MCED tests is currently unknown, but likely to be substantial. Further research is needed on the resource implications, risk of over-treatment and cost-effectiveness of implementing MCED tests for screening in the NHS.

The National Cancer Institute (USA) is launching the Vanguard Study on Multi-Cancer Detection which will begin enrolling healthy people aged 45 to 70 in a four-year pilot study from 2024 to assess the feasibility of a study to evaluate MCED tests.^129, 130^ Conclusions from the pilot study will inform the decision of whether to launch a longer term RCT, which may compare more than one MCED test with standard of care screening. Should this RCT go ahead with mortality and HRQoL outcomes, as well as assessment of the number and type of diagnostic workups needed after a positive test, and potential harms arising from the workups themselves, this would provide important information on the comparability of different tests. However, this study would be carried out in a US context, which has key differences to the UK (e.g., different population characteristics, cancer prevalence, health care system, and existing screening programmes) so its potential generalisability to the NHS is unclear.

Studies that capture patient-relevant outcomes are also required. A longitudinal observational design with a nested qualitative study to evaluate the psychological impact of the Galleri test (sIG(n)al)^138^ is embedded in the NHS-Galleri trial. Participants who have a cancer signal detected (expected number approximately 700) will be sent questionnaires at various timepoints to evaluate outcomes including anxiety, the psychological consequences of screening, reassurance/concern about the test result, understanding of results and help/health-seeking behaviour. Depending on response rates this may provide valuable insight into these important outcomes, although data will be collected within the context of a clinical trial and no translation of questionnaires will be available, which may lead to fewer responses from participants from diverse backgrounds. In addition, participants with a negative MCED test result will not be studied, which may limit the applicability of findings to the target UK screening population. Studies including participants representative of the UK screening population and with sufficiently long follow-up should be carried out to better understand the potential psychological and behavioural impacts of MCED tests in practice, both in those with positive and negative test results.

The setting up of a registry to collect and evaluate real-world evidence on MCED tests, to record the diagnostic pathway and patterns of care following a test and impact on relevant patient outcomes such as mortality, morbidity, adverse events and HRQoL, has also been suggested.^16^

As more MCED tests become available, it is important that appropriate studies are carried out and reported in sufficient detail for their diagnostic accuracy, feasibility and acceptability to be evaluated. Given the large variability in the number of cancers detected by different MCED tests, and the differences in accuracy for different cancers and stages within and between tests, comparison of the accuracy, costs and benefits of the different MCED tests against each other in a UK screening context would be valuable.

## 9 CONCLUSIONS

This comprehensive review summarised the existing evidence on 13 tests and technologies that aim to detect multiple cancers for screening of healthy populations. Although current available evidence does not support strong conclusions, studies reported promising accuracy evidence despite limitations. Additional studies are ongoing or planned which will address some current limitations.

RCTs with sufficiently long follow-up, reporting outcomes that are directly relevant to patients, such as mortality/morbidity, safety, HRQoL and impact of (true and false) positive and (false) negative results on the health system, are needed and some are planned or underway. Given the potential false reassurance of a false negative test result, studies with sufficiently long follow-up for detection of emerging cancers in those testing negative, and evaluating the impacts of a negative test on compliance with existing screening programmes, are essential for the proper assessment of the possible negative impacts of each test.

Given the limitations of current treatment strategies for some cancers, even if detected early, an MCED test that more accurately detects fewer, but more treatable cancers, and for which there is currently no national screening programme, may have greater overall benefits than the use of a test that detects many cancers with no effective treatment, or those already covered by existing screening programmes. No completed or ongoing study was found comparing the potential benefits to individuals or healthcare systems of different MCED tests against each other.

Decisions on implementation of MCED tests for screening in an asymptomatic population need to be underpinned by solid evidence, preferably RCTs carried out in a relevant population, setting, and with an appropriate length follow-up, so that an evidence-based evaluation can be carried out. At the moment, this evidence is lacking for all the tests evaluated in this report. Careful consideration is needed of which, if any, of the MCED tests currently available should be used, taking account of the current paucity of high-quality, relevant evidence on their accuracy, acceptability, cost/utility benefits and impact on the NHS.

## Data Availability

This study did not generate any new data as it used existing sources, and all data is contained within the manuscript. Any queries should be addressed to the corresponding author.

## Abbreviations

AICS: AminoIndex Cancer Screening
CA125: Cancer antigen 125
CAP: Cluster Randomized Trial of PSA Testing for Prostate Cancer
CCGA: Circulating Cell-free Genome Atlas
CDA: Cancer Differentiation Analysis
CDSR: Cochrane Database of Systematic Reviews
cfDNA: Cell-free DNA
CI: Confidence interval
CSO: Cancer Signal Origin
CT: Computed tomography
CTC: Circulating tumour cells
ctDNA: Circulating tumour DNA
DARE: Database of Abstracts of Reviews of Effects
DELFI: DNA Evaluation of Fragments for Early Interception
DNA: Deoxyribonucleic acid
EDI: Equality, Diversity and Inclusion
FN: False negative
FP: False positive
HRQoL: Health-related quality of life
HTA: Health Technology Assessment
INAHTA: International Health Technology Assessment database
MCED: Multi-cancer early detection
NA: Not applicable
NHS: National Health Service
NPV: Negative predictive value
PET-CT: Positron emission tomography-computed tomography
PPCS: Prospective Population-based Cohort Study
PPV: Positive predictive value
PRISMA: Preferred Reporting Items for Systematic Reviews and Meta-Analyses
PROMIS: Patient-Reported Outcomes Measurement Information System
QUADAS: Quality Assessment of Diagnostic Accuracy Studies
RCT: Randomised controlled trial
SPOT-MAS: Screening for the Presence Of Tumour by Methylation And Size
TN: True negative
TP: True positive
UKCTOCS: UK Collaborative Trial of Ovarian Cancer Screening

## ADDITIONAL INFORMATION

### Acknowledgements

The authors would like to thank Helen Fulbright, Information Specialist, for peer reviewing the MEDLINE search strategy, Professor Bob Philips (Professor of Paediatric Oncology at Hull York Medical School and the Centre for Reviews and Dissemination) for assistance with identifying content experts, and also Leo Stevens (Head of Communications and Engagement at Humber and Yorkshire Cancer Alliance) and Helen Roberts (Patient and Public Involvement Coordinator at University of Hull/Hull York Medical School) for their invaluable help in contacting PPI representatives.

The authors would also like to thank the following content experts and patient and public representatives for their valuable comments and feedback throughout the development of this report.

*Content experts contributing to the protocol and report*

Bethany Shinkins, Professor of Health Economics, Diagnosis and Screening, University of Warwick. Joanne Cairns, Research Fellow, Yorkshire Cancer Research Career Development Fellowship, Cancer Awareness, Screening and Diagnostic Pathways (CASP) Research Group, Hull York Medical School (HYMS).

Matthew Callister, Consultant Respiratory Physician, Leeds Teaching Hospitals.

Rachel Iveson, Cancer Diagnosis and Innovation Lead, Humber and North Yorkshire Cancer Alliance.

Una Macleod, Professor of Primary care Medicine, HYMS and Transforming Cancer Outcomes in Yorkshire.

*Patient and public representatives contributing to protocol or report*

Sian Balsom, Manager, Health Watch York.

Ron Stamp, Public representative; former Clinical Biochemist and NHS Research Director.

Linda Wolstenholme, Patient representative, Humber and North Yorkshire Cancer Alliance.

*Organisations/projects contributing to patient and public representative consultation exercise*

Healthwatch York

Humber and Yorkshire Cancer Alliance Transforming Cancer Outcomes in Yorkshire Project

Involve Hull, University of Hull/Hull York Medical School

*Individuals contributing to patient and public representative consultation exercise including, but not limited to:*

Linda Wolstenholme, Sue Tucker, Allyson Kent, June Pitt, Manoj Mistry, Arif Hoque, Lynne Wright, Sally Osgerby, Margaret Ogden, Arthur Spur, Howard Lester.

### CRediT Statement

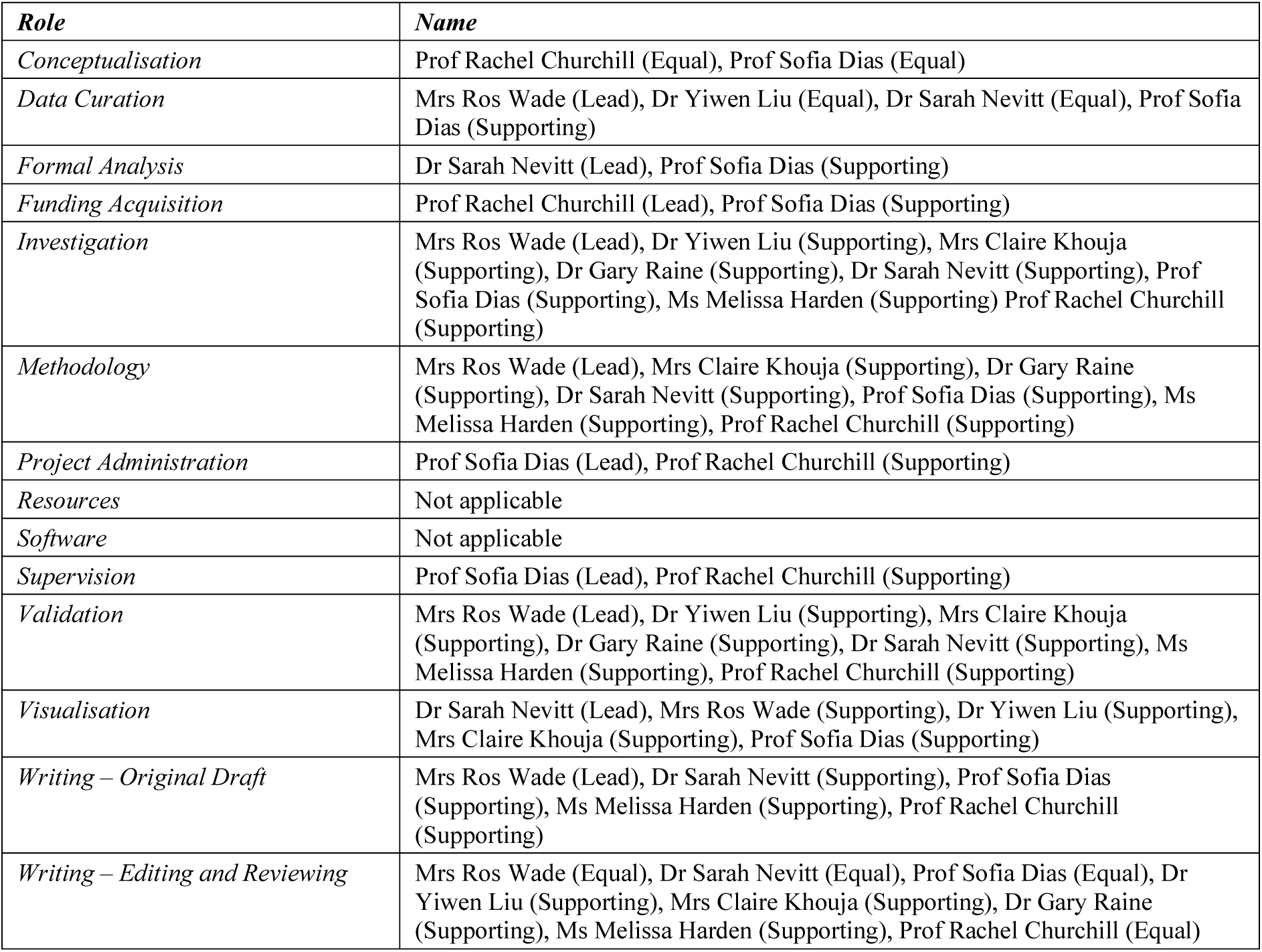

### Contributions of authors

Ros Wade and Sarah Nevitt contributed to the protocol, study selection, data extraction, study validity assessment and synthesis of the included studies. They also contributed to the interpretation of the results and the writing of the report.

Yiwen Liu contributed to study selection, data extraction, study validity assessment and synthesis of the included studies. She also contributed to the interpretation of the results and the writing of the report.

Melissa Harden contributed to the protocol, developed search strategies, conducted a range of searches to locate studies and wrote sections of the report.

Claire Khouja and Gary Raine contributed to the protocol, study selection, data extraction, study validity assessment and writing of the report.

Rachel Churchill contributed to the protocol, interpretation of the results, writing of the report and stakeholder engagement.

Sofia Dias contributed to the protocol, study selection, data extraction, validity assessment and synthesis of the included studies. She also contributed to the interpretation of the results and the writing of the report. Sofia had overall responsibility for the project.

All authors read and approved the final version of the report.

### Ethics statement

This review did not involve the collection or analysis of any data that was not included in previously published research in the public domain. Therefore, no ethical approval was required.

### Information Governance Statement

This is a systematic literature review and therefore, the current research did not handle any personal information.

### Disclaimer

This publication presents independent research commissioned by the National Institute for Health and Care Research (NIHR). The views and opinions expressed by authors in this publication are those of the authors and do not necessarily reflect those of the NHS, the NIHR, the NIHR Coordinating Centre, the NIHR Evidence Synthesis programme or the Department of Health and Social Care.

## APPENDICES

## APPENDIX 1. PRISMA CHECKLIST

**Table 9.**
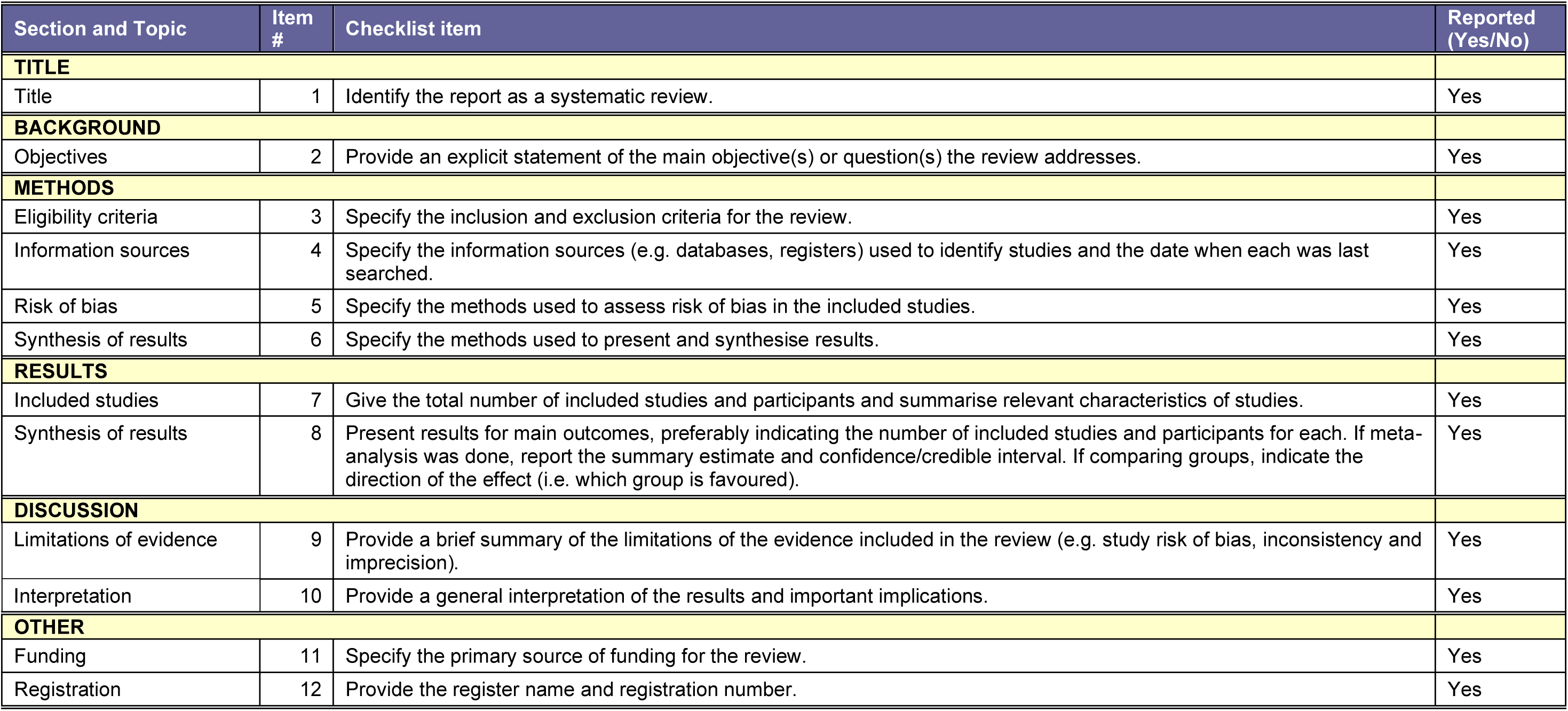
PRISMA 2020 abstract checklist.

| Section and Topic | Item # | Checklist item | Reported (Yes/No) |
| --- | --- | --- | --- |
| **TITLE** |  |  |  |
| Title | 1 | Identify the report as a systematic review. | Yes |
| **BACKGROUND** |  |  |  |
| Objectives | 2 | Provide an explicit statement of the main objective(s) or question(s) the review addresses. | Yes |
| **METHODS** |  |  |  |
| Eligibility criteria | 3 | Specify the inclusion and exclusion criteria for the review. | Yes |
| Information sources | 4 | Specify the information sources (e.g. databases, registers) used to identify studies and the date when each was last searched. | Yes |
| Risk of bias | 5 | Specify the methods used to assess risk of bias in the included studies. | Yes |
| Synthesis of results | 6 | Specify the methods used to present and synthesise results. | Yes |
| **RESULTS** |  |  |  |
| Included studies | 7 | Give the total number of included studies and participants and summarise relevant characteristics of studies. | Yes |
| Synthesis of results | 8 | Present results for main outcomes, preferably indicating the number of included studies and participants for each. If meta-analysis was done, report the summary estimate and confidence/credible interval. If comparing groups, indicate the direction of the effect (i.e. which group is favoured). | Yes |
| **DISCUSSION** |  |  |  |
| Limitations of evidence | 9 | Provide a brief summary of the limitations of the evidence included in the review (e.g. study risk of bias, inconsistency and imprecision). | Yes |
| Interpretation | 10 | Provide a general interpretation of the results and important implications. | Yes |
| **OTHER** |  |  |  |
| Funding | 11 | Specify the primary source of funding for the review. | Yes |
| Registration | 12 | Provide the register name and registration number. | Yes |

**Table 10.** PRISMA 2020 checklist.

| Section and Topic | Item # | Checklist item | Location where item is reported |
| --- | --- | --- | --- |
| **TITLE** |  |  |  |
| Title | 1 | Identify the report as a systematic review. | 1 |
| **ABSTRACT** |  |  |  |
| Abstract | 2 | See the PRISMA 2020 for Abstracts checklist. | 4-5 |
| **INTRODUCTION** |  |  |  |
| Rationale | 3 | Describe the rationale for the review in the context of existing knowledge. | 19 |
| Objectives | 4 | Provide an explicit statement of the objective(s) or question(s) the review addresses. | 19 |
| **METHODS 4-** |  |  |  |
| Eligibility criteria | 5 | Specify the inclusion and exclusion criteria for the review and how studies were grouped for the syntheses. | 19-21 |
| Information sources | 6 | Specify all databases, registers, websites, organisations, reference lists and other sources searched or consulted to identify studies. Specify the date when each source was last searched or consulted. | 21-22 |
| Search strategy | 7 | Present the full search strategies for all databases, registers and websites, including any filters and limits used. | 21-22, Appendix 2 |
| Selection process | 8 | Specify the methods used to decide whether a study met the inclusion criteria of the review, including how many reviewers screened each record and each report retrieved, whether they worked independently, and if applicable, details of automation tools used in the process. | 22 |
| Data collection process | 9 | Specify the methods used to collect data from reports, including how many reviewers collected data from each report, whether they worked independently, any processes for obtaining or confirming data from study investigators, and if applicable, details of automation tools used in the process. | 22-24 |
| Data items | 10a | List and define all outcomes for which data were sought. Specify whether all results that were compatible with each outcome domain in each study were sought (e.g. for all measures, time points, analyses), and if not, the methods used to decide which results to collect. | 23-24 |
|  | 10b | List and define all other variables for which data were sought (e.g. participant and intervention characteristics, funding sources). Describe any assumptions made about any missing or unclear information. | 22-23 |
| Study risk of bias assessment | 11 | Specify the methods used to assess risk of bias in the included studies, including details of the tool(s) used, how many reviewers assessed each study and whether they worked independently, and if applicable, details of automation tools used in the process. | 24 |
| Effect measures | 12 | Specify for each outcome the effect measure(s) (e.g. risk ratio, mean difference) used in the synthesis or presentation of results. | 23 |
| Synthesis methods | 13a | Describe the processes used to decide which studies were eligible for each synthesis (e.g. tabulating the study intervention characteristics and comparing against the planned groups for each synthesis (item #5)). | 24 |
|  | 13b | Describe any methods required to prepare the data for presentation or synthesis, such as handling of missing summary statistics, or data conversions. | 23-24 |
|  | 13c | Describe any methods used to tabulate or visually display results of individual studies and syntheses. | 24 |
|  | 13d | Describe any methods used to synthesize results and provide a rationale for the choice(s). If meta-analysis was performed, describe the model(s), method(s) to identify the presence and extent of statistical heterogeneity, and software package(s) used. | 24 |
|  | 13e | Describe any methods used to explore possible causes of heterogeneity among study results (e.g. subgroup analysis, meta-regression). | 24 |
|  | 13f | Describe any sensitivity analyses conducted to assess robustness of the synthesized results. | N/A |
| Reporting bias assessment | 14 | Describe any methods used to assess risk of bias due to missing results in a synthesis (arising from reporting biases). | N/A (no meta-analysis was done) |
| Certainty assessment | 15 | Describe any methods used to assess certainty (or confidence) in the body of evidence for an outcome. | 24 |
| **RESULTS** |  |  |  |
| Study selection | 16a | Describe the results of the search and selection process, from the number of records identified in the search to the number of studies included in the review, ideally using a flow diagram. | 25-27, Figure 1 |
|  | 16b | Cite studies that might appear to meet the inclusion criteria, but which were excluded, and explain why they were excluded. | 27, 52-53, Table 8, Appendix 2 |
| Study characteristics | 17 | Cite each included study and present its characteristics. | 30-32, Tables 1-2, Appendix 4 Table 10 |
| Risk of bias in studies | 18 | Present assessments of risk of bias for each included study. | 32-34, Figure 2, Table 3, Appendix 4 Table 11 |
| Results of individual studies | 19 | For all outcomes, present, for each study: (a) summary statistics for each group (where appropriate) and (b) an effect estimate and its precision (e.g. confidence/credible interval), ideally using structured tables or plots. | Tables 4-6, Figures 3-4 |
| Results of syntheses | 20a | For each synthesis, briefly summarise the characteristics and risk of bias among contributing studies. | Table 2, Appendix 4 Table 10 |
|  | 20b | Present results of all statistical syntheses conducted. If meta-analysis was done, present for each the summary estimate and its precision (e.g. confidence/credible interval) and measures of statistical heterogeneity. If comparing groups, describe the direction of the effect. | N/A (no meta-analysis was done, narrative synthesis results can be found on pages 34-44) |
|  | 20c | Present results of all investigations of possible causes of heterogeneity among study results. | 47 (subgroup analyses), Appendix 5 Tables 13-16, Appendix 6 Tables 17-18 |
|  | 20d | Present results of all sensitivity analyses conducted to assess the robustness of the synthesized results. | N/A |
| Reporting biases | 21 | Present assessments of risk of bias due to missing results (arising from reporting biases) for each synthesis assessed. | N/A (no meta-analysis was done) |
| Certainty of evidence | 22 | Present assessments of certainty (or confidence) in the body of evidence for each outcome assessed. | 34-36, 67 |
| **DISCUSSION** |  |  |  |
| Discussion | 23a | Provide a general interpretation of the results in the context of other evidence. | 63-66 |
|  | 23b | Discuss any limitations of the evidence included in the review. | 67 |
|  | 23c | Discuss any limitations of the review processes used. | 67 |
|  | 23d | Discuss implications of the results for practice, policy, and future research. | 67-69 |
| **OTHER INFORMATION** |  |  |  |
| Registration and protocol | 24a | Provide registration information for the review, including register name and registration number, or state that the review was not registered. | 5 |
|  | 24b | Indicate where the review protocol can be accessed, or state that a protocol was not prepared. | N/A (The protocol was prepared but not published). |
|  | 24c | Describe and explain any amendments to information provided at registration or in the protocol. | 24 (Additional analyses from stakeholder), 55-61 (additional stakeholder engagement after protocol stage) |
| Support | 25 | Describe sources of financial or non-financial support for the review, and the role of the funders or sponsors in the review. | 2-3, 5 |
| Competing interests | 26 | Declare any competing interests of review authors. | 2 |
| Availability of data, code and other materials | 27 | Report which of the following are publicly available and where they can be found: template data collection forms; data extracted from included studies; data used for all analyses; analytic code; any other materials used in the review. | All data extracted are provided in Tables 4-6, and Appendices 3-6 |

## APPENDIX 2. SEARCH STRATEGIES

### 1. Database searches

**MEDLINE ALL**

via Ovid http://ovidsp.ovid.com/

Date range: 1946 to September 13, 2023

Date searched: 14^th^ September 2023

Records retrieved: 2280

1. Neoplasms/ (504101)
2. ((cancer$ or neoplas$ or tumour$ or tumor$ or carcinoma$ or oncolog$ or malignan$ or precancer$) adj6 (multiple$ or many or several or numerous or various or varied or miscellaneous or mixed or diverse or different)).ti,ab. (481091)
3. (multicancer$ or multi-cancer$ or multitumo?r$ or multi-tumo?r$ or pan-cancer$ or pancancer$ or pan-tumo?r$ or pantumo?r$ or cross-cancer$ or crosscancer$ or cross-tumo?r$ or crosstumo?r$).ti,ab. (4793)
4. ((cancer$ or neoplas$ or tumour$ or tumor$ or carcinoma$ or oncolog$ or malignan$ or precancer$) adj3 (type or types)).ti,ab. (174370)
5. 1 or 2 or 3 or 4 (993603)
6. Liquid Biopsy/ (2716)
7. ((liquid$ or fluid$ or biofluid$ or bio-fluid$) adj3 biops$).ti,ab. (8392)
8. 6 or 7 (8976)
9. Biopsy/ or Biopsy, Fine-Needle/ (203652)
10. exp Blood/ (1196437)
11. 9 and 10 (9122)
12. ((blood or h?ematolog$ or plasma or serum) adj3 biops$).ti,ab. (5561)
13. 11 or 12 (14504)
14. Hematologic Tests/ (10175)
15. ((blood or h?ematolog$ or plasma or serum) adj2 (test or tests or testing or tested or assay$)).ti,ab. (79621)
16. 14 or 15 (88397)
17. Multiomics/ (822)
18. ((multiomic$ or multi-omic$ or panomic$ or pan-omic$ or integrative omic$) adj4 (test or tests or tested or testing or assay$ or biops$)).ti,ab. (114)
19. 17 or 18 (924)
20. ((Multi-analyte$ or multianalyte$) adj4 (detect$ or screen$ or test or tests or tested or testing or assay$ or biops$)).ti,ab. (571)
21. 8 or 13 or 16 or 19 or 20 (112181)
22. 5 and 21 (5730)
23. Mass Screening/ (116511)
24. Diagnostic Screening Programs/ (156)
25. early diagnosis/ (30350)
26. "Early Detection of Cancer"/ (38071)
27. (screen$ or detect$).ti. (656777)
28. ((early or earlystage or earli$ or first or initial or timely) adj3 (screen$ or detect$ or diagnos$ or test or tests or testing or tested)).ti,ab. (434798)
29. (screen$ adj3 (test$ or tool$ or method$ or strateg$ or modalit$ or technolog$ or program$ or service$ or policy or policies or guideline$ or population$)).ti,ab. (201334)
30. 23 or 24 or 25 or 26 or 27 or 28 or 29 (1188487)
31. 22 and 30 (1886)
32. ((cancer$ or neoplas$ or tumour$ or tumor$ or carcinoma$ or oncolog$ or malignan$ or precancer$) adj6 (multiple$ or many or several or numerous or various or varied or miscellaneous or mixed or diverse or different) adj6 (screen$ or detect$)).ti,ab. (12043)
33. ((cancer$ or neoplas$ or tumour$ or tumor$ or carcinoma$ or oncolog$ or malignan$ or precancer$) adj6 (type or types) adj6 (screen$ or detect$)).ti,ab. (4420)
34. 32 or 33 (15191)
35. 21 and 34 (606)
36. 31 or 35 (2018)
37. (((multi-cancer$ or multicancer$ or multi-tumo?r$ or multitumo?r$) adj6 (detect$ or screen$ or test or tests or tested or testing or assay$)) or MCED or MCDBT).ti,ab. (155)
38. ((multiple cancer$ or multiple tumo?r$) adj6 (detect$ or screen$ or test or tests or tested or testing or assay$)).ti,ab. (523)
39. ((pan-cancer$ or pancancer$ or pan-tumo?r$ or pantumo?r$) adj6 (detect$ or screen$ or test or tests or tested or testing or assay$)).ti,ab. (202)
40. ((cross-cancer$ or crosscancer$ or cross-tumo?r$ or crosstumo?r$) adj6 (detect$ or screen$ or test or tests or tested or testing or assay$)).ti,ab. (5)
41. ((multi-class cancer$ or multiclass cancer$ or multi-class tumo?r$ or multiclass tumo?r$) adj6 (detect$ or screen$ or test or tests or tested or testing or assay$)).ti,ab. (6)
42. 37 or 38 or 39 or 40 or 41 (869)
43. (Galleri or GalleriTM).mp. (7)
44. PanSEER$.mp. (3)
45. CancerSEEK$.mp. (7)
46. CancerEMC$.mp. (1)
47. (PanTum or PanTumDetect).mp. (3)
48. Epitope-detection in monocytes.mp. (12)
49. CancerRadar$.mp. (0)
50. (IvyGene$ or IvyGeneCORE$).mp. (0)
51. CancerLocator$.mp. (1)
52. CancerDetector$.mp. (1)
53. (EpiPanGI Dx$ or EpiPanGIDx$).mp. (1)
54. OverC.mp. (2)
55. DEEPGEN.mp. (6)
56. Dxcover$.mp. (1)
57. trucheck$.mp. (0)
58. Elypta$.mp. (0)
59. MiRXES$.mp. (6)
60. Freenome$.mp. (1)
61. 43 or 44 or 45 or 46 or 47 or 48 or 49 or 50 or 51 or 52 or 53 or 54 or 55 or 56 or 57 or 58 or 59 or 60 (47)
62. DELFI$.mp. (693)
63. Omni1$.mp. (24)
64. Signal-X$.mp. (48)
65. Harbinger$.mp. (2098)
66. EDIM$.mp. (180)
67. LUNAR$.mp. (4523)
68. MERCURY$.mp. (55377)
69. 62 or 63 or 64 or 65 or 66 or 67 or 68 (62897)
70. 22 and 69 (5)
71. 36 or 42 or 61 or 70 (2835)
72. exp animals/ not humans.sh. (5154669)
73. 71 not 72 (2804)
74. limit 73 to yr="2010 -Current" (2280)

**Key:**

/ = subject heading (MeSH heading)

sh = subject heading (MeSH heading)

exp = exploded subject heading (MeSH heading)

$ = truncation

? = optional wildcard – one or no characters

ti,ab = terms in title or abstract fields

mp = multi-purpose field search – searches several fields including title, original title, abstract, keyword, subject heading word

adj3 = terms within three words of each other (any order)

**Embase**

via Ovid http://ovidsp.ovid.com/

Date range: 1974 to 2023 September 13

Date searched: 14^th^ September 2023

Records retrieved: 5318

1. neoplasm/ (444533)
2. ((cancer$ or neoplas$ or tumour$ or tumor$ or carcinoma$ or oncolog$ or malignan$ or precancer$) adj6 (multiple$ or many or several or numerous or various or varied or miscellaneous or mixed or diverse or different)).ti,ab. (676542)
3. (multicancer$ or multi-cancer$ or multitumo?r$ or multi-tumo?r$ or pan-cancer$ or pancancer$ or pan-tumo?r$ or pantumo?r$ or cross-cancer$ or crosscancer$ or cross-tumo?r$ or crosstumo?r$).ti,ab. (7553)
4. ((cancer$ or neoplas$ or tumour$ or tumor$ or carcinoma$ or oncolog$ or malignan$ or precancer$) adj3 (type or types)).ti,ab. (253298)
5. 1 or 2 or 3 or 4 (1167319)
6. liquid biopsy/ (11133)
7. ((liquid$ or fluid$ or biofluid$ or bio-fluid$) adj3 biops$).ti,ab. (14049)
8. 6 or 7 (16738)
9. biopsy/ (178541)
10. exp blood/ (2566272)
11. 9 and 10 (29880)
12. ((blood or h?ematolog$ or plasma or serum) adj3 biops$).ti,ab. (10439)
13. 11 or 12 (38994)
14. blood examination/ (18548)
15. ((blood or h?ematolog$ or plasma or serum) adj2 (test or tests or testing or tested or assay$)).ti,ab. (127454)
16. 14 or 15 (142450)
17. ((multiomic$ or multi-omic$ or panomic$ or pan-omic$ or integrative omic$) adj4 (test or tests or tested or testing or assay$ or biops$)).ti,ab. (220)
18. ((Multi-analyte$ or multianalyte$) adj4 (detect$ or screen$ or test or tests or tested or testing or assay$ or biops$)).ti,ab. (777)
19. 8 or 13 or 16 or 17 or 18 (195146)
20. 5 and 19 (14344)
21. mass screening/ (61673)
22. cancer screening/ (97647)
23. early cancer diagnosis/ (13662)
24. (screen$ or detect$).ti. (792212)
25. ((early or earlystage or earli$ or first or initial or timely) adj3 (screen$ or detect$ or diagnos$ or test or tests or testing or tested)).ti,ab. (638156)
26. (screen$ adj3 (test$ or tool$ or method$ or strateg$ or modalit$ or technolog$ or program$ or service$ or policy or policies or guideline$ or population$)).ti,ab. (294581)
27. 22 or 23 or 24 or 25 or 26 (1533346)
28. 20 and 27 (4213)
29. ((cancer$ or neoplas$ or tumour$ or tumor$ or carcinoma$ or oncolog$ or malignan$ or precancer$) adj6 (multiple$ or many or several or numerous or various or varied or miscellaneous or mixed or diverse or different) adj6 (screen$ or detect$)).ti,ab. (17272)
30. ((cancer$ or neoplas$ or tumour$ or tumor$ or carcinoma$ or oncolog$ or malignan$ or precancer$) adj6 (type or types) adj6 (screen$ or detect$)).ti,ab. (6375)
31. 29 or 30 (21840)
32. 19 and 31 (1075)
33. 28 or 32 (4496)
34. (((multi-cancer$ or multicancer$ or multi-tumo?r$ or multitumo?r$) adj6 (detect$ or screen$ or test or tests or tested or testing or assay$)) or MCED or MCDBT).ti,ab. (339)
35. ((multiple cancer$ or multiple tumo?r$) adj6 (detect$ or screen$ or test or tests or tested or testing or assay$)).ti,ab. (849)
36. ((pan-cancer$ or pancancer$ or pan-tumo?r$ or pantumo?r$) adj6 (detect$ or screen$ or test or tests or tested or testing or assay$)).ti,ab. (463)
37. ((cross-cancer$ or crosscancer$ or cross-tumo?r$ or crosstumo?r$) adj6 (detect$ or screen$ or test or tests or tested or testing or assay$)).ti,ab. (10)
38. ((multi-class cancer$ or multiclass cancer$ or multi-class tumo?r$ or multiclass tumo?r$) adj6 (detect$ or screen$ or test or tests or tested or testing or assay$)).ti,ab. (10)
39. 34 or 35 or 36 or 37 or 38 (1587)
40. (Galleri or GalleriTM).mp. (28)
41. PanSEER$.mp. (7)
42. CancerSEEK$.mp. (17)
43. CancerEMC$.mp. (1)
44. (PanTum or PanTumDetect).mp. (6)
45. Epitope-detection in monocytes.mp. (18)
46. CancerRadar$.mp. (1)
47. (IvyGene$ or IvyGeneCORE$).mp. (7)
48. CancerLocator$.mp. (1)
49. CancerDetector$.mp. (1)
50. (EpiPanGI Dx$ or EpiPanGIDx$).mp. (2)
51. OverC.mp. (1)
52. DEEPGEN.mp. (13)
53. Dxcover$.mp. (9)
54. trucheck$.mp. (4)
55. Elypta$.mp. (1)
56. MiRXES$.mp. (42)
57. Freenome$.mp. (60)
58. 40 or 41 or 42 or 43 or 44 or 45 or 46 or 47 or 48 or 49 or 50 or 51 or 52 or 53 or 54 or 55 or 56 or 57 (205)
59. DELFI$.mp. (1238)
60. Omni1$.mp. (135)
61. Signal-X$.mp. (1058)
62. Harbinger$.mp. (2991)
63. EDIM$.mp. (265)
64. LUNAR$.mp. (8154)
65. MERCURY$.mp. (72070)
66. 59 or 60 or 61 or 62 or 63 or 64 or 65 (85868)
67. 20 and 66 (21)
68. 33 or 39 or 58 or 67 (6044)
69. (animal/ or animal experiment/ or animal model/ or animal tissue/ or nonhuman/) not exp human/ (6811834)
70. 68 not 69 (5933)
71. limit 70 to yr="2010 -Current" (5318)

**Key:**

/ = subject heading (Emtree heading)

exp = exploded subject heading (Emtree heading)

$ = truncation

? = optional wildcard – one or no characters

ti,ab = terms in title or abstract fields

mp = multi-purpose field search – searches several fields including title, original title, abstract, keyword, subject heading word, candidate terms, device trade name, device manufacturer.

adj3 = terms within three words of each other (any order)

**Cochrane Library**

via Wiley http://onlinelibrary.wiley.com/

**Cochrane Central Register of Controlled Trials (CENTRAL)**: Issue 8 of 12, Aug 2023

Records retrieved: 147

**Cochrane Database of Systematic Reviews (CDSR):** Issue 9 of 12, Sep 2023

Records retrieved: 5

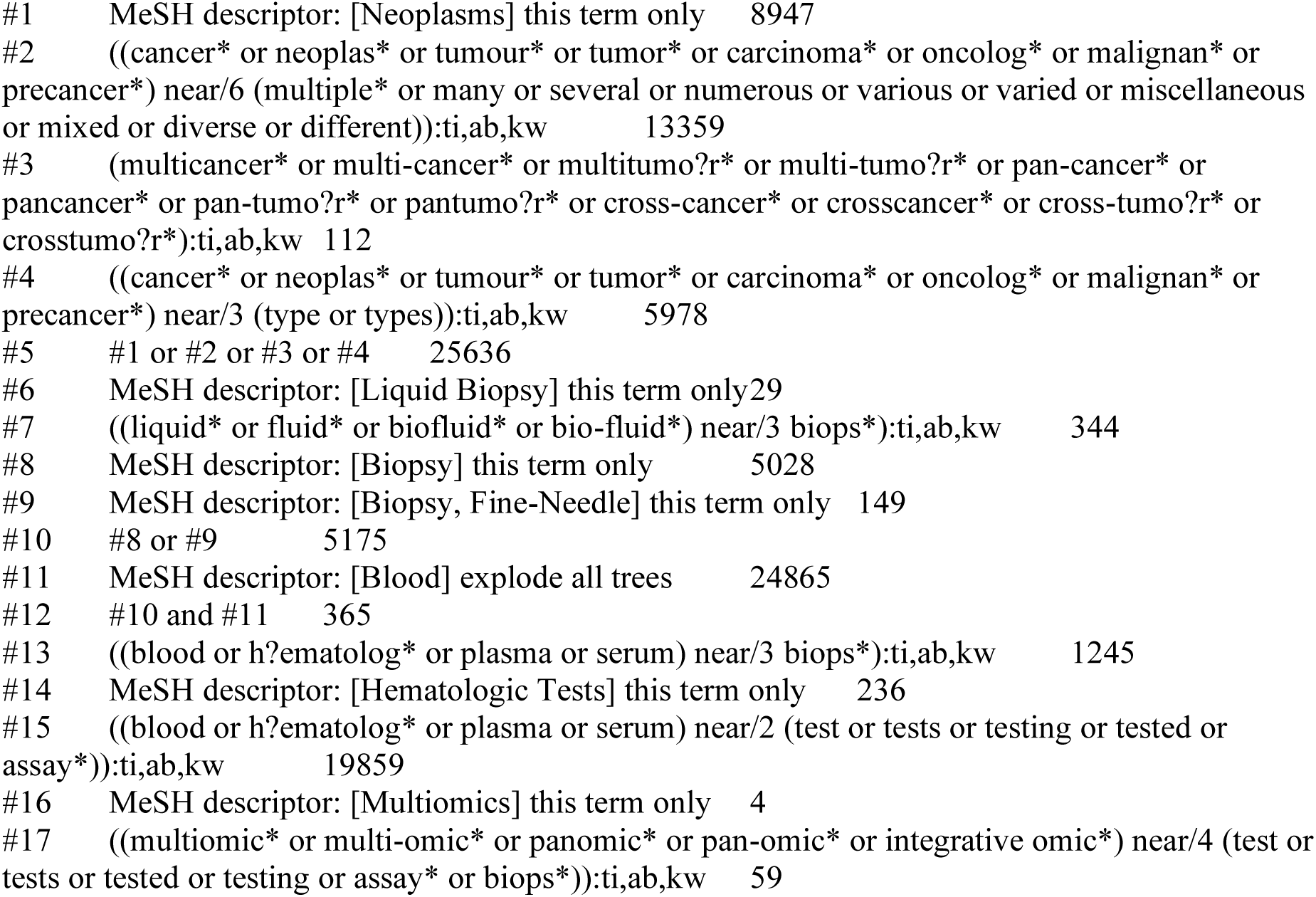

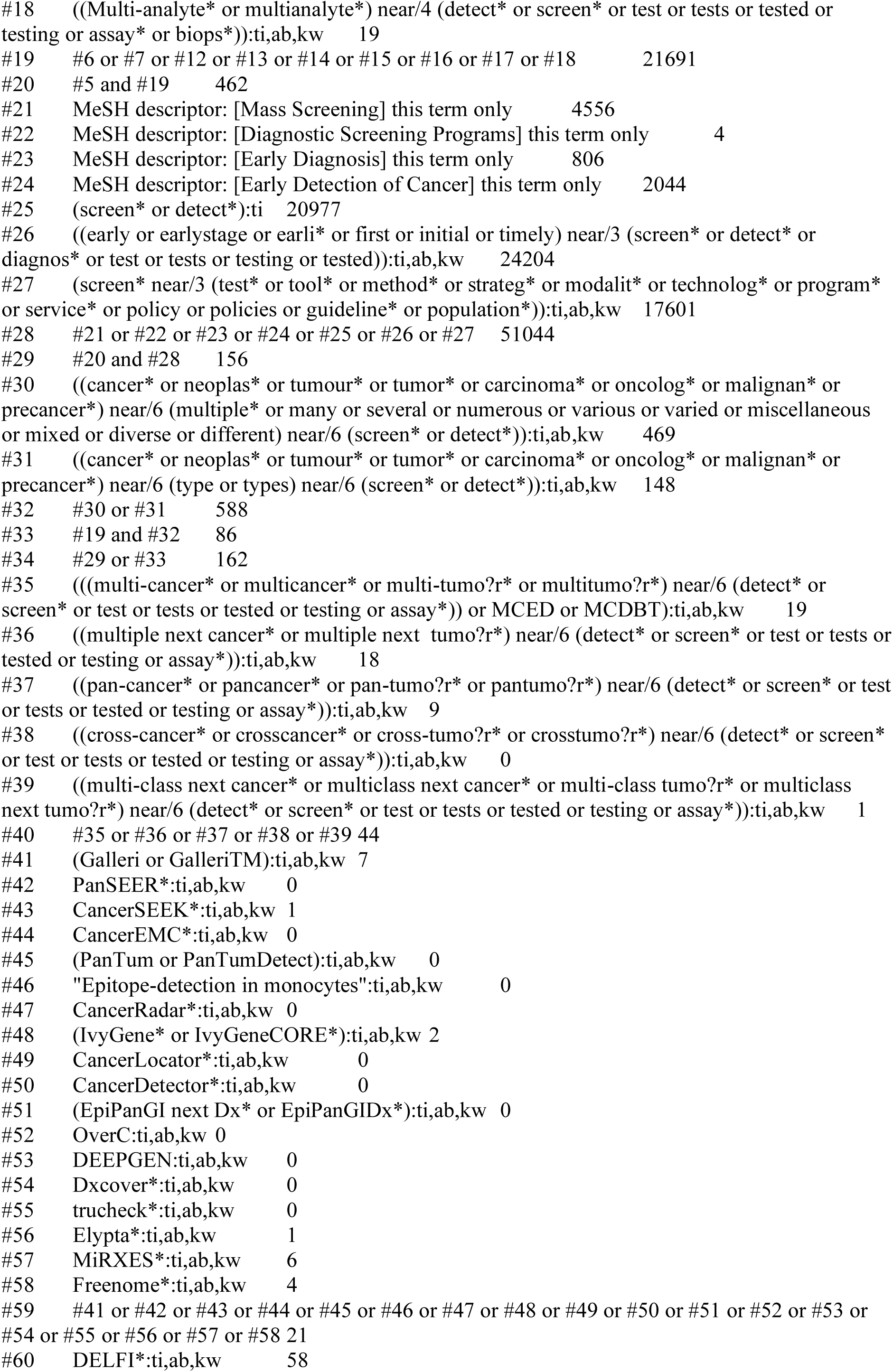

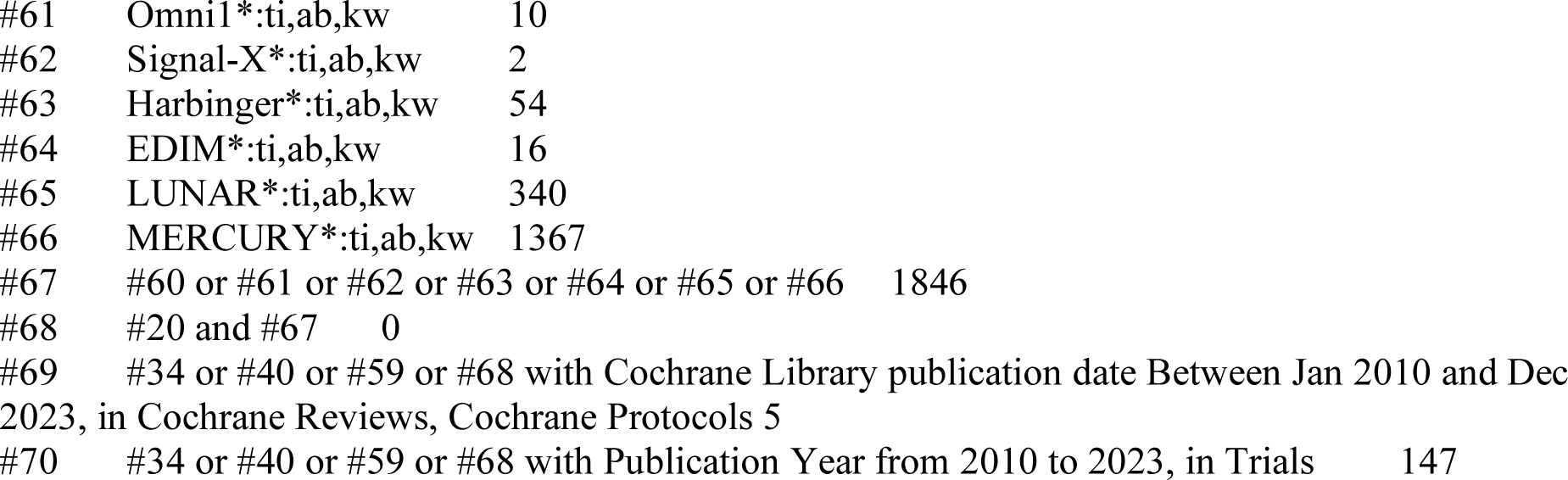

**Key:**

MeSH descriptor = subject heading (MeSH heading)

* = truncation

? = wildcard - zero or one characters

ti,ab,kw = terms in title, abstract or keyword fields

near/3 = terms within three words of each other (any order)

next = terms are next to each other

**Science Citation Index (SCI)**

**Conference Proceedings Citation Index – Science (CP-SCI)**

via Web of Science, Clarivate Analytics https://clarivate.com/

Date searched: 14th September 2023

Date range SCI: 1900 – present

Date range CP-SCI: 1990 – present

Records retrieved: 3635

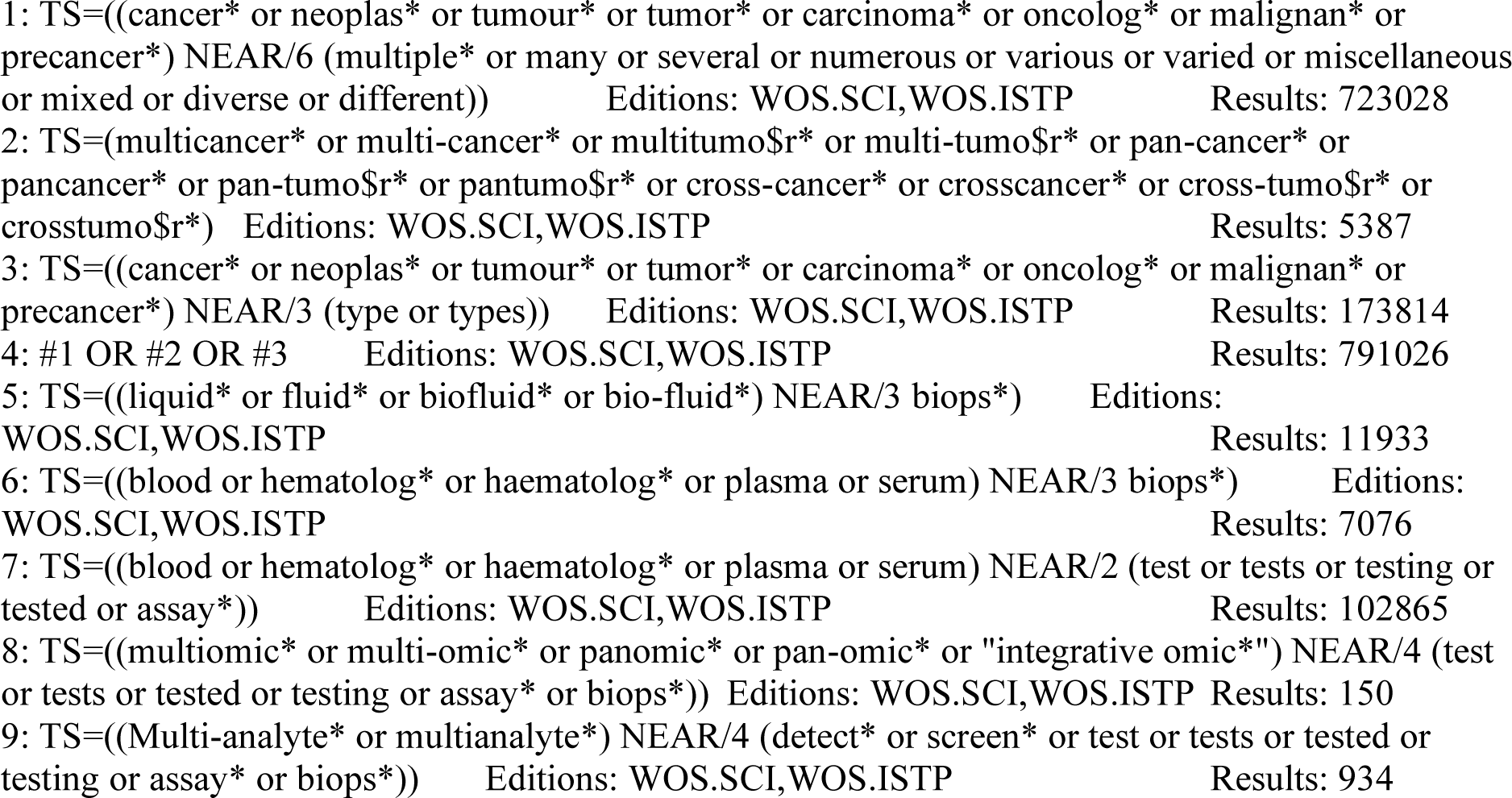

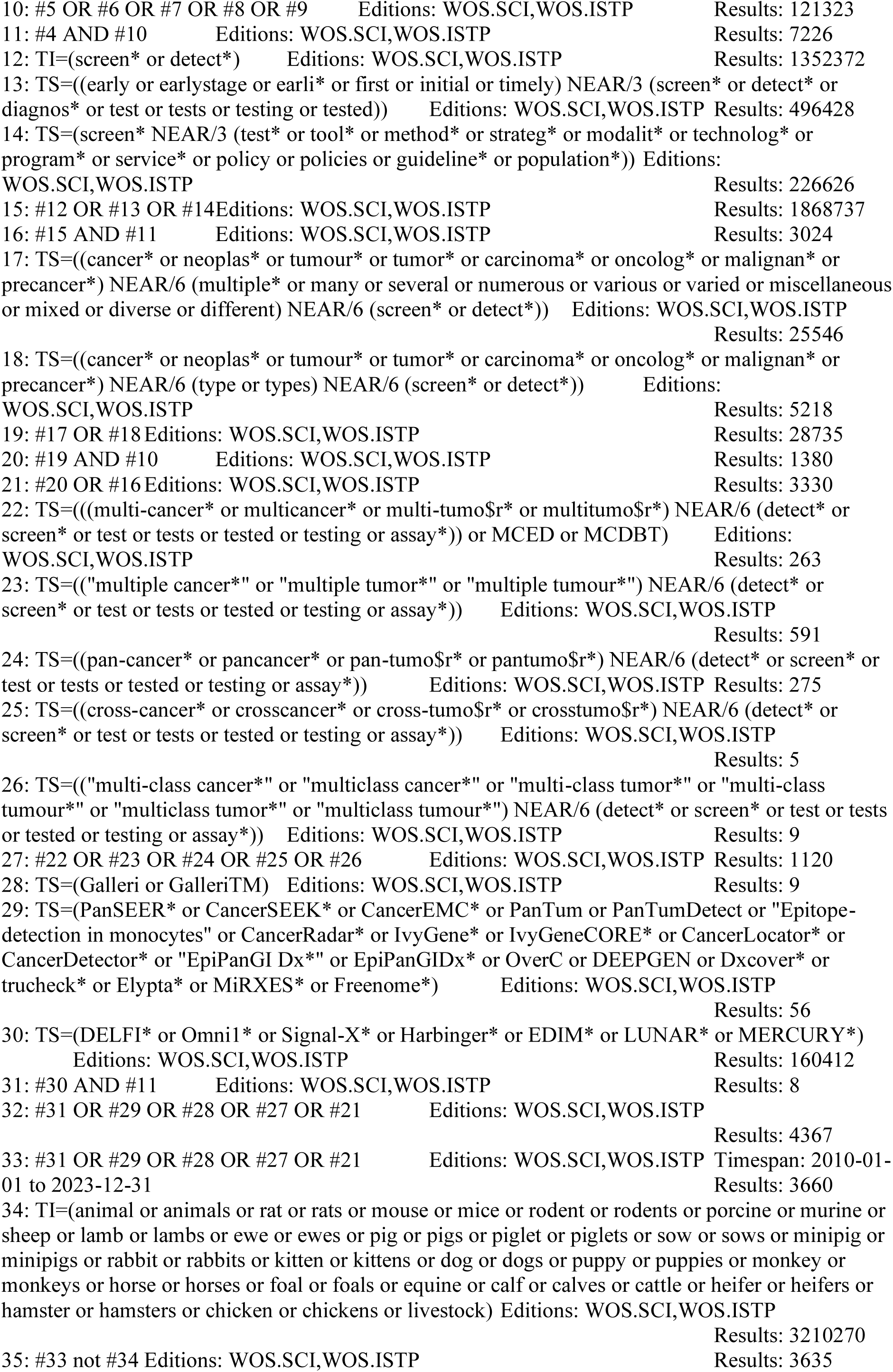

**Key:**

TS = topic tag; searches in title, abstract, author keywords and keywords plus fields

TI = search in title field

* = truncation

$ = represents zero or one character

NEAR/3 = terms within three words of each other (any order)

**EB Health - KSR Evidence**

via Ovid http://ovidsp.ovid.com/

Date range: 2015 to 2023 Week 37

Date searched: 14^th^ September 2023

Records retrieved: 45

1. ((cancer$ or neoplas$ or tumour$ or tumor$ or carcinoma$ or oncolog$ or malignan$ or precancer$) adj6 (multiple$ or many or several or numerous or various or varied or miscellaneous or mixed or diverse or different)).af. (5847)
2. (multicancer$ or multi-cancer$ or multitumo?r$ or multi-tumo?r$ or pan-cancer$ or pancancer$ or pan-tumo?r$ or pantumo?r$ or cross-cancer$ or crosscancer$ or cross-tumo?r$ or crosstumo?r$).af. (44)
3. ((cancer$ or neoplas$ or tumour$ or tumor$ or carcinoma$ or oncolog$ or malignan$ or precancer$) adj3 (type or types)).af. (3123)
4. 1 or 2 or 3 (7502)
5. ((liquid$ or fluid$ or biofluid$ or bio-fluid$) adj3 biops$).af. (143)
6. ((blood or h?ematolog$ or plasma or serum) adj3 biops$).af. (29)
7. ((blood or h?ematolog$ or plasma or serum) adj2 (test or tests or testing or tested or assay$)).af. (579)
8. ((multiomic$ or multi-omic$ or panomic$ or pan-omic$ or integrative omic$) adj4 (test or tests or tested or testing or assay$ or biops$)).af. (1)
9. ((Multi-analyte$ or multianalyte$) adj4 (detect$ or screen$ or test or tests or tested or testing or assay$ or biops$)).af. (1)
10. 5 or 6 or 7 or 8 or 9 (735)
11. 4 and 10 (52)
12. (screen$ or detect$).af. (53046)
13. ((early or earlystage or earli$ or first or initial or timely) adj3 (screen$ or detect$ or diagnos$ or test or tests or testing or tested)).af. (5713)
14. (screen$ adj3 (test$ or tool$ or method$ or strateg$ or modalit$ or technolog$ or program$ or service$ or policy or policies or guideline$ or population$)).af. (4258)
15. 12 or 13 or 14 (54744)
16. 11 and 15 (38)
17. ((cancer$ or neoplas$ or tumour$ or tumor$ or carcinoma$ or oncolog$ or malignan$ or precancer$) adj6 (multiple$ or many or several or numerous or various or varied or miscellaneous or mixed or diverse or different) adj6 (screen$ or detect$)).af. (196)
18. ((cancer$ or neoplas$ or tumour$ or tumor$ or carcinoma$ or oncolog$ or malignan$ or precancer$) adj6 (type or types) adj6 (screen$ or detect$)).af. (102)
19. 17 or 18 (277)
20. 10 and 19 (16)
21. 16 or 20 (39)
22. (((multi-cancer$ or multicancer$ or multi-tumo?r$ or multitumo?r$) adj6 (detect$ or screen$ or test or tests or tested or testing or assay$)) or MCED or MCDBT).af. (2)
23. ((multiple cancer$ or multiple tumo?r$) adj6 (detect$ or screen$ or test or tests or tested or testing or assay$)).af. (8)
24. ((pan-cancer$ or pancancer$ or pan-tumo?r$ or pantumo?r$) adj6 (detect$ or screen$ or test or tests or tested or testing or assay$)).af. (0)
25. ((cross-cancer$ or crosscancer$ or cross-tumo?r$ or crosstumo?r$) adj6 (detect$ or screen$ or test or tests or tested or testing or assay$)).af. (0)
26. ((multi-class cancer$ or multiclass cancer$ or multi-class tumo?r$ or multiclass tumo?r$) adj6 (detect$ or screen$ or test or tests or tested or testing or assay$)).af. (0)
27. 22 or 23 or 24 or 25 or 26 (8)
28. (Galleri or GalleriTM).af. (0)
29. PanSEER$.af. (0)
30. CancerSEEK$.af. (0)
31. CancerEMC$.af. (0)
32. (PanTum or PanTumDetect).af. (0)
33. "Epitope-detection in monocytes".af. (0)
34. CancerRadar$.af. (0)
35. (IvyGene$ or IvyGeneCORE$).af. (0)
36. CancerLocator$.af. (0)
37. CancerDetector$.af. (0)
38. (EpiPanGI Dx$ or EpiPanGIDx$).af. (0)
39. OverC.af. (0)
40. DEEPGEN.af. (0)
41. Dxcover$.af. (0)
42. trucheck$.af. (0)
43. Elypta$.af. (0)
44. MiRXES$.af. (0)
45. Freenome$.af. (0)
46. 28 or 29 or 30 or 31 or 32 or 33 or 34 or 35 or 36 or 37 or 38 or 39 or 40 or 41 or 42 or 43 or 44 or 45 (0)
47. DELFI$.af. (12)
48. Omni1$.af. (0)
49. Signal-X$.af. (0)
50. Harbinger$.af. (18)
51. EDIM$.af. (6)
52. LUNAR$.af. (19)
53. MERCURY$.af. (136)
54. 47 or 48 or 49 or 50 or 51 or 52 or 53 (191)
55. 11 and 54 (0)
56. 21 or 27 or 46 or 55 (45)
57. limit 56 to yr="2010 -Current" (45)

**Key**:

$ = truncation

? = optional wildcard – one or no characters

af = terms in any field

adj3 = terms within three words of each other (any order)

**Database of Abstracts of Reviews of Effects (DARE)**

**Health Technology Assessment (HTA) database**

via http://www.crd.york.ac.uk/CRDWeb/

Date range DARE: Inception – 31^st^ March 2015

Date range HTA database: Inception – 31^st^ March 2018

Date searched: 14^th^ September 2023

Records retrieved: 5

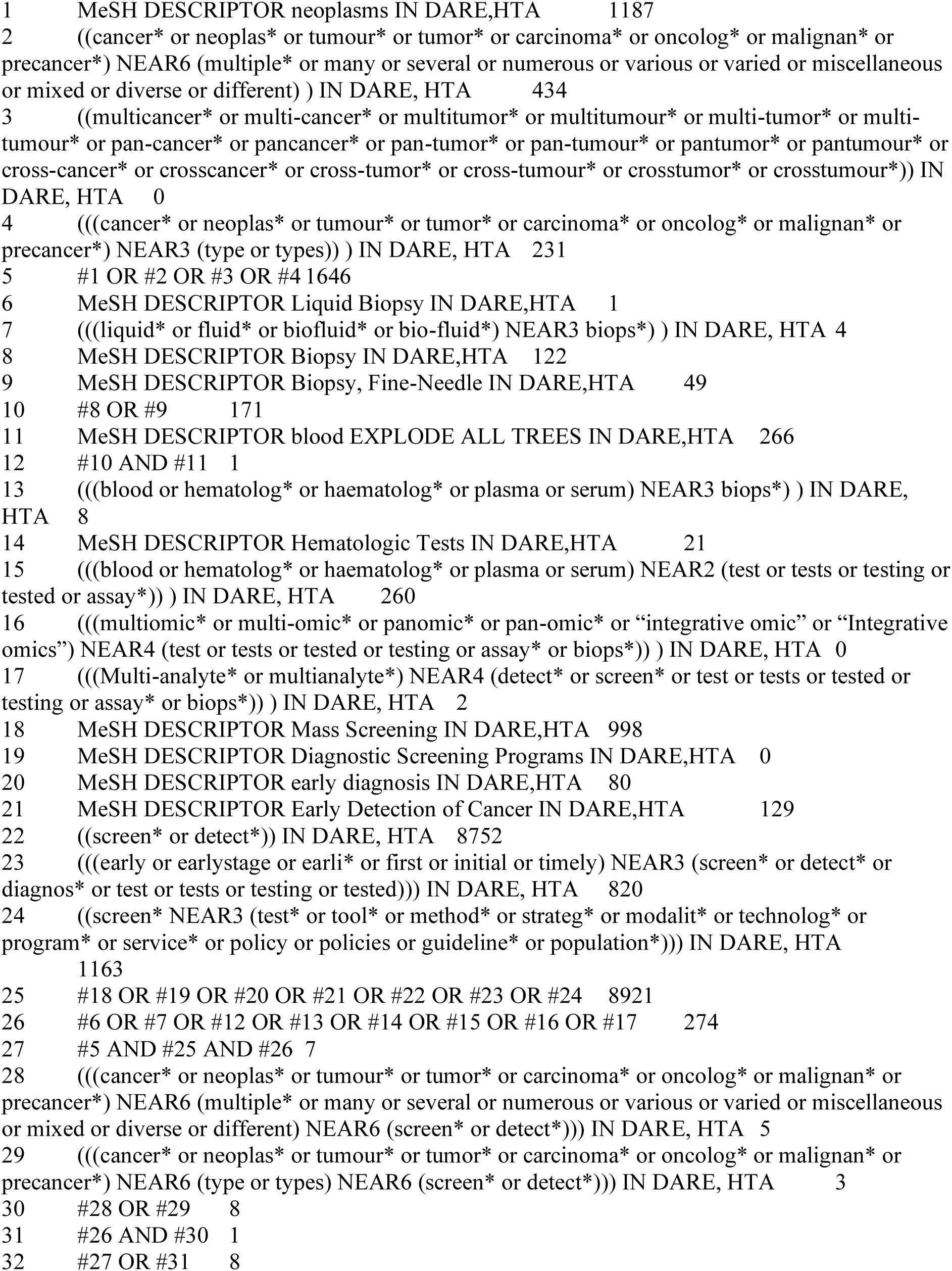

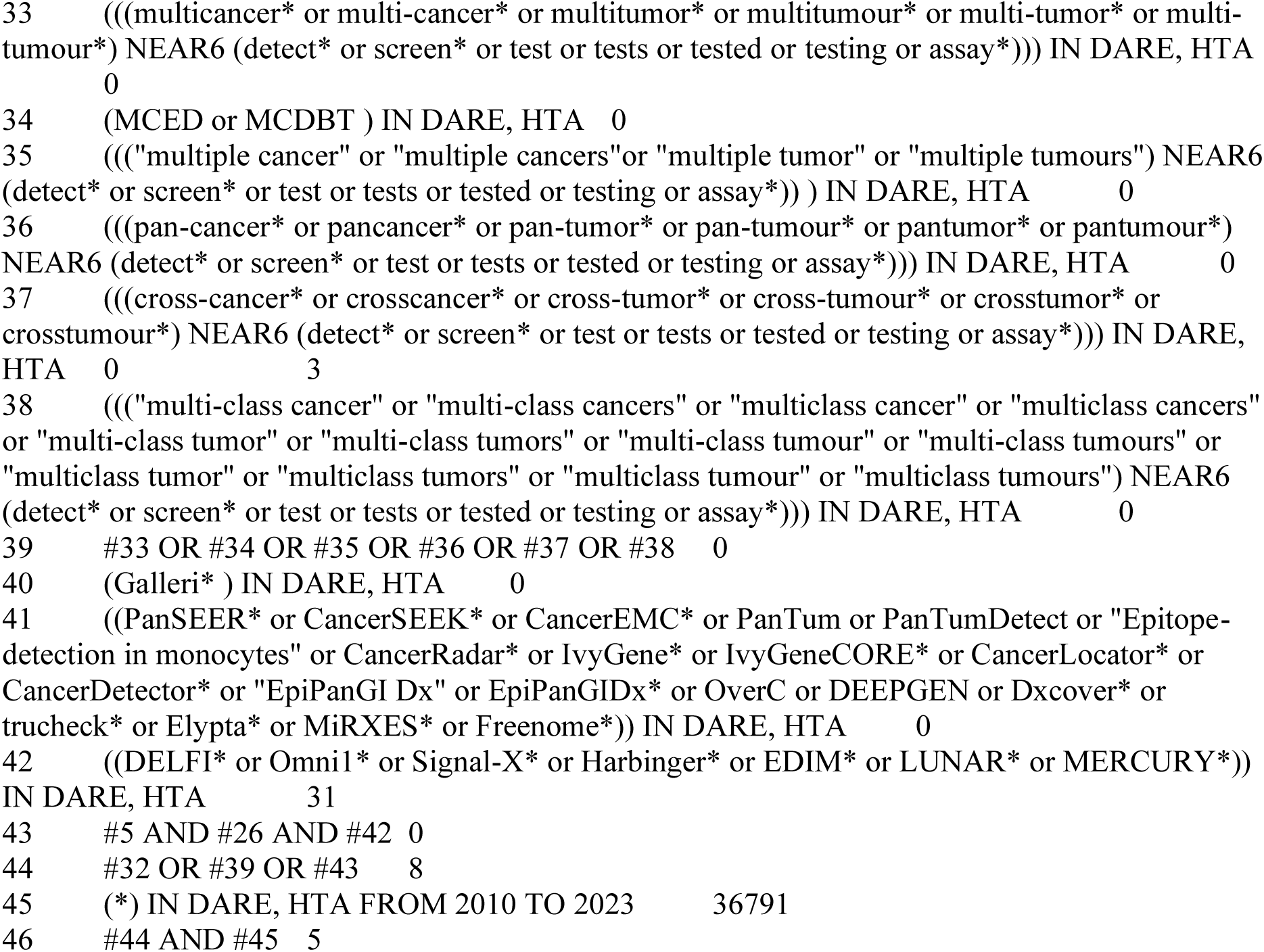

**Key:**

MeSH DESCRIPTOR = subject heading (MeSH heading)

* = truncation

adj3 = terms within three words of each other (order specified)

**International Health Technology Assessment (HTA) database**

via https://database.inahta.org/

Date range: Inception – 14^th^ September 2023

Date searched: 15^th^ September 2023 Records retrieved: 46

1. ((((screen* or detect* or diagnos* or test or tests or testing or tested)[Title] OR (screen* or detect* or diagnos* or test or tests or testing or tested)[abs] OR (screen* or detect* or diagnos* or test or tests or testing or tested)[Keywords]) AND ((early or earlystage or earli* or first or initial or timely)[Title] OR (early or earlystage or earli* or first or initial or timely)[abs] OR (early or earlystage or earli* or first or initial or timely)[Keywords])) OR (((test* or tool* or method* or strateg* or modalit* or technolog* or program* or service* or policy or policies or guideline* or population*)[Title] OR (test* or tool* or method* or strateg* or modalit* or technolog* or program* or service* or policy or policies or guideline* or population*)[abs] OR (test* or tool* or method* or strateg* or modalit* or technolog* or program* or service* or policy or policies or guideline* or population*)[Keywords]) AND ((screen*)[Title] OR (screen*)[abs] OR (screen*)[Keywords])) OR ((screen* or detect*)[Title]) OR ("Early Detection of Cancer"[mh]) OR ("Early Diagnosis"[mh]) OR ("Diagnostic Screening Programs"[mh]) OR ("Mass Screening"[mh])) AND ((((((Multi-analyte* or multianalyte*)[Title] OR (Multi-analyte* or multianalyte*)[abs] OR (Multi-analyte* or multianalyte*)[Keywords]) OR ((multiomic* or multi-omic* or panomic* or pan-omic* or "integrative omic" or "Integrative omics")[Title] OR (multiomic* or multi-omic* or panomic* or pan-omic* or "integrative omic" or "Integrative omics")[abs] OR (multiomic* or multi-omic* or panomic* or pan-omic* or "integrative omic" or "Integrative omics")[Keywords]) OR ("Multiomics"[mh])) AND ((biops* or test or tests or testing or tested or assay*)[Title] OR (biops* or test or tests or testing or tested or assay*)[abs] OR (biops* or test or tests or testing or tested or assay*)[Keywords])) OR ("Hematologic Tests"[mh]) OR (((biops* or test or tests or testing or tested or assay*)[Title] OR (biops* or test or tests or testing or tested or assay*)[abs] OR (biops* or test or tests or testing or tested or assay*)[Keywords]) AND ((blood or hematolog* or haematolog* or plasma or serum)[Title] OR (blood or hematolog* or haematolog* or plasma or serum)[abs] OR (blood or hematolog* or haematolog* or plasma or serum)[Keywords])) OR (("Blood"[mhe]) AND (("Biopsy, Fine-Needle"[mh]) OR ("Biopsy"[mh]))) OR (((biops*)[Title] OR (biops*)[abs] OR (biops*)[Keywords]) AND ((liquid* or fluid* or biofluid* or bio-fluid*)[Title] OR (liquid* or fluid* or biofluid* or bio-fluid*)[abs] OR (liquid* or fluid* or biofluid* or bio-fluid*)[Keywords])) OR ("Liquid Biopsy"[mh])) AND (((multicancer* or multi-cancer* or multitumor* or multitumour* or multi-tumor* or multi-tumour* or pan-cancer* or pancancer* or pan-tumor* or pan-tumour* or pantumor* or pantumour* or cross-cancer* or crosscancer* or cross-tumor* or cross-tumour* or crosstumor* or crosstumour*)[Title] OR (multicancer* or multi-cancer* or multitumor* or multitumour* or multi-tumor* or multi-tumour* or pan-cancer* or pancancer* or pan-tumor* or pan-tumour* or pantumor* or pantumour* or cross-cancer* or crosscancer* or cross-tumor* or cross-tumour* or crosstumor* or crosstumour*)[abs] OR (multicancer* or multi-cancer* or multitumor* or multitumour* or multi-tumor* or multi-tumour* or pan-cancer* or pancancer* or pan-tumor* or pan-tumour* or pantumor* or pantumour* or cross-cancer* or crosscancer* or cross-tumor* or cross-tumour* or crosstumor* or crosstumour*)[Keywords]) OR ((((type or types)[Title] OR (type or types)[abs] OR (type or types)[Keywords]) OR ((multiple* or many or several or numerous or various or varied or miscellaneous or mixed or diverse or different)[Title] OR (multiple* or many or several or numerous or various or varied or miscellaneous or mixed or diverse or different)[abs] OR (multiple* or many or several or numerous or various or varied or miscellaneous or mixed or diverse or different)[Keywords])) AND ((cancer* or neoplas* or tumour* or tumor* or carcinoma* or oncolog* or malignan* or precancer*)[Title] OR (cancer* or neoplas* or tumour* or tumor* or carcinoma* or oncolog* or malignan* or precancer*)[abs] OR (cancer* or neoplas* or tumour* or tumor* or carcinoma* or oncolog* or malignan* or precancer*)[Keywords])) OR ("Neoplasms"[mh]))) limit: 2010 to 2023, 44 hits
2. (Galleri or GalleriTM or PanSEER* or CancerSEEK* or CancerEMC* or PanTum or PanTumDetect or "Epitope-detection in monocytes" or CancerRadar* or IvyGene* or IvyGeneCORE* or CancerLocator* or CancerDetector* or "EpiPanGI Dx" or EpiPanGIDx* or OverC or DEEPGEN or Dxcover* or trucheck* or Elypta* or MiRXES* or Freenome*)[Title] OR (Galleri or GalleriTM or PanSEER* or CancerSEEK* or CancerEMC* or PanTum or PanTumDetect or "Epitope-detection in monocytes" or CancerRadar* or IvyGene* or IvyGeneCORE* or CancerLocator* or CancerDetector* or "EpiPanGI Dx" or EpiPanGIDx* or OverC or DEEPGEN or Dxcover* or trucheck* or Elypta* or MiRXES* or Freenome*)[abs] OR (Galleri or GalleriTM or PanSEER* or CancerSEEK* or CancerEMC* or PanTum or PanTumDetect or "Epitope-detection in monocytes" or CancerRadar* or IvyGene* or IvyGeneCORE* or CancerLocator* or CancerDetector* or "EpiPanGI Dx" or EpiPanGIDx* or OverC or DEEPGEN or Dxcover* or trucheck* or Elypta* or MiRXES* or Freenome*)[Keywords] limit: 2010 to 2023, 2 hits
3. ((DELFI* or Omni1* or Signal-X* or Harbinger* or EDIM* or LUNAR* or MERCURY*)[Title] OR (DELFI* or Omni1* or Signal-X* or Harbinger* or EDIM* or LUNAR* or MERCURY*)[abs] OR (DELFI* or Omni1* or Signal-X* or Harbinger* or EDIM* or LUNAR* or MERCURY*)[Keywords]) AND ((((((Multi-analyte* or multianalyte*)[Title] OR (Multi-analyte* or multianalyte*)[abs] OR (Multi-analyte* or multianalyte*)[Keywords]) OR ((multiomic* or multi-omic* or panomic* or pan-omic* or "integrative omic" or "Integrative omics")[Title] OR (multiomic* or multi-omic* or panomic* or pan-omic* or "integrative omic" or "Integrative omics")[abs] OR (multiomic* or multi-omic* or panomic* or pan-omic* or "integrative omic" or "Integrative omics")[Keywords]) OR ("Multiomics"[mh])) AND ((biops* or test or tests or testing or tested or assay*)[Title] OR (biops* or test or tests or testing or tested or assay*)[abs] OR (biops* or test or tests or testing or tested or assay*)[Keywords])) OR ("Hematologic Tests"[mh]) OR (((biops* or test or tests or testing or tested or assay*)[Title] OR (biops* or test or tests or testing or tested or assay*)[abs] OR (biops* or test or tests or testing or tested or assay*)[Keywords]) AND ((blood or hematolog* or haematolog* or plasma or serum)[Title] OR (blood or hematolog* or haematolog* or plasma or serum)[abs] OR (blood or hematolog* or haematolog* or plasma or serum)[Keywords])) OR (("Blood"[mhe]) AND (("Biopsy, Fine-Needle"[mh]) OR ("Biopsy"[mh]))) OR (((biops*)[Title] OR (biops*)[abs] OR (biops*)[Keywords]) AND ((liquid* or fluid* or biofluid* or bio-fluid*)[Title] OR (liquid* or fluid* or biofluid* or bio-fluid*)[abs] OR (liquid* or fluid* or biofluid* or bio-fluid*)[Keywords])) OR ("Liquid Biopsy"[mh])) AND (((multicancer* or multi-cancer* or multitumor* or multitumour* or multi-tumor* or multi-tumour* or pan-cancer* or pancancer* or pan-tumor* or pan-tumour* or pantumor* or pantumour* or cross-cancer* or crosscancer* or cross-tumor* or cross-tumour* or crosstumor* or crosstumour*)[Title] OR (multicancer* or multi-cancer* or multitumor* or multitumour* or multi-tumor* or multi-tumour* or pan-cancer* or pancancer* or pan-tumor* or pan-tumour* or pantumor* or pantumour* or cross-cancer* or crosscancer* or cross-tumor* or cross-tumour* or crosstumor* or crosstumour*)[abs] OR (multicancer* or multi-cancer* or multitumor* or multitumour* or multi-tumor* or multi-tumour* or pan-cancer* or pancancer* or pan-tumor* or pan-tumour* or pantumor* or pantumour* or cross-cancer* or crosscancer* or cross-tumor* or cross-tumour* or crosstumor* or crosstumour*)[Keywords]) OR ((((type or types)[Title] OR (type or types)[abs] OR (type or types)[Keywords]) OR ((multiple* or many or several or numerous or various or varied or miscellaneous or mixed or diverse or different)[Title] OR (multiple* or many or several or numerous or various or varied or miscellaneous or mixed or diverse or different)[abs] OR (multiple* or many or several or numerous or various or varied or miscellaneous or mixed or diverse or different)[Keywords])) AND ((cancer* or neoplas* or tumour* or tumor* or carcinoma* or oncolog* or malignan* or precancer*)[Title] OR (cancer* or neoplas* or tumour* or tumor* or carcinoma* or oncolog* or malignan* or precancer*)[abs] OR (cancer* or neoplas* or tumour* or tumor* or carcinoma* or oncolog* or malignan* or precancer*)[Keywords])) OR ("Neoplasms"[mh]))) 0 hits

Key:

[Keywords] = search of keywords field

[abs] = search of abstract field

[Title] = search of title field

[mh] = subject heading search

* = truncation

**PROSPERO**

via https://www.crd.york.ac.uk/prospero/

Date searched: 15^th^ September 2023

Records retrieved: 71

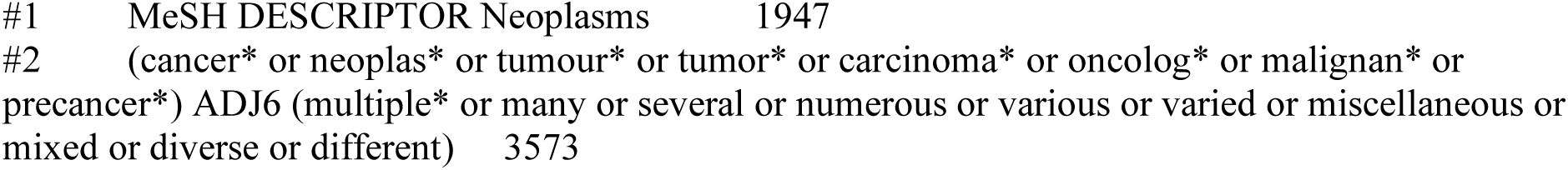

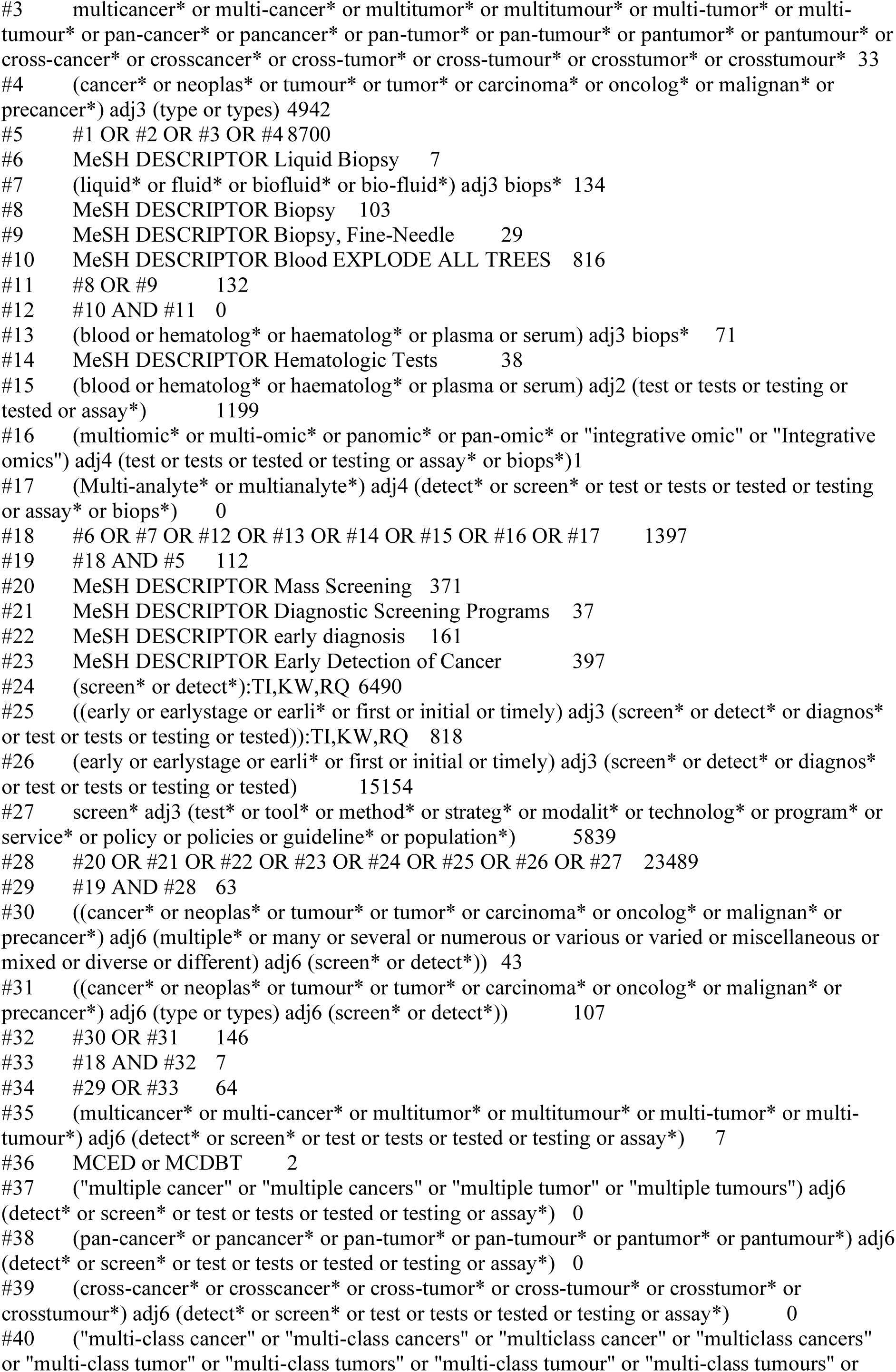

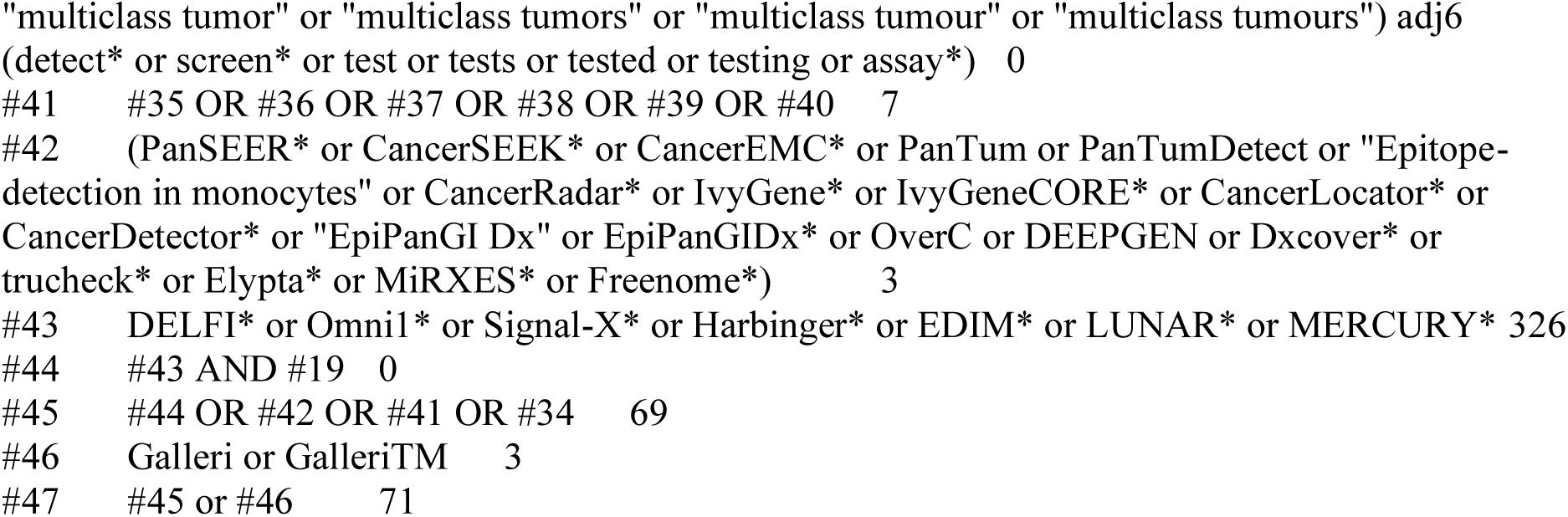

**Key:**

MeSH DESCRIPTOR = subject heading (MeSH heading)

* = truncation

TI,KW,RQ = terms in title, keyword or research question field

adj3 = terms within 3 words of each other (order specified)

**ClinicalTrials.gov**

https://clinicaltrials.gov/ct2/

Date searched: 15^th^ September 2023 Records retrieved: 325

1. 208 Studies found for: ("liquid biopsy" OR "blood test" OR "haematological test" OR "hematological test" OR "plasma test" OR "serum test") AND (screen OR screened OR screening OR detect OR detection) | (cancer OR neoplasm OR tumour OR tumor) AND (multiple OR many OR several OR numerous OR various OR varied OR miscellaneous OR mixed OR diverse OR different)
2. 2 Studies found for: ("liquid biopsy" OR "blood test" OR "haematological test" OR "hematological test" OR "plasma test" OR "serum test") AND (screen OR screened OR screening OR detect OR detection) | ("cancer type" OR "cancer types" OR "tumour type" OR "tumour types" OR "tumor type" OR "tumor types")
3. 12 Studies found for: (multiomic OR multi-omic OR multianalyte OR multi-analyte) AND (test OR tests OR tested OR testing OR assay OR biopsy) | (cancer OR neoplasm OR tumour OR tumor) AND (multiple OR many OR several OR numerous OR various OR varied OR miscellaneous OR mixed OR diverse OR different)
4. No Studies found for: (multiomic OR multi-omic OR multianalyte OR multi-analyte) AND (test OR tests OR tested OR testing OR assay OR biopsy) | ("cancer type" OR "cancer types" OR "tumour type" OR "tumour types" OR "tumor type" OR "tumor types")
5. 26 Studies found for: (detect OR detection OR screen OR screened OR screening OR test OR assay) | (multicancer OR multi-cancer OR multitumor OR multitumour OR multi-tumor OR multi-tumour)
6. 5 Studies found for: MCED OR MCDBT
7. 13 Studies found for: (detect OR detection OR screen OR screened OR screening OR test OR assay) | (pan-cancer OR pancancer OR pan-tumor OR pan-tumour OR pantumor OR pantumour)
8. 11 Studies found for: (detect OR detection OR screen OR screened OR screening OR test OR assay) | ("multiple cancer" OR "multiple cancers" OR "multiple tumor" OR "multiple tumors" OR "multiple tumour" OR "multiple tumours")
9. 5 Studies found for: (Galleri OR GalleriTM OR PanSEER OR CancerSEEK OR CancerEMC OR PanTum OR PanTumDetect OR "Epitope-detection in monocytes" OR CancerRadar OR IvyGene OR IvyGeneCORE OR CancerLocator OR CancerDetector OR "EpiPanGI Dx" OR EpiPanGIDx OR OverC OR DEEPGEN)
10. 17 Studies found for: Dxcover OR trucheck OR Elypta OR MiRXES OR Freenome OR "Harbinger health test" OR EDIM OR "MERCURY test"
11. 26 Studies found for: (DELFI OR Omni1 OR Signal-X OR LUNAR) AND (detect OR detection OR screen OR screened OR screening OR test OR assay or biopsy) | (cancer OR neoplasm OR tumour OR tumor)

**WHO International Clinical Trials Registry Platform (ICTRP)**

https://trialsearch.who.int/Default.aspx

Date searched: 18^th^ September 2023

Records retrieved: 266

Basic search interface used. No date limits available in basic search interface, therefore results from all years downloaded and records pre-2010 removed in EndNote.

1. 12 records for 12 trials found for: (cancer* OR neoplas* OR tumour* OR tumor*) AND (multiple* OR many OR several OR numerous OR various OR varied OR miscellaneous OR mixed OR diverse OR different) AND (liquid biops*)
2. 23 records for 17 trials found for: (cancer* OR neoplas* OR tumour* OR tumor*) AND (type OR types) AND (liquid biops*)
3. 212 records for 204 trials found for: (cancer* OR neoplas* OR tumour* OR tumor*) AND (multiple* OR many OR several OR numerous OR various OR varied OR miscellaneous OR mixed OR diverse OR different) AND (blood OR haematolog* OR hematolog* OR plasma OR serum) AND (screen* OR detect* OR test* OR assay*)
4. 2 records for 2 trials found for: (cancer* OR neoplas* OR tumour* OR tumor*) AND (multiple* OR many OR several OR numerous OR various OR varied OR miscellaneous OR mixed OR diverse OR different) AND (multiomic* OR multi-omic* OR multianalyte* OR multi-analyte*)
5. 29 records for 29 trials found for: (multicancer* OR multi-cancer* OR multitumor* OR multitumour* OR multi-tumor* OR multi-tumour*) AND (detect* OR screen* OR test OR tests OR tested OR testing OR assay*)
6. 9 records for 9 trials found for: ("multiple cancer" OR "multiple cancers" OR "multiple tumor" OR "multiple tumors" OR "multiple tumour" OR "multiple tumours") AND (detect* OR screen* OR test OR tests OR tested OR testing OR assay*)
7. 9 records for 9 trials found for: (pan-cancer* OR pancancer* OR pan-tumor* OR pan-tumour* OR pantumor* OR pantumour*) AND (detect* OR screen* OR test OR tests OR tested OR testing OR assay*)
8. No results were found for: (cross-cancer* OR crosscancer* OR cross-tumor* OR cross-tumour* OR crosstumor* OR crosstumour*) AND (detect* OR screen* OR test OR tests OR tested OR testing OR assay*)
9. 9 records for 9 trials found for: (Galleri OR GalleriTM OR PanSEER OR CancerSEEK OR CancerEMC OR PanTum OR PanTumDetect OR Epitope-detection in monocytes OR CancerRadar OR IvyGene OR IvyGeneCORE OR CancerLocator OR CancerDetector OR “EpiPanGI Dx” OR EpiPanGIDx OR OverC OR DEEPGEN)
10. 12 records for 12 trials found for: Dxcover OR trucheck OR Elypta OR MiRXES OR Freenome OR “Harbinger health test” OR EDIM OR “MERCURY test”
11. 10 records for 10 trials found for: (DELFI OR Omni1 OR Signal-X OR LUNAR) AND (cancer* OR neoplas* OR tumour* OR tumor*)
12. No Studies found for: (multiomic OR multi-omic OR multianalyte OR multi-analyte) AND (test OR tests OR tested OR testing OR assay OR biopsy) | ("cancer type" OR "cancer types" OR "tumour type" OR "tumour types" OR "tumor type" OR "tumor types")

### 2. Website searches

**HTA Agencies**

Date searched: 19^th^ September 2023 Records retrieved: 12

Browsed or searched the following HTA Agency websites to check for additional reports not found through database searches. A date limit of 2010 was applied.

**Agency for Healthcare Research and Quality (AHRQ), USA**

1. https://www.ahrq.gov/research/findings/ta/index.html Browsed list of technology assessments, topic refinements and archive of technology assessments – 3 relevant reports found
2. https://www.ahrq.gov/research/findings/evidence-based-reports/search.html Filtered list to cancer and browsed 133 results – 1 relevant report found

**Adelaide Health Technology Assessment (AHTA), AUSTRALIA**

https://health.adelaide.edu.au/adelaide-health-technology-assessment/research-services/publications/

Browsed following lists (2010 onwards):

reports and monographs – none relevant

protocols – none relevant

Technology Briefs and Prioritising summaries – 2 relevant reports found

Presentations and abstracts – none relevant

**Agency for Care Effectiveness – Singapore**

https://www.ace-hta.gov.sg/

Browsed lists of technology guidance, horizon scanning reports, scientific publications – 1 relevant report found

**Austrian Institute for Health Technology Assessment**

https://eprints.aihta.at/

Search terms used:

1. liquid biopsy – 24 results browsed, none relevant
2. multicancer – 0
3. multi-cancer – 0
4. cancer screening – 69 results browsed, none relevant

**Canadian Agency for Drugs and Technologies in Health (CADTH), CANADA**

https://www.cadth.ca/

General search https://www.cadth.ca/search?s=&facets_query=&page=0

1. liquid biopsy – 52 results, 4 potentially relevant
2. multi-cancer early detection - 3 results, all duplicates with 1.
3. MCED – 3 results, all duplicates with 1.

Browsed projects in progress page and topics under consideration page – none relevant

**Health Information and Quality Authority, IRELAND HIQA**

https://www.hiqa.ie/reports-and-publications/health-technology-assessments

Browsed all 126 health technology assessments – none relevant

**Scottish Health Technologies Group**

https://shtg.scot/our-advice/

Browsed all publications 2010-2023 – 1 relevant report found in progress: https://shtg.scot/what-we-do/work-programme/

Browsed all publications – none relevant

**Health Technology Wales**

https://healthtechnology.wales/reports-guidance/

1. liquid biopsy – 4 results – none relevant
2. multi-cancer early detection – 52 results – none relevant Browsed all 251 reports – none relevant

**National Institute for Health and Care Excellence**

https://www.nice.org.uk/

General search box:

1. “liquid biopsy” – 2 results, none relevant.
2. “multicancer” – 0 results.
3. “multi-cancer” – 0 results.
4. https://www.nice.org.uk/guidance/published Searched for cancer, limited to 2010 to current, filtered to Diagnostic guidance – 11 results browsed for relevance, none relevant in development – 4 results browsed for relevance, none relevant awaiting development – 101 results browsed for relevance, none relevant
5. https://www.nice.org.uk/guidance/published Searched for cancer, limited to 2010 to current, filtered to Medtech innovation briefings – 24 results browsed for relevance, none relevant

**National Institute for Health Research Journals Library**

https://www.journalslibrary.nihr.ac.uk/search/#/

1. liquid biopsy – 79 results browsed, none relevant
2. muticancer – 0
3. multi-cancer – 0
4. cancer screening, limited to HTA assessments – 363 results browsed, 1 relevant https://www.journalslibrary.nihr.ac.uk/hta/hta24660/#/abstract

**Belgian Health Care Knowledge Centre**

https://www.kce.fgov.be/en/all-reports-0

1. cancer - Filtered to HTA reports – browsed 19 reports – none relevant

**Test manufacturer website searches**

After screening, the included studies were examined to produce a list of company names and their tests. The website of each company (where available) was located and browsed to find further relevant references relating to multi-cancer early detection tests used for screening published from 2020 onwards.

1. Company: Adela Test: No name https://www.adelabio.com/ Date searched: 11^th^ October 2023
2. Company: Ajinomoto Group Test: AminoIndex Cancer Screening (AICS) https://www.ajinomoto.com/innovation/action/aminoindex Date searched: 10^th^ October 2023
3. Company: AnPac Bio-Medical Science Test: No name https://www.anpacbio.com/ Date searched: 10^th^ October2023
4. Company: AVRT Test: Aristotle https://avrtnow.com/aristotle/ Date searched: 10^th^ October 2023
5. Company: Burning Rock DX Test: OverC https://us.brbiotech.com/ Date searched: 10^th^ October 2023
6. Company: Datar Cancer Genetics Tests: TruCheck, Trueblood, EasyCheck https://datarpgx.com/ Date searched: 10^th^ October 2023
7. Company: Elypta Test: No name https://www.elypta.com/ Date searched: 10^th^ October 2023
8. Company: Exact Sciences Test: CancerSEEK https://www.exactsciences.com/ Date searched: 10^th^ October 2023
9. Company: Gene Solutions Test: SPOT-MAS https://genesolutions.vn/en/product/spot-mas/ Date searched: 10^th^ October 2023
10. Company: GenePlus Bejing Test: No name https://en.geneplus.cn/home Date searched: 10^th^ October 2023
11. Company: Geneseeq Test: Mercury https://na.geneseeq.com/ Date searched: 11^th^ October 2023
12. Company: GRAIL Test: Galleri https://grail.com/ https://www.galleri.com/ Date searched: 11^th^ October 2023
13. Company: Guardant Test: Guardant LUNAR-2 (also known as Shield) https://guardanthealth.com/ Date searched: 10^th^ October 2023
14. Company: Harbinger Health Test: Harbinger Health Test https://www.harbinger-health.com/ Date searched: 11^th^ October 2023
15. Company: RMDM Group Test: PanTum test https://rmdm.group/ Date searched: 11^th^ October 2023
16. Company: SeekIn Test: OncoSeek https://www.seekincancer.com/ Date searched: 10^th^ October 2023
17. Company: Singlera Genomics Test: PanSeerX https://singleraoncology.com/ Date searched: 10^th^ October 2023

Websites could not be located for the following companies: Nanjing Shihe Jiyin, Carcimun Biotech, and Shenzhen Kerida Health Technology. In addition, the names of companies producing the following tests could not be found: SpecGastro test and CancerD24.

## APPENDIX 3. PPI INVITE

### Multi-cancer early detection tests for general population screening

General population cancer screening is only available for some cancers (cervical, breast, bowel). Additionally, in some areas of England and Wales people at high risk of developing lung cancer can receive a lung health check. Breast, prostate, lung, and bowel cancers together account for just over half of all new cancers diagnosed. Most other cancers are detected after presentation of symptoms. Many of these will be diagnosed at stages III and IV. This means treatment options may be more limited.

New tests that look for signs of cancer in blood (blood-based multi-cancer early detection tests - MCEDs) are being developed; they aim to detect multiple different cancers at an early stage, when they are potentially more treatable. The NHS Long Term Plan ambition seeks to diagnose 75% of cancers at stage I or II, to enable more effective treatment. An MCED test embedded within a national population-based screening programme, in addition to existing cancer screening programmes, may increase the number of cancers diagnosed at an earlier stage. This could potentially improve the likelihood of successful treatment and increase survival rates.

The possible introduction of such a tests raises a number of questions. Several research projects have been commissioned to explore the impact of these tests within the context of a general population screening programme. We would like to explore some of these questions with patients and the public, particularly those who might be eligible for this type of screening and those with lived experience of a cancer diagnosis.

We are planning an online focus group involving a number of organisations to discuss these on **Friday 17^th^ November between 1 and 2.30pm.**

Some of the things that we want to explore on this call are:

- How willing people are to take up MCED screening opportunities
- What impact this might have on other screening programmes
- What concerns people have about false negative and false positive results
- How people feel about universal versus targeted use of screening tools

## APPENDIX 4. LIST OF EXCLUDED STUDIES WITH RATIONALE

### Excluded on Intervention (n=130)

1. cfDNA Assay Prospective Observational Validation for Early Cancer Detection and Minimal Residual Disease.
2. Collecting Blood Samples From Patients With and Without Cancer to Evaluate Tests for Early Cancer Detection.
3. Development and Validation of Harbinger Health Test for Early Cancer Detection.
4. Multi-Cancer Early Detection (MCED) of Firefighters.
5. PAN-study: Pan-Cancer Early Detection Study (PAN).
6. PERformance of Multi-Cancer Early-detectIon Based on Various Biomarkers in fEmale Cancers, PERCEIVEII.
7. PERformance of Multi-Cancer Early-detectIon Based on Various Biomarkers in fEmale Cancers, PERCEIVE-I.
8. PRediction Of Five Usual Tumors Using Blood Test for Risk Assessment and Early Detection.
9. Prospective Screening and Differentiating Common Cancers Using Peripheral Blood Cell-Free DNA Sequencing.
10. Screening for High Frequency Malignant Disease.
11. The FuSion Program: A Prospective and Multicenter Cohort Study of Pan-Cancer Screening in Chinese Population.
12. The Jinling Cohort.
13. The PREDICT Study: Prospective Early Detection In a Population at High-risk for Common Malignant Tumor.
14. The STRIVE Study: Development of a Blood Test for Early Detection of Multiple Cancer Types.
15. Clinical Study of Pan-cancer DNA Methylation Test in Plasma.
16. LEVANTIS-0087A: GAGomes for Multi-Cancer Early Detection in Asymptomatic Adults (LEV87A).
17. LEVANTIS-0093A: GAGomes for Multi-Cancer Early Detection in High-Risk Adults (LEV93A).
18. Non-invasive Liquid Biopsy Analysis of Epigenomics Signatures in Multiple Cancer Types.
19. Pan-canceR Early DetectIon projeCT.
20. Pan-canceR Early-Stage deteCtion by lIquid Biopsy tEchNique projecT.
21. Project CADENCE (CAncer Detected Early caN be CurEd).
22. The Sanderson Study: A Case Control Study for the Development of Multiomics Blood Tests for Cancer Screening.
23. Akolkar D, Patil D, Crook T, et al. Circulating ensembles of tumor-associated cells: A redoubtable new systemic hallmark of cancer. International Journal of Cancer 2020;146:3485-94. doi: https://dx.doi.org/10.1002/ijc.32815
24. Alexander G, Lin W, Ramaiah M, et al. Analytical validation of a multi-cancer early detection test with tissue localization using a cell-free DNA-based targeted methylation assay. Cancer Research Conference: American Association for Cancer Research Annual Meeting, AACR 2020;80 doi: https://dx.doi.org/10.1158/1538-7445.AM2020-721
25. Alexander GE, Jung B, Ji L, et al. Analytical performance of a cfDNA-based targeted methylation multi-cancer early detection test for population-scale screening. Cancer Research Conference: AACR Annual Meeting 2021;81 doi: https://dx.doi.org/10.1158/1538-7445.AM2021-112
26. Antonowicz S, Kumar S, Wiggins T, et al. Diagnostic metabolomic blood tests for endoluminal gastrointestinal cancer - a systematic review and assessment of quality. Cancer Epidemiol Biomarkers Prev 2016;25:6-15. doi: http://dx.doi.org/10.1158/1055-9965.EPI-15-0524
27. Baker M, Cameron JM, Sala A, et al. Multicancer early detection with a spectroscopic liquid biopsy platform. Journal of Clinical Oncology Conference: Annual Meeting of the American Society of Clinical Oncology, ASCO 2022;40 doi: https://dx.doi.org/10.1200/JCO.2022.40.16_suppl.3034
28. Bao H, Wang Z, Ma X, et al. Letter to the Editor: An ultra-sensitive assay using cell-free DNA fragmentomics for multi-cancer early detection. Molecular Cancer 2022;21:129. doi: https://dx.doi.org/10.1186/s12943-022-01594-w
29. Bergamaschi A, Collins F, Ellison C, et al. Changes in DNA hydroxymethylation for the detection of multiple cancers in plasma cellfree DNA. Journal of Clinical Oncology Conference 2019;37 doi: https://dx.doi.org/10.1200/JCO.2019.37.15_suppl.3058
30. Best M, Sol N, Kooi I, et al. Allowance of tumor-educated platelets for multiclass liquid biopsy-based diagnosis of cancer. Journal of Clinical Oncology Conference 2015;33
31. Bratulic S, Limeta A, Dabestani S, et al. Noninvasive detection of any-stage cancer using free glycosaminoglycans. Proceedings of the National Academy of Sciences of the United States of America 2022;119:e2115328119. doi: https://dx.doi.org/10.1073/pnas.2115328119
32. Bryce AH, Liu MC, Seiden MV, et al. Performance of a cell-free DNA-based multi-cancer detection test as a tool for diagnostic resolution of symptomatic cancers. Cancer Research Conference: AACR Annual Meeting 2021;81 doi: https://dx.doi.org/10.1158/1538-7445.AM2021-LB058
33. Budnik B, Amirkhani H, Forouzanfar MH, et al. A novel proteomics-based plasma test for early detection of multiple cancers in the general population. medRxiv 2023;08 doi: https://dx.doi.org/10.1101/2023.05.06.23289613
34. Cameron JM, Antoniou G, Brennan PM, et al. Early colorectal cancer detection with a spectroscopic liquid biopsy. Cancer Research Conference: American Association for Cancer Research Annual Meeting, ACCR 2023;83 doi: https://dx.doi.org/10.1158/1538-7445.AM2023-6506
35. Cameron JM, Sala A, Antoniou G, et al. Multi-cancer early detection with a spectroscopic liquid biopsy platform. Cancer Research Conference: American Association for Cancer Research Annual Meeting, ACCR 2020;82 doi: https://dx.doi.org/10.1158/1538-7445.AM2022-5920
36. Carey J, Leal A, Chesnick B, et al. Detecting cancer using genome-wide cfDNA nucleosomal fragmentation in a prospective multi cancer cohort. Cancer Research Conference: AACR Annual Meeting 2021;81 doi: https://dx.doi.org/10.1158/1538-7445.AM2021-570
37. Che H, Jatsenko T, Lenaerts L, et al. Pan-cancer detection and typing by mining patterns in large genome-wide cell-free DNA sequencing datasets. medRxiv 2022;20 doi: https://dx.doi.org/10.1101/2022.02.16.22268780
38. Chen J, Yang Y, Wang Z, et al. A Multicancer Malignant Pleural Effusion Diagnostic Test Using Hexokinase 2 and Single-Cell Sequencing. Clinical Chemistry 2022;68:680-90. doi: https://dx.doi.org/10.1093/clinchem/hvac003
39. Chen X, Dong Z, Hubbell E, et al. Prognostic Significance of Blood-Based Multi-cancer Detection in Plasma Cell-Free DNA. Clinical Cancer Research 2021;27:4221-29. doi: https://dx.doi.org/10.1158/1078-0432.CCR-21-0417
40. Chen X, Gole J, Gore A, et al. Non-invasive early detection of cancer four years before conventional diagnosis using a blood test. Nature communications 2020;11:3475. doi: https://dx.doi.org/10.1038/s41467-020-17316-z
41. ChiCtr. PanTum technique for the detection of peripheral blood APO10 and TKTL1 in the diagnosis of high incidence of malignant tumors in Chinese population, 2020.
42. ChiCtr. A prospective, multicenter cohort study of pan-cancer screening in Chinese population, 2021.
43. ChiCtr. SZ-PILOT Study: Prospective observational study of the YiDiXueTM multi-cancer early detection Kit in multi-cancer early screening in normal people, 2022.
44. Constancio V, Nunes SP, Moreira-Barbosa C, et al. Early detection of the major male cancer types in blood-based liquid biopsies using a DNA methylation panel. Clinical Epigenetics 2019;11:175. doi: https://dx.doi.org/10.1186/s13148-019-0779-x
45. Cree IA. Plasma cfDNA for early cancer detection. Tumor Biology 2016;37(Supplement 1):S13. doi: https://dx.doi.org/10.1007/s13277-016-5287-4
46. Cree IA, Uttley L, Buckley W, et al. The evidence base for circulating tumour DNA blood-based biomarkers for the early detection of cancer: a systematic mapping review. BMC Cancer 2017;17:697. doi: https://dx.doi.org/10.1186/s12885-017-3693-7
47. CTRI. A trial for confirming the accuracy of PanTum test for solid tumor detection, 2022.
48. CTRI. A simple blood test to understand presence or absence of cancer. 2023. http://www.ctri.nic.in/Clinicaltrials/pmaindet2.php?trialid=81990 (accessed 2023).
49. CTRI. A simple blood test to understand presence or absence of cancer. 2023. http://www.ctri.nic.in/Clinicaltrials/pmaindet2.php?trialid=87700 (accessed 2023).
50. Desai M, Shchegrov SR, Chai S, et al. Analytical validation of a tissue-free, multicancer, post-diagnosis cancer research test that uses cellfree DNA methylation profiling. Cancer Research Conference: American Association for Cancer Research Annual Meeting, ACCR 2023;83 doi: https://dx.doi.org/10.1158/1538-7445.AM2023-LB297
51. Dev HS, Lach R, Park G, et al. Early detection assay using ctDNA methylation for hard-to-detect cases including prostate and renal cancer. European Urology 2023;83(Supplement 1):S533. doi: https://dx.doi.org/10.1016/S0302-2838%2823%2900414-1
52. Douville C, Cohen JD, Ptak J, et al. Assessing aneuploidy with repetitive element sequencing. Proceedings of the National Academy of Sciences of the United States of America 2020;117:4858-63. https://dx.doi.org/10.1073/pnas.1910041117
53. Douville C, Nobles C, Hwang HJ, et al. 73P Multi-cancer early detection through evaluation of aneuploidy, methylation, and protein biomarkers in plasma. Annals of Oncology 2022;33(Supplement 7):S575. doi: https://dx.doi.org/10.1016/j.annonc.2022.07.106
54. Gao Q, Li B, Cai S, et al. LBA3 Early detection and localization of multiple cancers using a blood-based methylation assay (ELSA-seq). Annals of Oncology 2020;31(Supplement 6):S1358. doi: https://dx.doi.org/10.1016/j.annonc.2020.10.292
55. Gao Q, Li B, Cai S, et al. Early detection and localization ofmultiple cancers using a blood-based methylation assay (ELSA-seq). Journal of Clinical Oncology Conference 2021;39 doi: https://dx.doi.org/10.1200/JCO.2021.39.3_suppl.459
56. Gao Q, Wang C, Yang X, et al. A multi-cancer early detection model based on liquid biopsy of multi-omics biomarkers: A proof of concept study (PROMISE study). Annals of Oncology 2022;33(Supplement 7):S963-S64. doi: https://dx.doi.org/10.1016/j.annonc.2022.07.1035
57. Gao Q, Zhang Y, Xu J, et al. Clinical validation of a multicancer detection blood test by circulating cell-free DNA (cfDNA) methylation sequencing: The THUNDER study. Journal of Clinical Oncology Conference: Annual Meeting of the American Society of Clinical Oncology, ASCO 2022;40 doi: https://dx.doi.org/10.1200/JCO.2022.40.16_suppl.10544
58. Gatto F, Bratulic S, Cavarretta ITR, et al. Detection of any-stage cancer using plasma and urine glycosaminoglycans. Journal of Clinical Oncology Conference: Annual Meeting of the American Society of Clinical Oncology, ASCO 2021;39 doi: https://dx.doi.org/10.1200/JCO.2021.39.15_suppl.3034
59. Greenwald ZR, El-Zein M, Bouten S, et al. Mobile Screening Units for the Early Detection of Cancer: A Systematic Review. Cancer Epidemiology, Biomarkers & Prevention 2017;26:1679-94. doi: https://dx.doi.org/10.1158/1055-9965.EPI-17-0454
60. Han T, Hong Y, Zhihua P, et al. An ultrasensitive method for noninvasive pan-cancer early detection based on targeted methylation sequencing of cellfree DNA. Journal of Clinical Oncology Conference: Annual Meeting of the American Society of Clinical Oncology, ASCO 2021;39 doi: https://dx.doi.org/10.1200/JCO.2021.39.15-suppl.10544
61. Han T, Liu T, Suxing L, et al. An ultrasensitive approach for cancer screening and tissue of origin prediction based on targeted methylation sequencing of cell-free DNA. Journal of Clinical Oncology Conference: Annual Meeting of the American Society of Clinical Oncology, ASCO 2022;40 doi: https://dx.doi.org/10.1200/JCO.2022.40.16_suppl.10553
62. Hartman AR, Oxnard G, Klein E, et al. Multicancer detection of early-stage cancers with simultaneous tissue localization using a plasma cfDNA-based targeted methylation assay. Clinical Cancer Research Conference: AACR Special Conference on Advances in Liquid Biopsies Miami, FL United States 2020;26 doi: https://dx.doi.org/10.1158/1557-3265.LiqBiop20-IA02
63. Hashimoto K, Inada M, Yamamoto Y, et al. Preliminary evaluation of miR-1307-3p in human serum for detection of 13 types of solid cancer using microRNA chip. Heliyon 2021;7:7. https://dx.doi.org/10.1016/j.heliyon.2021.e07919
64. He Y, Valouev A, Xiong L, et al. Highly sensitive blood-based multi-cancer screening device with tiered specificity based on diagnostic workflow. Cancer Research Conference: American Association for Cancer Research Annual Meeting, ACCR 2023;83 doi: https://dx.doi.org/10.1158/1538-7445.AM2023-3331
65. Hinestrosa JP, Kurzrock R, Lewis JM, et al. Early-stage multi-cancer detection using an extracellular vesicle protein-based blood test. Communication medicale 2022;2:29. doi: https://dx.doi.org/10.1038/s43856-022-00088-6
66. Hongling Y, Qian Z, Qunzhi Z, et al. The diagnoThe diagnostic accuracy of liquid exosomal miRNAs for cancer detection: a meta-analysis. 2020
67. Horst C, Dickson J, Tisi S, et al. P41.04 The SUMMIT Study: Pulmonary Nodule and Incidental Findings in the First 10,000 Participants of a Population-Based Low-Dose CT Screening Study. Journal of Thoracic Oncology 2021;16(3 Supplement):S473-S74. doi: https://dx.doi.org/10.1016/j.jtho.2021.01.818
68. Hsieh JCH, Liao CT, Wang HM, et al. Evaluation of circulating miRNAs for earlier cancer detection through machine-learning expression profiling. Journal of Clinical Oncology Conference 2020;38 doi: https://dx.doi.org/10.1200/JCO.2020.38.15_suppl.1559
69. Huang JY, Soupir AC, Schlick BD, et al. Cancer Detection and Classification by CpG Island Hypermethylation Signatures in Plasma Cell-Free DNA. Cancers 2021;13:18. doi: https://dx.doi.org/10.3390/cancers13225611
70. Jain V, Chaitali W, Namrata B. Quantitation of total circulating cell-free DNA as a screening modality for cancer. Pravara Medical Review 2019;11(4):14-20.
71. Jamshidi A, Liu MC, Klein EA, et al. Evaluation of cell-free DNA approaches for multi-cancer early detection. Cancer Cell 2022;40:1537-49.e12. doi: https://dx.doi.org/10.1016/j.ccell.2022.10.022
72. Jones C, Gray E, Gavan S, et al. A systematic review of model-based economic evaluations of stratified early detection interventions in cancer; taking account of risk-estimation, threshold setting and clinical protocols. 2019
73. Jurmeister P. [Early diagnosis and localization of cancer by liquid biopsy]. Pathologe 2018;39:328-29. doi: https://dx.doi.org/10.1007/s00292-018-0454-6
74. Katerov S, Vaccaro A, Hennek J, et al. Accurate multi-cancer detection using methylated DNA markers and proteins in plasma. Cancer Research Conference: AACR Annual Meeting 2021;81 doi: https://dx.doi.org/10.1158/1538-7445.AM2021-111
75. Kct. A Prospective, Multi-center Clinical Study to Establish Multi-Cancer Early Detection Platform through the Analysis of Whole Genome Sequencing of Circulating DNA in Cancer Patients and Healthy Volunteers, 2023.
76. Kim A, Chung KC, Keir C, et al. PCN225 Patient-Reported Outcomes Associated with Cancer Screening: A Systematic Review. Value in Health 2021;24(Supplement 1):S62. doi: https://dx.doi.org/10.1016/j.jval.2021.04.315
77. Kinross J, Kruusmaa K, Bitenc M, et al. A panel of methylation markers for multi-cancer detection from plasma. Annals of Oncology 2020;31(Supplement 4):S280. doi: https://dx.doi.org/10.1016/j.annonc.2020.08.218
78. Klein EA, Hubbell E, Maddala T, et al. Development of a comprehensive cell-freeDNA (cfDNA) assay for early detection of multiple tumor types: The Circulating Cellfree Genome Atlas (CCGA) study. Journal of Clinical Oncology Conference 2018;36 doi: https://dx.doi.org/10.1200/JCO.2018.36.15_suppl.12021
79. Kurtzman K, Oxnard G, Klein E, et al. PR01.08 Simultaneous Multi-Cancer Detection and Tissue of Origin Prediction Via Targeted Bisulfite Sequencing of Plasma Cell-Free DNA. Journal of Thoracic Oncology 2021;16(1 Supplement):S43-S44. doi: https://dx.doi.org/10.1016/j.jtho.2020.10.085
80. Kurtzman KN, Bryce AH, Liu MC, et al. Multi-cancer detection test to aid head and neck cancer diagnosis. Otolaryngology - Head and Neck Surgery 2021;165(1 SUPPL):P211-P12. doi: https://dx.doi.org/10.1177/01945998211030910d
81. Kurtzman KN, Oxnard G, Klein E, et al. Multi-Cancer Detection of Early-Stage Cancers with Simultaneous Tissue Localization Using a Plasma Circulating Tumor Cell-Free DNA-Based Targeted Methylation Assay. Gastroenterology 2020;158(6 Supplement 1):S-642. doi: https://dx.doi.org/10.1016/S0016-5085%2820%2932300-3
82. Li B, Su J, Zhang G, et al. Analytical performance of ELSA-seq, a bloodbased test for early detection of multiple cancers. Cancer Research Conference: American Association for Cancer Research Annual Meeting, ACCR 2020;82 doi: https://dx.doi.org/10.1158/1538-7445.AM2022-5116
83. Liu L, Toung JM, Jassowicz AF, et al. Targeted methylation sequencing of plasma cell-free DNA for cancer detection and classification. Annals of Oncology 2018;29:1445-53. doi: https://dx.doi.org/10.1093/annonc/mdy119
84. Liu L, Toung JM, Vijayaraghavan R, et al. A highly sensitive method for noninvasive cancer profiling through targeted methylation sequencing of circulating cell-free DNA. Cancer Research Conference: American Association for Cancer Research Annual Meeting 2017;77 doi: https://dx.doi.org/10.1158/1538-7445.AM2017-5381
85. Liu MC, Bryce AH, Seiden MV, et al. Performance of a Multi-Cancer Detection Test as a Tool for Diagnostic Resolution of Symptomatic Gynecological Cancers. Journal of Minimally Invasive Gynecology 2021;28(11 Supplement):S45-S46. doi: https://dx.doi.org/10.1016/j.jmig.2021.09.407
86. Liu MC, Jamshidi A, Klein EA, et al. 1123O Evaluation of cell-free DNA approaches for multi-cancer early detection. Annals of Oncology 2021;32(Supplement 5):S921. doi: https://dx.doi.org/10.1016/j.annonc.2021.08.765
87. Liu MC, Jamshidi A, Venn O, et al. Genome-wide cell-free DNA (cfDNA) methylation signatures and effect on tissue of origin (TOO) performance. Journal of Clinical Oncology Conference 2019;37 doi: https://dx.doi.org/10.1200/JCO.2019.37.15-suppl.3049
88. Liu MC, Klein E, Hubbell E, et al. Plasma cell-free DNA (cfDNA) assays for early multi-cancer detection: The circulating cell-free genome atlas (CCGA) study. Annals of Oncology 2018;29(Supplement 8):viii14. doi: https://dx.doi.org/10.1093/annonc/mdy268.048
89. Liu MC, Oxnard GR, Klein EA, et al. Sensitive and specific multi-cancer detection and localization using methylation signatures in cell-free DNA. Annals of Oncology 2020;31:745-59. doi: https://dx.doi.org/10.1016/j.annonc.2020.02.011
90. Liu Q, Shaknovich R, Chen X, et al. cfDNA methylation profiling distinguishes lineage-specific hematologic malignancies. Cancer Research Conference: American Association for Cancer Research Annual Meeting, AACR 2020;80 doi: https://dx.doi.org/10.1158/1538-7445.AM2020-139
91. Nakles-Taylor R, Rosenthal SH, Cheng LL, et al. Frequency of pathogenic and likely pathogenic variants in breast and ovarian cancer genes identified in a 34-gene hereditary multi-cancer panel at a diagnostic reference laboratory. Familial Cancer 2022;21(3):281. doi: https://dx.doi.org/10.1007/s10689-021-00273-x
92. NCT. Prospective Screening Programme for Malignant Tumors. In: First Affiliated Hospital SY-SU, Sixth Affiliated H, Sun Yat-sen U, eds., 2020.
93. NCT. AssesSment of Early-deteCtion basEd oN liquiD Biopsy in Hepatobiliary Cancer Malignancies. In: Guangzhou BR, ed., 2021.
94. NCT. The Unintrusive Detection of EaRly-stage Cancers. In: Guangzhou Burning Rock Dx C, Ltd, eds., 2021.
95. Nguyen H, Raymond VM, Vento-Gaudens E, et al. Screening for high frequency malignant disease (SHIELD). Journal of Clinical Oncology Conference: Annual Meeting of the American Society of Clinical Oncology, ASCO 2022;40 doi: https://dx.doi.org/10.1200/JCO.2022.40.16_suppl.TPS1602
96. Nimgaonkar A, Segurado O, Tsai WS, et al. A novel circulating tumor cell blood test for early detection of colorectal, prostate, and breast cancers: Results from 709 samples. Journal of Clinical Oncology Conference 2018;36 doi: https://dx.doi.org/10.1200/JCO.2018.36.15_suppl.e13549
97. Nunes SP, Moreira-Barbosa C, Salta S, et al. Cell-Free DNA Methylation of Selected Genes Allows for Early Detection of the Major Cancers in Women. Cancers 2018;10:26. doi: https://dx.doi.org/10.3390/cancers10100357
98. Oxnard GR, Klein EA, Seiden M, et al. Simultaneous multi-cancer detection and tissue of origin (TOO) localization using targeted bisulfite sequencing of plasma cell-free DNA (cfDNA). Journal of Global Oncology 2019;5(Supplement):44. doi: https://dx.doi.org/10.1200/JGO.2019.5.suppl.44
99. Oxnard GR, Klein EA, Seiden MV, et al. Simultaneous multi-cancer detection and tissue of origin (TOO) localization using targeted bisulfite sequencing of plasma cell-free DNA (cfDNA). Annals of Oncology 2019;30(Supplement 5):v912. doi: https://dx.doi.org/10.1093/annonc/mdz394.074
100. Prieur A, Kepenekian V, Mazard T, et al. Progastrin, a new blood biomarker for multiple cancers allowing a new strategy for screening, early detection and monitoring. Journal of Global Oncology 2018;4(Supplement 2):211s. doi: https://dx.doi.org/10.1200/jgo.18.85400
101. Prieur A, Mazard T, Assenat E, et al. Progastrin: A new specific early cancer screening biomarker. Journal of Clinical Oncology Conference 2017;35
102. Quagliarini E, Digiacomo L, Caputo D, et al. Magnetic Levitation of Personalized Nanoparticle-Protein Corona as an Effective Tool for Cancer Detection. Nanomaterials 2022;12:19. doi: https://dx.doi.org/10.3390/nano12091397
103. Raymond V, Nguyen H, Cotton L, et al. PP01.20 Trial in Progress: Screening for High Frequency Malignant Disease (SHIELD). Journal of Thoracic Oncology 2023;18(3 Supplement):e19-19. doi: https://dx.doi.org/10.1016/j.jtho.2022.09.046
104. Ris F, Hellan M, Douissard J, et al. Blood-Based Multi-Cancer Detection Using a Novel Variant Calling Assay (DEEPGENTM): Early Clinical Results. Cancers 2021;13:15. doi: https://dx.doi.org/10.3390/cancers13164104
105. Roy D, Taggart D, Zheng L, et al. Circulating cell-free DNA methylation assay: Towards early detection of multiple cancer types. Cancer Research Conference: American Association for Cancer Research Annual Meeting 2019;79 doi: https://dx.doi.org/10.1158/1538-7445.SABCS18-837
106. Saman S, Stagno MJ, Warmann SW, et al. Biomarkers Apo10 and TKTL1: Epitope-detection in monocytes (EDIM) as a new diagnostic approach for cholangiocellular, pancreatic and colorectal carcinoma. Cancer Biomarkers: Section A of Disease Markers 2020;27:129-37. doi: https://dx.doi.org/10.3233/CBM-190414
107. Schwaederle M, Husain H, Fanta PT, et al. Detection rate of actionable mutations in diverse cancers using a biopsy-free (blood) circulating tumor cell DNA assay. Oncotarget 2016;7:9707-17. doi: https://dx.doi.org/10.18632/oncotarget.7110
108. Seneviratne L, Evans S, Pulicharam J, et al. Discovery of a core-panel of markers for a blood-assay for cancer detection utilizing cf DNA methylation changes. Journal of Clinical Oncology Conference 2020;38 doi: https://dx.doi.org/10.1200/JCO.2020.38.15_suppl.1522
109. Shao Y, Bao H, Wang Z, et al. An ultra-sensitive assay using cell-free DNA fragmentomics for multi-cancer early detection. Journal of Clinical Oncology Conference: Annual Meeting of the American Society of Clinical Oncology, ASCO 2022;40 doi: https://dx.doi.org/10.1200/JCO.2022.40.16_suppl.3037
110. Song G, Wang L, Tang J, et al. Circulating metabolites as potential biomarkers for the early detection and prognosis surveillance of gastrointestinal cancers. Metabolomics 2023;19:36. doi: https://dx.doi.org/10.1007/s11306-023-02002-0
111. Stackpole M, Zeng W, Liu CC, et al. Multi-feature ensemble learning on cell-free dna for accurately detecting and locating cancer. Cancer Research Conference: AACR Annual Meeting 2021;81 doi: https://dx.doi.org/10.1158/1538-7445.AM2021-24
112. Suo C, Zhao R, Jiang Y, et al. The FuSion Project of Pan-Cancer Early Screening in Chinese-An integrative study by Fudan University and Singlera. Cancer Research Conference: American Association for Cancer Research Annual Meeting, ACCR 2023;83 doi: https://dx.doi.org/10.1158/1538-7445.AM2023-4194
113. Thierry AR, Tanos R, Otandault A, et al. Towards a screening test for cancer by circulating DNA analysis. Journal of Clinical Oncology Conference 2019;37 doi: https://dx.doi.org/10.1200/JCO.2019.37.15_suppl.e13146
114. Tisi S, Dickson J, Horst C, et al. SUMMIT study: protocolised management of pulmonary incidental findings in a lung cancer screening cohort. Lung Cancer 2020;139(Supplement 1):S5. doi: https://dx.doi.org/10.1016/S0169-5002%2820%2930039-8
115. Tomeva E, Switzeny OJ, Heitzinger C, et al. Comprehensive Approach to Distinguish Patients with Solid Tumors from Healthy Controls by Combining Androgen Receptor Mutation p.H875Y with Cell-Free DNA Methylation and Circulating miRNAs. Cancers 2022;14:17. doi: https://dx.doi.org/10.3390/cancers14020462
116. Valouev A, Zotenko E, Snyder M, et al. Development of a highly sensitive multicancer, targeted, cell-free DNA epigenomic assay for integrated screening of lung and colorectal cancer. Journal of Clinical Oncology Conference: Annual Meeting of the American Society of Clinical Oncology, ASCO 2022;40 doi: https://dx.doi.org/10.1200/JCO.2022.40.16_suppl.3542
117. Wang F, Li X, Li M, et al. Ultra-short cell-free DNA fragments enhance cancer early detection in a multi-analyte blood test combining mutation, protein and fragmentomics. Clinical Chemistry & Laboratory Medicine 2023;08:08. doi: https://dx.doi.org/10.1515/cclm-2023-0541
118. Wang R, Wen H, Xu YC, et al. Circulating MicroRNAs as a Novel Class of Diagnostic Biomarkers in Gastrointestinal Tumors Detection: A Meta-Analysis Based on 42 Articles. Plos One 2014;9:13. doi: https://dx.doi.org/10.1371/journal.pone.0113401
119. Wen H, Feng Z, Ge H, et al. Multi-cancer early detection in gynaecological malignancies based on integrating multi-omics assays by liquid biopsy: A prospective study. Annals of Oncology 2022;33(Supplement 7):S821-S22. doi: https://dx.doi.org/10.1016/j.annonc.2022.07.731
120. Wen YH, Chang PY, Hsu CM, et al. Cancer screening through a multi-analyte serum biomarker panel during health check-up examinations: Results from a 12-year experience. Clinica Chimica Acta 2015;450:273-6. doi: https://dx.doi.org/10.1016/j.cca.2015.09.004
121. Wolpin BM, Richards DA, Cohn AL, et al. Performance of a blood-based test for the detection of multiple cancer types. Journal of Clinical Oncology Conference 2020;38 doi: https://dx.doi.org/10.1200/JCO.2020.38.4_suppl.283
122. Wong D, Luo P, Oldfield L, et al. Integrated analysis of cell-free DNA for the early detection of cancer in people with Li-Fraumeni Syndrome. medRxiv 2022;11 doi: https://dx.doi.org/10.1101/2022.10.07.22280848
123. Zhang A, Hu H. Development and validation of a novel circulating cell-free microRNA diagnostic model with high accuracy for multi-cancer early detection. Cancer Research Conference: American Association for Cancer Research Annual Meeting, ACCR 2020;82 doi: https://dx.doi.org/10.1158/1538-7445.AM2022-5931
124. Zhang A, Hu H. A Novel Blood-Based microRNA Diagnostic Model with High Accuracy for Multi-Cancer Early Detection. Cancers 2022;14:11. doi: https://dx.doi.org/10.3390/cancers14061450
125. Zhang H, Zhao L, Jiang J, et al. Multiplexed nanomaterial-assisted laser desorption/ionization for pan-cancer diagnosis and classification. Nature communications 2022;13:617. doi: https://dx.doi.org/10.1038/s41467-021-26642-9
126. Zhang Y, Zhao H, Bi X, et al. Evaluation of a multi-level, multi-parameter detection method for digestive system cancer diagnosis. Journal of Clinical Oncology Conference 2015;33 doi: https://dx.doi.org/10.1200/jco.2015.33.15_suppl.e12578
127. Zhang YL, Yao Y, Xu YP, et al. Pan-cancer circulating tumor DNA detection in over 10,000 Chinese patients (vol 12, 11, 2021). Nature Communications 2021;12:1. doi: https://dx.doi.org/10.1038/s41467-021-21285-2
128. Zhang Z, Chang WJ, Cai JB, et al. Multi-cancer detection and tissue of origin determination based on 5-hydroxymethylcytosine biomarkers in circulating cell-free DNA. Journal of Clinical Oncology 2021;39:2. doi: https://dx.doi.org/10.1200/JCO.2021.39.15_suppl.3123
129. Zheng J, Li Z, Jiang R, et al. Development of a novel liquid biopsy test to diagnose and locate gastrointestinal cancers. Journal of Clinical Oncology Conference 2020;38 doi: https://dx.doi.org/10.1200/JCO.2020.38.15_suppl.1557
130. Zhou X, Cheng Z, Dong MY, et al. Tumor fractions deciphered from circulating cell-free DNA methylation for cancer early diagnosis. Nature Communications 2022;13:13. doi: https://dx.doi.org/10.1038/s41467-022-35320-3

### Excluded on study design (n=29)

1. Agency for Healthcare R, Quality. Role of Liquid Biopsy in Detection and Management of Cancer in the Medicare Population ID: MYOE58. 2021
2. Cervena K, Vodicka P, Vymetalkova V. Diagnostic and prognostic impact of cell-free DNA in human cancers: Systematic review. *Mutation Research-Reviews in Mutation Research* 2019;781:100-29. doi: https://dx.doi.org/10.1016/j.mrrev.2019.05.002
3. Chang ET, Hubbell E, Klein EA. Multicancer Early Detection. *Clinical Gastroenterology & Hepatology* 2023;07:07. doi: https://dx.doi.org/10.1016/j.cgh.2023.03.039
4. Cohen S, Reichert H, Kansal AR, et al. Pcn272 Improved Efficiency of Cancer Screening with a Multi-Cancer Early Detection Test. *Value in Health* 2020;23(Supplement 1):S71. doi: https://dx.doi.org/10.1016/j.jval.2020.04.1738
5. Cohn AL, Seiden M, Kurtzman KN, et al. The Circulating Cell-free Genome Atlas (CCGA) Study: Follow-up (F/U) on non-cancer participants with cancer-like cell-free DNA signals. *Journal of Clinical Oncology Conference* 2019;37 doi: https://dx.doi.org/10.1200/JCO.2019.37.15_suppl.5574
6. Dive C. Liquid biopsies for the management of cancer patient treatment and for early detection of cancer. *Molecular Cancer Therapeutics Conference: AACR NCI EORTC International Conference: Molecular Targets and Cancer Therapeutics* 2017;17 doi: https://dx.doi.org/10.1158/1535-7163.TARG-17-CN08-03
7. Fagery M, Ijzerman M, Khorshidi Hadi A, et al. Clinical Evidence of Multi-cancer Early Detection (MCED) Blood-based Liquid Biopsy for Early Cancer Detection: A Systematic Literature Review. 2023
8. Hackshaw A, Berg CD. An efficient randomised trial design for multi-cancer screening blood tests: nested enhanced mortality outcomes of screening trial (vol 22, pg 1360, 2021). *Lancet Oncology* 2021;22:E472-E72.
9. Hackshaw A, Cohen SS, Reichert H, et al. Estimating the population health impact of a multi-cancer early detection genomic blood test to complement existing screening in the US and UK. *British Journal of Cancer* 2021;125:1432-42. doi: https://dx.doi.org/10.1038/s41416-021-01498-4
10. Hanna M, Dey N, Grady WM. Response to letter about Multicancer Early Detection Assays. *Clinical gastroenterology and hepatology: the official clinical practice journal of the American Gastroenterological Association* 2023;19 doi: https://dx.doi.org/10.1016/j.cgh.2023.05.012
11. Hubbell E, Clarke CA, Aravanis AM, et al. Modeled Reductions in Late-stage Cancer with a Multi-Cancer Early Detection Test. *Cancer Epidemiology, Biomarkers & Prevention* 2021;30:460-68. doi: https://dx.doi.org/10.1158/1055-9965.EPI-20-1134
12. Jia S, Xie L, Li L, et al. The values of liquid biopsy as a screening tool of cancer: a systematic review. 2020
13. Jia S, Xie L, Li L, et al. Values of liquid biopsy in early detection of cancer: results from meta-analysis. *Expert Review of Molecular Diagnostics* 2021;21:417-27. doi: https://dx.doi.org/10.1080/14737159.2021.1910025
14. Jiao B, Gulati R, Katki HA, et al. A Quantitative Framework to Study Potential Benefits and Harms of Multi-Cancer Early Detection Testing. *Cancer Epidemiology, Biomarkers & Prevention* 2022;31:38-44. doi: https://dx.doi.org/10.1158/1055-9965.EPI-21-0380
15. Jørgensen Nanna E, Sopina L. Economic evaluations of ctDNA in cancer diagnosis: a systematic review protocol. 2022
16. Kansal A, Shaul A, Ye W, et al. Cost-effectiveness of a multi-cancer early detection test in individuals with a personal or family history of cancer. *Journal of Managed Care and Specialty Pharmacy* 2022;28(10 A-Supplement):S117-S18.
17. Kim A, Cong Z, Jazieh A, et al. Estimating the incremental population health impact of a multi-cancer early detection test to complement existing screening among populations with an elevated risk for cancer with additional risk factors in the United States. *Journal of Managed Care and Specialty Pharmacy* 2022;28(10 A-Supplement):S35.
18. Klein EA. Re: Sabrina H. Rossi, Grant D. Stewart. Re: Clinical Validation of a Targeted Methylation-based Multi-cancer Early Detection Test Using an Independent Validation Set. Eur Urol. 2022;82:442-443. *European Urology* 2022;82:e144. doi: https://dx.doi.org/10.1016/j.eururo.2022.07.033
19. Kramer A, Schuuring E, Vessies DCL, et al. A Micro-Costing Framework for Circulating Tumor DNA Testing in Dutch Clinical Practice. *Journal of Molecular Diagnostics* 2023;25:36-45. doi: https://dx.doi.org/10.1016/j.jmoldx.2022.10.004
20. Lin GA, Phillips KA, Fendrick AM. Reading the crystal ball: Primary care implications while awaiting outcomes for multi-cancer early detection tests. *Healthcare* 2023;11:100705. doi: https://dx.doi.org/10.1016/j.hjdsi.2023.100705
21. Nakamura Y. SY22-4 Current and future paradigms of liquid biopsy for cancer care. *Annals of Oncology* 2022;33(Supplement 6):S446. doi: https://dx.doi.org/10.1016/j.annonc.2022.05.484
22. Oh Y, Park JH, Chung LIY, et al. Systematic review and meta-analysis of the accuracy of tumor origin detection in blood-based multicancer early detection (MCED) in the general population. *Cancer Research Conference: American Association for Cancer Research Annual Meeting, ACCR* 2023;83 doi: https://dx.doi.org/10.1158/1538-7445.AM2023-784
23. Ortendahl J, Lee J, Hubbell E, et al. Pcn2 Projected Lifetime Clinical Value of a Multicancer Early Detection Test. *Value in Health* 2020;23(Supplement 1):S22. doi: https://dx.doi.org/10.1016/j.jval.2020.04.1510
24. Park JH, Oh Y, Chung LIY, et al. Systematic review and meta-analysis of the accuracy and applicability of blood-based multi-cancer early detection (MCED) in the general population. *Cancer Research Conference: American Association for Cancer Research Annual Meeting, ACCR* 2023;83 doi: https://dx.doi.org/10.1158/1538-7445.AM2023-783
25. Rodríguez-Ces Ana M, Rapado-González Ó, Salgado-Barreira Á, et al. Liquid biopsies based on cell-free DNA integrity as a biomarker for cancer diagnosis. 2021
26. Sasieni P, Smittenaar R, Hubbell E, et al. Modelled mortality benefits of multi-cancer early detection screening in England. *British Journal of Cancer* 2023;129:72-80. doi: https://dx.doi.org/10.1038/s41416-023-02243-9
27. Tafazzoli A, Ramsey SD, Shaul A, et al. The Potential Value-Based Price of a Multi-Cancer Early Detection Genomic Blood Test to Complement Current Single Cancer Screening in the USA. *Pharmacoeconomics* 2022;40:1107-17. doi: https://dx.doi.org/10.1007/s40273-022-01181-3
28. Uhe I, Hagen ME, Ris F, et al. Cell-free DNA liquid biopsy for early detection of gastrointestinal cancers: A systematic review. *World Journal of Gastrointestinal Oncology* 2021;13:1799-812. doi: https://dx.doi.org/10.4251/wjgo.v13.i11.1799
29. Zhang L, Shen M, Wei Y, et al. A systematic review of cost-effectiveness of multi-cancer screening. 2023

### Excluded on population (n=4)

1. Early Detection of de Novo Cancer in Liver Transplant Recipients.
2. Garcia-Corbacho J, Ruiz IV, Angelats L, et al. First-results of the CLIMB360 study, a prospective molecular screening program across multiple cancer types based on circulating tumor DNA (ctDNA). *Annals of Oncology* 2021;32:S396-S97. https://dx.doi.org/10.1016/j.annonc.2021.08.372
3. Horst C, Dickson J, Tisi S, et al. SUMMIT study: protocolised management of pulmonary nodules in a lung cancer screening cohort. *Lung Cancer* 2020;139(Supplement 1):S3. doi: https://dx.doi.org/10.1016/S0169-5002%2820%2930034-9
4. Jassowicz A, Liu L, Huang H, et al. Targeted methylation sequencing of plasma cell-free DNA identifies patients with advanced breast, colorectal, non-small cell lung cancer, melanoma with poor outcomes. *Annals of Oncology* 2017;28(Supplement 5):v34-v35. doi: https://dx.doi.org/10.1093/annonc/mdx363.041

### Excluded on outcomes (n=12)

1. Detecting Cancers Earlier Through Elective Plasma-based CancerSEEK Testing.
2. Drks. Detecting cancers Earlier Through Elective plasma-based CancerSEEK Testing – Ascertaining Serial Cancer patients to Enable New Diagnostic II (DETECT-ASCEND2), 2023.
3. Jones C, Parker A, Warren A, et al. Mammography utilization among women with a negative circulating tumor DNA-based early cancer detection test. *Journal of Clinical Oncology Conference* 2020;38 doi: https://dx.doi.org/10.1200/JCO.2020.38.15_suppl.1563
4. Lim S, Guneta V, Chow S, et al. Synergizing Biobanking Processes Between Academia And Commercial Biobanks. *Biopreservation and Biobanking* 2023;21(3):A50. doi: https://dx.doi.org/10.1089/bio.2023.29118.abstracts
5. Sasieni P, Clarke CA, Hubbell E. 1135P Impact of MCED screening interval on reduction in late-stage cancer diagnosis and mortality. *Annals of Oncology* 2021;32(Supplement 5):S925. doi: https://dx.doi.org/10.1016/j.annonc.2021.08.777
6. Tafazzoli A, Ramsey SD, Shaul A, et al. POSB44 Drivers of Value-Based Price (VBP) for a Multi-Cancer Early Detection (MCED) Test. *Value in Health* 2022;25(1 Supplement):S68. doi: https://dx.doi.org/10.1016/j.jval.2021.11.317
7. Tafazzoli A, Ramsey SD, Shaul A, et al. EE5 Assessment of Value Based Price (VBP) for a Multi-Cancer Early Detection (MCED) Test in a Medicare Population. *Value in Health* 2022;25(7 Supplement):S335-S36. doi: https://dx.doi.org/10.1016/j.jval.2022.04.257
8. Vrba L, Futscher B, Bernert R, et al. Sentinel-10: A New Multi-Cancer Early Detection Test. *Journal of Molecular Diagnostics* 2022;24:S107-S07.
9. Xu L, Wang J, Ma W, et al. Validation of a high performing blood test for multiple major cancer screenings. *Journal of Clinical Oncology Conference: Annual Meeting of the American Society of Clinical Oncology, ASCO* 2021;39 doi: https://dx.doi.org/10.1200/JCO.2021.39.15-suppl.10561
10. Xu L, Wang J, Yang T, et al. Toward the development of a $100screening test for 6 major cancer types. *Cancer Research Conference: American Association for Cancer Research Annual Meeting, AACR* 2020;80 doi: https://dx.doi.org/10.1158/1538-7445.AM2020-4601
11. Xu LH, Wang J, Ma WF, et al. A high performance blood test for multiple cancer early screening. *Cancer Research* 2021;81:1.
12. You B, Kepenekian V, Prieur A, et al. Progastrin, a new blood biomarker for the diagnostic and therapeutic monitoring, in gastro-intestinal cancers: A BIG-RENAPE project. *Annals of Oncology* 2018;29(Supplement 8):viii37. doi: https://dx.doi.org/10.1093/annonc/mdy269.117

### Excluded duplicate (n=1)

1. Marlow L, Schmeising-Barnes N, Warwick J, et al. Psychological Impact of the Galleri Test (sIG(n)al): Protocol for a longitudinal evaluation of the psychological impact of receiving a cancer signal in the NHS-Galleri Trial. *medRxiv* 2023;12 doi: https://dx.doi.org/10.1101/2023.06.12.23291276

## APPENDIX 5. DETAILS OF THE INCLUDED STUDIES

**Table 11.**
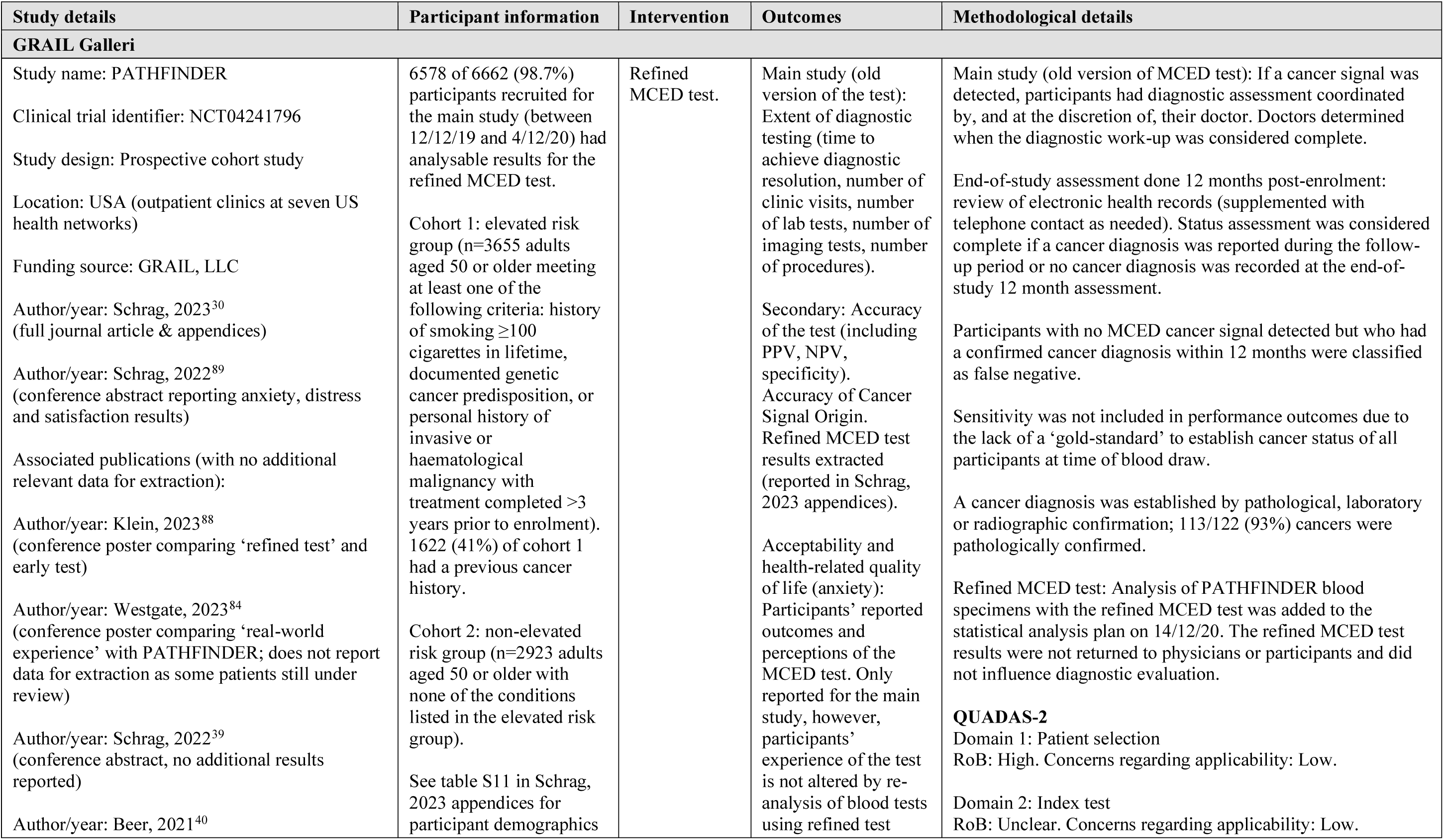
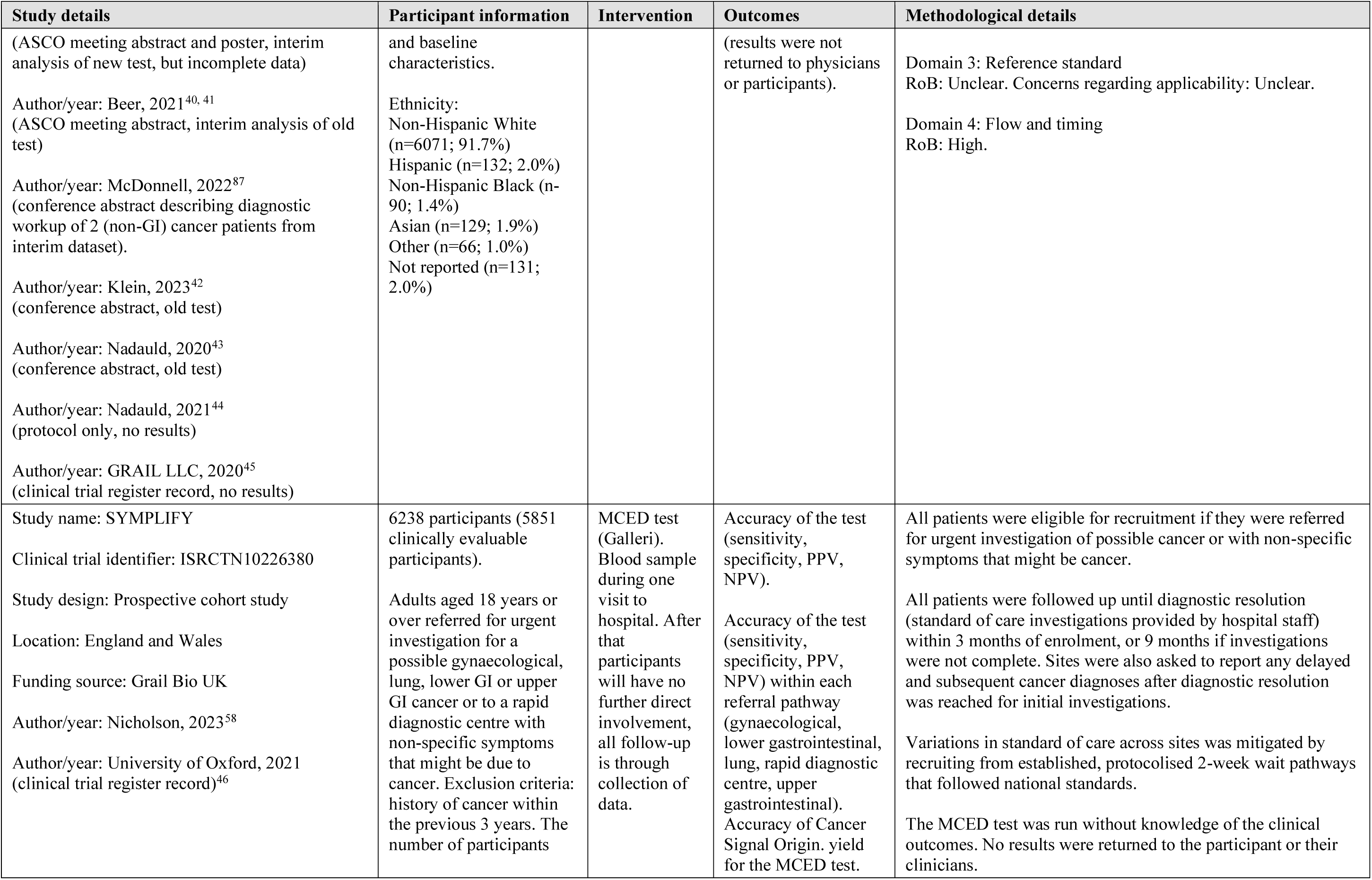
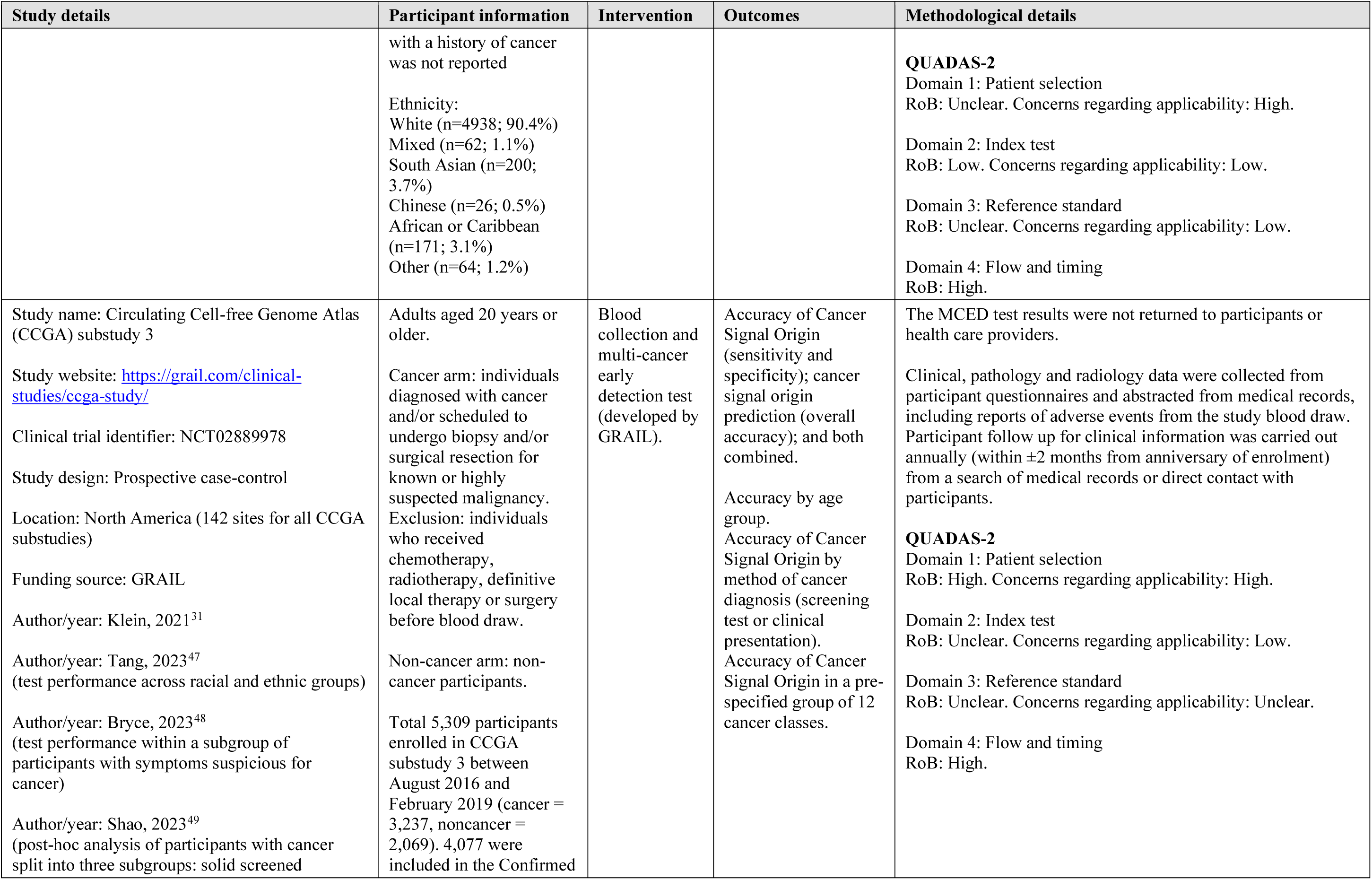
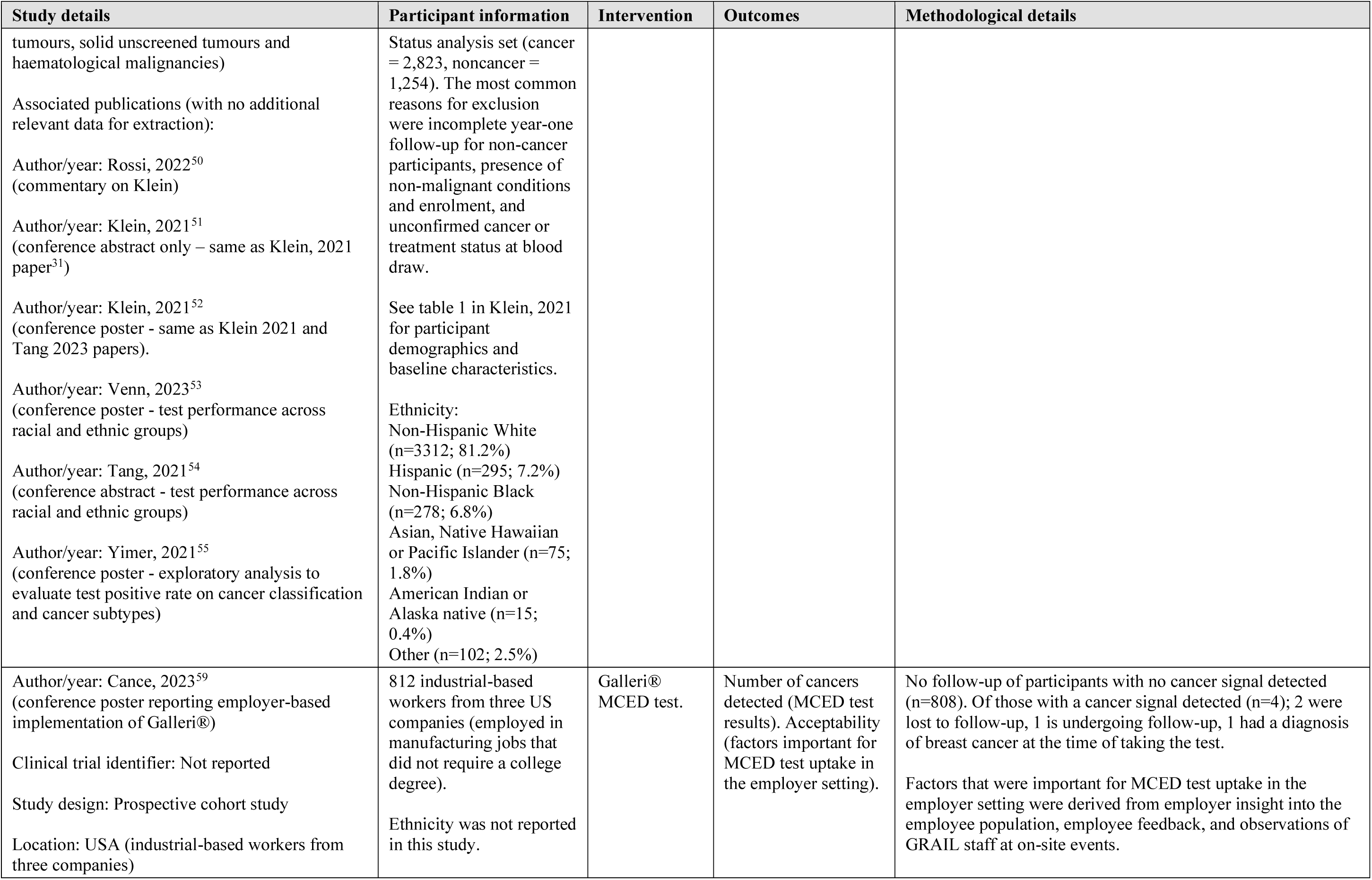
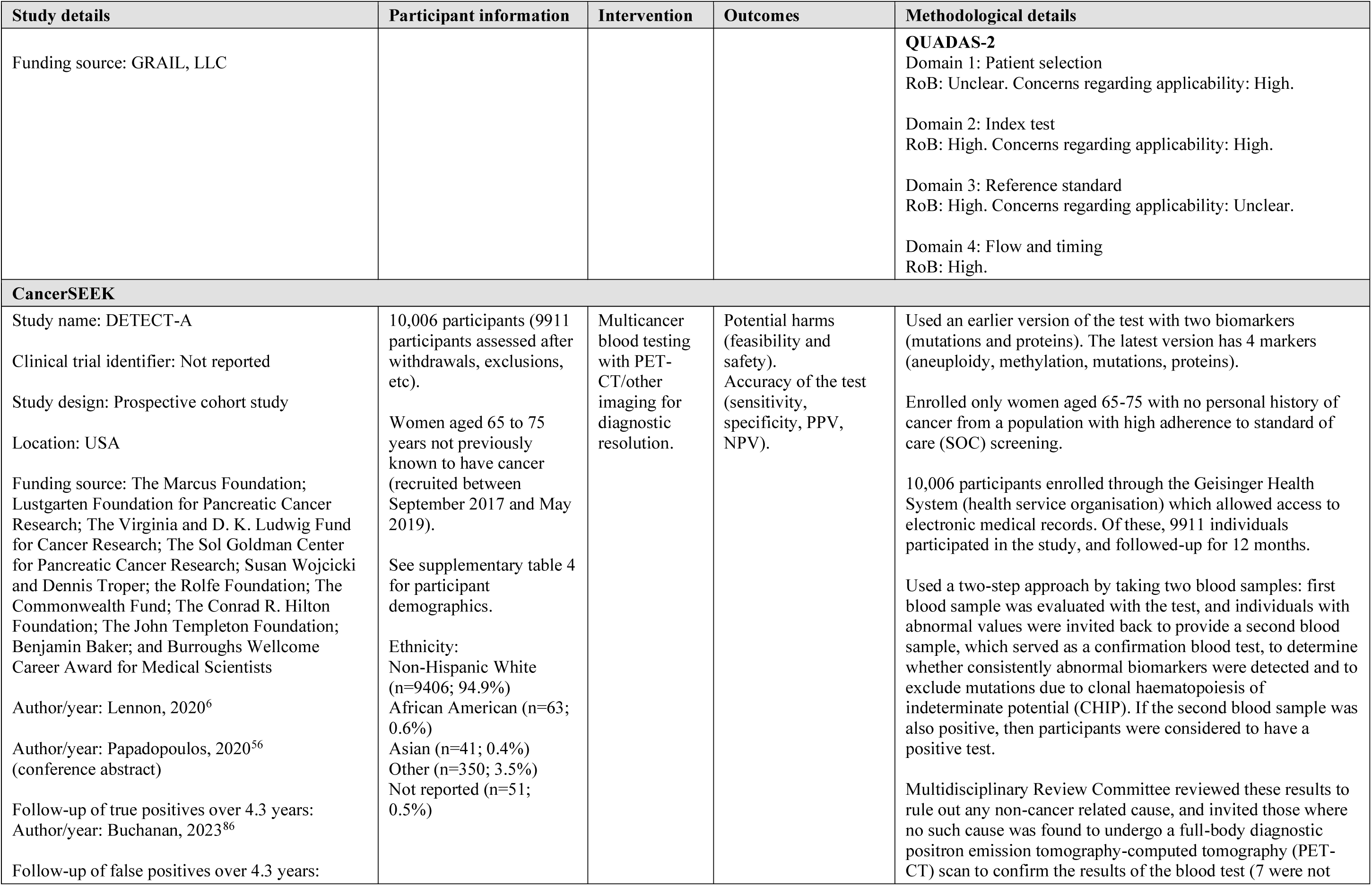
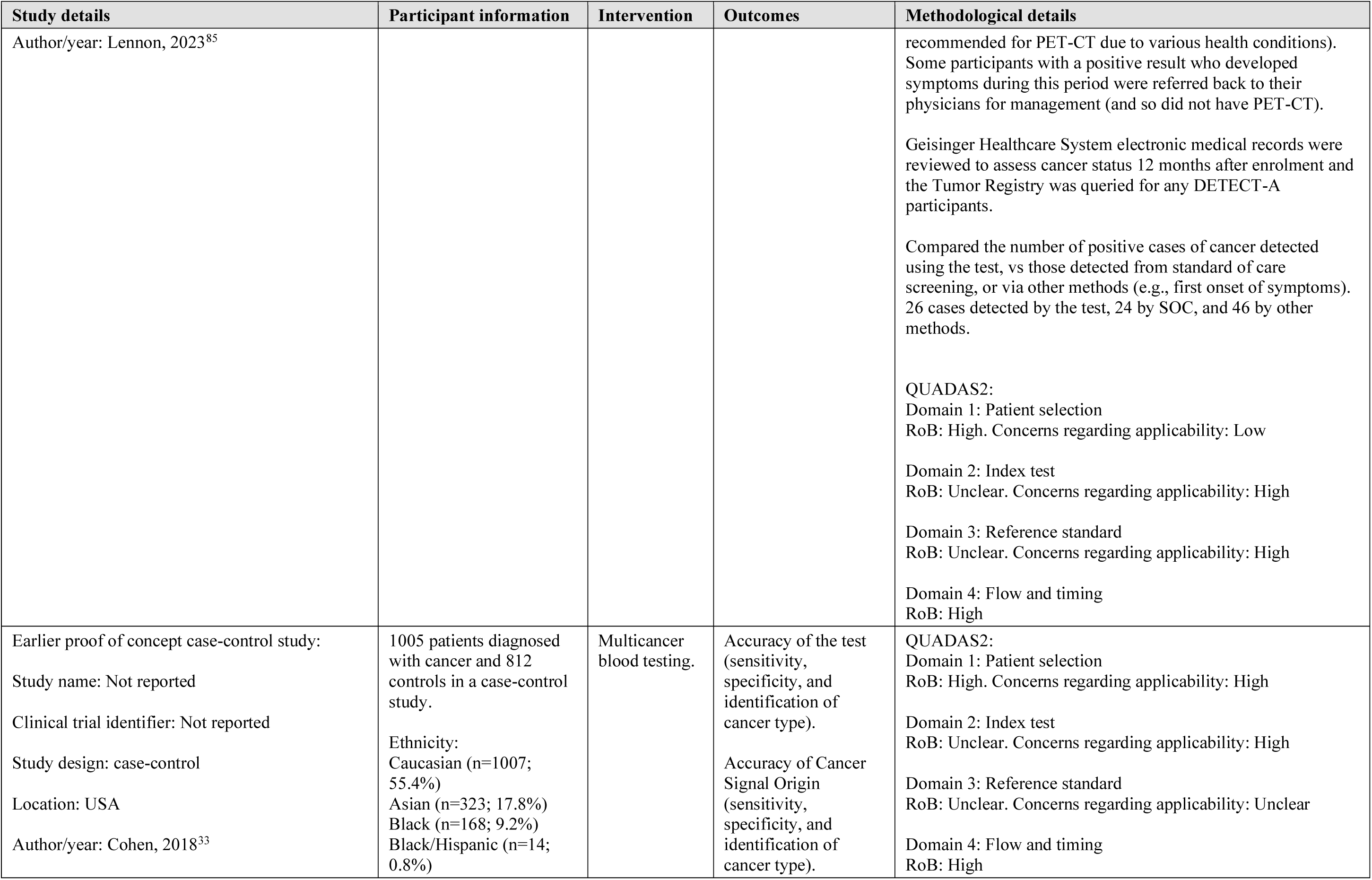
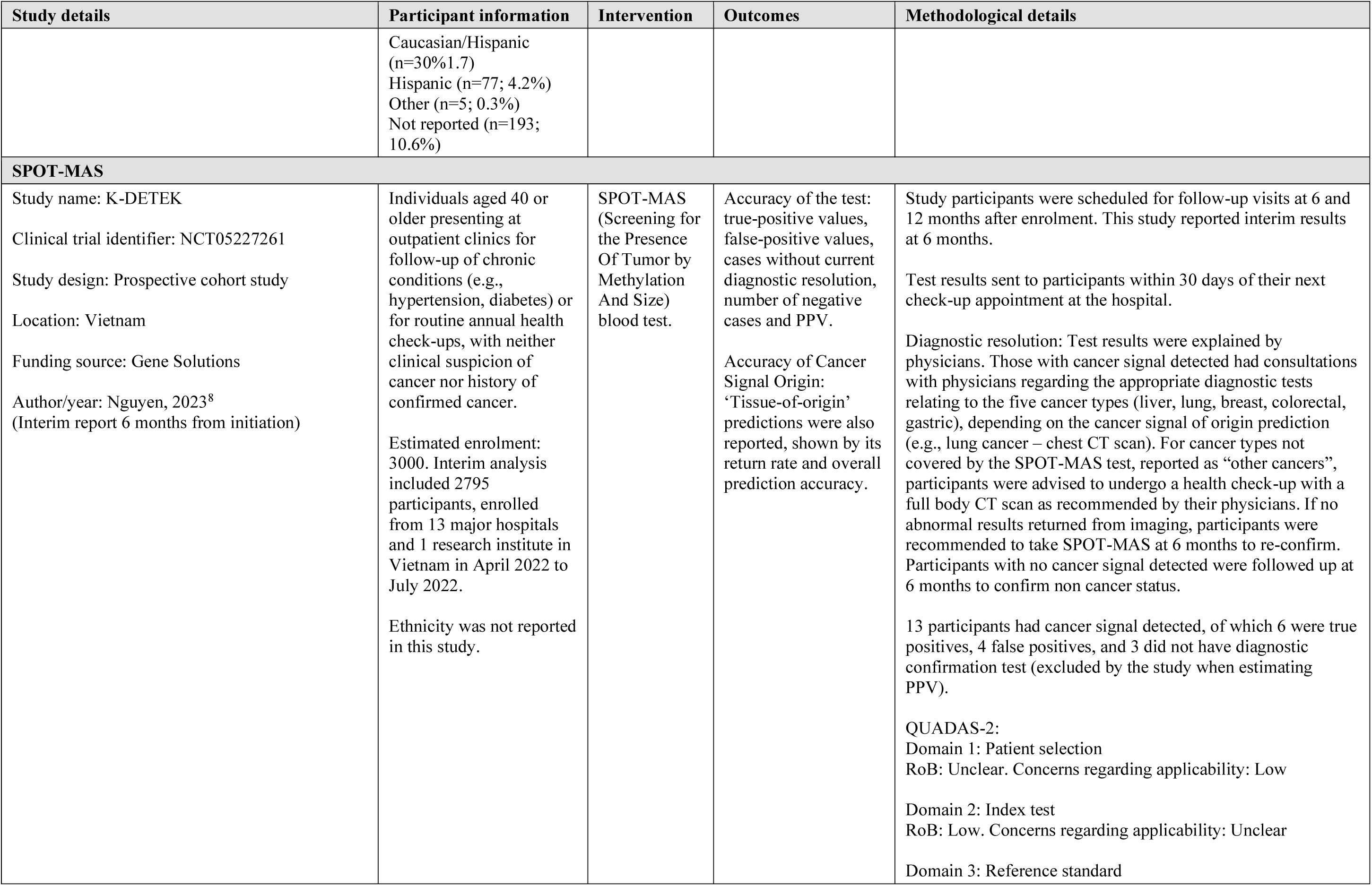
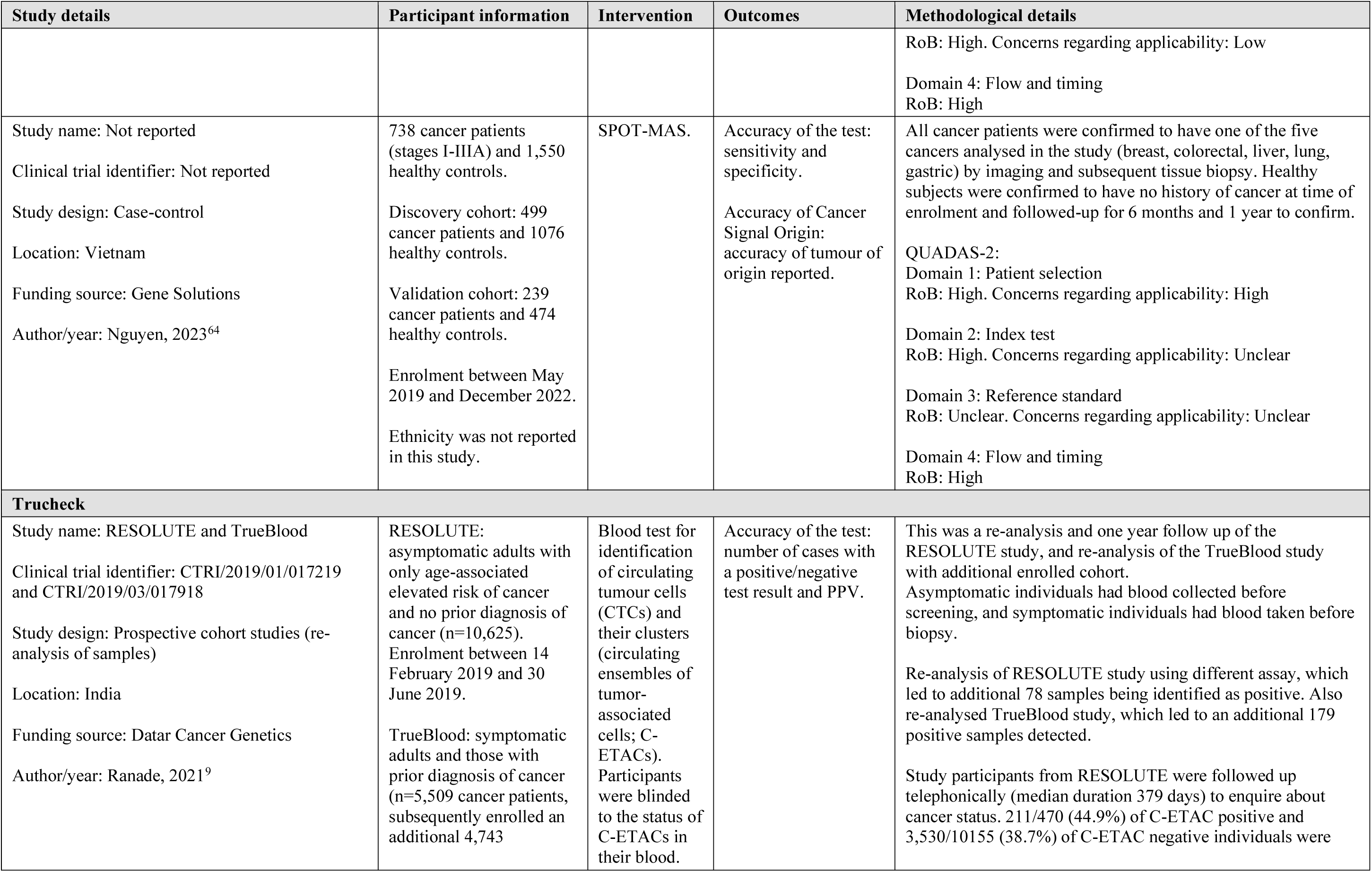
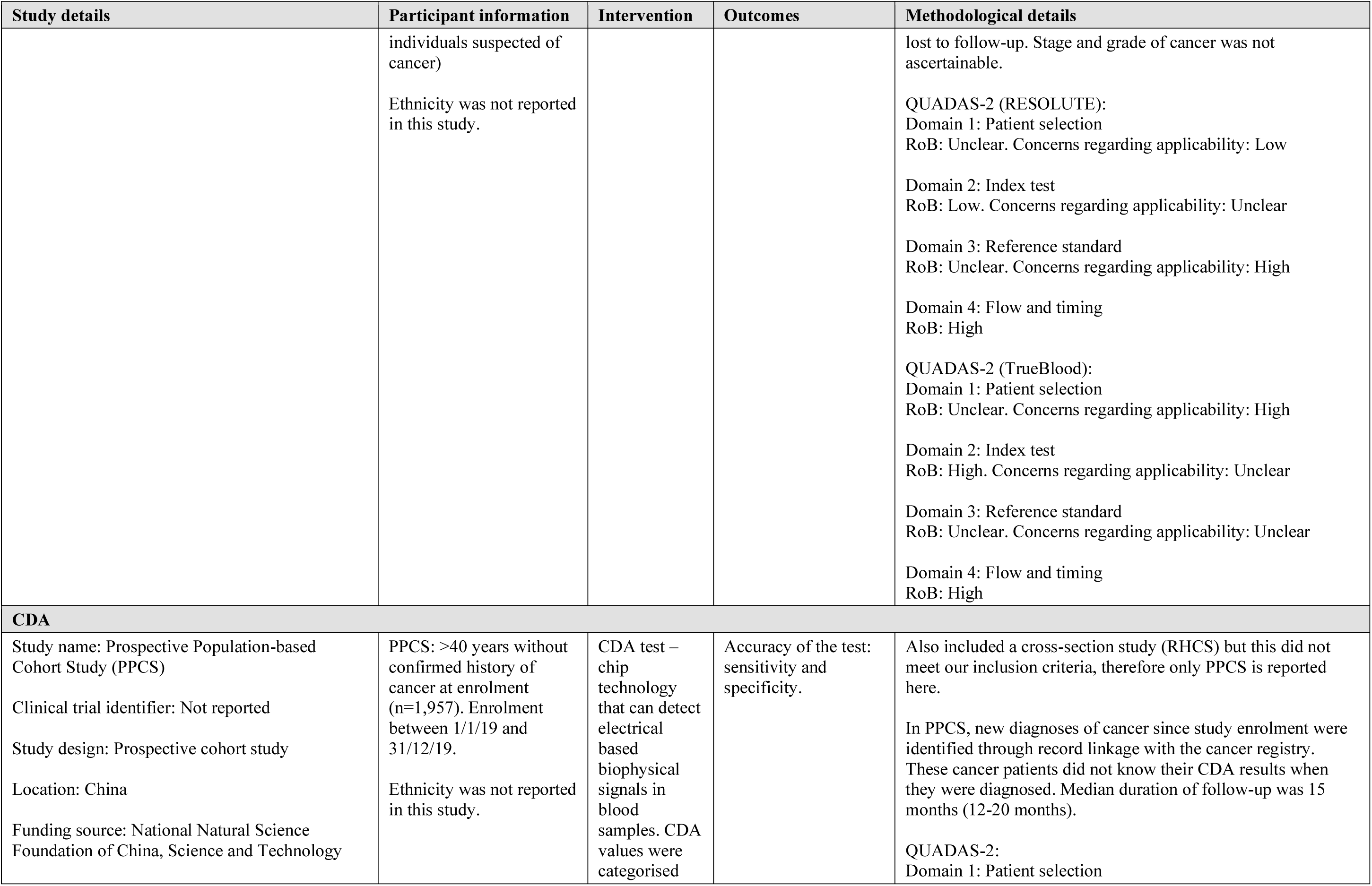
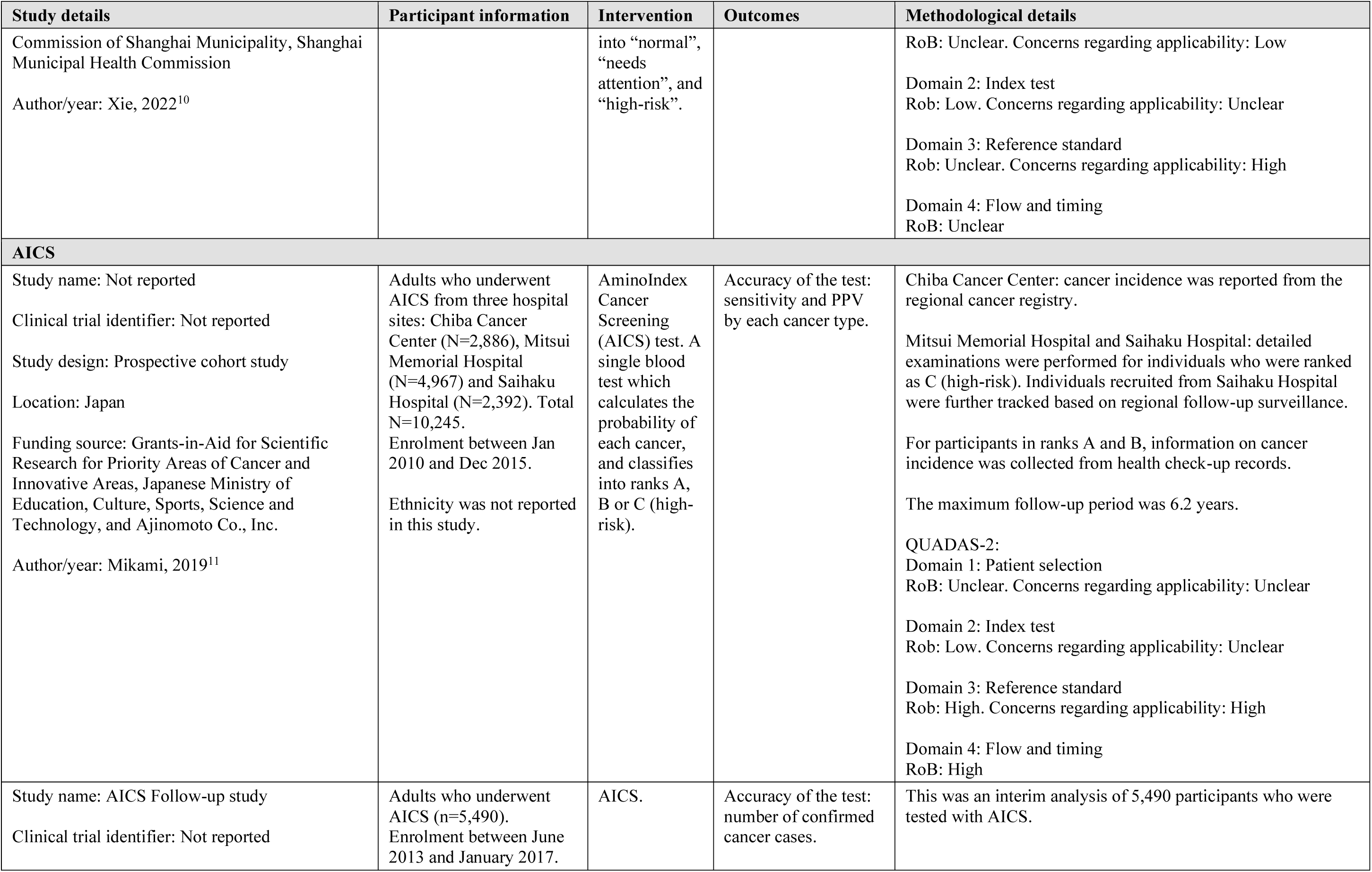
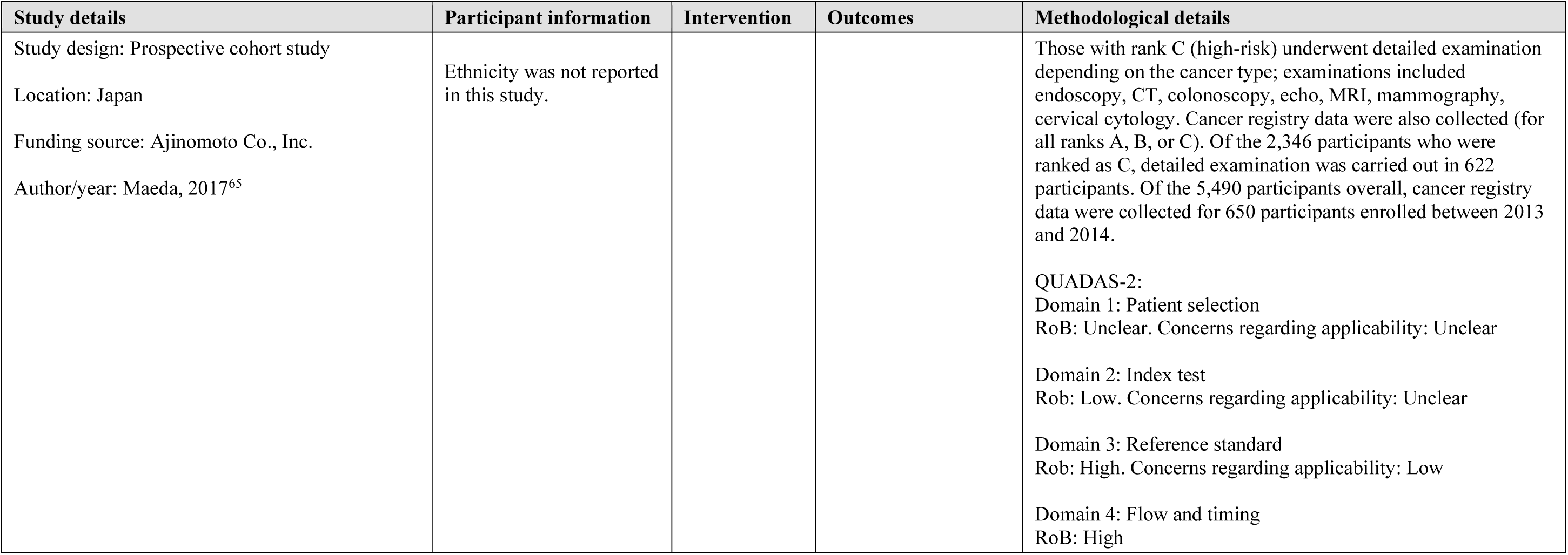
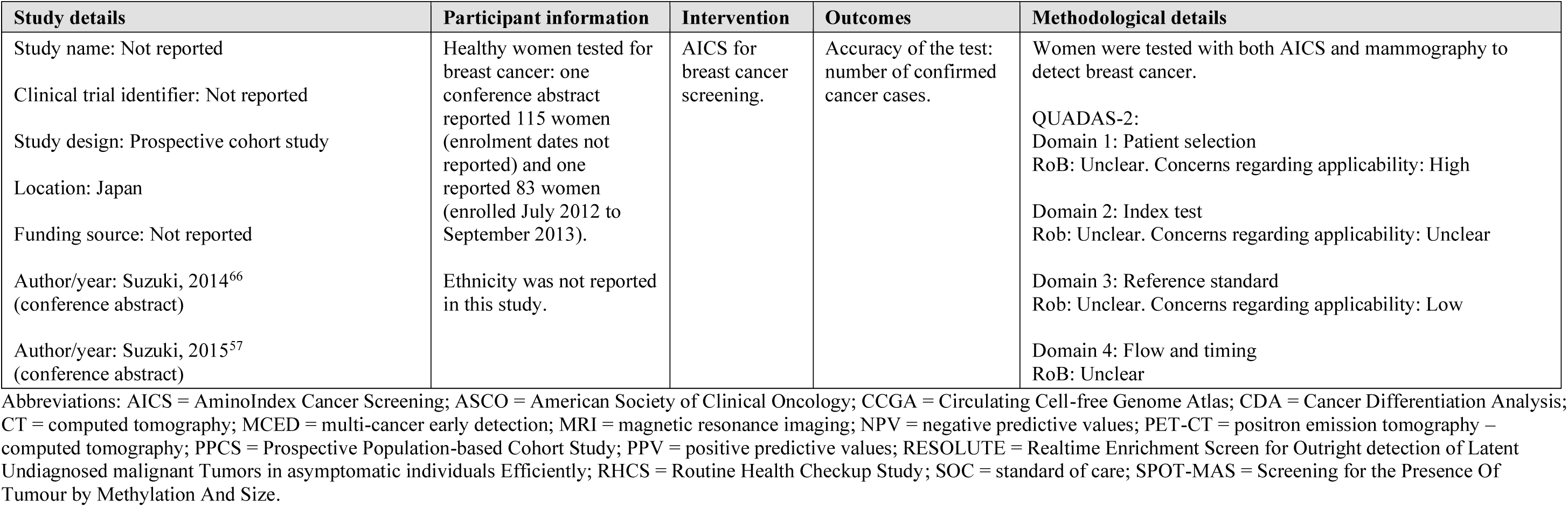
Characteristics of the included studies for each MCED test.

## APPENDIX 6. INCLUDED STUDIES OF MCED TECHNOLOGIES AT AN UNCLEAR STAGE OF DEVELOPMENT

**Table 12.** Characteristics of the included studies of MCED technologies at an unclear stage of development.

| Study details | Participant information | Review outcomes assessed/results | QUADAS-2 overall result |
| --- | --- | --- | --- |
| **Aristotle - StageZero Life Sciences** |  |  |  |
| Case-control (Dempsey, 2020),<sup>67</sup> United States | Cancer arm (n=1013)<br>Non-cancer arm (n=1832; including 1042 healthy controls and 790 with other health conditions)<br>Ethnicity was not reported in this study. | Accuracy of the test (sensitivity, PPV and NPV across 11 types of cancer):<br>Sensitivity: cervical, nasopharynx and stomach cancer reported highest sensitivity at 100%, lowest reported for colon at 55.6%; PPV ranged from 5.6% for liver to 77.7% for breast; NPV ranged from 96.7% for colon polyps to 100% for bladder, cervical, endometrial, liver, nasopharynx, ovarian and stomach. | Risk of bias: High<br>Applicability concerns: High |
| **CancerenD24 – manufacturer unknown** |  |  |  |
| Case-control (Arber, 2017),<sup>68</sup> Israel | Not reported<br>Ethnicity was not reported in this study (cancer patients and healthy controls were matched on ethnicity). | Accuracy of the test (across 7 types of cancer):<br>For colorectal cancer: sensitivity: 79.2%, specificity: 74.7%, PPV: 38%, NPV: 94.8%<br>For pancreatic cancer: sensitivity: 70%, specificity: 75.9%, PPV: 17.1%, NPV: 97.3%<br>Other outcomes reported: Sensitivity and specificity for haematological malignancies were also statistically significant (but not reported). The test was unable to discriminate patients with cervical, stomach and lung cancer and healthy subjects. | Risk of bias: High<br>Applicability concerns: High |
| Case-control (Massarwi, 2019),<sup>69</sup> Israel | Not reported<br>Ethnicity was not reported in this study. | Accuracy of the test: sensitivity (across 5 types of cancer): 94% for all cancers (bladder: 100%, pancreas: 89%, colorectal: 100%, colon adenoma: 87%, stomach: 100%); specificity: 84% (healthy subjects), 74% (healthy subjects with family history), 95% (healthy subjects without family history); NPV also reported (ranged from 93% to 100% depending on cancer type and whether there is family history). | Risk of bias: High<br>Applicability concerns: High |
| Case-control (Shapira, 2020),<sup>70</sup> Israel | Cancer arm (n=222)<br>Non-cancer arm (n=745)<br>Ethnicity was not reported in this study. | Accuracy of the test: specificity (across 17 types of cancer): 68.6%, sensitivity and NPV reported for each type of cancer (sensitivity ranged from 38.0% [bladder] to 100% [oesophageal, Squamous cell carcinoma, and stomach]). | Risk of bias: High<br>Applicability concerns: High |
| Case-control (Shapira, 2021),<sup>71</sup> Israel | Cancer arm (n=552)<br>Non-cancer arm (n=724)<br>Ethnicity was not reported in this study. | Accuracy of the test: sensitivity (across 8 cancer types): 84% (haematological), 80% (lung), 73% (breast), 71% (head and neck and GI cancers) | Risk of bias: High<br>Applicability concerns: High |
| Case-control (Madah, 2023),<sup>72</sup> Israel | Cancer arm (n=464)<br>Non-cancer arm (n=1138; matched on age, gender and medical history)<br>Ethnicity was not reported in this study. | Accuracy of the test (across 21 major cancer types): sensitivity: 87%, specificity: 87% | Risk of bias: High<br>Applicability concerns: High |
| **OncoSeek – SeekIn Inc** |  |  |  |
| Case-control (Luan, 2023),<sup>73</sup> China | Two independent validation cohorts:<br>Cohort 1:<br>Cancer arm (n=363)<br>Non-cancer arm (n=5556)<br>Nov 2012 to May 2022<br>Cohort 2 (same data as Cohen 2018, CancerSEEK):<br>Cancer arm (n=1005)<br>Non-cancer arm (n=812)<br>Ethnicity was not reported in cohort 1, and cohort 2 examined the same participants as Cohen 2018. | Accuracy of the test (supplements table S3):<br>In cohort 1: sensitivity: 47.4% (42.1%-52.7%), specificity: 90.0% (89.2%-90.8%), PPV: 23.7% (20.6%-26.9%), NPV: 96.3% (95.8%-96.8%)<br>In cohort 2: sensitivity: 49.3% (46.1%-52.4%), specificity: 90.1% (87.9%-92.1%), PPV: 86.1% (83.0%-88.8%), NPV: 58.9% (56.1%-61.7%) | Risk of bias: High<br>Applicability concerns: High |
| Case-control (Mao, 2023),<sup>74</sup> China | Cancer arm (n=1959)<br>Non-cancer arm: (n=7423)<br>Divided into one training and two independent validation cohorts<br>Ethnicity was not reported in this study. | Note: results were not provided separately for training and validation cohorts, only that it was consistent between them.<br>Accuracy of the test (across 9 common cancer types): sensitivity: 51.7% (49.4%-53.9%), sensitivity for pancreatic cancer: 77.6% (69.3%-84.6%). Sensitivity ranged from 37.1% to 77.6% across breast, colorectal, liver, lung, lymphoma, oesophagus, ovary, pancreas and stomach cancers.<br>Specificity: 92.9% (95% CI 92.3 to 93.5)<br>Accuracy of CSO: 66.8% (within true positives) | Risk of bias: High<br>Applicability concerns: High |
| **SeekInCare – SeekIn Inc** |  |  |  |
| Case-control and prospective cohort study (Mao 2023),<sup>75</sup> China | Case-control:<br>Cancer arm (n=615; stage I-IV, 8 common cancers, 19 uncommon cancers)<br>Non-cancer arm (n=898)<br>Real-world cohort: 1212 subjects (median follow-up time: 753 days [range 78-1669 days])<br>Ethnicity was not reported in this study. | Accuracy of the test:<br>Case-control: sensitivity: 69.4% (stage I: 50.3%, stage II: 64%, stage III: 73.8%, stage IV: 86.2%), specificity: 98.0%, sensitivity by each type of cancer (breast: 45.1%, stomach: 50.0%, lung: 63.4%, colorectum: 69.4%, lymphoma: 70.5%, liver: 81.4%, pancreas: 82.4%, leukemia: 90.9%)<br>Real-world cohort: sensitivity: 72.2%, specificity: 96.1%, PPV: 22.0%, NPV: 99.6% | Case-control:<br>Risk of bias: High<br>Applicability concerns: High<br>Cohort study:<br>Risk of bias: High<br>Applicability concerns: Unclear |
| SeekIn Inc news article: case-control and prospective cohort study (SeekIn Inc, 2022),<sup>76</sup> location not reported | Case-control:<br>Cancer arm (n=616; stage I-IV 8 common cancers, 19 other types)<br>Non-cancer arm (n=898)<br>Real-world cohort: 604 subjects (median follow-up time: 404 days)<br>Ethnicity was not reported in this study. | Accuracy of the test:<br>Case-control: sensitivity: 68.0% (stage I: 49.0%, stage II: 61.3%, stage III: 72.5%, stage IV: 85.4%), specificity: 98.0%<br>Real-world cohort (detected 12 cancer cases): sensitivity: 92.3%, specificity: 97.7%, PPV: 57.1%, NPV: 99.7% | Case-control:<br>Risk of bias: High<br>Applicability concerns: High<br>Cohort study:<br>Risk of bias: Unclear<br>Applicability concerns: Unclear |
| **OverC – Burning Rock Biotech** |  |  |  |
| THUNDER (Gao, 2023)<sup>77</sup><br>Case-control, China | Independent validation sample (age-matched):<br>Cancer arm (n=473)<br>Non-cancer arm (n=473). Apr 2021 to Nov 2021<br>Ethnicity was not reported in this study. | Accuracy of the test:<br>sensitivity: 69.1% (64.8%-73.3%), specificity: 98.9% (97.6%-99.7%); sensitivity by stage: stage I 35.4% (26.6%-45.0%), stage II 54.5% (43.6%-65.2%), stage III 82.4% (75.1%-88.3%), stage IV 93.8% (88.2%-97.3%).<br>Accuracy of CSO: first CSO correct: 83.2% (78.7%-87.1%), first or second CSO correct: 91.7% (88.2%-94.5%)<br>Subgroup analysis by age and sex. | Risk of bias: High<br>Applicability concerns: High |
| THUNDER-II (Gao, 2021)<sup>78</sup><br>Case-control, China | Independent validation sample:<br>Cancer arm (n=202)<br>Non-cancer arm (n=158; including 76 healthy controls and 82 high-risk individuals)<br>Ethnicity was not reported in this study. | Accuracy of the test: sensitivity: 74.8% (68.1%-80.5%), specificity: 98.1% (94.1%-99.5%); sensitivity by stage: Stage I 53.0% (40.4%-76.3%), stage II 73.3% (57.8%-84.9%), stage III 90.4% (78.2%-96.4%), stage IV 92.3% (78.0%-98.0%).<br>Accuracy of CSO: 80.8% (73.4%-86.6%) | Risk of bias: High<br>Applicability concerns: High |
| **Carcimun-test – Carcimun Biotech** |  |  |  |
| Case-control (Salat, 2022),<sup>81</sup> Austria | Cancer patients (across 17 cancer types, undergoing surgery) and healthy controls<br>Cancer arm (n=170);<br>Non-cancer arm (n=137)<br>Ethnicity was not reported in this study. | Accuracy of the test: accuracy: 90.0%, sensitivity: 88.8%, specificity: 91.2%, PPV: 92.0%, NPV: 87.0%<br>Mortality: 5-year all-cause mortality was similar among cancer patients who were true positives and false negatives, suggesting the test had missed clinically relevant cancers.<br>Potential harms: no adverse effects observed for the blood withdrawal. | Risk of bias: High<br>Applicability concerns: High |
| **SpecGastro test – manufacturer unknown** |  |  |  |
| Case-control (Ma, 2022),<sup>82</sup> China | Gastrointestinal cancer patients (n=282; 98 colorectal cancer, 136 gastric cancer, 48 oesophageal cancer) and 195 controls<br>Ethnicity was not reported in this study. | Accuracy of the test: sensitivity: 76.6% (71.1%-81.3%), specificity: 89.2% (83.8%-93.1%). Sensitivity by each cancer type: colorectal (87.8% [79.2%-93.2%]), gastric (69.9% [61.3%-77.3%]), oesophageal (72.9% [57.9%-84.3%]) | Risk of bias: High<br>Applicability concerns: High |

Abbreviations: CSO = Cancer Signal Origin; GI = gastrointestinal; NPV = negative predictive values; PPV = positive predictive values; THUNDER = The Unintrusive Detection of Early-stage cancers.

**Table 13.**
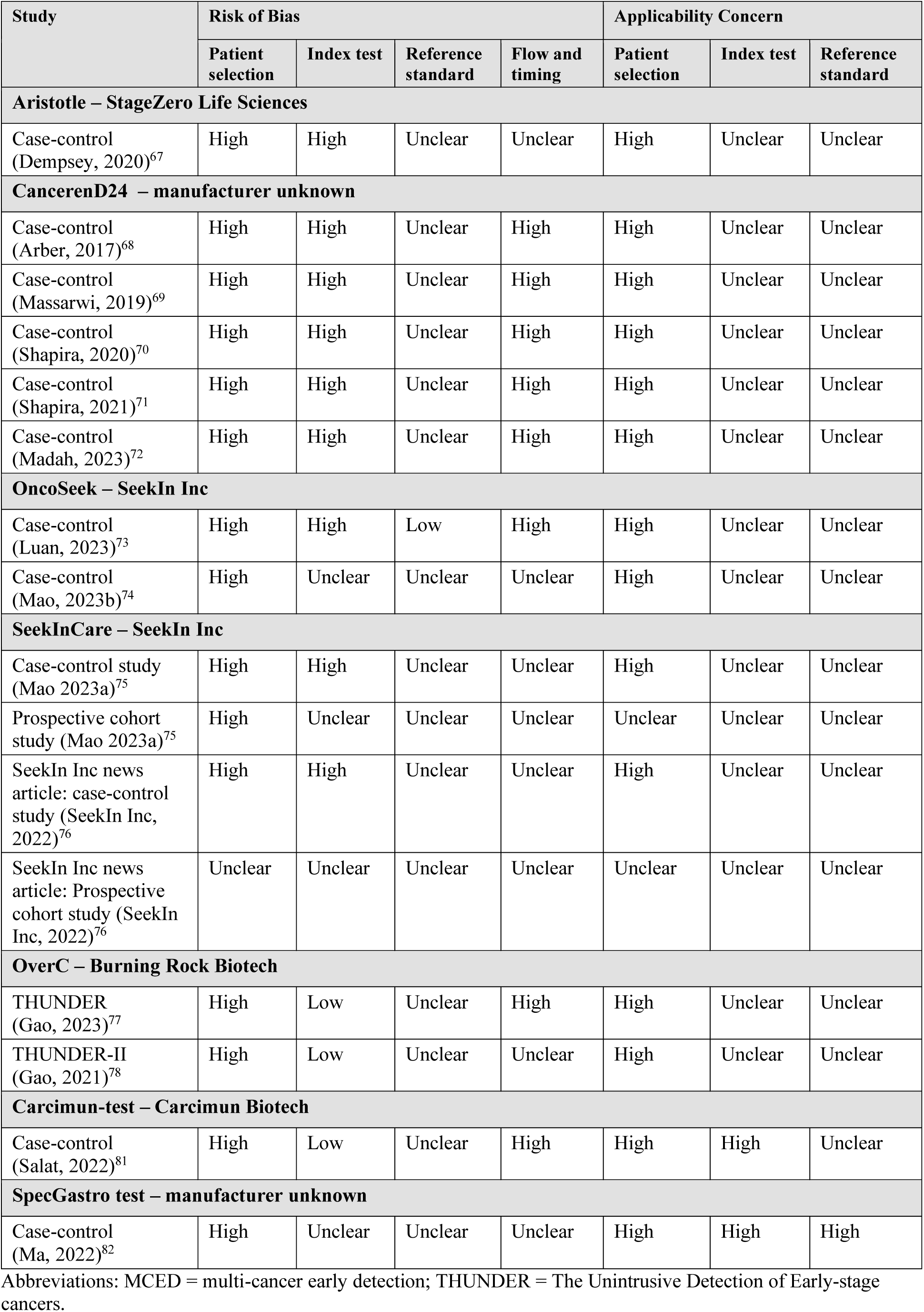
QUADAS-2 assessment results for the included studies of MCED technologies at an unclear stage of development.

## APPENDIX 7. CANCER TYPES DETECTED BY INCLUDED TESTS

**Table 14.**
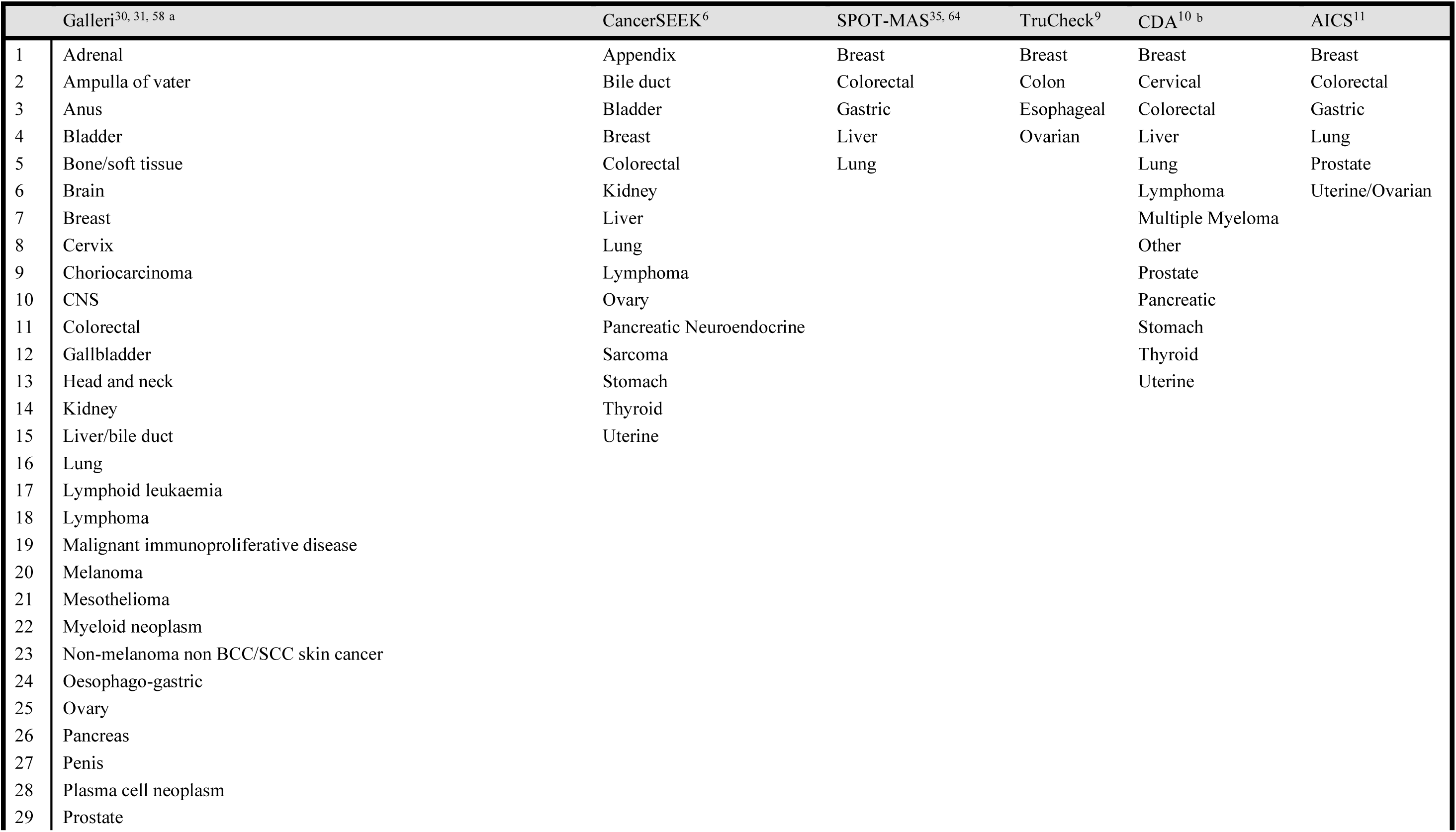
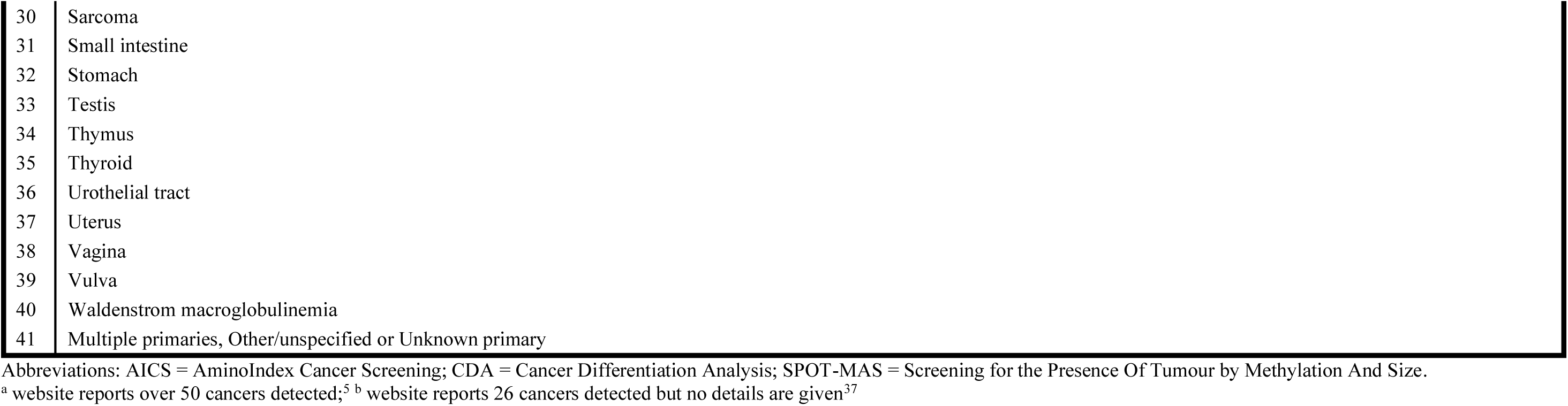
List of cancer types detected by included tests.

|  | Galleri<sup>30, 31, 58 a</sup> | CancerSEEK<sup>6</sup> | SPOT-MAS<sup>35, 64</sup> | TruCheck<sup>9</sup> | CDA<sup>10 b</sup> | AICS<sup>11</sup> |
| --- | --- | --- | --- | --- | --- | --- |
| 1 | Adrenal | Appendix | Breast | Breast | Breast | Breast |
| 2 | Ampulla of Vater | Bile duct | Colorectal | Colon | Cervical | Colorectal |
| 3 | Anus | Bladder | Gastric | Esophageal | Colorectal | Gastric |
| 4 | Bladder | Breast | Liver | Ovarian | Liver | Lung |
| 5 | Bone/soft tissue | Colorectal | Lung |  | Lung | Prostate |
| 6 | Brain | Kidney |  |  | Lymphoma | Uterine/Ovarian |
| 7 | Breast | Liver |  |  | Multiple Myeloma |  |
| 8 | Cervix | Lung |  |  | Other |  |
| 9 | Choriocarcinoma | Lymphoma |  |  | Prostate |  |
| 10 | CNS | Ovary |  |  | Pancreatic |  |
| 11 | Colorectal | Pancreatic Neuroendocrine |  |  | Stomach |  |
| 12 | Gallbladder | Sarcoma |  |  | Thyroid |  |
| 13 | Head and neck | Stomach |  |  | Uterine |  |
| 14 | Kidney | Thyroid |  |  |  |  |
| 15 | Liver/bile duct | Uterine |  |  |  |  |
| 16 | Lung |  |  |  |  |  |
| 17 | Lymphoid leukaemia |  |  |  |  |  |
| 18 | Lymphoma |  |  |  |  |  |
| 19 | Malignant immunoproliferative disease |  |  |  |  |  |
| 20 | Melanoma |  |  |  |  |  |
| 21 | Mesothelioma |  |  |  |  |  |
| 22 | Myeloid neoplasm |  |  |  |  |  |
| 23 | Non-melanoma non BCC/SCC skin cancer |  |  |  |  |  |
| 24 | Oesophago-gastric |  |  |  |  |  |
| 25 | Ovary |  |  |  |  |  |
| 26 | Pancreas |  |  |  |  |  |
| 27 | Penis |  |  |  |  |  |
| 28 | Plasma cell neoplasm |  |  |  |  |  |
| 29 | Prostate |  |  |  |  |  |

|  |  |
| --- | --- |
| 30 | Sarcoma |
| 31 | Small intestine |
| 32 | Stomach |
| 33 | Testis |
| 34 | Thymus |
| 35 | Thyroid |
| 36 | Urothelial tract |
| 37 | Uterus |
| 38 | Vagina |
| 39 | Vulva |
| 40 | Waldenstrom macroglobulinemia |
| 41 | Multiple primaries, Other/unspecified or Unknown primary |

Abbreviations: AICS = AminoIndex Cancer Screening; CDA = Cancer Differentiation Analysis; SPOT-MAS = Screening for the Presence Of Tumour by Methylation And Size.
<sup>a</sup> website reports over 50 cancers detected;<sup>5</sup> <sup>b</sup> website reports 26 cancers detected but no details are given<sup>37</sup>

**Table 15.**
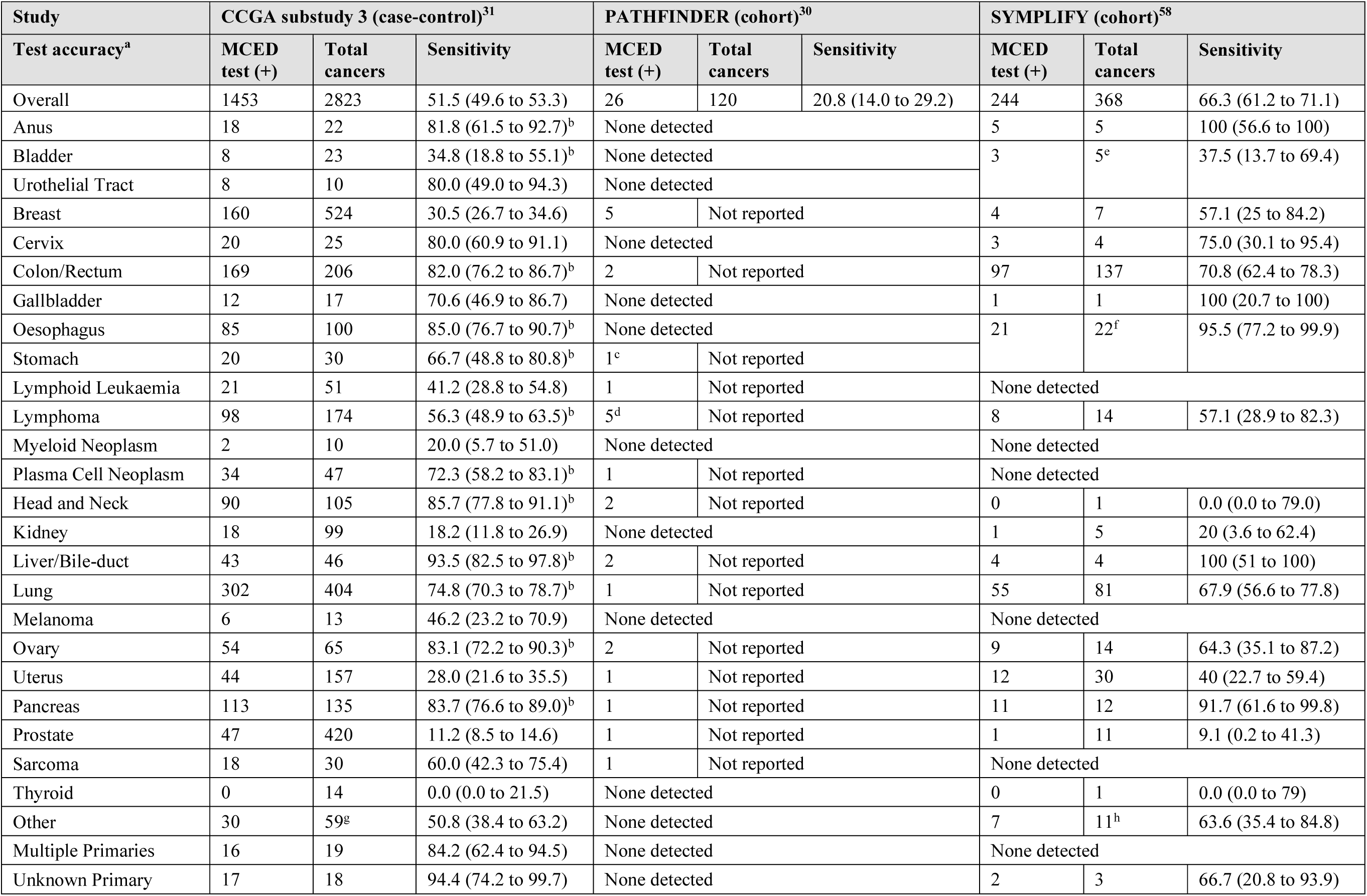
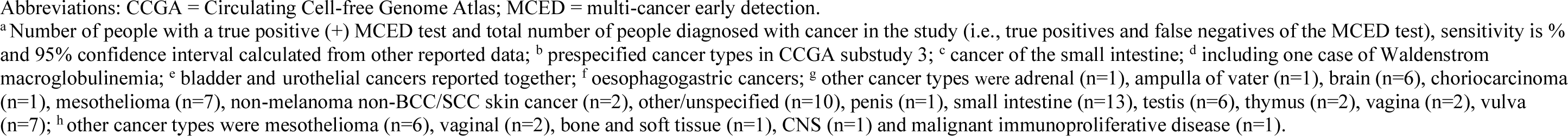
Number and proportion of cancers detected by the GRAIL MCED test (Galleri) for different cancer types.

| Study | CCGA substudy 3 (case-control)<sup>31</sup> |  |  | PATHFINDER (cohort)<sup>30</sup> |  |  | SYMPHIFY (cohort)<sup>58</sup> |  |  |
| --- | --- | --- | --- | --- | --- | --- | --- | --- | --- |
| Test accuracy<sup>a</sup> | MCED test (+) | Total cancers | Sensitivity | MCED test (+) | Total cancers | Sensitivity | MCED test (+) | Total cancers | Sensitivity |
| Overall | 1453 | 2823 | 51.5 (49.6 to 53.3) | 26 | 120 | 20.8 (14.0 to 29.2) | 244 | 368 | 66.3 (61.2 to 71.1) |
| Anus | 18 | 22 | 81.8 (61.5 to 92.7)<sup>b</sup> | None detected |  |  | 5 | 5 | 100 (56.6 to 100) |
| Bladder | 8 | 23 | 34.8 (18.8 to 55.1)<sup>b</sup> | None detected |  |  | 3 | 5<sup>e</sup> | 37.5 (13.7 to 69.4) |
| Urothelial Tract | 8 | 10 | 80.0 (49.0 to 94.3) | None detected |  |  |  |  |  |
| Breast | 160 | 524 | 30.5 (26.7 to 34.6) | 5 | Not reported |  | 4 | 7 | 57.1 (25 to 84.2) |
| Cervix | 20 | 25 | 80.0 (60.9 to 91.1) | None detected |  |  | 3 | 4 | 75.0 (30.1 to 95.4) |
| Colon/Rectum | 169 | 206 | 82.0 (76.2 to 86.7)<sup>b</sup> | 2 | Not reported |  | 97 | 137 | 70.8 (62.4 to 78.3) |
| Gallbladder | 12 | 17 | 70.6 (46.9 to 86.7) | None detected |  |  | 1 | 1 | 100 (20.7 to 100) |
| Oesophagus | 85 | 100 | 85.0 (76.7 to 90.7)<sup>b</sup> | None detected |  |  | 21 | 22<sup>f</sup> | 95.5 (77.2 to 99.9) |
| Stomach | 20 | 30 | 66.7 (48.8 to 80.8)<sup>b</sup> | 1<sup>c</sup> | Not reported |  |  |  |  |
| Lymphoid Leukaemia | 21 | 51 | 41.2 (28.8 to 54.8) | 1 | Not reported |  | None detected |  |  |
| Lymphoma | 98 | 174 | 56.3 (48.9 to 63.5)<sup>b</sup> | 5<sup>d</sup> | Not reported |  | 8 | 14 | 57.1 (28.9 to 82.3) |
| Myeloid Neoplasm | 2 | 10 | 20.0 (5.7 to 51.0) | None detected |  |  | None detected |  |  |
| Plasma Cell Neoplasm | 34 | 47 | 72.3 (58.2 to 83.1)<sup>b</sup> | 1 | Not reported |  | None detected |  |  |
| Head and Neck | 90 | 105 | 85.7 (77.8 to 91.1)<sup>b</sup> | 2 | Not reported |  | 0 | 1 | 0.0 (0.0 to 79.0) |
| Kidney | 18 | 99 | 18.2 (11.8 to 26.9) | None detected |  |  | 1 | 5 | 20 (3.6 to 62.4) |
| Liver/Bile-duct | 43 | 46 | 93.5 (82.5 to 97.8)<sup>b</sup> | 2 | Not reported |  | 4 | 4 | 100 (51 to 100) |
| Lung | 302 | 404 | 74.8 (70.3 to 78.7)<sup>b</sup> | 1 | Not reported |  | 55 | 81 | 67.9 (56.6 to 77.8) |
| Melanoma | 6 | 13 | 46.2 (23.2 to 70.9) | None detected |  |  | None detected |  |  |
| Ovary | 54 | 65 | 83.1 (72.2 to 90.3)<sup>b</sup> | 2 | Not reported |  | 9 | 14 | 64.3 (35.1 to 87.2) |
| Uterus | 44 | 157 | 28.0 (21.6 to 35.5) | 1 | Not reported |  | 12 | 30 | 40 (22.7 to 59.4) |
| Pancreas | 113 | 135 | 83.7 (76.6 to 89.0)<sup>b</sup> | 1 | Not reported |  | 11 | 12 | 91.7 (61.6 to 99.8) |
| Prostate | 47 | 420 | 11.2 (8.5 to 14.6) | 1 | Not reported |  | 1 | 11 | 9.1 (0.2 to 41.3) |
| Sarcoma | 18 | 30 | 60.0 (42.3 to 75.4) | 1 | Not reported |  | None detected |  |  |
| Thyroid | 0 | 14 | 0.0 (0.0 to 21.5) | None detected |  |  | 0 | 1 | 0.0 (0.0 to 79) |
| Other | 30 | 59<sup>g</sup> | 50.8 (38.4 to 63.2) | None detected |  |  | 7 | 11<sup>h</sup> | 63.6 (35.4 to 84.8) |
| Multiple Primaries | 16 | 19 | 84.2 (62.4 to 94.5) | None detected |  |  | None detected |  |  |
| Unknown Primary | 17 | 18 | 94.4 (74.2 to 99.7) | None detected |  |  | 2 | 3 | 66.7 (20.8 to 93.9) |

Abbreviations: CCGA = Circulating Cell-free Genome Atlas; MCED = multi-cancer early detection.
<sup>a</sup>Number of people with a true positive (+) MCED test and total number of people diagnosed with cancer in the study (i.e., true positives and false negatives of the MCED test), sensitivity is % and 95% confidence interval calculated from other reported data; <sup>b</sup> prespecified cancer types in CCGA substudy 3; <sup>c</sup> cancer of the small intestine; <sup>d</sup> including one case of Waldenstrom macroglobulinemia; <sup>e</sup> bladder and urothelial cancers reported together; <sup>f</sup> oesophagogastric cancers; <sup>g</sup> other cancer types were adrenal (n=1), ampulla of vater (n=1), brain (n=6), choriocarcinoma (n=1), mesothelioma (n=7), non-melanoma non-BCC/SCC skin cancer (n=2), other/unspecified (n=10), penis (n=1), small intestine (n=13), testis (n=6), thymus (n=2), vagina (n=2), vulva (n=7); <sup>h</sup> other cancer types were mesothelioma (n=6), vaginal (n=2), bone and soft tissue (n=1), CNS (n=1) and malignant immunoproliferative disease (n=1).

**Table 16.** Number and proportion of cancers detected by the CancerSEEK test.

| Study | Cohen 2018 (case-control)<sup>33</sup> |  |  | DETECT-A (cohort)<sup>6</sup> |  |  |
| --- | --- | --- | --- | --- | --- | --- |
| Test accuracy<sup>a</sup> | MCED test (+) | Total cancers | Sensitivity | MCED test (+) | Total cancers | Sensitivity |
| Overall | 626 | 1005 | 62.3 (59.3 to 65.3) | 26 | 70 | 27.1 (18.5 to 37.1) |
| Bladder | None detected |  |  | 0 | 1 | 0.0 (0.0 to 79) |
| Breast | 70 | 209 | 33.5 (27.4 to 40.1) | 1 | 27 | 3.7 (0.7 to 18.3) |
| Colon/Rectum | 252 | 388 | 64.9 (60.1 to 69.5) | 2 | 3 | 66.7 (20.8 to 93.9) |
| Oesophagus | 31 | 45 | 68.9 (54.3 to 80.5) | None detected |  |  |
| Stomach | 49 | 68 | 72.1 (60.4 to 81.3) | 0 | 3 | 0.0 (0.0 to 56) |
| Lymphoma | None detected |  |  | 2 | 4 | 50.0 (15 to 85) |
| Kidney | None detected |  |  | 1 | 2 | 50.0 (9.5 to 90.5) |
| Liver/Bile-duct | 43 | 44 | 97.7 (88.2 to 99.6) | 0 | 2 | 0.0 (0.0 to 65.8) |
| Lung | 61 | 104 | 58.7 (49 to 67.6) | 9 | 21 | 42.9 (24.5 to 63.5) |
| Ovary | 53 | 54 | 98.1 (90.2 to 99.7) | 6 | 7 | 85.7 (48.7 to 97.4) |
| Uterus | None detected |  |  | 2 | 15 | 13 (3.7 to 37.9) |
| Pancreas | 67 | 93 | 72.0 (62.2 to 80.1) | 0 | 2 | 0.0 (0.0 to 65.8) |
| Sarcoma | None detected |  |  | 0 | 2 | 0.0 (0.0 to 65.8) |
| Thyroid | None detected |  |  | 1 | 5 | 20 (3.6 to 62.4) |
| Other | None detected |  |  | 2 | 2<sup>b</sup> | 100 (34.2 to 100) |

Abbreviations: MCED = multi-cancer early detection.
<sup>a</sup>Number of people with a true positive (+) MCED test and total number of people diagnosed with cancer in the study (i.e., true positives and false negatives of the MCED test), sensitivity is % and 95% confidence interval calculated from other reported data; <sup>b</sup> other cancer types were appendix (n=1) and unknown carcinoma (n=1).

**Table 17.** Number and proportion of cancers detected by the SPOT-MAS test.

| Study | Nyugen 2023 (case-control)<sup>64</sup> |  |  | K-DETEK (cohort)<sup>8</sup> |  |  |
| --- | --- | --- | --- | --- | --- | --- |
| Test accuracy<sup>a</sup> | MCED test (+) | Total cancers | Sensitivity | MCED test (+) | Total cancers | Sensitivity |
| Overall | 173 | 239 | 72.4 (66.3 to 78.0) | 6 | 6 | 100 (54.1 to 100) |
| Breast | 33 | 67 | 49.3 (37.7 to 60.9) | 1 | Not reported |  |
| Colon/Rectum | 44 | 53 | 83.0 (70.8 to 90.8) | None detected |  |  |
| Gastric | 19 | 31 | 61.3 (43.8 to 76.3) | 1 | Not reported |  |
| Liver/Bile-duct | 83 | 91 | 91.2 (83.6 to 95.5) | 3 | Not reported |  |
| Lung | 36 | 43 | 83.7 (70.0 to 91.9) | None detected |  |  |
| Other | None detected |  |  | 1<sup>b</sup> | Not reported |  |

Abbreviations: MCED = multi-cancer early detection.
<sup>a</sup>Number of people with a true positive (+) MCED test and total number of people diagnosed with cancer in the study (i.e., true positives and false negatives of the MCED test), sensitivity is % and 95% confidence interval calculated from other reported data; <sup>b</sup> other cancer type was endometrial (n=1).

**Table 18.**
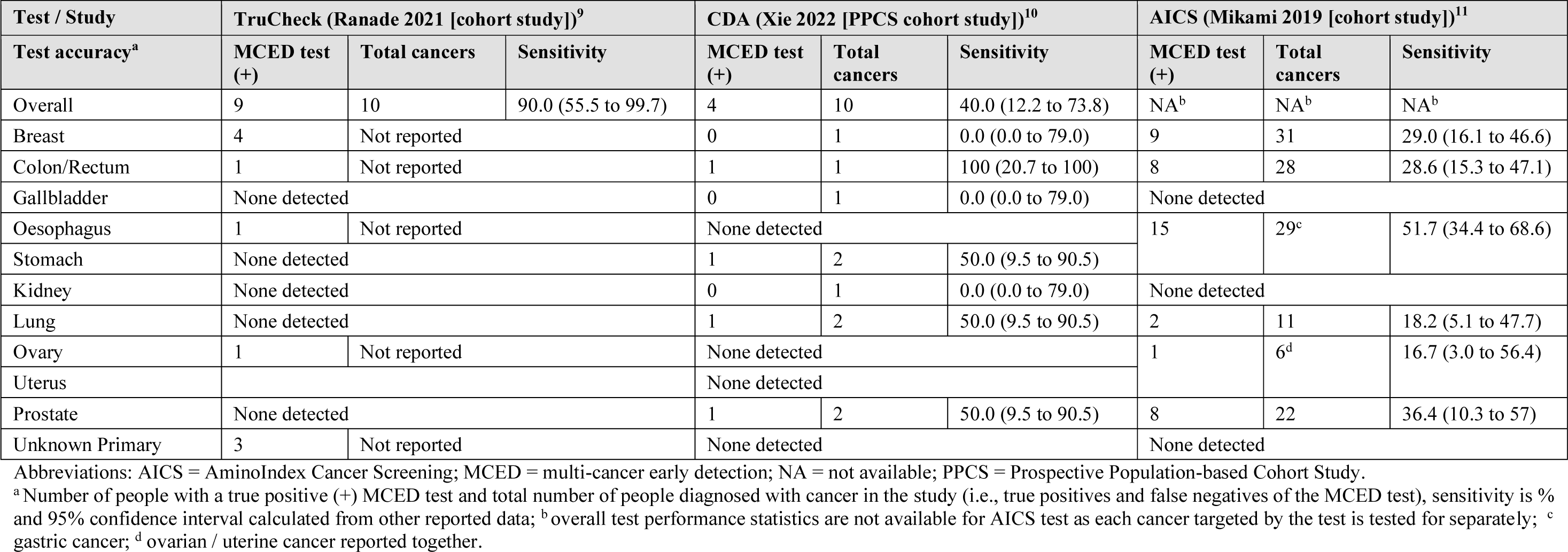
Number and proportion of cancers detected by the TruCheck, CDA and AICS tests.

| Test / Study | TruCheck (Ranade 2021 [cohort study])<sup>9</sup> |  |  | CDA (Xie 2022 [PPCS cohort study])<sup>10</sup> |  |  | AICS (Mikami 2019 [cohort study])<sup>11</sup> |  |  |
| --- | --- | --- | --- | --- | --- | --- | --- | --- | --- |
| Test accuracy<sup>a</sup> | MCED test (+) | Total cancers | Sensitivity | MCED test (+) | Total cancers | Sensitivity | MCED test (+) | Total cancers | Sensitivity |
| Overall | 9 | 10 | 90.0 (55.5 to 99.7) | 4 | 10 | 40.0 (12.2 to 73.8) | NA<sup>b</sup> | NA<sup>b</sup> | NA<sup>b</sup> |
| Breast | 4 | Not reported |  | 0 | 1 | 0.0 (0.0 to 79.0) | 9 | 31 | 29.0 (16.1 to 46.6) |
| Colon/Rectum | 1 | Not reported |  | 1 | 1 | 100 (20.7 to 100) | 8 | 28 | 28.6 (15.3 to 47.1) |
| Gallbladder | None detected |  |  | 0 | 1 | 0.0 (0.0 to 79.0) | None detected |  |  |
| Oesophagus | 1 | Not reported |  | None detected |  |  | 15 | 29<sup>c</sup> | 51.7 (34.4 to 68.6) |
| Stomach | None detected |  |  | 1 | 2 | 50.0 (9.5 to 90.5) |  |  |  |
| Kidney | None detected |  |  | 0 | 1 | 0.0 (0.0 to 79.0) | None detected |  |  |
| Lung | None detected |  |  | 1 | 2 | 50.0 (9.5 to 90.5) | 2 | 11 | 18.2 (5.1 to 47.7) |
| Ovary | 1 | Not reported |  | None detected |  |  | 1 | 6<sup>d</sup> | 16.7 (3.0 to 56.4) |
| Uterus |  |  |  | None detected |  |  |  |  |  |
| Prostate | None detected |  |  | 1 | 2 | 50.0 (9.5 to 90.5) | 8 | 22 | 36.4 (10.3 to 57) |
| Unknown Primary | 3 | Not reported |  | None detected |  |  | None detected |  |  |

Abbreviations: AICS = AminoIndex Cancer Screening; MCED = multi-cancer early detection; NA = not available; PPCS = Prospective Population-based Cohort Study.
<sup>a</sup>Number of people with a true positive (+) MCED test and total number of people diagnosed with cancer in the study (i.e., true positives and false negatives of the MCED test), sensitivity is % and 95% confidence interval calculated from other reported data; <sup>b</sup> overall test performance statistics are not available for AICS test as each cancer targeted by the test is tested for separately; <sup>c</sup> gastric cancer; <sup>d</sup> ovarian / uterine cancer reported together.

## APPENDIX 8. TEST PERFORMANCE OF GRAIL MCED TEST AND CANCERSEEK BY SUBGROUPS

**Table 19.** Test performance statistics of the refined MCED test (Galleri) in the PATHFINDER study by risk cohorts.

|  | All patients | ≥50 y With Additional Risk | ≥50 y Without Additional Risk |
| --- | --- | --- | --- |
| Number analysed<sup>a</sup> | 6369 | 3532 | 2837 |
| Total cancers (n) | 120 | 77 | 43 |
| TP (n) | 25 | 18 | 7 |
| FP (n) | 33 | 22 | 11 |
| FN (n) | 95 | 59 | 36 |
| TN (n) | 6216 | 3433 | 2783 |
| **Accuracy of the test, % (95% confidence interval)** |  |  |  |
| Sensitivity | 20.8 (14.0 to 29.2)<sup>b</sup> | 23.4 (14.5 to 34.4)<sup>b</sup> | 16.3 (6.8 to 30.7)<sup>b</sup> |
| Specificity | 99.5 (99.3 to 99.6) | 99.4 (99.0 to 99.6) | 99.6 (99.3 to 99.8) |
| PPV | 43.1 (31.2 to 55.9) | 45.0 (30.7 to 60.2) | 38.9 (20.3 to 61.4) |
| NPV | 98.5 (98.2 to 98.8) | 98.3 (97.8 to 98.7) | 98.7 (98.2 to 99.1) |
| First CSO correct | 84.0 (65.3 to 93.6) | 88.9 (67.2 to 96.9) | 71.4 (35.9 to 91.8) |
| First or second CSO correct | 88.0 (70.0 to 95.8) | 88.9 (67.2 to 96.9) | 85.7 (48.7 to 99.3) |

Abbreviations: CSO = cancer signal origin; FN = false negatives; FP = false positives; MCED = multi cancer early detection; NPV = negative predictive value; PPV = positive predictive value; TN = true negatives; TP = true positives.
<sup>a</sup> Complete analysis set, those who received the MCED test, with follow-up information and/or diagnostic resolution, <sup>b</sup> values calculated from other reported data.

**Table 20.** Test performance by age and ethnicity in the CCGA substudy 3 of GRAIL MCED test and Cohen 2018 study of CancerSEEK.

| Test performance %<br>(95% CI)<sup>a</sup> | CCGA substudy 3 (case-control)<sup>31</sup> |  |  |  |  | Cohen 2018 (case-control)<sup>33</sup> |  |  |  |  |
| --- | --- | --- | --- | --- | --- | --- | --- | --- | --- | --- |
|  | MCED tests (+) | Total cancers | Sensitivity | Specificity | First CSO accuracy | MCED tests (+) | Total cancers | Sensitivity | Specificity | First CSO accuracy |
| Overall | 1453 | 2823 | 51.5 (49.6 to 53.3) | 99.5 (99.0 to 99.8) | 88.7 (87.0 to 90.2) | 626 | 1005 | 62.3 (59.3 to 65.3) | 99.1 (98.5 to 99.8) | 67.7 (64.0 to 71.3) |
| < 50 years | 21 | 385 | 55.1 (50.1 to 60.0) | 99.8 (98.6 to 100.0) | 87.1 (81.9 to 91.0) | 85 | 152 | 55.9 (48.0 to 63.6) | 99.7 (98.3 to 99.9) | 63.5 (52.9 to 73.0) |
| ≥ 50 years | 1241 | 2438 | 50.9 (48.9 to 52.9) | 99.4 (98.6 to 99.7) | 89.0 (87.1 to 90.6) | 541 | 853 | 63.4 (60.1 to 66.6) | 98.7 (97.3 to 99.4) | 68.4 (64.4 to 72.2) |
| ≥ 65 years | 725 | 1331 | 54.5 (51.9 to 57.2) | 99.4 (97.9 to 99.8) | 88.5 (86.0 to 90.7) | 299 | 475 | 62.9 (58.5 to 67.2) | 98.6 (95.9 to 99.5) | 66.6 (61.0 to 71.7) |
| 50 – 79 years | Not reported |  |  |  |  | 494 | 775 | 63.7 (60.3 to 67.1) | 99.1 (97.7 to 99.7) | 66.8 (62.5 to 70.8) |
| White<sup>b</sup> | 1193 | 2316 | 50.5 (48.4 to 52.5) | 99.6 (99.0 to 99.8) | Not reported | 365 | 675 | 54.1 (50.3 to 57.8) | 98.5 (96.5 to 99.4) | 72.6 (67.8 to 76.9) |
| Hispanic (all races) | 121 | 192 | 63.0 (56.0 to 69.5) | 98.1 (93.2 to 99.5) | Not reported | 1 | 1 | 100 (20.7 to 100) | 100 (96.9 to 100) | 100 (20.7 to 100) |
| Black<sup>b</sup> | 104 | 193 | 53.9 (46.8 to 60.8) | 100 (95.7 to 100) | Not reported | 11 | 14 | 78.6 (52.4 to 92.4) | 98.7 (95.4 to 99.6) | 63.6 (35.4 to 84.8) |
| Unknown | 34 | 65 | 52.3 (40.4 to 64.0) | 100 (89.6 to 100) | Not reported | 7 | 14 | 50 (26.8 to 73.2) | 100 (98 to 100) | 71.4 (35.9 to 91.8) |
| Other | 25 | 57<sup>c</sup> | 43.9 (31.8 to 56.7) | 100 (89.6 to 100) | Not reported | 239 | 323<sup>d</sup> | 70.4 (68.9 to 78.5) | 100 (85.1 to 100) | 60.3 (53.9 to 66.2) |

Abbreviations: CCGA = Circulating Cell-free Genome Atlas; CI = confidence interval; CSO = cancer signal origin.
<sup>a</sup>Number of people with a true positive (+) MCED test and total number of people diagnosed with cancer in the study (i.e., true positives and false negatives of the MCED test), sensitivity, specificity and first CSO accuracy are % and 95% CI, calculated from other reported data in Klein 2021,<sup>31</sup> Tang 2023 (CCGA substudy)<sup>47</sup> and Cohen 2018 (Table S4 and S10)<sup>33</sup>. Some categories of ethnicity combined compared to those reported in the original study reports, to align subgroups across the two tests, <sup>b</sup> non-Hispanic; <sup>c</sup> includes American Indian or Alaska native, Asian, native Hawaiian or Pacific islander; <sup>d</sup> Asian.

## APPENDIX 9. GRIPP2 SHORT FORM TABLE

| Section and topic | Item | Reported on page No |
| --- | --- | --- |
| 1: Aim | Report the aim of PPI in the study | 24-25 – Stakeholder involvement |
| 2: Methods | Provide a clear description of the methods used for PPI in the study | 24-25 – Stakeholder involvement |
| 3: Study results | Outcomes - Report the results of PPI in the study, including both positive and negative outcomes | 55-59 – Stakeholder Engagement<br>60-61 – Patient and Public Involvement |
| 4: Discussion and conclusions | Outcomes - Comment on the extent to which PPI influenced the study overall. Describe positive and negative effects | 4 – Abstract<br>10 - Plain language summary<br>11-12, 15-16 - Scientific summary<br>24-25 – Stakeholder involvement<br>55-59 - Stakeholder Engagement<br>63, 67 – Discussion<br>69-70 - Conclusion |
| 5: Reflections/critical perspective | Comment critically on PPI input in the study, reflecting on the things that went well and those that did not, so others can learn from this experience | 60-61 - Patient and Public Involvement |

## REFERENCES

1. NHS. NHS screening. 2021. Available from: https://www.nhs.uk/conditions/nhs-screening/ [accessed 7 September 2023].

2. NHS. Lung health checks. 2023. Available from: https://www.nhs.uk/conditions/lung-health-checks/ [accessed 7 September 2023].

3. Cancer Research UK. Cancer incidence for common cancers. Available from: https://www.cancerresearchuk.org/health-professional/cancer-statistics/incidence/common-cancers-compared [accessed 7 September 2023].

4. Grail. About Galleri. A new way to screen for more cancers. Available from: https://www.galleri.com/what-is-galleri [accessed 7th September 2023].

5. Grail. Breakthrough test performance. 2023. Available from: https://www.galleri.com/hcp/galleri-test-performance [accessed 21st September 2023].

6. Lennon AM, Buchanan AH, Kinde I, Warren A, Honushefsky A, Cohain AT, et al. Feasibility of blood testing combined with PET-CT to screen for cancer and guide intervention. Science 2020;369:03.

7. Gao Q, Zeng Q, Wang Z, Li C, Xu Y, Cui P, et al. Circulating cell-free DNA for cancer early detection. The Innovation 2022;3:100259.

8. Nguyen THH, Lu YT, Le VH, Bui VQ, Nguyen LH, Pham NH, et al. Clinical validation of a ctDNA-based assay for multi-cancer detection: an interim report from a Vietnamese longitudinal prospective cohort study of 2795 participants. Cancer Invest 2023;41:232–48.

9. Ranade A, Bhatt A, Page R, Limaye S, Crook T, Akolkar D, et al. Hallmark circulating tumor-associated cell clusters signify 230 times higher one-year cancer risk. Cancer Prev Res (Phila) 2021;14:11–6.

10. Xie L, Du X, Wang S, Shi P, Qian Y, Zhang W, et al. Development and evaluation of cancer differentiation analysis technology: a novel biophysics-based cancer screening method. Expert Rev Mol Diagn 2022;22:111–7.

11. Mikami H, Kimura O, Yamamoto H, Kikuchi S, Nakamura Y, Ando T, et al. A multicentre clinical validation of AminoIndex Cancer Screening (AICS). Scientific Reports 2019;9:7.

12. Brito-Rocha T, Constancio V, Henrique R, Jeronimo C. Shifting the cancer screening paradigm: the rising potential of blood-based multi-cancer early detection tests. Cells 2023;12:935.

13. NHS. The NHS Long Term Plan. 2019. Available from: https://www.longtermplan.nhs.uk/publication/nhs-long-term-plan/ [accessed 7 September 2023].

14. Dhruva SS, Smith-Bindman R, Redberg RF. The need for randomized clinical trials demonstrating reduction in all-cause mortality with blood tests for cancer screening. JAMA Intern Med 2023;183:1051–3.

15. Lee R, Robbins HA. PATHFINDER: another step on the uncharted path to multicancer screening. Lancet 2023;402:1213–5.

16. Etzioni R, Gulati R, Weiss NS. Multicancer early detection: Learning from the past to meet the future. J Natl Cancer Inst 2022;114:349–52.

17. Wise J. A blood test for multiple cancers: game changer or overhyped? BMJ 2022;378:o2279.

18. Mahase E. Clinicians raise concerns over pilot of blood test for multiple cancers. BMJ 2023;383:2268.

19. Centre for Reviews and Dissemination. Systematic reviews. CRD’s guidance for undertaking reviews in healthcare. York: CRD, University of York; 2009.

20. Page MJ, McKenzie JE, Bossuyt PM, Boutron I, Hoffmann TC, Mulrow CD, et al. The PRISMA 2020 statement: an updated guideline for reporting systematic reviews. BMJ 2021;372:n71.

21. Thomas J, Graziosi S, Brunton J, Ghouze Z, O’Driscoll P, Bond M, et al. EPPI-Reviewer: advanced software for systematic reviews, maps and evidence synthesis. London: EPPI Centre, UCL Social Research Institute, University College London; 2022.

22. Stevenson M, Sergeant E, Firestone S. epiR: tools for the analysis of epidemiological data. R package version 2.0.65. 2023. Available from: https://CRAN.R-project.org/package=epiR [accessed 24th October 2023].

23. R Core Team. R: a language and environment for statistical computing. R Foundation for Statistical Computing, Vienna, Austria 2023. Available from: https://www.R-project.org/ [accessed 24th October 2023].

24. Whiting PF, Rutjes AWS, Westwood ME, Mallett S, Deeks JJ, Reitsma JB, et al. QUADAS-2: A revised tool for the quality assessment of diagnostic accuracy studies. Ann Intern Med 2011;155:529–36.

25. Wickham H. ggplot2: elegant graphics for data analysis. [program] New York: Springer-Verlag; 2016.

26. Wade R, Nevitt S, Harden M, Dias S, Raine G, Khouja C, et al. *Multi-cancer early detection tests for screening*: PROSPERO 2023 CRD42023467901; 2023. Available from: https://www.crd.york.ac.uk/prospero/display_record.php?ID=CRD42023467901 [accessed 30th October 2023]

27. Healthwatch York. Available from: https://www.healthwatchyork.co.uk/ [accessed 22nd September 2023].

28. Hull York Medical School. Transforming cancer outcomes in Yorkshire. Available from: https://www.hyms.ac.uk/research/transform [accessed 22nd September 2023].

29. Humber and North Yorkshire Cancer Alliance. About us. Available from: https://hnycanceralliance.org.uk/about-us/ [accessed 22nd September 2023].

30. Schrag D, Beer TM, McDonnell CH, Nadauld L, Dilaveri CA, Reid R, et al. Blood-based tests for multicancer early detection (PATHFINDER): a prospective cohort study. Lancet 2023;402:1251–60.

31. Klein EA, Richards D, Cohn A, Tummala M, Lapham R, Cosgrove D, et al. Clinical validation of a targeted methylation-based multi-cancer early detection test using an independent validation set. Ann Oncol 2021;32:1167–77.

32. Liu MC, Oxnard GR, Klein EA, Swanton C, Seiden MV. Sensitive and specific multi-cancer detection and localization using methylation signatures in cell-free DNA. Ann Oncol 2020;31:745–59.

33. Cohen JD, Li L, Wang Y, Thoburn C, Afsari B, Danilova L, et al. Detection and localization of surgically resectable cancers with a multi-analyte blood test. Science 2018;359:926–30.

34. Beer TM. Shifting the paradigm for early cancer detection: a multi-biomarker class approach to MCED testing. Video of presentation from the 2023 ASCO annual meeting. Exact Sciences; 2023. Available from: https://www.exactsciences.com/Pipeline-and-Data/multi-cancer-early-detection/resources [accessed 19th October 2023].

35. Gene Solutions. SPOT-MAS multi cancer early detection. Available from: https://genesolutions.vn/en/product/spot-mas/ [accessed 24th October 2023].

36. Datar Cancer Genetics. Datar Cancer Genetics. Non-invasive solutions in cancer. Available from: https://datarpgx.com/ [accessed 24th October 2023].

37. Anpac Bio-Medical Science Company Limited. Anpac Bio. Available from: https://www.anpacbio.com/ [accessed 24th October 2023].

38. Ajinomoto Group. AminoIndex®. Available from: https://www.ajinomoto.com/innovation/action/aminoindex [accessed 24th October 2023].

39. Schrag D, McDonnell CH, Nadauld L, Dilaveri CA, Klein EA, Reid R, et al. A prospective study of a multi-cancer early detection blood test. Ann Oncol 2022;33:S961.

40. Beer TM, McDonnell CH, Nadauld L, Liu MC, Klein EA, Reid RL, et al. A prespecified interim analysis of the PATHFINDER study: Performance of a multicancer early detection test in support of clinical implementation. J Clin Oncol 2021;39:2.

41. Beer TM, McDonnell CH, Nadauld L, Liu MC, Klein EA, Reid RL, et al. Interim results of PATHFINDER, a clinical use study using a methylation-based multi-cancer early detection test. J Clin Oncol 2021;39:3010.

42. Klein E, Beer T. Assessing implementation of a multi-cancer early detection blood test for population screening in a clinical practice setting: prospective PATHFINDER cohort study. Cancer Prev Res (Phila) 2023;16:IA022.

43. Nadauld L, McDonnell CH, Liu MC, Klein E, Beer TM, Schrag D, et al. The PATHFINDER study: assessment of the implementation of an investigational multi-cancerearly detection test into clinical practice. Cancer Res 2020;80:CT291.

44. Nadauld LD, McDonnell CH, 3rd, Beer TM, Liu MC, Klein EA, Hudnut A, et al. The PATHFINDER study: assessment of the implementation of an investigational multi-cancer early detection test into clinical practice. Cancers (Basel) 2021;13:3501.

45. GRAIL LLC. Assessment of the implementation of an investigational multi-cancer early detection test into clinical practice. ClinicalTrials.gov. Bethesda (MD): National Library of Medicine (US); 2020. Available from: https://classic.clinicaltrials.gov/show/NCT04241796 [accessed 28th September 2023].

46. University of Oxford. SYMPLIFY – assessing a multi-cancer early detection test in individuals referred with signs and symptoms of cancer. ISCRTN Registry, BioMed Central Ltd.; 2021. Available from: https://www.isrctn.com/ISRCTN10226380 [accessed 28th September 2023].

47. Tang WHW, Yimer H, Tummala M, Shao S, Chung G, Clement J, et al. Performance of a targeted methylation-based multi-cancer early detection test by race and ethnicity. Prev Med 2023;167:107384.

48. Bryce AH, Thiel DD, Seiden MV, Richards D, Luan Y, Coignet M, et al. Performance of a cell-free DNA-based multi-cancer detection test in individuals presenting with symptoms suspicious for cancers. JCO Precis Oncol 2023;7:e2200679.

49. Shao SH, Allen B, Clement J, Chung G, Gao J, Hubbell E, et al. Multi-cancer early detection test sensitivity for cancers with and without current population-level screening options. Tumori 2023;109:335–41.

50. Rossi SH, Stewart GD. Re: Clinical validation of a targeted methylation-based multi-cancer early detection test using an independent validation set. Eur Urol 2022;82:442-3.

51. Klein EA, Richards D, Cohn A, Tummala M, Lapham R, Cosgrove D, et al. Clinical validation of a targeted methylation-based multi-cancer early detection test. Cancer Res 2021;81:LB013.

52. Klein EA, Richards D, Cohn A, Tummala M, Lapham R, Cosgrove D, et al. Detectingcancer signal across multiple cancers with one blood draw: validating a Multi-Cancer Early Detection (MCED) yest. In: American Academy of Family Physicians (AAFP) Family Medicine Experience (FMX), September 28-October 2, 2021. Anaheim, CA; 2021.

53. Venn O, Bredno J, Thornton A, Chang C, Hubbell E, Kurtzman K, et al. Robustness of a targeted methylation-based Multi-Cancer Early Detection (MCED) test to population differences in self-reported ethnicity. In: AACR Conference on The Science of Cancer Health Disparities in Racial/Ethnic Minorities and the Medically Underserved, September 29-October 2, 2023. Orlando, Florida; 2023.

54. Tang WHW, Yimer HA, Tummala MK, Shao S, Chung GG, Clement JM, et al. Performance of a targeted methylation-based multi-cancer early detection test by race/ethnicity. J Clin Oncol 2021;39:2.

55. Yimer HA, Tang WHW, Tummala MK, Shao S, Chung GG, Couch F, et al. Detection of cancer signal for over 50 AJCC cancer types with a multi-cancer early-detection test. J Clin Oncol 2021;39:2.

56. Papadopoulos N. A first-of-its-kind prospective study of amulti-cancer blood test to screen and manage 10,000 women with no history of cancer. Cancer Res 2020;80:CT022.

57. Suzuki M. Breast cancer screening by evaluating amino acid levels in the blood. Breast 2015;24:S69.

58. Nicholson BD, Oke J, Virdee PS, Harris DA, O’Doherty C, Park JE, et al. Multi-cancer early detection test in symptomatic patients referred for cancer investigation in England and Wales (SYMPLIFY): a large-scale, observational cohort study. Lancet Oncol 2023;24:733–43.

59. Cance W, Lilyestrom W, Johnson T, Winn R, Jemal A. Employer-based implementation of Galleri® multi-cancer early detection testing to address socioeconomic disparities in receipt of screening. In: AACR Conference on The Science of Cancer Health Disparities in Racial/Ethnic Minorities and the Medically Underserved, September 29-October 2, 2023. Orlando, Florida; 2023. p. C108.

60. Neal RD, Johnson P, Clarke CA, Hamilton SA, Zhang N, Kumar H, et al. Cell-free DNA-based multi-cancer early detection test in an asymptomatic screening population (NHS-Galleri): design of a pragmatic, prospective randomised controlled trial. Cancers (Basel) 2022;14:4818.

61. GRAIL LLC. PATHFINDER 2: A multi-cancer early detection study. ClinicalTrials.gov. Bethesda (MD): National Library of Medicine (US); 2021. Available from: https://classic.clinicaltrials.gov/show/NCT05155605 [accessed 13th October 2023].

62. GRAIL LLC. REFLECTION: a clinical practice learning program for Galleri®. ClinicalTrials.gov. Bethesda (MD): National Library of Medicine (US); 2022. Available from: https://classic.clinicaltrials.gov/show/NCT05205967 [accessed 13th October 2023].

63. University College London. The SUMMIT study: a cancer screening study. ClinicalTrials.gov. Bethesda (MD): National Library of Medicine (US); 2019. Available from: https://classic.clinicaltrials.gov/show/NCT03934866 [accessed 13th October 2023].

64. Nguyen VT, Nguyen TH, Doan NN, Pham TM, Nguyen GT, Tran TT, et al. Multimodal analysis of methylomics and fragmentomics in plasma cell-free DNA for multi-cancer early detection and localization. medRxiv [Preprint] 1 August 2023 [cited 13 October 2023]. Available from: 10.1101/2023.04.12.23288460

65. Maeda S, Miyagi E, Yao M, Ichikawa Y, Rino Y. AminoIndex^TM^ Cancer Screening (AICS) follow-up study, interim analysis report. Cancer Sci 2017;109:P-2386.

66. Suzuki M. Usefulness of AminoIndex Cancer Screening (AICS) for breast cancer screening. Eur J Cancer 2014;50:S64.

67. Dempsey AA, Tripp JH, Chao S, Stamatiou D, Pilcz T, Ying J, et al. Aristotle: a single blood test for pancancer screening. J Clin Oncol 2020;38:e15037.

68. Arber N, Shapira S, Kazanov D, Fokra A, Sally Z, Zigdon S, et al. CD24 predictive levels-a simple novel blood test for early detection of various malignancies. United European Gastroenterology Journal 2017;5(5 Supplement 1):A765.

69. Massarwi S, Shapira S, Kazanov D, Asido S, Hay-Levy M, Matatov G, et al. CD 24 for early detection and surveillance of cancer using a universal simple blood test. United European Gastroenterol J 2019;7:407.

70. Shapira S, Kazanov D, Shimon MB, Levy MH, Mdah F, Asido S, et al. The dark age of single organ screening is over: CD24 is a novel universal simple blood test for early detection of cancer. J Clin Oncol 2020;38:e15591.

71. Shapira S, Kazanov D, Shimon MB, Mdah F, Levy MH, Galazan L, et al. The dark age of single organ screening is over: CD24 Is a novel universal pan-cancer blood test for early detection of cancer. Gastroenterology 2021;160:S-130.

72. Madah F, Kazanov D, Motlaq M, Argaman L, Shaked M, Shenberg G, et al. The dark age of single organ screening is over: CD24 is a novel universal pan-cancer blood test for early detection of cancer. Cancer Res 2023;83:3342.

73. Luan Y, Zhong G, Li S, Wu W, Liu X, Zhu D, et al. A panel of seven protein tumour markers for effective and affordable multi-cancer early detection by artificial intelligence: a large-scale and multicentre case-control study. EClinicalMedicine 2023;61:102041.

74. Mao M, Luan Y, Zhong G, Li S, Wu W, Liu X, et al. A panel of seven protein tumour markers for effective and affordable multi-cancer early detection by artificial intelligence. JCO Glob Oncol 2023;9:155.

75. Mao M, Li S, Ren Q, Luan Y, Liang W, Geng S, et al. Integrating multi-omics features for blood-based multi-cancer early detection. JCO Glob Oncol 2023;9:156.

76. Cision. SeekIn presents new data supporting its pan-cancer early detection test at the Early Detection of Cancer Conference 2022. Cision; 2022. Available from: https://www.prnewswire.com/news-releases/seekin-presents-new-data-supporting-its-pan-cancer-early-detection-test-at-the-early-detection-of-cancer-conference-2022-301652662.html [accessed 26th October 2023].

77. Gao Q, Lin YP, Li BS, Wang GQ, Dong LQ, Shen BY, et al. Unintrusive multi-cancer detection by circulating cell-free DNA methylation sequencing (THUNDER): development and independent validation studies. Ann Oncol 2023;34:486–95.

78. Gao Q, Li B, Xu J, Fang S, Qiu F, Su J, et al. Analysis of epigenomic signatures in cell-free DNA (cfDNA) from cancer patients and high-risk controls: a blinded test cohort of THUNDER-II study. J Clin Oncol 2021;39:e22518.

79. Guangzhou Burning Rock Dx. Multi-cancer early-detection test in asymptomatic individuals (PREVENT). ClinicalTrials.gov. Bethesda (MD): National Library of Medicine (US); 2022. Available from: https://classic.clinicaltrials.gov/show/NCT05227534 [accessed 28th September 2023].

80. Shanghai Zhongshan Hospital. Pan-canceR Early DetectIon projeCT (PREDICT). ClinicalTrials.gov. Bethesda (MD): National Library of Medicine (US); 2021. Available from: https://classic.clinicaltrials.gov/show/NCT04817306 [accessed 17th October 2023].

81. Salat A, Voigt W, von und zu Zwerger B. Prospective and single-blinded evaluation of the multi-cancer Carcimun-test. Precision Cancer Medicine 2022;5 10.21037/pcm-21-35.

82. Ma Y, Zhao G, Wang K, Song L, Xiong S, Li H, et al. Clinical performance of a liquid biopsy test based on the detection of multiple DNA methylation biomarkers for early detection of gastrointestinal cancers. J Clin Oncol 2022;40:e16096.

83. McGuinness LA. robvis: an R package and web application for visualising risk-of-bias assessments. 2019. Available from: https://CRAN.R-project.org/package=robvis [accessed 27th November 2023].

84. Westgate C, Kingsbury D, Poliak M, Lipton J, McMillin M, Malinow LB, et al. Early real-world experience with a multi-cancer early detection test In: American Society of Clinical Oncology (ASCO), June 2–6, 2023. Chicago, IL; 2023.

85. Lennon AM, Buchanan AH, Rego SP, A CO, Elias PZ, Sadler JR, et al. Outcomes in participants with a false positive multi-cancer early detection (MCED) test: results from >4 years follow-up from DETECT-A, the first large, prospective, interventional MCED study. J Clin Oncol 2023;41:3039.

86. Buchanan AH, Lennon AM, Rego SP, Choudhry OA, Elias PZ, Sadler JR, et al. Long-term clinical outcomes of cancers diagnosed following detection by a blood-based multi-cancer early detection (MCED) test. J Clin Oncol 2023;41:3037.

87. McDonnell CH, Hudnut AG, Behl D, Ang R, Spinelli B, Jacobs D, et al. Diagnostic workup following a multicancer early detection test with a cancer signal origin prediction. J Clin Oncol 2022;40:e15037.

88. Klein EA, McDonnell CH, Nadauld L, Dilaveri CA, Reid R, Marinac CR, et al. Clinical evaluation of Cancer Signal Origin (CSO) prediction and diagnostic resolution following multi-cancer early detection testing. In: American Society of Clinical Oncology (ASCO), June 2–6, 2023. Chicago, IL; 2023.

89. Schrag D, Beer TM, McDonnell CH, Nadauld L, Dilaveri CA, Klein EA, et al. Evaluation of anxiety, distress and satisfaction with a multi-cancer early detection test. Ann Oncol 2022;33:S963.

90. GRAIL. Written evidence submitted by GRAIL (FCR0024) to the Health and Social Care Committe Future Cancer inquiry: UK Parliament Health and Social Care Committee; 2023. Available from: https://committees.parliament.uk/writtenevidence/120627/pdf/ [accessed 30th October 2023].

91. Cristiano S, Leal A, Phallen J, Fiksel J, Adleff V, Bruhm DC, et al. Genome-wide cell-free DNA fragmentation in patients with cancer. Nature 2019;570:385–9.

92. Annapragada AV, Medina JE, Lof P, Mathios D, Foda ZH, Noe M, et al. Early detection of ovarian cancer using cell-free DNA fragmentomes. Cancer Res 2023;83:773.

93. Xu L, Wang J, Yang T, Tao J, Liu X, Ye Z, et al. Toward the development of a $100 screening test for 6 major cancer types. Cancer Res 2020;80:4061.

94. Xu L, Wang J, Ma W, Liu X, Li S, Chen S, et al. Validation of a high performing blood test for multiple major cancer screenings. J Clin Oncol 2021;39:10561.

95. Sun Yat-sen University Cancer Center. PanTum technique for the detection of peripheral blood APO10 and TKTL1 in the diagnosis of high incidence of malignant tumors in Chinese population. Chinese Clinical Trial Registry; 2020. Available from: http://www.chictr.org.cn/showproj.aspx?proj=64757 [accessed 28th September 2023].

96. Millennium Oncology India Private Limited. A trial for confirming the accuracy of PanTum test for solid tumor detection. Clinical Trials Registry India; 2022. Available from: http://www.ctri.nic.in/Clinicaltrials/pmaindet2.php?trialid=73694 [accessed 28th September 2023].

97. Guardant Health Inc. Screening for high frequency malignant disease. ClinicalTrials.gov. Bethesda (MD): National Library of Medicine (US); 2021. Available from: https://classic.clinicaltrials.gov/show/NCT05117840 [accessed 13th October 2023].

98. Nguyen H, Raymond VM, Vento-Gaudens E, Marino E, Lang K, Eagle C. Screening for high frequency malignant disease (SHIELD). J Clin Oncol 2022;40:TPS1602.

99. Raymond V, Nguyen H, Cotton L, Vento-Gaudens E, Eagle C. PP01.20 Trial in progress: screening for high frequency malignant disease (SHIELD). J Thorac Oncol 2023;18:e19.

100. Harbinger Health. Development and validation of Harbinger Health Test for early cancer detection. ClinicalTrials.gov. Bethesda (MD): National Library of Medicine (US); 2022. Available from: https://classic.clinicaltrials.gov/show/NCT05435066 [accessed 17th October 2023].

101. Adela Inc. cfDNA assay prospective observational validation for early cancer detection and minimal residual disease. ClinicalTrials.gov. Bethesda (MD): National Library of Medicine (US); 2022. Available from: https://classic.clinicaltrials.gov/show/NCT05366881 [accessed 29th September 2023].

102. Newman AM, Bratman SV, To J, Wynne JF, Eclov NC, Modlin LA, et al. An ultrasensitive method for quantitating circulating tumor DNA with broad patient coverage. Nat Med 2014;20:548–54.

103. Mazzone PJ, Wong KK, Tsay JCJ, Pass HI, Vachani A, Ryan A, et al. Prospective evaluation of cell-free DNA fragmentomes for lung cancer detection. Cancer Res 2023;83:5766.

104. CASCADE-LUNG: cancer screening assay using DELFI; a clinical validation study in lung. ClinicalTrials.gov. Bethesda (MD): National Library of Medicine (US); Available from: https://classic.clinicaltrials.gov/show/NCT05306288 [accessed 15th September 2023].

105. Early detection of de novo cancer in liver transplant recipients. ClinicalTrials.gov. Bethesda (MD): National Library of Medicine (US); Available from: https://classic.clinicaltrials.gov/show/NCT05492617 [accessed 15th September 2023].

106. Cameron JM, Sala A, Antoniou G, Brennan PM, Butler HJ, Conn JJ, et al. A spectroscopic liquid biopsy for the earlier detection of multiple cancer types. Br J Cancer 2023:10.1038/s41416-023-02423-7.

107. Vincere Cancer Centre. Multi-Cancer Early Detection (MCED) of firefighters. ClinicalTrials.gov. Bethesda (MD): National Library of Medicine (US); 2023. Available from: https://classic.clinicaltrials.gov/show/NCT05780957 [accessed 28th September 2023].

108. Harbinger Health. Press release - Harbinger Health raises $140 million in series B funding to accelerate advancement of its screening platform for early-stage cancers. Harbinger Health; 2023. Available from: https://www.harbinger-health.com/news/harbinger-health-raises-140-million-in-series-b-funding-to-accelerate-advancement-of-its-screening-platform-for-early-stage-cancers [accessed 26th October 2023].

109. Bao H, Wang Z, Ma X, Guo W, Zhang X, Tang W, et al. Letter to the Editor: An ultra-sensitive assay using cell-free DNA fragmentomics for multi-cancer early detection. Mol Cancer 2022;21:129.

110. Nanjing Shihejiyin Technology. The Jinling cohort. ClinicalTrials.gov. Bethesda (MD): National Library of Medicine (US); 2023. Available from: https://classic.clinicaltrials.gov/show/NCT06011694 [accessed 13th October 2023].

111. Zhang H, Zhao L, Jiang J, Zheng J, Yang L, Li Y, et al. Multiplexed nanomaterial-assisted laser desorption/ionization for pan-cancer diagnosis and classification. Nature communications 2022;13:617.

112. Fudan University Taizhou Institute of Health Sciences. A prospective, multicenter cohort study of pan-cancer screening in Chinese population. Chinese Clinical Trial Registry; 2021. Available from: http://www.chictr.org.cn/showproj.aspx?proj=141068 [accessed 30th September 2023].

113. Singlera Genomics. The FuSion Program: a prospective and multicenter cohort study of pan-cancer screening in Chinese population. ClinicalTrials.gov. Bethesda (MD): National Library of Medicine (US); 2021. Available from: https://classic.clinicaltrials.gov/show/NCT05159544 [accessed 13th October 2023].

114. Suo C, Zhao R, Jiang Y, Zhang Y, He Q, Su Z, et al. The FuSion Project of pan-cancer early screening in chinese - an integrative study by Fudan University and Singlera. Cancer Res 2023;83:4194.

115. PanTum Detect. PanTum Detect; 2023. Available from: https://www.pantumdetect.com/ [accessed 26th October 2023].

116. Saman S, Stagno MJ, Warmann SW, Malek NP, Plentz RR, Schmid E. Biomarkers Apo10 and TKTL1: Epitope-detection in monocytes (EDIM) as a new diagnostic approach for cholangiocellular, pancreatic and colorectal carcinoma. Cancer Biomark 2020;27:129–37.

117. Epigeneres Biotech Pvt Ltd. A simple blood test to understand presence or absence of cancer. Clinical Trials Registry India; 2023. Available from: http://www.ctri.nic.in/Clinicaltrials/pmaindet2.php?trialid=87700 [accessed 24th October 2023].

118. Phallen J, Sausen M, Adleff V, Leal A, Hruban C, White J, et al. Direct detection of early-stage cancers using circulating tumor DNA. Sci Transl Med 2017;9:eaan2415.

119. Peking University Shenzhen Hospital. SZ-PILOT study: prospective observational study of the YiDiXueTM multi-cancer early detection kit in multi-cancer early screening in normal people. Chinese Clinical Trial Registry; 2022. Available from: https://www.chictr.org.cn/showproj.html?proj=187882 [accessed 30th September 2023].

120. Staniszewska S, Brett J, Simera I, Seers K, Mockford C, Goodlad S, et al. GRIPP2 reporting checklists: tools to improve reporting of patient and public involvement in research. BMJ 2017;358:j3453.

121. Baker D, Middleton E. Cervical screening and health inequality in England in the 1990s. J Epidemiol Community Health 2003;57:417–23.

122. Chancellor M, Modi J, Muhammad R, Batioja K, Garrett E, Waters P, et al. Health inequities in mammography: a scoping review. Eur J Radiol 2023;160:110693.

123. Mosquera I, Mendizabal N, Martín U, Bacigalupe A, Aldasoro E, Portillo I. Inequalities in participation in colorectal cancer screening programmes: a systematic review. Eur J Public Health 2020;30:416–25.

124. McLeod M, Kvizhinadze G, Boyd M, Barendregt J, Sarfati D, Wilson N, et al. Colorectal cancer screening: how health gains and cost-effectiveness vary by ethnic group, the impact on health inequalities, and the optimal age range to screen. Cancer Epidemiol Biomarkers Prev 2017;26:1391–400.

125. Sarfati D, Shaw C, Simmonds S. Commentary: Inequalities in cancer screening programmes. Int J Epidemiol 2010;39:766–8.

126. Alam Z, Cairns JM, Scott M, Dean JA, Janda M. Interventions to increase cervical screening uptake among immigrant women: a systematic review and meta-analysis. PLoS One 2023;18:e0281976.

127. Cairns JM, Greenley S, Bamidele O, Weller D. A scoping review of risk-stratified bowel screening: current evidence, future directions. Cancer Causes Control 2022;33:653–85.

128. De Mil R, Guillaume E, Launay L, Guittet L, Dejardin O, Bouvier V, et al. Cost-effectiveness analysis of a mobile mammography unit for breast cancer screening to reduce geographic and social health inequalities. Value Health 2019;22:1111–8.

129. National Cancer Institute Division of Cancer Prevention. Multi-Cancer Detection (MCD) research. National Cancer Institute, U.S. National Institutes of Health; Available from: https://prevention.cancer.gov/major-programs/multi-cancer-detection-mcd-research [accessed 30th October 2023].

130. Kaiser J. ‘The complexities are staggering.’ U.S. plans huge trial of blood tests for multiple cancers. Science 2022:10.1126/science.add6151.

131. Duffy MJ, Diamandis EP, Crown J. Circulating tumor DNA (ctDNA) as a pan-cancer screening test: is it finally on the horizon? Clin Chem Lab Med 2021;59:1353–61.

132. Callister MEJ, Crosbie EJ, Crosbie PAJ, Robbins HA. Evaluating multi-cancer early detection tests: an argument for the outcome of recurrence-updated stage. Br J Cancer 2023;129:1209–11.

133. Menon U, Gentry-Maharaj A, Burnell M, Singh N, Ryan A, Karpinskyj C, et al. Ovarian cancer population screening and mortality after long-term follow-up in the UK Collaborative Trial of Ovarian Cancer Screening (UKCTOCS): a randomised controlled trial. Lancet 2021;397:2182–93.

134. Martin RM, Donovan JL, Turner EL, Metcalfe C, Young GJ, Walsh EI, et al. Effect of a low-intensity PSA-based screening intervention on prostate cancer mortality: the CAP randomized clinical trial. JAMA 2018;319:883–95.

135. LeeVan E, Pinsky P. Predictive performance of cell-free nucleic acid-based multi-cancer early detection tests: a systematic review. Clin Chem 2023:hvad134.

136. Pons-Belda OD, Fernandez-Uriarte A, Diamandis EP. Can circulating tumor DNA support a successful screening test for early cancer detection? The Grail paradigm. Diagnostics (Basel) 2021;11:2171.

137. Brentnall AR, Mathews C, Beare S, Ching J, Sleeth M, Sasieni P. Dynamic data-enabled stratified sampling for trial invitations with application in NHS-Galleri. Clin Trial 2023;20:425–33.

138. Marlow LA, Schmeising-Barnes N, Warwick J, Waller J. Psychological impact of the Galleri test (sIG(n)al): Protocol for a longitudinal evaluation of the psychological impact of receiving a cancer signal in the NHS-Galleri trial. BMJ Open 2023;13:e072657.

